# Integrative Proteome- and Phenome-Wide Assessment Uncovers Causal Protein Drivers and Drug Targets for Heterogeneous Kidney Diseases

**DOI:** 10.1101/2025.10.09.25337438

**Authors:** Jefferson L. Triozzi, Fatih Mamak, Otis D. Wilson, Hua-Chang Chen, Zhihong Yu, Kai Gravel-Pucillo, Brian R. Ferolito, Kelly Cho, John Michael Gaziano, Sumitra Muralidhar, T. Alp Ikizler, Cassianne Robinson-Cohen, Ayush Giri, Ran Tao, Alexandre C. Pereira, Adriana M. Hung, the VA Million Veteran Program

**Author notes:** **Corresponding Author:** Adriana M. Hung, MD, MPH, Division of Nephrology and Hypertension, Department of Medicine, Vanderbilt University Medical Center, 1161 21st Ave South, MCN S-3223, Nashville, TN 37232-2372.

## Abstract

Interpreting proteomic associations with chronic kidney disease (CKD) is challenging due to the disease’s clinical heterogeneity and complex overlap with systemic conditions. We present a framework that identifies causal circulating protein drivers of CKD and delineates their subtype-specific and systemic effects using electronic health record (EHR) data at biobank scale. Using proteome-wide Mendelian randomization, we instrumented cis-acting protein quantitative trait loci for 2,807 circulating proteins and tested them against detailed, EHR-based kidney function outcomes in 464,631 Million Veteran Program participants. Proteins were mapped to nine kidney disease subtypes defined by genome-wide association meta-analyses from the Million Veteran Program, UK Biobank, and FinnGen. Phenome-wide association studies across 1,020 traits distinguished renal versus extra-renal associations. This integrative strategy prioritizes 93 proteins with proteome-wide significance for kidney outcomes, demonstrates subtype-specific relevance, exposes systemic associations, and maps therapeutic targets to nominate candidates for drug development and repurposing.

## Introduction

Drug-target Mendelian Randomization (MR) is a strategy for validating therapeutic mechanisms, identifying drug repurposing opportunities, and nominating novel drug targets for complex traits such as kidney diseases^1–3^. It leverages genetic variants as instrumental variables to infer causal relationships between drug-target modulation and clinical outcomes, often using gene or protein expression data^1^. In proteomics-based MR, cis-acting protein quantitative trait loci (cis-pQTLs) serve as genetic instruments to estimate the effects of circulating protein levels on disease endpoints^4^. In nephrology, the current benchmark is creatinine-based estimated glomerular filtration rate (eGFR) due to its routine measurement and broad availability in clinical and research settings.^5^ Large-scale genome-wide association studies (GWAS) of eGFR have yielded fundamental insights into the genetics of kidney function with high statistical power^6^. However, eGFR is a static snapshot that may overlook dynamic, longitudinal measures of the onset and progression of kidney disease. Moreover, eGFR is an aggregate of diverse pathophysiological processes, limiting its ability to capture the mechanistic heterogeneity that defines subtypes of kidney disease.

These limitations are relevant for proteome-wide association studies of kidney traits, where causal inference can be vulnerable to misclassification of outcomes, mediation through intermediate traits, and other confounders. Circulating proteins exerting effects beyond the kidney, such as those influencing cardiometabolic, immunologic, or other pathways, make it difficult to determine whether a Mendelian randomization signal reflects a direct renal mechanism^7^. Such effects may reflect a shared upstream biology affecting both kidney and other tissues, a renal effect with systemic consequences, or a systemic effect with renal consequences. This uncertainty is a gap between genomic discovery and actionable insight into how modulating a given protein would affect kidney function.

To address these challenges, a comprehensive MR framework is required. It should incorporate kidney outcomes derived from longitudinal electronic health records, enabling the capture of disease onset, progression, and subtype-specific associations. It should also characterize systemic pleiotropy by mapping the associations of genetic instruments across the phenome, thereby distinguishing MR-inferred drug targets that act directly on the kidney from those with on-target or off-target effects in other organ systems. High-throughput phenotyping, using diagnosis-codes linked to large biobanks, can help enhance precision by “zooming in” on specific kidney disease subtypes and “zooming out” on extra-renal pathways across the phenome.^8,9^

In this study, we conducted a comprehensive proteome-wide association study (PWAS) to evaluate causal relationships between circulating protein levels and kidney function decline (**Figure 1**). First, we performed proteome-wide MR using cis-pQTLs from four large-scale proteomic platforms covering 2,807 circulating proteins. These instruments were tested against longitudinal, electronic health record-derived kidney outcomes in 464,000 Million Veteran Program (MVP) participants, including eGFR, incident chronic kidney disease (CKD), progression to end-stage kidney disease (ESKD), and annualized relative eGFR slope decline estimated via linear mixed-effects models. Aggregate association testing combined evidence across the kidney outcomes to identify proteins with significant causal effects. Second, we applied two-sample MR to map each protein to nine kidney disease subtypes, defined by genome-wide association study meta-analyses from the MVP, UK Biobank, and FinnGen. Third, we evaluated systematic phenotype pleiotropy for each protein using phenome-wide association studies (PheWAS)^9^. Finally, we prioritized therapeutic opportunities by linking significant proteins to established drug target databases, constructing networks demonstrating the relationship between drugs, proteins, kidney disease, and systemic conditions. The renal effects of drugs targeting these proteins or their closely related ligands were interpreted in this context.

**Figure 1.**
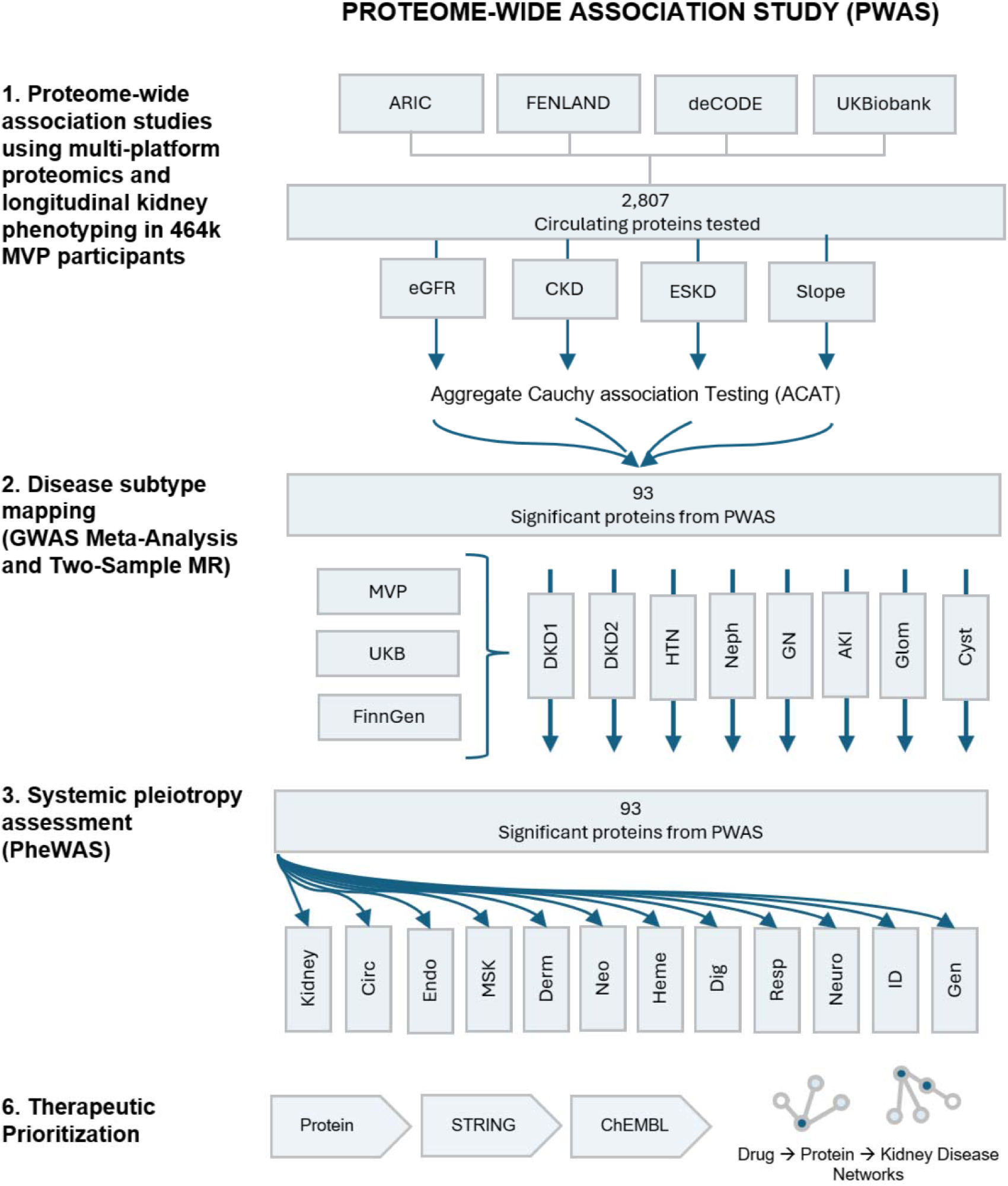
Roadmap of study design. There was a sequential analytic framework applied in this study. We first performed a proteome-wide Mendelian randomization study of 2,807 cis-pQTL instruments derived from four proteomics cohorts (ARIC, Fenland, deCODE, UK Biobank) against a multi-trait kidney outcome built from the Million Veteran Program electronic health records, integrating baseline eGFR, incident chronic kidney disease (CKD), progression to end-stage kidney disease (ESKD), and annualized eGFR slope. Proteins reaching proteome-wide significance (n = 93) were next interrogated in two-sample MR analyses across 9 ICD-code defined kidney disease subtypes using a genome-wide association meta-analysis combining FinnGen, MVP and UK Biobank datasets. The same proteins (n = 93) were queried against 1020 phecodes in the MVP gwPheWAS to evaluate extra-renal pleiotropy. Therapeutic potential of proteins were discussed based on high-confidence protein-protein-interaction map in STRING and druggability annotation using ChEMBL.

## Results

The PWAS included up to 464,631 European ancestry individuals in the VA MVP (**Table 1**). Participants were predominantly male Veterans (mean age 64.3 ± 13.8 years, 92.6 % male) with a body mass index in the obese range (30.1 ± 6.0 kg/m^2^) and a high burden of cardiometabolic comorbidities. The clinical load was heavier in the case subsets (diabetes and hypertension affected 39.3% and 75.8% of chronic kidney disease cases and 59.9% and 88.5% of end-stage kidney disease cases, respectively). Baseline risk factors were moderately controlled (mean systolic/diastolic blood pressure 129/76 mm Hg, mean HbA1c 6.2%), and serial creatinine measurements for 109,570 participants enabled estimation of annualized eGFR slope.

**Table 1.** Baseline characteristics of the Million Veteran Program study population in the proteome-wide association study.

|  | eGFR | eGFR Slope | Chronic Kidney Disease |  | End-Stage Kidney Disease |  |
| --- | --- | --- | --- | --- | --- | --- |
| Sample Size | N = 464631 | N = 109570 | N = 130263 Cases | N = 327645 Controls | N = 6723 Cases | N = 457908 Controls |
| Age, mean (SD) | 64.27 (13.76) | 67 (9) | 72.35 (9.48) | 60.98 (13.92) | 68.25 (9.86) | 64.21 (13.80) |
| Female, N (%) | 34414 (7.4) | 3,917 (3.5) | 6184 (4.75) | 28032 (8.6) | 198 (2.9) | 34216 (7.5) |
| Male, N (%) | 430216 (92.6) | 105,653 (96.5) | 124079 (95.3) | 299612 (91.4) | 6525 (97.1) | 423691 (92.5) |
| Systolic blood pressure, mm Hg, mean (SD) | 129.38 (15.59) | 134 (18) | 130.63 (16.61) | 128.73 (14.97) | 134.65 (19.39) | 129.30 (15.50) |
| Diastolic blood pressure, mm Hg, mean (SD) | 75.83 (10.25) | 83 (10) | 73.45 (10.48) | 76.91 (9.92) | 72.97 (11.59) | 75.88 (10.22) |
| HbA1c, %, mean (SD) | 6.19 (1.22) | 6.6 (1.0) | 6.51 (1.28) | 6.04 (1.14) | 6.85 (1.60) | 6.18 (1.21) |
| Body mass index, kg/m2, mean (SD) | 30.11 (5.96) | 32 (6) | 30.33 (5.94) | 30.0 (5.96) | 30.82 (6.38) | 30.10 (5.95) |
| Diabetes, N (%) | 116716 (25.1) | 40.80% | 51162 (39.3) | 61524 (18.8) | 4030 (59.9) | 112686 (24.6) |
| Hypertension, N (%) | 266978 (57.7) | 50% | 98,746 (75.8) | 162285 (49.5) | 5947 (88.5) | 261031 (57.0) |

### Main Findings

We assembled 2,807 unique plasma proteins instrumented by cis-pQTLs within ± 500 kb of each gene’s transcription start site from GWAS from four proteomic datasets (ARIC, Fenland, deCODE, and UK Biobank. Each protein was interrogated for associations with four components of the primary kidney outcome (eGFR, eGFR slope, CKD, and ESKD) (**Figure 2**, **Supplementary** Figures 1-4). Because the kidney outcomes are biologically related and therefore not statistically independent, we combined the MR *P*-values for every protein using aggregate Cauchy association testing (ACAT), yielding one summary *P*-value per protein (**Supplementary Table 1-5**). Applying a Bonferroni threshold, we identified 93 proteins with experiment-wide significance. Of these, 38 proteins were statistically significant in ≥ two proteomic datasets, 19 in ≥ three proteomic datasets, and 7 in all four proteomic datasets (*AGER, GSTA1, IDI2, INHBB, INHBC, MFAP4, MST1*) (**Figure 3**).

**Figure 2.**
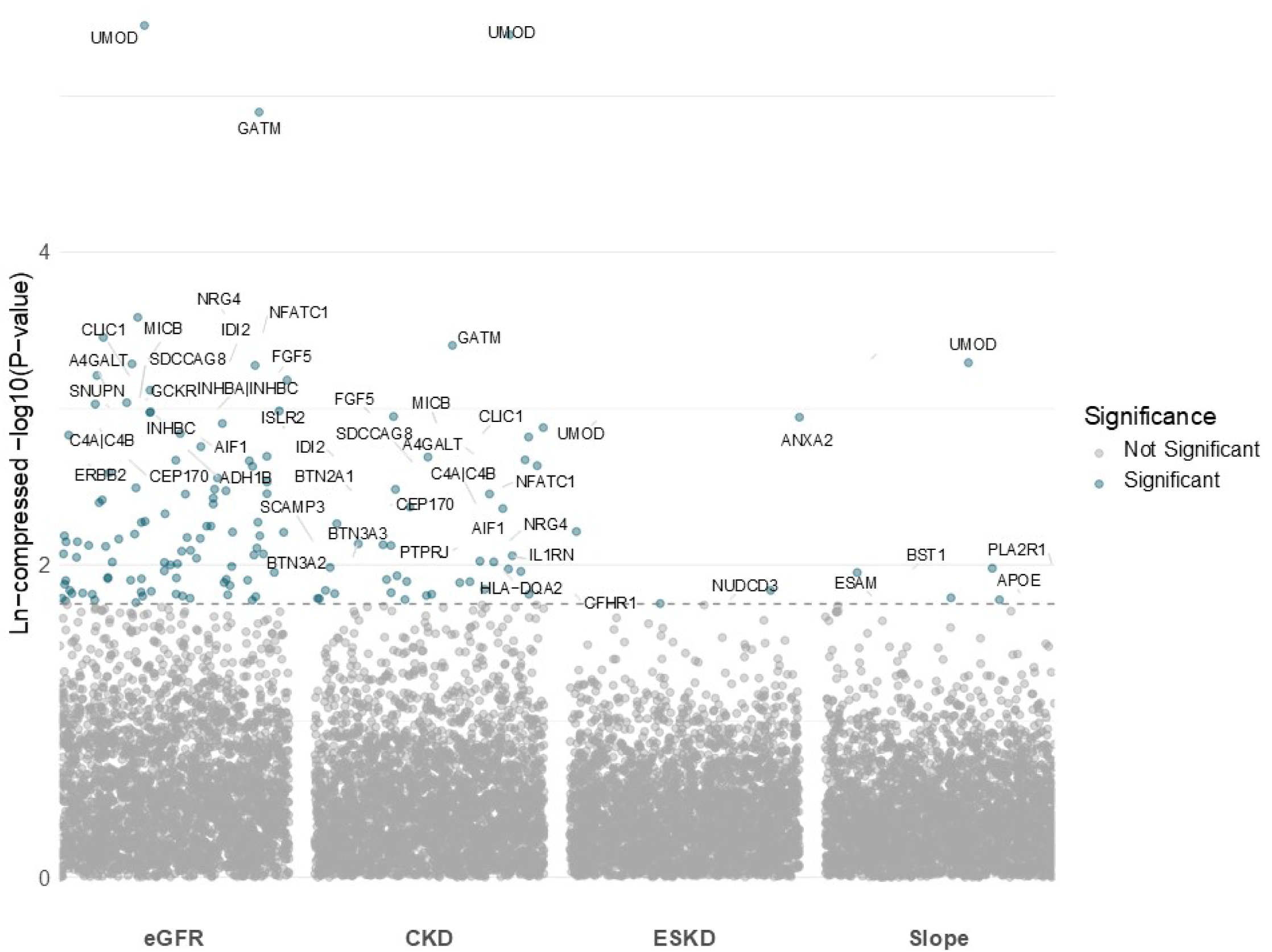
Outcome-stratified Manhattan plot of proteome-wide Mendelian randomization associations between circulating proteins and kidney function outcomes. Each dot is a Mendelian randomization (MR) association between a circulating protein for a given component of the primary kidney outcome: estimated glomerular filtration rate (eGFR), chronic kidney disease (CKD), end-stage kidney disease (ESKD) and annual eGFR slope. The x-axis is partitioned into the four components of the primary kidney outcome, and the y-axis is a natural log compressed -log10(*P*-value). Points are colored blue when they surpass the Bonferroni-corrected threshold (dashed line).

**Figure 3.**
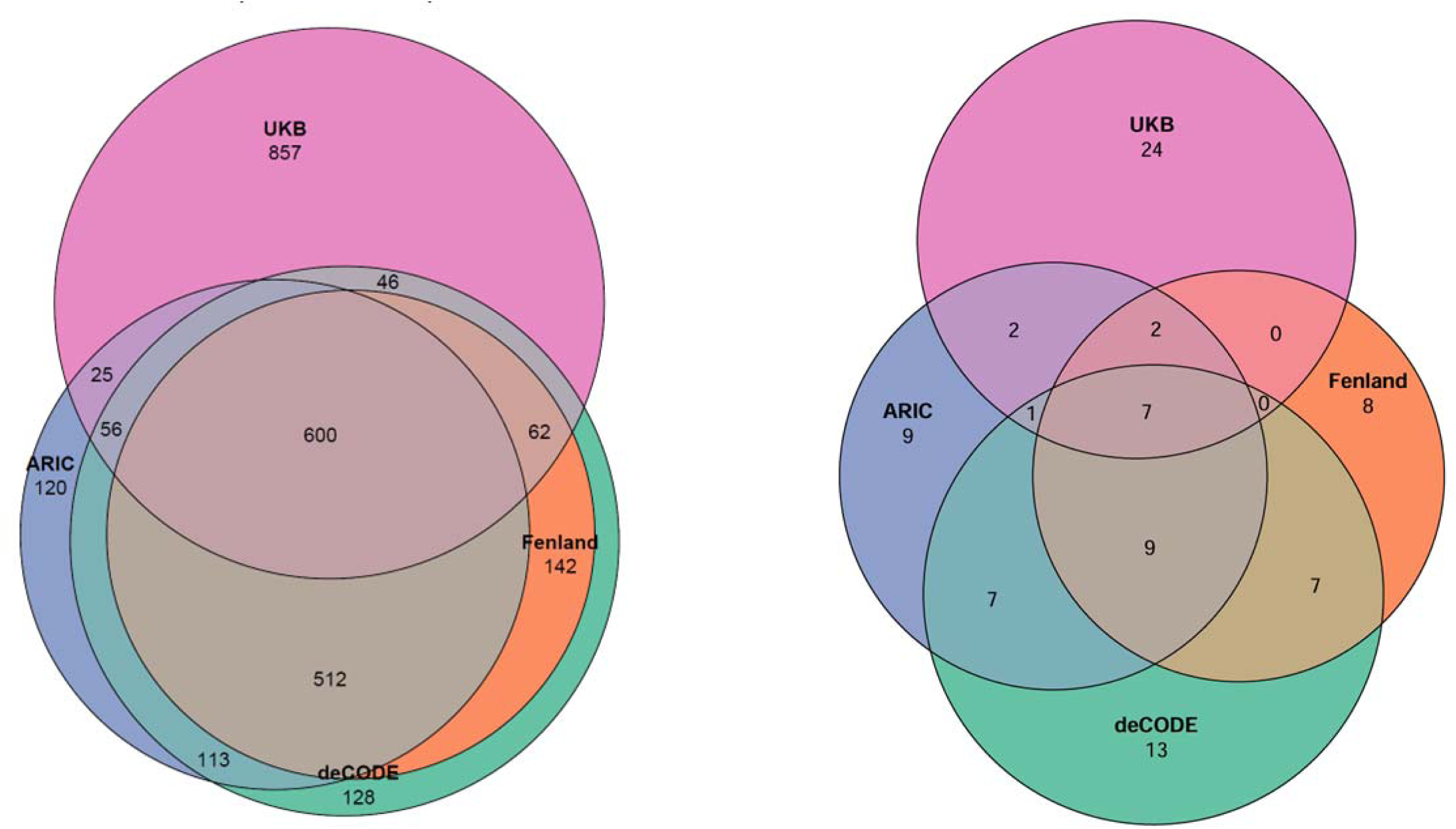
Overlap of genetically instrumented plasma proteins measured across four proteomic datasets (left) and those that were significantly associated with the primary kidney outcome across four proteomic datasets (right). Venn diagrams illustrate the overlap among plasma proteins analyzed (left panel) and the subset identified as significantly associated with the primary kidney outcome (right panel) across four independent proteomic datasets: Atherosclerosis Risk in Communities (ARIC), Fenland Study (Fenland), deCODE genetics (deCODE), and UK Biobank Olink proteomics (UKB). Numbers indicate unique or shared proteins identified in each intersection between proteomic datasets.

### Sensitivity Analyses

The 93 circulating plasma proteins identified in the PWAS underwent further prioritization based on heterogeneity and pleiotropy testing, cross-endpoint consistency, genetic colocalization, and external replication in the CKDGen Consortium. For multi-SNP pQTL instruments, we applied Cochran’s Q (consistency across SNP effects) and MR-Egger (pleiotropy). There were 70/93 proteins demonstrating consistency of MR effects, defined as uniformly protective (higher eGFR, slower decline, lower CKD/ESKD risk) or uniformly detrimental (lower eGFR, faster decline, higher CKD/ESKD risk) (**Supplementary Table 6, Supplementary** Figures 5-8). When stratified by dataset, the proportion of proteins with directional consistency was highest in ARIC (75.0%), followed by deCODESoma (73.9%), Fenland (71.1%), and UKBBOlink (69.4%). These differences could reflect variability in study design, population, the underlying biology of the associations, or the proteomic measurement platform. Colocalization supported a shared causal variant for 28/93 proteins (**Supplementary Table 7**). There were 50 proteins (54%) replicated externally, using aggregate association across eGFR, CKD, i25, and rapid3 kidney outcomes in the CKDGen Consortium (**Supplementary Table 8**). Overall, 12 statistically significant proteins passed all sensitivity testing criteria: *ANGPTL3*, *ADH1B*, *FAM213A*, *GCKR*, *GSTA1*, *IDI2*, *INHBA*, *INHBC*, *MANBA*, *MICB*, *MST1*, and *NRG4*.

### Effect modification by diabetes status

Recognizing diabetes as a critical modulator of kidney disease and systemic pleiotropy, we further stratified our endpoint analyses by diabetes status (N =116,716 DM, N = 347,915 non-DM) (**Supplementary** Figure 9)^10^. Among the proteins significantly associated with kidney function in the main analysis, *UMOD, CLIC1, BTN2A1, AGER, MICB, C4A|C4B, BST1, INHBC* exhibited significant heterogeneity (*P*-value < 0.05/93) in their genetic associations with a kidney outcome.

### Mapping to kidney disease subtypes with GWAS meta-analyses and two-Sample MR of nine kidney disease subtypes

We re-tested the 93 PWAS proteins using two-sample cis-MR against 9 ICD-coded kidney phenotypes from MVP, UK Biobank, and FinnGen (Bonferroni P < 5.6×10^-3^) (**Figure 4**). 48/93 proteins (51.6%) were associated with ≥1 subtype. Total significant associations by subtype were: Hypertensive Kidney Disease 23, Acute Renal Failure 16, Type 2 DKD 12, Other nephropathies 12, Nephrotic Syndrome 9, Glomerulonephritis 6, Type 1 DKD 6, Glomerular disease, NOS 5, and Cystic Kidney Disease 4. 24 proteins were specific to a single category, including examples such as ADH1B, ANXA11 (AKI); PSG5 (Cystic); RCL (Nephrotic Syndrome); ATRAID (Other nephropathies); GCKR, LTB (Type 1 DKD); and GCDH, GSS, MANBA, NCAN (Type 2 DKD). Type 1 DKD overlapped most with glomerular categories (Glomerulonephritis 4/6 = 66.7%, Nephrotic Syndrome 4/6 = 66.7%, Other nephropathies 4/6 = 66.7%), whereas Type 2 DKD showed overlap with the hypertensive/acute kidney injury subtypes (Hypertensive 4/12 = 33.3%, AKI 3/12 = 25.0%, with ≤ 2 associations with glomerular categories). Hypertensive kidney disease, glomerular diseases and other nephropathies demonstrated overlap. If those categories were removed, then 12 additional proteins would show specific subtype associations including AKI (ANXA2, FAM213A, FGF5, GPX1, MST1), Cystic (A4GALT), nephrotic (AGER, CFHR1), Type 2 DKD (APOE, GSTA1, INHBC, PCK1).

**Figure 4.**
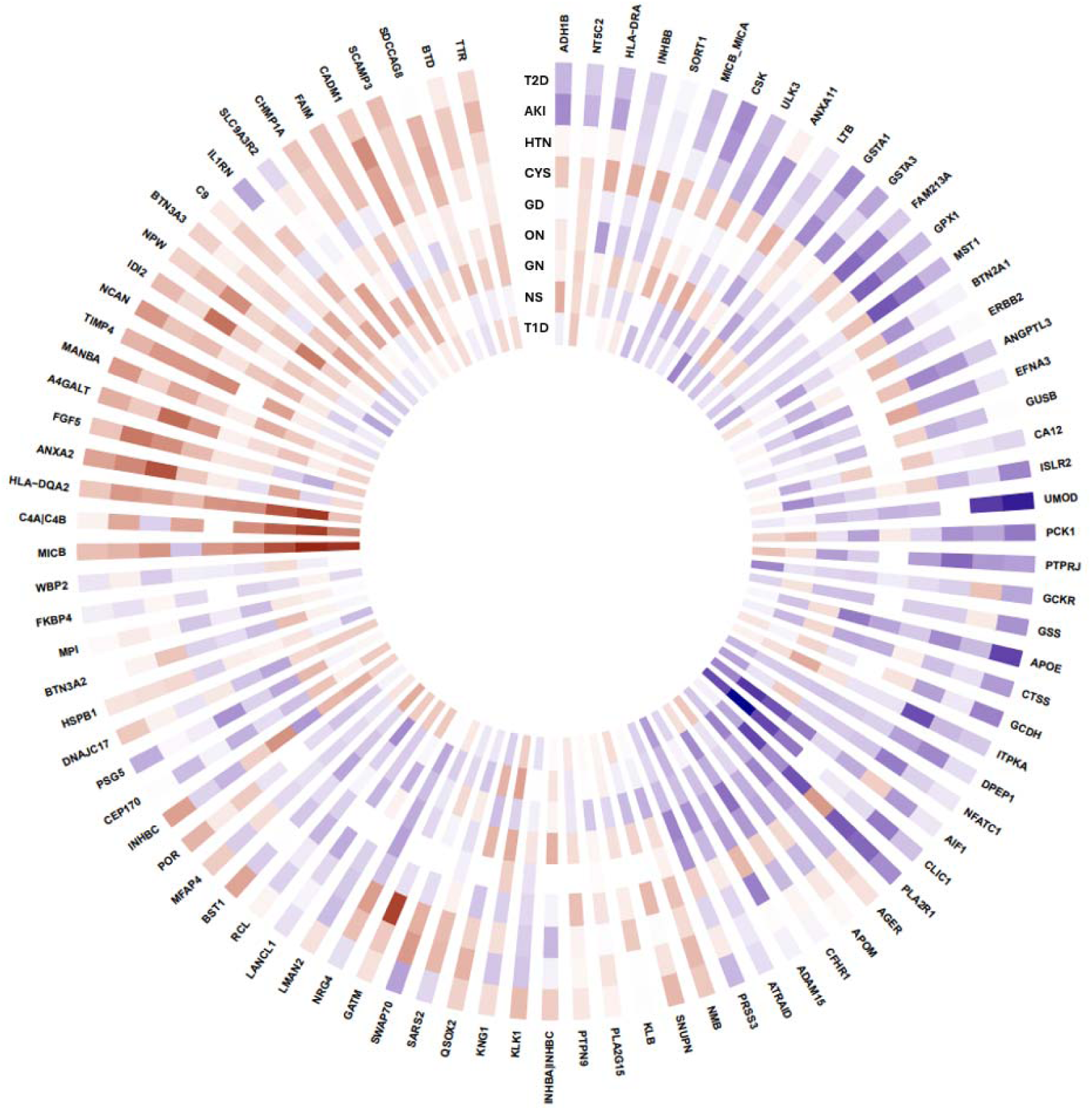
Heatmap of two-sample Mendelian randomization associations between 93 proteins from the proteome-wide analysis with 9 kidney disease subtypes. There were 93 plasma proteins that achieved statistical significance in our proteome-wide association study (outermost gene name labels). The heatmap summarizes the Mendelian-randomization associations between those proteins with 9 kidney disease subtypes based on GWAS meta-analysis utcomes from the MVP, UKB, and FinnGen. The blue indicates a positive beta association and the red indicates a negative beta association. The intensity of the color is proportional to the natural log compressed -log10(*P*-value). The HLA-A was removed from the heatmap because the markers were not present in the kidney summary statistic files. AKI—acute kidney injury, CYS—cystic kidney disease, GD—glomerular diseases, GN—glomerulonephritis, NS—Nephrotic syndrome, ON—Other nephropathies, T1D—Type 1 Diabetic Kidney Disease, T2D—Type 2 Diabetic Kidney disease, HTN—Hypertensive kidney disease. Full data presented in **Supplementary Table 10**.

### Assessment of systemic pleiotropy with PheWAS

Clinical phenotypes were defined using phecodes, which map International Classification of Diseases (ICD) codes to disease traits.^9^ PheWAS analysis was performed using an established methodology wherein logistic regression models adjusting for age, sex, and first 5 principal components were used to assess the association between each pQTL and 1020 phecodes across 11 clinical domains. To determine kidney specificity, we compiled the list of kidney-related phecodes as a separate 12^th^ kidney domain (**Figure 5, Supplementary Table 12**).

**Figure 5.**
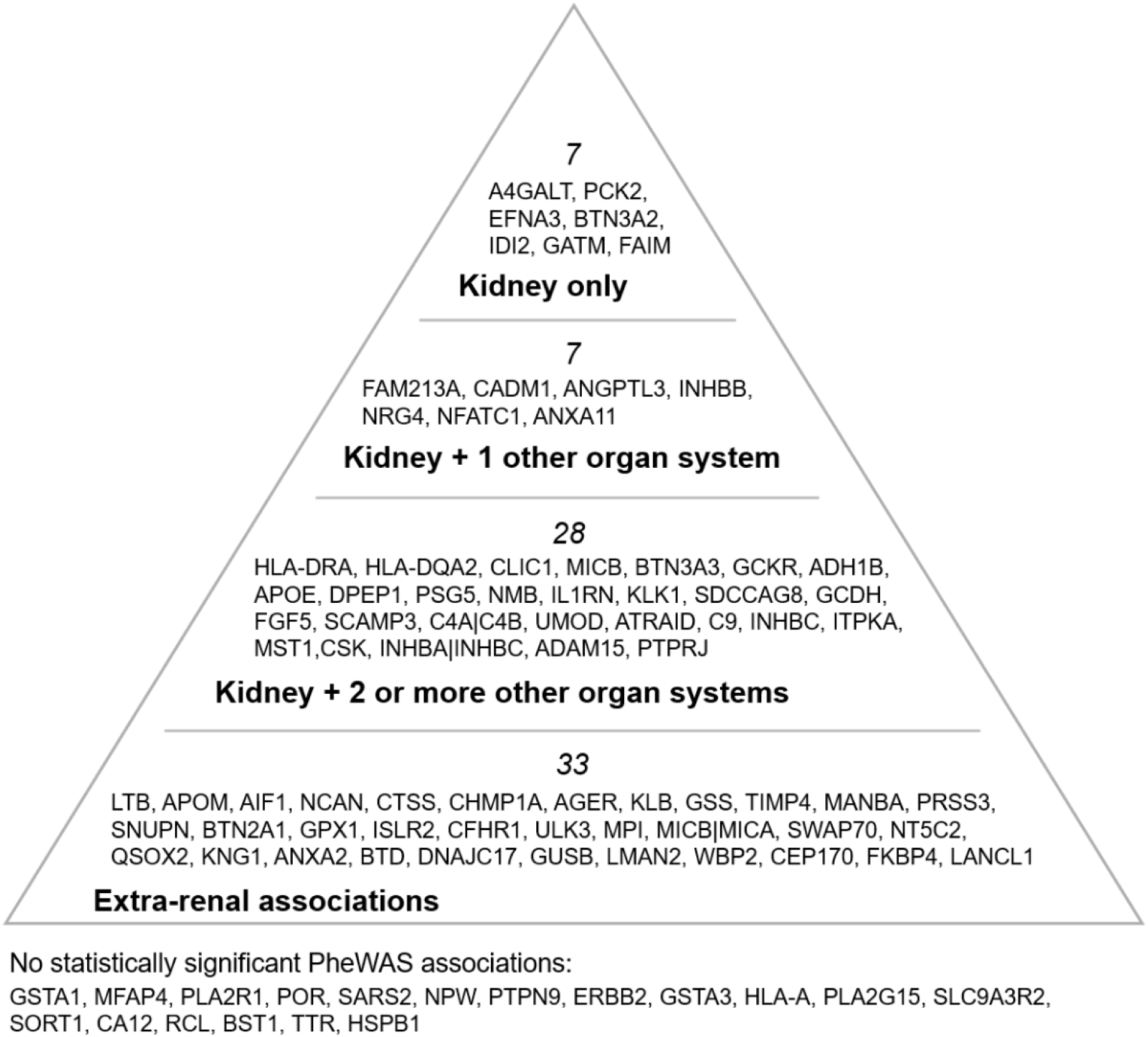
Hierarchical pyramid of organ-system pleiotropy revealed by phenome-wide association studies of 93 circulating protein QTLs. This pyramid summarizes results from phenome-wide association studies (PheWAS) in which each of 93 cis-pQTLs from the proteome-wide association study were tested via logistic-regression models adjusted for age, sex and the first ten principal components of ancestry against 1020 PheCodes from MVP. The apex lists eight proteins whose variants are exclusively associated with kidney PheCodes (listed in **Supplementary Table 12**). The next tiers contains proteins that show kidney associations plus signals in one or more other organ systems. There were 19 proteins that were under-powered for any significant PheWAS hit. Together, the figure depicts a spectrum from kidney-specific to highly pleiotropic proteins.

There were 71 proteins demonstrating statistically significant PheWAS associations (*P*-value < 4.9×10^-5^), with the most frequent protein-phecode associations found in the kidney domain (n ≈ 40), endocrine/metabolic domain (n ≈ 30), and circulatory domain (n ≈ 25) (**Supplementary Table 10**, **Supplementary** Figure 10). There were 7 proteins that exclusively associated only with the kidney domain supporting their proximal role in kidney disease (e.g., *A4GALT, PCK2, EFNA3, BTN3A2, IDI2, GATM, FAIM*). Beyond these, the majority of PheWAS associations were unique to individual proteins, reflecting substantial heterogeneity and pleiotropy (**Supplementary** Figure 11). Nineteen proteins did not yield any statistically significant PheWAS. This could either indicate kidney-specificity or small variant effect sizes that fall below PheWAS power.

### Protein-protein interaction and druggability assessments

To assess the therapeutic tractability of PWAS-identified proteins, we first queried the ChEMBL database to determine whether any were direct targets of preclinical, investigational, or approved drugs (**Supplementary Table 13**)^11,12^. We expanded the search using STRING Protein-Protein Interaction Networks to identify high-confidence interactors (interaction score > 0.80) for each primary PWAS protein and assessed whether these secondary interactor proteins were drug targets^13^. In total, the drug-protein-kidney disease networks comprised 259 unique drugs, 118 unique protein targets, and 367 unique drug-protein target pairs (**Supplementary** Figure 12). Of the 93 primary PWAS proteins, 34 (36.6%) had at least one pharmacologic entry point either directly or indirectly. Twelve (12.9%) were direct drug targets (*ADH1B*, *AGER*, *ANGPTL3*, *CA12*, *CTSS*, *DPEP1*, *ERBB2*, *HSPB1*, *INHBA*, *KLB*, *LTB*, *TTR*), with the remainder accessible through secondary protein interactors.

## Discussion

This integrative proteome-wide association study delivers a map of causal protein drivers and druggable pathways across the spectrum of CKD. The proteome-wide Mendelian randomization analysis identified circulating proteins that were associated with a comprehensive kidney outcome based on longitudinal electronic health records, and each protein is mapped to specific kidney disease subtypes and evaluated for systemic pleiotropy. This prioritizes potential therapeutic targets or drug safety signals for kidney diseases.

Overall, 12 of the 93 statistically significant proteins demonstrated robustness across all sensitivity analyses (*ANGPTL3*, *ADH1B*, *FAM213A*, *GCKR*, *GSTA1*, *IDI2*, *INHBA*, *INHBC*, *MANBA*, *MICB*, *MST1*, and *NRG4*). However, there was variability in their renal specificity. For example, circulating ADH1B was a robust finding in the main analysis and was associated with acute renal failure, yet showed extensive cardiometabolic associations, arguing against renal specificity^14,15^. In contrast, 7 proteins demonstrated only kidney-related associations, supporting their proximal kidney biology (*A4GALT, PCK2, EFNA3, BTN3A2, FAIM, IDI2*, and *GATM*). Some of these represent known biomarkers of kidney disease (*IDI2* as a robust indicator of kidney function in multiple prior studies^16,17^, *EFNA3* gene variants have been determinants of kidney function and albuminuria^6,18^, and *BTN3A2* is an immune-regulatory protein enriched in immune cells and genetically linked to CKD, including IgA nephropathy)^2^. Other renal-specific findings have biological plausibility. *A4GALT* synthesizes glycosphingolipid at the renal podocyte/endothelial interface, and is investigated as a target for Fabry disease^19,20^. *PCK2* underpins renal proximal tubule gluconeogenesis and is differentially expressed in proximal tubules in diabetic kidney disease^21^. *FAIM1,* an endogenous inhibitor of Fas-mediated apoptosis, is a novel finding but warrants further investigation given that preventing tubular cell apoptosis can limit renal injury and fibrosis^22,23^. Notably, *GATM*, the enzyme glycine amidinotransferase, catalyzes the first, rate-limiting step of creatine biosynthesis and may represent a biased creatinine-based eGFR without reflecting true changes in filtration.

Proteins with tractable drug targets converged on inflammatory and fibrotic biology (e.g., the “final common pathway” of CKD^24^). These domains were related to cytokines and immune signaling (*IL1RN, LTB, NFATC1, CSK, MST1, PSG5*), the complement system (*C4A, C4B, C9, CFHR1)*, TGF-B superfamily (*INHBA, INHBB, INHBC*), pattern-recognition (*AGER*), kallikrein-kinin and vascular inflammation (*KNG1*), stress/redox processes (*HSPB1*), and fibrotic signaling (*ERBB2*). Several networks recapitulated pathways already targeted by late-stage or approved CKD therapies, serving as internal positive controls (such as the inhibition of *C3*-related disease by pegcetacoplan, or the inhibition of the *PPP3CA-NFAT* axis by calcineurin inhibitors **Fig 6A**)^25–27^. We nominate other anti-inflammatory pathways, currently under development for non-renal indications, as promising candidates for CKD drug repurposing. One such pathway is the IL1- IL1RN signaling pathway, which shows an association with diabetic kidney disease in our study and is already therapeutically targeted in gout. Preclinical models demonstrating that IL1 blockade reverses or prevents albuminuria, inflammation, and fibrosis in diabetic and hypertensive kidney disease^28,29^. A randomized clinical trial using rilonacept in stage 3/4 CKD showed significant vascular function improvement, underscoring feasibility for future kidney- outcome trials^30^. Another promising axis is activin/inhibin signaling (*INHBA, INHBB, INHBC*). Preclinical studies show that trapping activin-receptor ligands reduce proteinuria and renal fibrosis in CKD models^31^. Although not kidney-approved, agents with related pharmacology, such as sotatercept (approved for pulmonary arterial hypertension but also studied in dialysis) and bimagrumab (e.g., an investigational dual activin receptor A/B antagonists that improves lean body mass), may warrant evaluation as anti-fibrotic strategy for CKD^32,33^.

**Figure 6.**
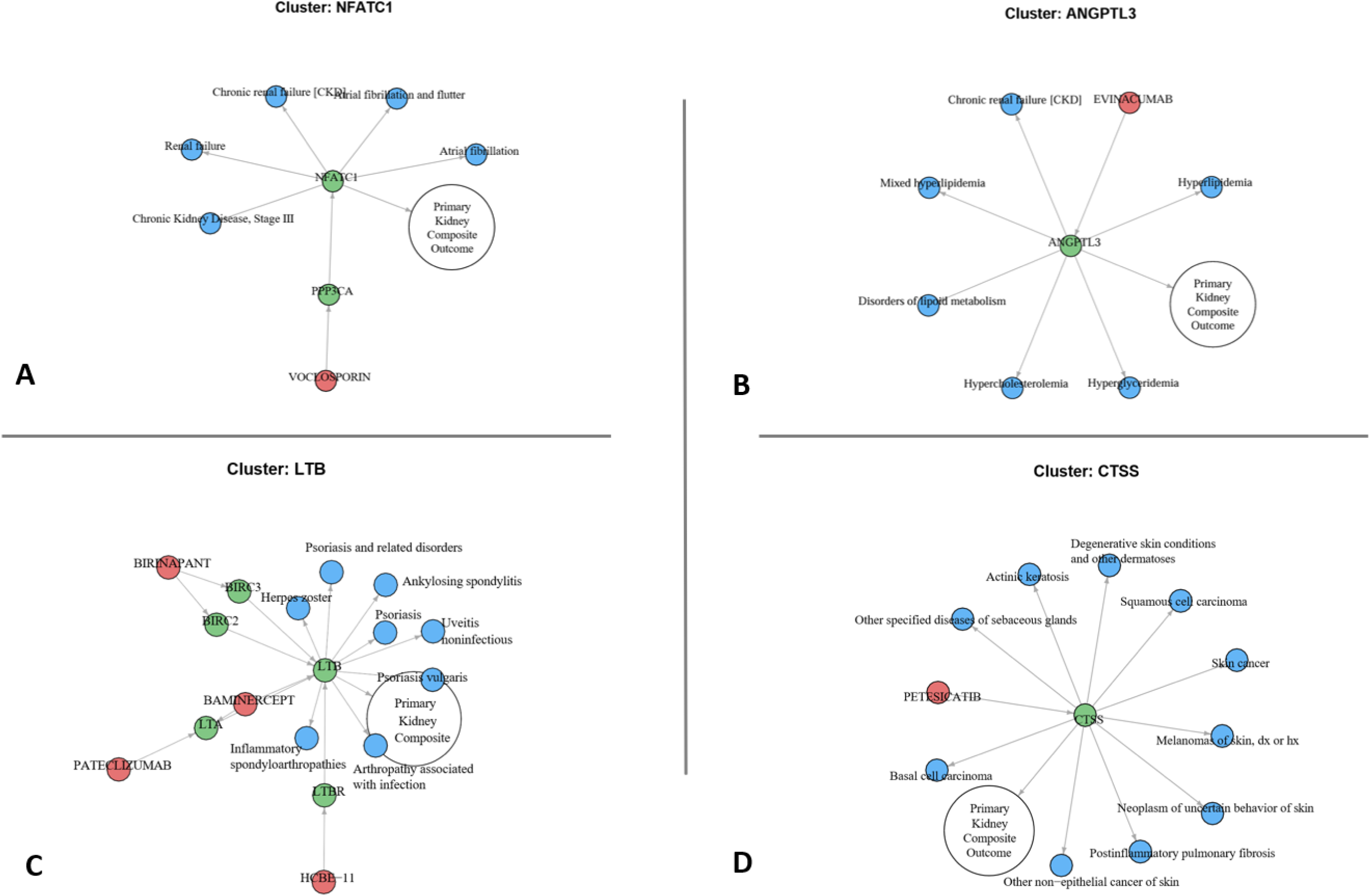
Drug-protein-kidney disease network vignettes, demonstrating potential drug repurposing and on-target safety liabilities. Each network is centered on a protein (green) linked to the study’s primary kidney outcome (white) and to phenome-wide associations (blue), with approved or investigational drugs (red) mapped to the protein or a closely related ligand. **(A)** The calcineurin axis (PPP3CA-NFATC1) is recovered as an internal positive control demonstrating a kidney protective mechanism for voclosporin. **(B)** ANGPTL3 shows convergent associations with dyslipidemia and CKD nominating its inhibition for renal indications beyond lipid lowering. **(C)** LTB and its receptor axis (LTBR-LTBR) show convergent associations with kidney diseases and autoimmune phenotypes (for example psoriasis, spondyloarthropathies, uveitis), consistent with prior studies indicating a kidney benefit in immune-complex nephritis. **(D)** Cathepsin S (CTSS) is linked to the kidney composite and has a clinical inhibitor, but the phenome context highlights multiple cutaneous neoplasms, consistent with prior studies indicating a kidney benefit with potential risk of cutaneous malignancy.

Proteins with shared renal and extra-renal associations could represent *on-target benefit* (shared biology where the same causal pathway drives phenotypes in multiple organs and thus might yield multiple therapeutic benefits). For example, Angiopoietin-like 3 (*ANGPTL3*) exemplifies shared biological pathways between dyslipidemia and CKD as reflected in our PheWAS findings (**Fig 6B**). *ANGPTL3* is a hepatokine that inhibits lipoprotein and endothelial lipases, and human loss-of-function and pharmacologic antagonism lower triglycerides and low-density lipoprotein and are associated with reduced coronary disease risk^34,35^. Beyond its role in lipid metabolism, *ANGPTL3* has direct effects on kidney structure and function. ANGPTL3 increases glomerular endothelial permeability and remodels podocyte actin, with overexpression causing proteinuria and podocyte injury, whereas its blockade is renoprotective in several models^36,37^. These observations nominate *ANGPTL3* inhibition beyond lipid lowering but also as a target to mitigate lipotoxic stress and podocyte injury in CKD. Another example of shared biology is Lymphotoxin beta (*LTB*), a membrane protein that is part of the TNF superfamily B^38^. In our study *LTB-LTBR* was associated with increased CKD, but was dominated in PheWAS by autoimmune associations such as psoriasis, inflammatory arthritis, and uveitis (**Fig 6C**). Indeed, *LTB-LTBR* signaling is up-regulated in human kidneys with inflammatory glomerulonephritis, and *LTBR* blockade in mouse lupus models improves kidney function^39^. The *LTB-LTBR* axis is known to be both pathogenic and drug-responsive across multiple autoimmune-affected organs (skin, joints, eyes), reinforcing its relevance to autoimmunity and highlighting its potential as a therapeutic target in immune-mediated CKD^40,41^.

In contrast we can identify potential *on-target safety liabilities* (safety trade-offs benefiting the kidneys but harming other organ systems). For example, cathepsin S (*CTSS*) emerged as a drug target with a benefit for kidneys but additional associations with cutaneous malignancy. *CTSS* facilitates MHC-II antigen presentation, and may promote microvascular dysfunction and amplify profibrotic signaling^42^. Selective *CTSS* inhibition in preclinical models reduces albuminuria, glomerulosclerosis, inflammation, and fibrotic remodeling^43^. Clinically, the *CTSS* inhibitor petesicatib has shown pharmacodynamic immunomodulation with acceptable short-term tolerability^44^ (**Fig 6D**). However, prior studies suggest that cathepsin blockade may modify anti-tumor immune surveillance, aligning with our PheWAS signals with cutaneous malignancy^45^. Although no malignancy signal has yet emerged in short-term trials, the convergence of mechanistic plausibility and genetic association supports proactive dermatologic monitoring in future *CTSS*-targeted therapies^46^.

The strengths of our design include a longitudinal, multi-endpoint kidney outcome that captures both disease onset and progression in electronic health records, providing a detailed and clinically-relevant kidney function outcomes. The use of multiple, large-scale proteomics platforms allows for cross-validation and the inclusion of multiple sensitivity analysis, including the replication of the main findings in an independent CKDGen cohort reinforce robustness. We leveraged high-throughput phenotyping projects (MVP, UK Biobank, FinnGen) to map significant findings with diagnosis code-based kidney subtypes. Phenome-wide testing separated proteins with broad systemic effects from those with kidney-restricted associations, placing protein signals in a broader clinical context^47^. Compared with prior gene- or variant-level scorecards that mainly weigh regulatory potential, our proteome-to-phenome framework separates kidney-specific signals from those with extra-renal or systemic effects, which helps with therapeutic prioritization and evaluation of benefits or safety liabilities^48^.

Several caveats remain. Standard MR assumptions apply. Despite the usage of cis-acting variants, colocalization, and multiple sensitivity analyses, pQTL instruments can still have residual horizontal pleiotropy, linkage disequilibrium with neighboring genes, and weak-instrument bias. Phenome-wide analyses may be underpowered for low-frequency variants and rely on ICD-based phecodes that aggregate heterogeneous entities (e.g., “nephrotic syndrome”), which could dilute the subtype-specific effects. In several instances the same instruments showed strong, reproducible associations across datasets but with opposite directions. This likely reflects differences in sampling along the disease course, population and comorbidities, and assay/platform behavior. Also, MR estimates lifelong, genetically proxied exposure whereas measured protein-kidney associations are dynamic. We therefore prioritize the magnitude and reproducibility of association and assign direction using convergent evidence (cross-endpoint consistency within a dataset, colocalization, replication, phenome-wide context, and prior biology). Genetic perturbations are not equivalent to pharmacology, so “druggability” inferences are hypothesis-generating. The discovery pQTL resources are predominantly European ancestry, limiting generalizability. Finally, current plasma assays incompletely capture low-abundance or kidney-restricted proteins. Future work should extend pQTL instruments to kidney-tissue platforms.

In conclusion, this proteome-to-phenome study integrated longitudinal electronic health record outcomes, kidney-subtype GWAS meta-analyses, and phenome-wide association studies to yield potentially causal, clinically-relevant protein drivers of CKD. We nominate 93 circulating proteins, many with subtype-specific effects, external replication and colocalization support, and potential pharmacologic targets. By examining the renal effects within systemic contexts, the framework helps to interpret on-target benefits and safety liabilities, helping to inform drug target selection, drug repurposing, and drug safety monitoring. Genetic evidence serves as motivation to advance clinical development of novel therapies for kidney diseases.

## Methods

### Overview of analysis

We implemented a multi-step analytic pipeline to prioritize pQTL instruments for their potential causal associations with kidney diseases (**Figure 1**). The main analysis was a proteome-wide Mendelian Randomization (MR) which assessed the associations between genetic variants associated with circulating proteins with a primary kidney outcome in the Million Veteran Program (MVP). The exposures were genetic variants influencing plasma protein levels leveraging proteomic data from four cohorts (ARIC, Fenland, deCODE, UK Biobank). The primary kidney outcome included four components: estimated glomerular filtration rate (eGFR, a continuous measure of kidney filtration), incident chronic kidney disease (CKD, defined as persistently reduced eGFR for ≥90 days), progression to end-stage kidney disease (ESKD, defined by dialysis initiation, kidney transplant, or sustained eGFR <15 mL/min/1.73 m² for ≥90 days), and annualized eGFR slope (the rate of kidney function decline modeled via linear mixed-effects regression). For each protein we conducted four separate two sample MR tests yielding four *P*-values. Because these four components are biologically related and not independent, ACAT combined four *P*-values into a single summary *P*-value in order to identify proteins with overall significance across any of the four components^49^. We performed subgroup analyses stratified by diabetes status and formally compared subgroup-specific genetic effect estimates using a z-test for heterogeneity to identify diabetes-dependent variation in associations. Significant findings from the proteome-wide MR analyses were subsequently subjected to further validation. We assessed the consistency of protein associations across the four kidney function endpoints, performed genetic colocalization analyses between each identified pQTL and the corresponding kidney function loci to evaluate shared genetic architecture, and sought replication of these findings in an independent external dataset from the CKDGen consortium. We performed targeted MR analyses of these protein-associated variants against predefined kidney disease-specific outcomes, by combining GWAS meta-analyses for 9 kidney disease subtypes from MVP, UK Biobank, and FinnGen (e.g., diabetic kidney disease, hypertensive kidney disease, glomerular diseases, glomerulonephritis, nephrotic syndrome, cystic kidney disease, acute renal failure). Finally, we performed PheWAS of protein-associated variants across the MVP. Drugs targeting these proteins using ChEMBL resource, or proteins closely related biologically in protein-protein interaction networks based on STRING, were interpreted within this context.

### Study Population

The VA Million Veteran Program (MVP) is a national cohort launched in 2011 designed to study the contributions of genetics, lifestyle, and military exposures to health and disease among US Veterans^50^. Blood biospecimens were collected for DNA isolation and genotyping, and the biorepository was linked with the VA electronic health record (EHR), which includes diagnosis codes (International Classification of Diseases ninth revision [ICD-9] and tenth revision [ICD- 10]), laboratory measures, and detailed survey questionnaires collected at the time of enrollment for all Veterans followed in the healthcare system up to September 2019^51,52^.

Specimen collection and genotype quality control have been described in detail elsewhere. In brief, blood specimens were collected at recruitment sites across the country and then shipped within 24 hours to the VA Central Biorepository in Boston, MA for processing and storage. Study participants were genotyped using a customized Affymetrix Axiom biobank array (the MVP 1.0 Genotyping Array), containing over 730,000 variants^53^. Duplicate samples were excluded as well as samples with observed heterozygosity greater than the expected heterozygosity, missing genotype call rate greater than 2.5%, or incongruence between sex inferred from genetic information and gender extracted from phenotype data. The harmonized race/ethnicity and genetic ancestry (HARE) approach, developed and validated by MVP, was used to assign individuals to ancestral groups^54^. This machine learning algorithm leverages information from both the self-reported race/ethnicity data from the MVP Baseline survey and genotype data to categorized Veterans into mutually exclusive groups.

### Clinical Covariates

Ambulatory medications were collected using outpatient pharmacy files. VA laboratory data, vital status files, and administrative diagnosis and procedure codes were collected. Diagnoses were defined using the International Classification of Diseases (ICD) versions 9 (ICD-9) and 10 (ICD- 10). Procedures were defined using ICD-9 and ICD-10 Procedure and Current Procedural Terminology (CPT) codes. Estimated glomerular filtration rate (eGFR) was calculated using the Chronic Kidney Disease Epidemiology Collaboration (CKD-EPI) Equation. Hematuria and proteinuria were defined using urine analysis (dipstick) data. Proteinuria was defined using urine dipstick measurement, with normal defined as a urine dipstick reading of negative or trace and positive defined as a urine dipstick reading as ≥ 1+. We did not include microalbuminuria due to missingness in 50% of the cohort. Diabetes was defined as requiring both 1) single instance of diabetes ICD code at a face-to-face PCP outpatient visit or the use of two diabetes ICD code on two different days and 2) outpatient prescription of a diabetes drug at any time.

### Exposures

In total, the analysis included 2,807 unique plasma protein exposures derived from four large-scale cohorts: the Atherosclerosis Risk in Communities (ARIC) study, the Fenland study (Fenland), the deCODE study (deCODE), and the UK Biobank (UKB). Within each proteomic dataset, assays were mapped to a UniProt identifier and HGNC gene symbol: ARIC = 1508, Fenland = 1489, deCODE = 1659, UKBB = 1690. We selected pQTLs located within cis-regions defined as genomic regions within ± 500 kilobases (kb) of the transcription start sites of their respective target genes to enrich for direct genetic effects on nearby gene expression. All analyses included variants with a minor allele frequency (MAF) greater than 1%. The ARIC study is an ongoing, community-based cohort initiated between 1987 and 1989, enrolling 15,792 individuals from four geographic regions across the United States. pQTL analyses included 7,213 participants and targeted 4,657 proteins measured from samples collected during the cohort’s third visit on the SomaScan v4 platform^55^. The Fenland study is a population-based cohort comprising generally healthy individuals residing in Cambridgeshire, United Kingdom^56^. pQTLs were derived from a genome-proteome-wide scan of 10,708 unrelated participants of European ancestry with 4,979 aptamers targeting 4,775 distinct proteins with on the SomaScan v4 platform.^57^ The deCODE cohort consists of plasma samples from Icelandic participants collected between August 2000 and January 2019 through the population-based deCODE Health study and other genetic research initiatives^58^. pQTLs from the deCODE Icelandic cohort were generated in 35,559 genotyped Icelanders whose protein abundances were quantified with 4,907 assays targeting 4,719 unique proteins after quality control using SomaScan v4. The UKB is a prospective cohort that recruited approximately 500,000 individuals aged 40 to 69 years across the United Kingdom from 2006 to 2010.^59^ Relative abundances of 2,923 plasma proteins were measured in approximately 54,000 participants with the Olink Explore 3072 proximity-extension assay as part of the UK Biobank Pharma Proteomics Project^60^.

### Outcomes

#### Primary kidney outcome

We conducted a multi-trait kidney function analysis, combining association *P*-values for eGFR, incident CKD, ESKD, and eGFR slope with Aggregated Cauchy Association Testing (ACAT) to obtain gene-level significance across this correlated outcome set. Estimated glomerular filtration rate (eGFR) is a continuous measure reflecting overall kidney filtration function calculated using serum creatinine using the race-free Chronic Kidney Disease Epidemiology Collaboration (CKD-EPI) creatinine equation^61^. Incident chronic kidney disease (CKD) was defined as a sustained reduction in eGFR below 60 mL/min/1.73 m² persisting for ≥90 days, identified from repeated outpatient creatinine-based measurements, a validated definition by guidelines^62,63^. Progression to end-stage kidney disease (ESKD) was defined by initiation of dialysis, kidney transplantation, or persistently low eGFR <15 mL/min/1.73 m² for ≥90 consecutive days, based on specific dialysis and transplantation procedure codes, as previously described. Annualized eGFR slope was calculated using linear mixed-effects models to estimate each participant’s rate of kidney function decline over time, as previously described^64^.

#### Genome-wide association study for the primary kidney outcome

We conducted four independent GWAS (one each for baseline eGFR, annualized eGFR slope, incident CKD, and incident ESKD) within the MVP and further repeated these GWASes stratified by diabetes mellitus (DM) status. All models included age at blood draw, sex, and the first ten ancestry-specific principal components (PCs). Baseline eGFR (first outpatient creatinine after cohort entry) and eGFR slope were analyzed with linear regression, while incident CKD and ESKD were analyzed with logistic regression. CKD cases required two eGFR values < 60 mL/min/1.73 m2 at least 90 days apart. ESKD cases required chronic dialysis, kidney transplantation, or two eGFR values < 15 mL/min/1.73m2 ≥90 days apart, identified with ICD-9- CM, ICD-10-CM, ICD-9-Procedure, ICD-10-PCs, and CPT codes. Controls were event-free at censoring and had ≥2 outpatient encounters during follow-up.

### Statistical Analyses

#### Main analysis: a proteome-wide Mendelian randomization study

We used a proteome-wide Mendelian randomization (MR) framework^65^. pQTL instruments from ARIC, Fenland, deCODE, and UKB were aligned to the kidney outcome GWASes (baseline eGFR, eGFR slope, incident CKD or ESKD) using effect/other alleles. If a variant was absent, we imputed missing allele frequencies from Ensembl biomaRt substituting a proxy r² ≥ 0.9 in 1000 Genomes EUR population and with similar MAF% ∼2%). Two-sample MR was performed in R: inverse-variance-weighted (IVW, multiplicative random-effects) models were fitted when ≥2

SNPs were available, otherwise a Wald ratio was used. Associations were performed at the protein level based on the UniProt accession ID and HGNC gene symbol of each protein exposure. We applied a Bonferroni threshold of P < 0.05 / n for each dataset where n is the number of proteins tested within the dataset.

#### Heterogeneity and pleiotropy testing

Instrument validity was evaluated with Cochran’s Q (heterogeneity P ≥ 0.05) and MR-Egger intercept (pleiotropy P ≥ 0.05). Instruments failing either test were flagged and their MR estimates interpreted cautiously.

#### Endpoint consistency/assessment of directionality

To evaluate the robustness of associations between circulating proteins and kidney health, we examined the directionality of Mendelian randomization (MR) effect estimates for each protein across the four biologically related components of the kidney outcome: baseline eGFR, eGFR decline (slope), incident CKD, and incident ESKD. Directional consistency was assessed for all 93 proteins that met the proteome-wide significance threshold in the primary analysis). For this sensitivity analysis, we applied a Bonferroni-corrected of *P*-value < 0.0125 for each protein-outcome pair threshold to account for the four clinical endpoints.

#### Genetic colocalization

We conducted genetic colocalization analyses to determine whether proteins identified from the main analysis share causal genetic variants with kidney function traits derived from the kidney outcome GWASes. For each significant pQTL identified from the main analysis, we extracted genetic variants within a ± 500 kb region around the sentinel SNP. Variants were harmonized between pQTL and GWAS datasets by matching chromosome positions, reference and alternate alleles, and minor allele frequencies. Bayesian colocalization analyses were implemented using the *coloc* R package, employing default prior probabilities (p1 = 1×10^-4^, p1 = 1×10^-4^, p_12_ = 1×10^-5^)^66^. Colocalization was confirmed if the posterior probability of a shared causal variant (PP4) exceeded 0.8.

#### External replication

We attempted to replicate all proteins that met the experiment-wide significance threshold in the MVP proteome-wide MR. Replication analyses were performed in the Chronic Kidney Disease Genetics Consortium (CKDGen), an international consortium established to identify genetic loci associated with kidney function and disease through large-scale meta-analyses of GWAS^67^. Summary statistics were used for cross-sectional eGFR (meta-analysis, n > 1 million) and CKD (European ancestry meta-analysis, n = 480,698; 41,395 cases). Additionally, longitudinal kidney function decline outcomes utilized GWAS results for “Rapid3” (eGFR decline ≥3 mL/min/1.73m²/year; 34,874 cases, 107,090 controls) and “CKDi25” (incident CKD defined as ≥25% decline in eGFR with follow-up eGFR <60 mL/min/1.73m2; 19,901 cases, 175,244 controls).^68^ Within each protein we combined the four single-outcome MR *P*-values using the Aggregate Cauchy Association Test (ACAT) analogous to the MVP PWAS. Replication was declared when the ACAT *P*-value met a Bonferroni-corrected *P*-valueD<D0.05 / n, where n is total number of proteins attempting replication.

#### Effect modification of diabetes mellitus

We further assessed whether diabetes mellitus (DM) status modified the effect of statistically significant proteins from the main analysis. We repeated this MR in the DM and the non-DM subgroups separately for each protein-kidney outcome pair and used a statistical test of heterogeneity (Z-test), calculated as the difference in subgroup-specific MR estimates divided by the standard error of that difference.

#### Kidney disease-specific analysis

We performed targeted MR using significant proteins from the main analysis as the exposures and a set of 9 specific kidney disease subtypes as the outcomes: diabetic kidney disease (type 1 and type 2), hypertensive kidney disease, glomerular disease, glomerulonephritis, nephrotic syndrome, cystic kidney disease, and acute renal failure. These ICD-code based outcomes were derived from meta-analyses of GWAS summary statistics from the Million Veteran Program (MVP), FinnGen^69^, and UK Biobank (UKB)^59,70^ generated within the MVP Data Core. The MVP GWASes used generalized linear mixed models to account for participant relatedness using a GPU-optimized version of the SAIGE package implemented on the U.S. Department of Energy Summit supercomputer^11^. The same MR approach (IVW or Wald ratio) was employed. Definitions of each kidney subtype definition, cases, and controls are provided in **Supplementary Table 9**. A Bonferroni threshold was applied to account for 9 kidney disease subtypes of *P*-value < 5.6×10^-3^.

#### Phenome-wide association studies (PheWAS)

We assessed clinical pleiotropy for protein-instrumenting variants by conducting a PheWAS across the MVP. For each of the sentinel proteins that met proteome-wide significance in the proteome-wide Mendelian-randomization screen, we extracted every conditionally independent cis-pQTL from the discovery cohorts. Each SNP was then tested against 931 electronic-health-record derived phenotypes (phecodes) across 11 domains that were curated by the MVP Data Core^9^. Each phecode represents ICD codes grouped into clinically relevant phenotypes for clinical studies ^10^. For each phecode, participants with ≥2 phecode-mapped ICD-9 or ICD-10 codes were defined as cases, whereas those with no instance of a phecode-mapped ICD-9 or ICD-10 code were defined as controls. Populations where the phecode comprises < 200 cases or controls were more likely to result in unstable results and were excluded. Associations were modeled with additive logistic regression. All models were adjusted for age at encounter, sex, and the first ten ancestry principal components to control for population stratification. Results were corrected for multiple testing with the Bonferroni adjustment across the full phenome (*P*- value < 0.05 / 1020).

#### Protein-protein interaction context and druggability screen

We queried the human STRING knowledgebase (v 12.5) through the STRINGdb Bioconductor interface (v 2.8.4).^13^ For each lead protein that was significant with our primary kidney outcome we obtained all first-shell interactors with a combined confidence score ≥ 0.9 (equivalent to 900 in STRING), and yielded a non-redundant, high-confidence PPI network. Druggability was evaluated by overlaying this network with ChEMBL annotations to flag proteins that are already direct drug targets and proteins whose immediate interactors are druggable^11,12^. Finally, we assembled integrated networks linking drugs, proteins, the primary kidney outcome, and PheWAS results to visualize the relationships.

## Acknowledgements

We are grateful to the MVP study participants for their contributions to science and all the enrolling sites listed in the Core Acknowledgement (**Appendix**) and the MVP staff and MVP program office. J.L.T. and A.M.H. had full access to all the data in the study and assumed responsibility for data integrity. We are also grateful to Vanderbilt University Medical Center for their support to our staff and faculty for their research endeavor.

## Disclaimer

This publication does not represent the views of the Department of Veteran Affairs or the United States Government. This research is based on data from the Million Veteran Program, Office of Research and Development, Veterans Health Administration.

## Data Availability

The VA MVP individual level data will not be shared through dbGAP; the Office of Research and Development of the Department of Veterans Affairs needs to be contacted for access requests for that data.

