## Supplemental Figures for "Integrative Proteome- and Phenome-Wide Assessment Uncovers Causal Protein Drivers and Drug Targets for Heterogeneous Kidney Diseases"

**Supplementary Figures**

**Supplementary Figure 1.** Proteome‑wide Mendelian randomization of ARIC proteins and kidney outcomes.

**Supplementary Figure 2.** Proteome‑wide Mendelian randomization of Fenland proteins and kidney outcomes.

**Supplementary Figure 3.** Proteome‑wide Mendelian randomization of deCODE proteins and kidney outcomes.

**Supplementary Figure 4.** Proteome‑wide Mendelian randomization of UK Biobank proteins and kidney outcomes.

**Supplementary Figure 5.** Forest plots of ARIC cis‑pQTLs that reached study‑wide significance and their association with the four MVP kidney outcomes.

**Supplementary Figure 6.** Forest plots of Fenland cis‑pQTLs that reached study‑wide significance and their association with the four MVP kidney outcomes.

**Supplementary Figure 7.** Forest plots of deCODE cis‑pQTLs that reached study‑wide significance and their association with the four MVP kidney outcomes.

**Supplementary Figure 8.** Forest plots of UK Biobank cis‑pQTLs that reached study‑wide significance and their association with the four MVP kidney outcomes.

**Supplementary Figure 9.** Scatter plot illustrates diabetes-dependent variability in genetic associations between plasma proteins and the four components of kidney outcome.

**Supplementary Figure 10.** UpSet plot of phenome‑wide protein associations across clinical domains.

**Supplementary Figure 11.** PheWAS plots for significant proteins from the proteome-wide analysis.

**Supplementary Figure 12.** Drug-protein-kidney disease network plots.

**Appendex 1.** VA Million Veteran Program: Core Acknowledgements for Publications.

**Supplementary Figure 1. Proteome‑wide Mendelian randomization of ARIC proteins and kidney outcomes.** Volcano plots display the Mendelian randomisation estimates for plasma proteins in ARIC against four kidney phenotypes adjudicated in the Million Veteran Program (MVP): baseline estimated glomerular filtration rate (eGFR), chronic kidney disease (CKD), end‑stage kidney disease (ESKD), and annualized eGFR slope. The x‑axis shows the MR β‑coefficient the y‑axis shows the -log10 P-value. Points colored by effect direction (red =increased protein associated with lower eGFR, CKD, ESKD, Slope, blue = increased protein associated with higher eGFR, CKD, ESKD, Slope). Genes surpassing the Bonferroni threshold (horizontal grey line) are labelled.
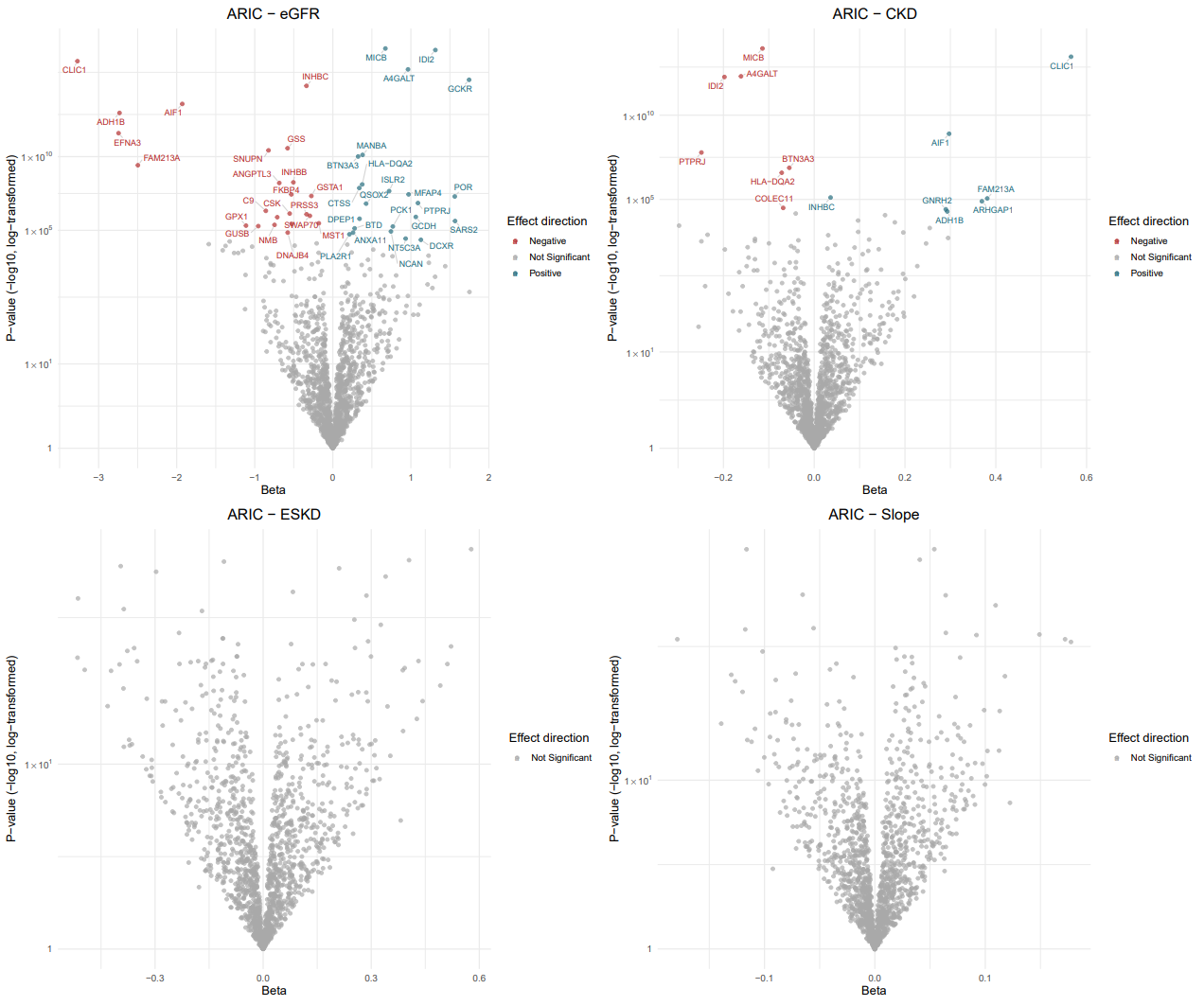

**Supplementary Figure 2. Proteome‑wide Mendelian randomization of Fenland proteins and kidney outcomes.** Volcano plots display the Mendelian randomisation estimates for plasma proteins in Fenland against four kidney phenotypes adjudicated in the Million Veteran Program (MVP): baseline estimated glomerular filtration rate (eGFR), chronic kidney disease (CKD), end‑stage kidney disease (ESKD), and annualized eGFR slope. The x‑axis shows the MR β‑coefficient the y‑axis shows the -log10 P-value. Points colored by effect direction (red =increased protein associated with lower eGFR, CKD, ESKD, Slope; blue = increased protein associated with higher eGFR, CKD, ESKD, Slope).

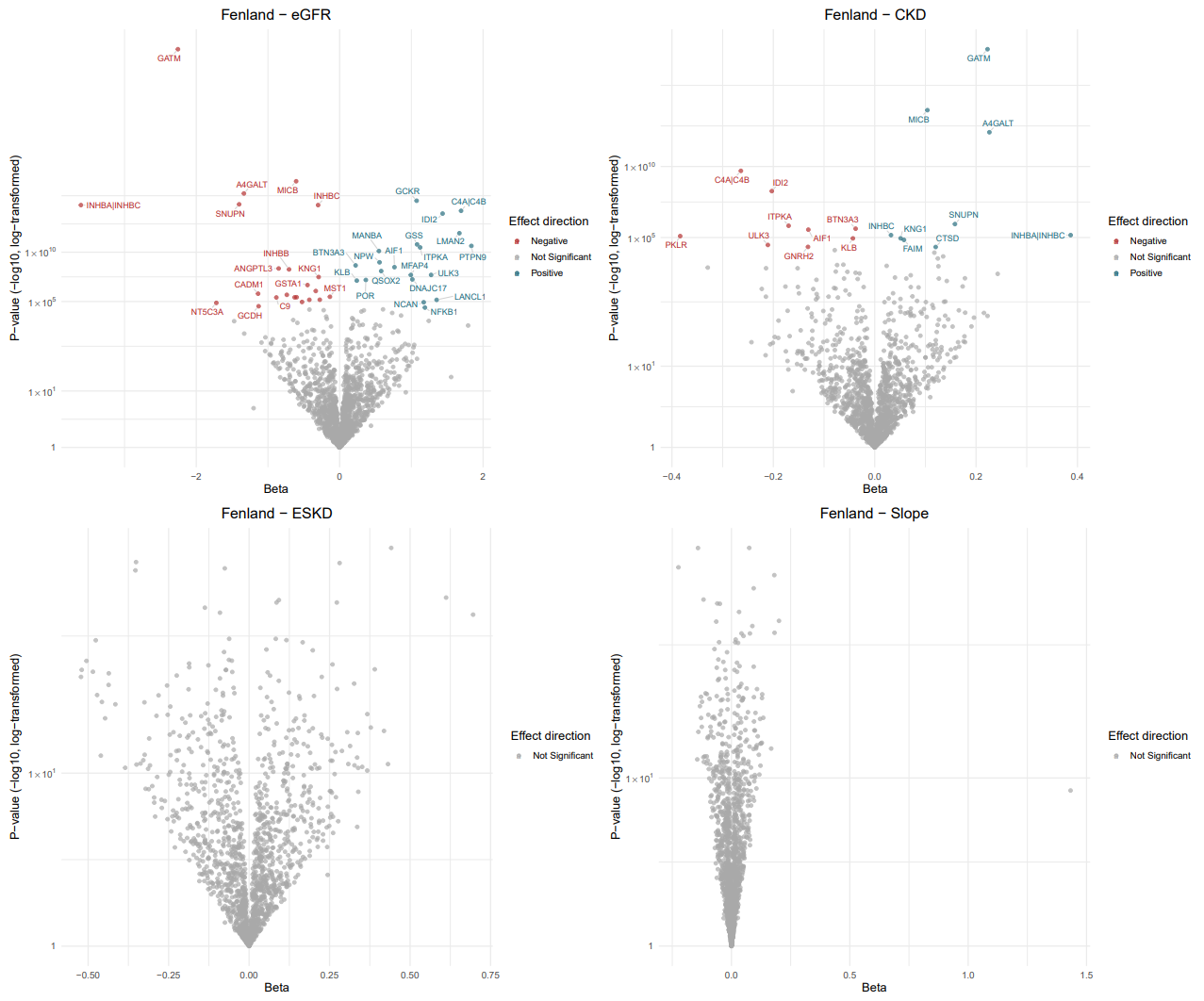

**Supplementary Figure 3 Proteome‑wide Mendelian randomization of deCODE proteins and kidney outcomes.** Volcano plots display the Mendelian randomisation estimates for plasma proteins in deCODE against four kidney phenotypes adjudicated in the Million Veteran Program (MVP): baseline estimated glomerular filtration rate (eGFR), chronic kidney disease (CKD), end‑stage kidney disease (ESKD), and annualized eGFR slope. The x‑axis shows the MR β‑coefficient the y‑axis shows the -log10 P-value. Points colored by effect direction (red =increased protein associated with lower eGFR, CKD, ESKD, Slope, blue = increased protein associated with higher eGFR, CKD, ESKD, Slope).

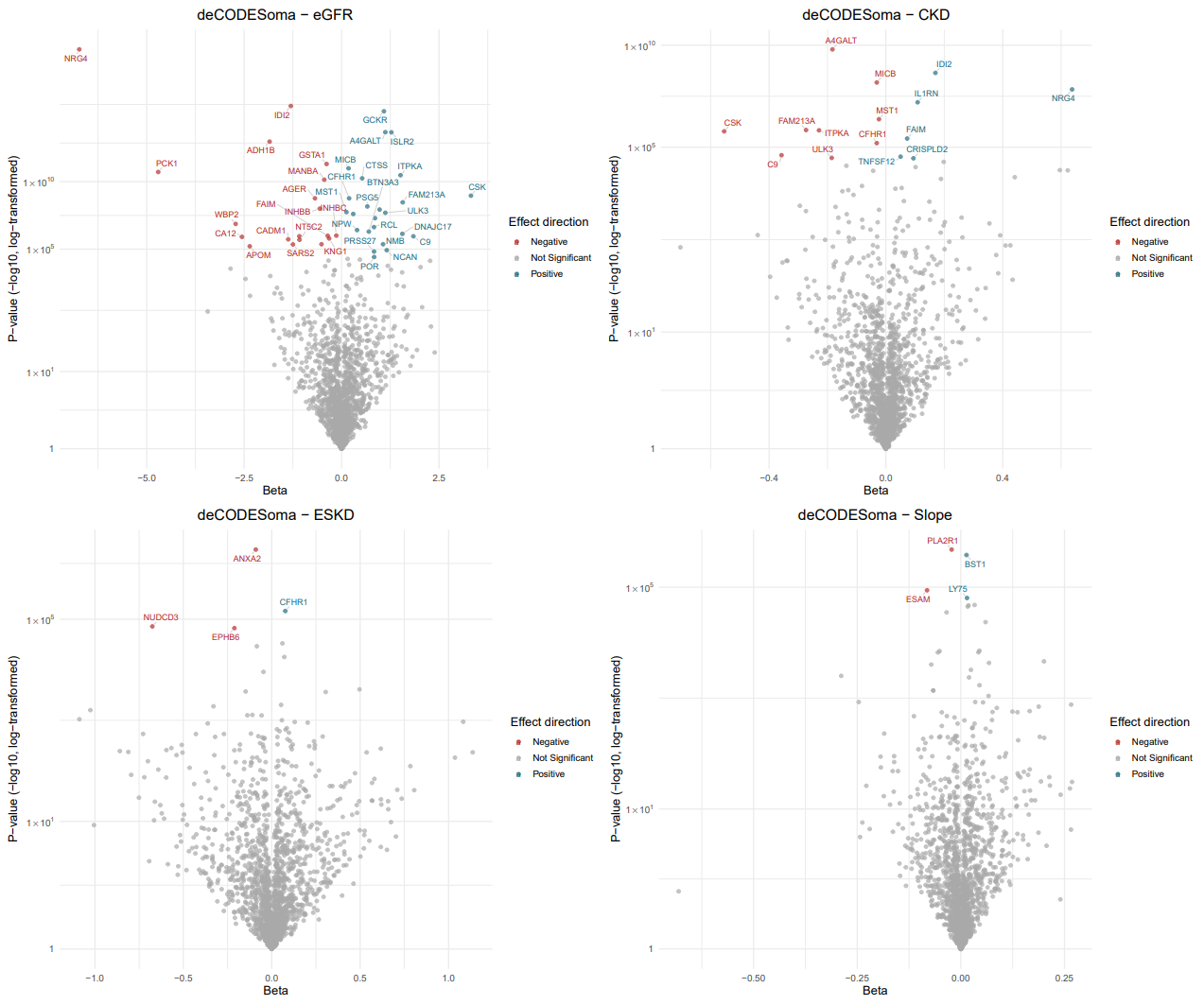

**Supplementary Figure 4. Proteome‑wide Mendelian randomization of UK Biobank proteins and kidney outcomes.** Volcano plots display the Mendelian randomisation estimates for plasma proteins in UK Biobank against four kidney phenotypes adjudicated in the Million Veteran Program (MVP): baseline estimated glomerular filtration rate (eGFR), chronic kidney disease (CKD), end‑stage kidney disease (ESKD), and annualized eGFR slope. The x‑axis shows the MR β‑coefficient the y‑axis shows the -log10 P-value. Points colored by effect direction (red =increased protein associated with lower eGFR, CKD, ESKD, Slope, blue = increased protein associated with higher eGFR, CKD, ESKD, Slope).

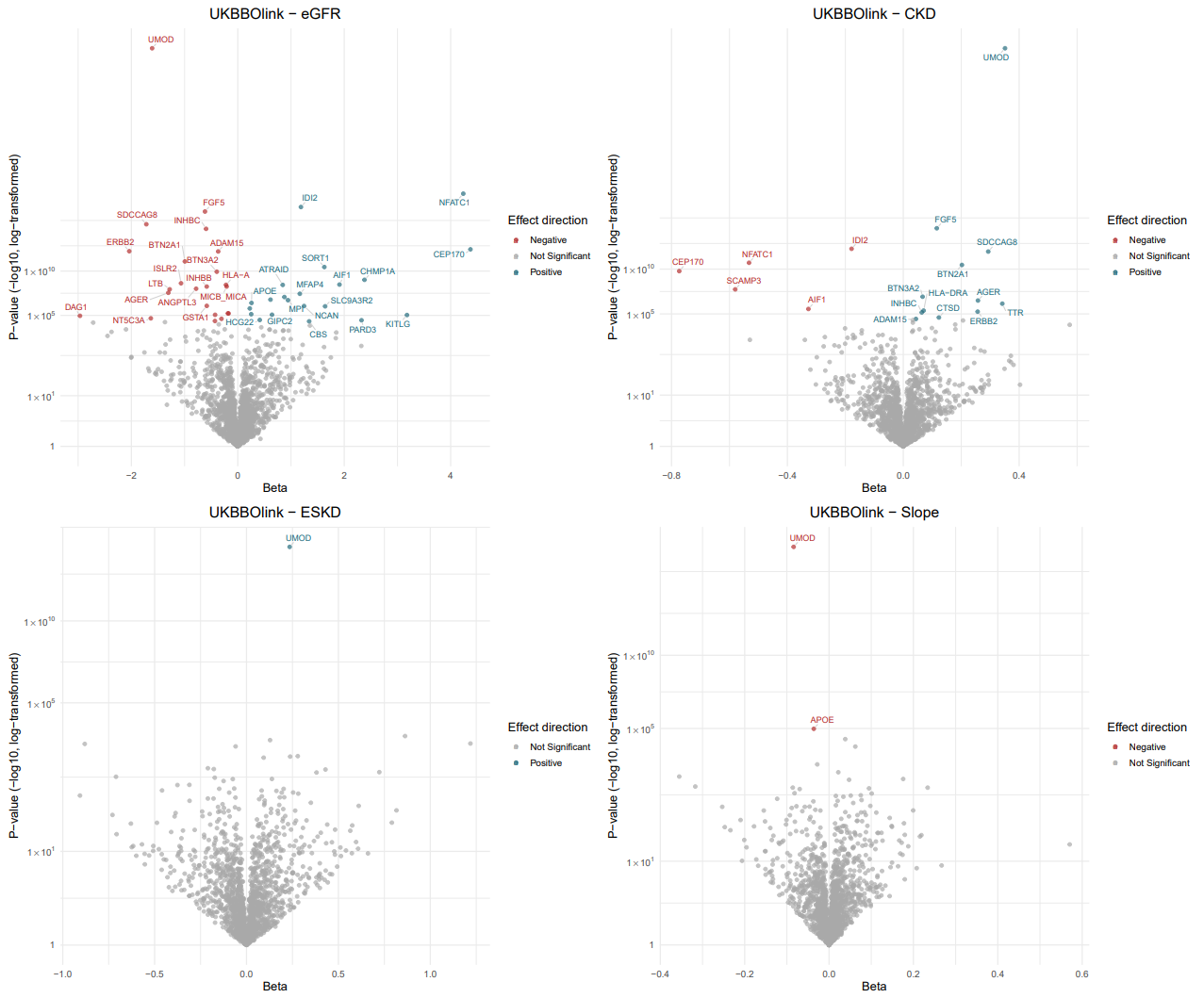

**Supplementary Figure 5. Forest plots of ARIC cis‑pQTLs that reached study‑wide significance and their association with the four MVP kidney outcomes.**

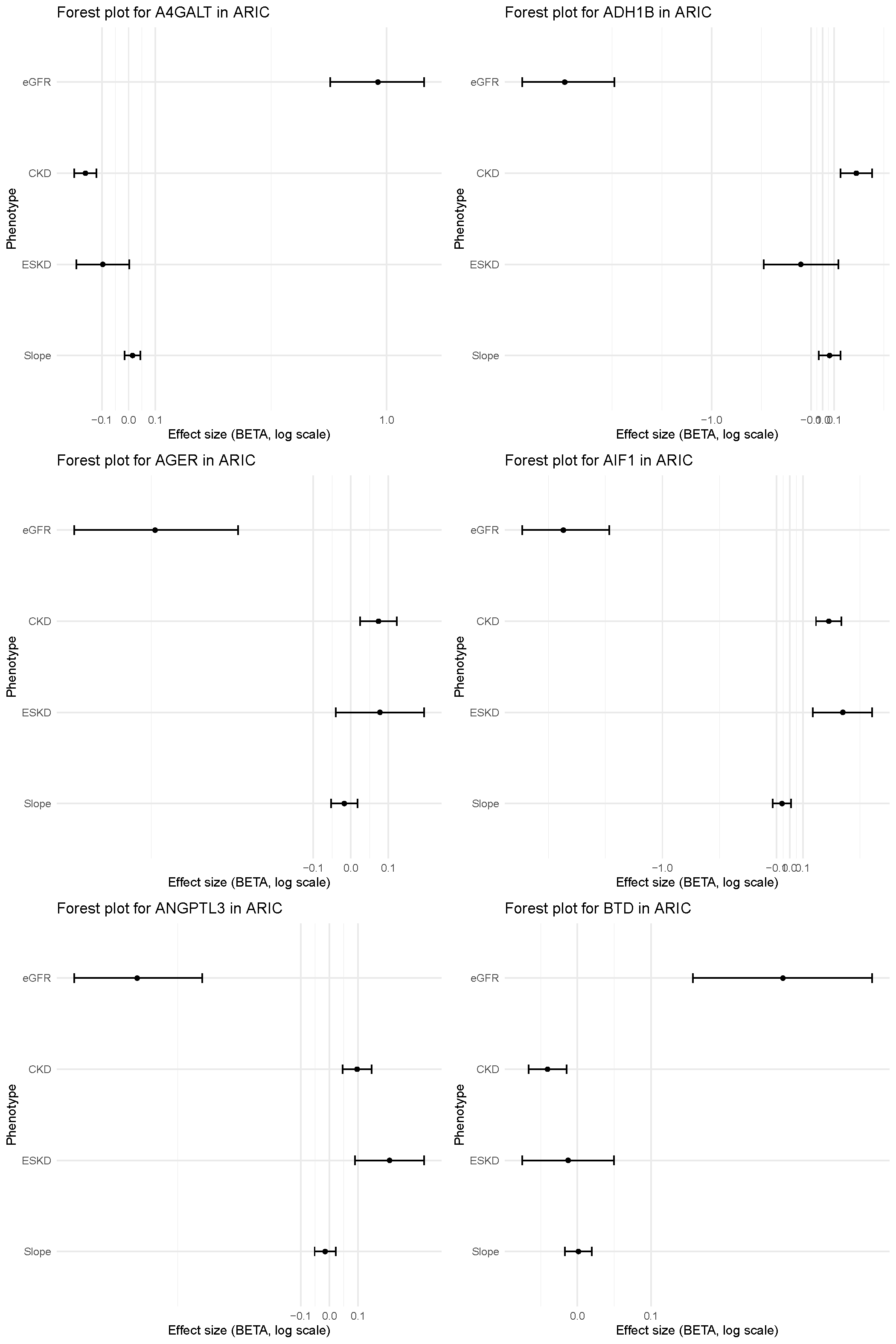

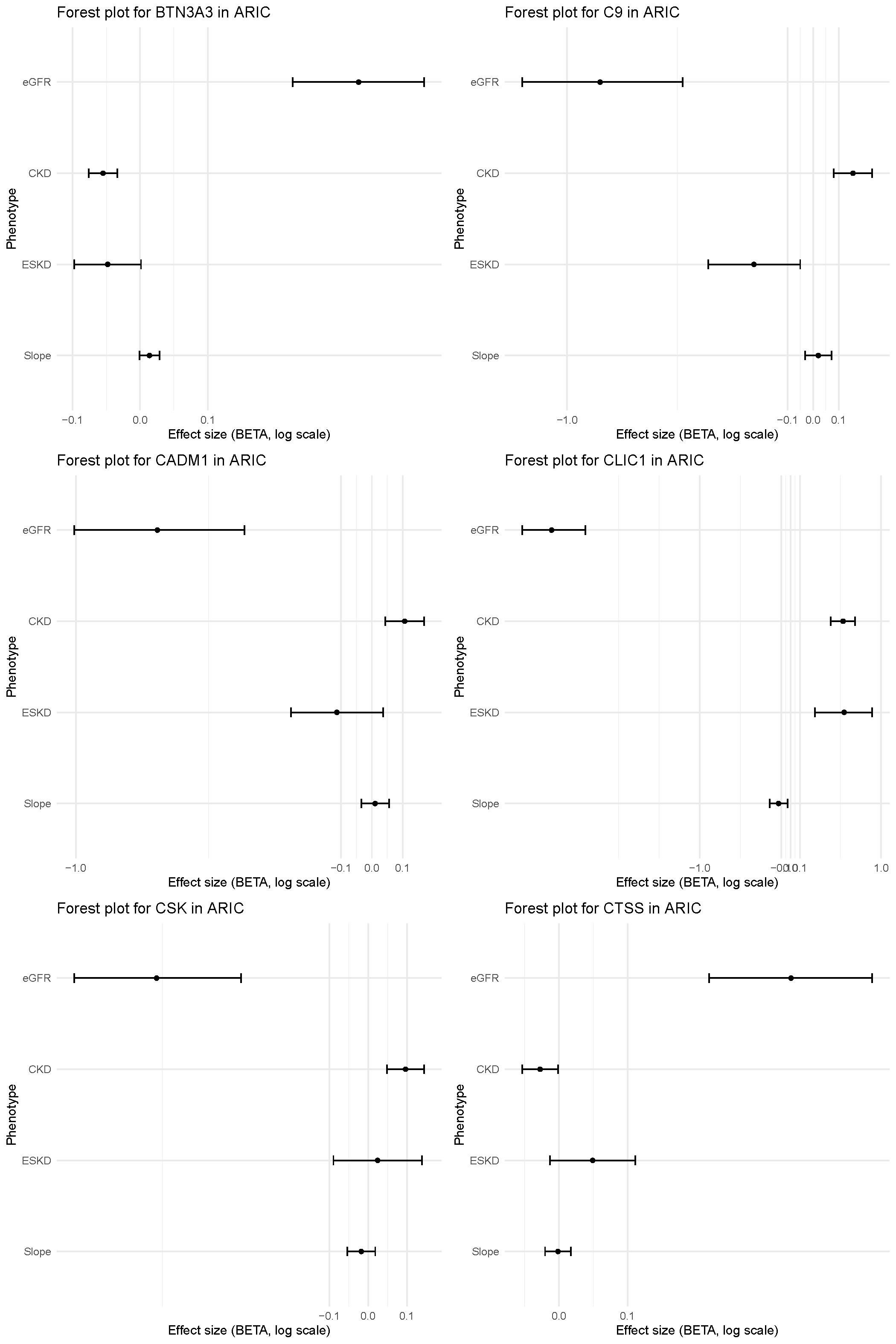

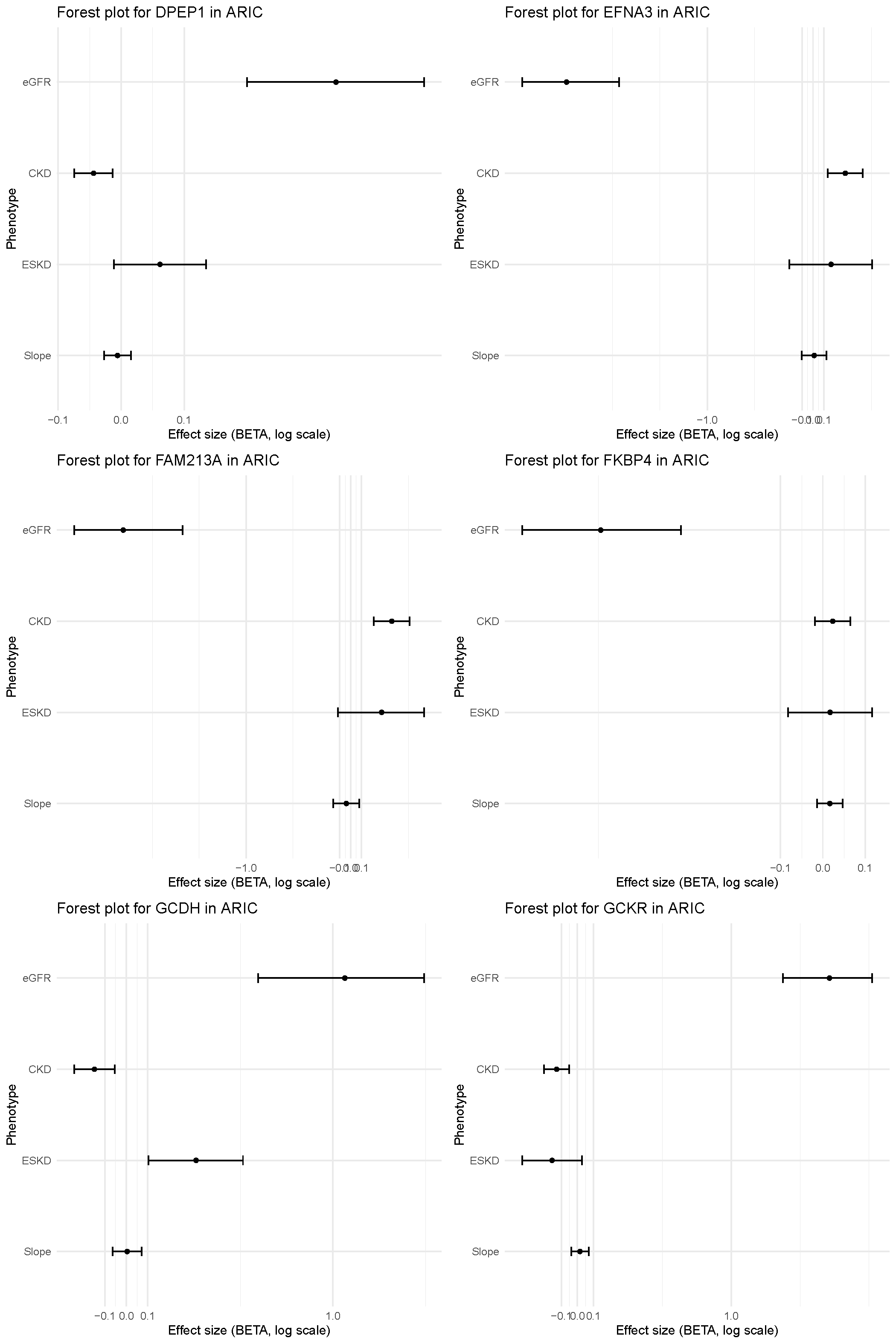

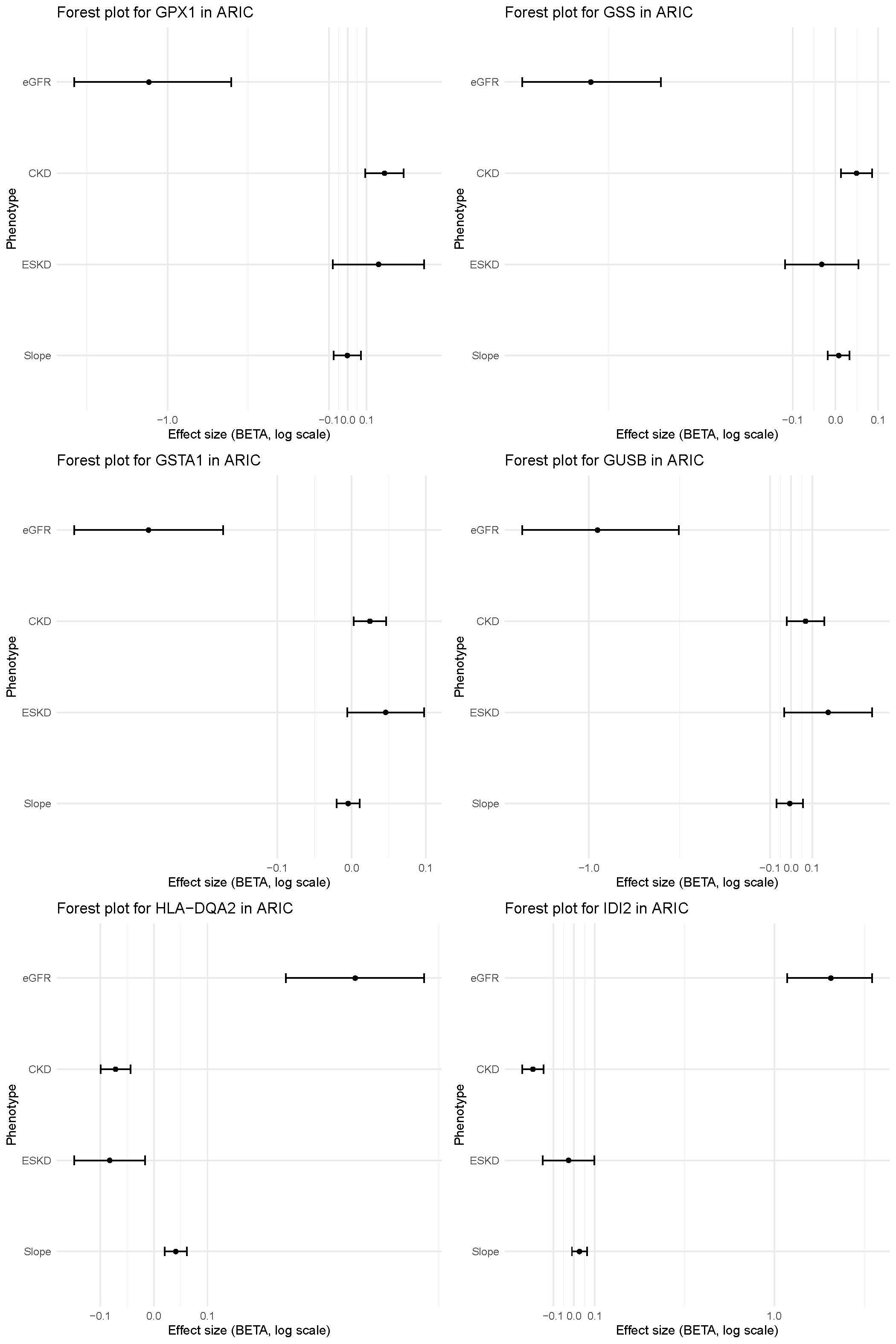

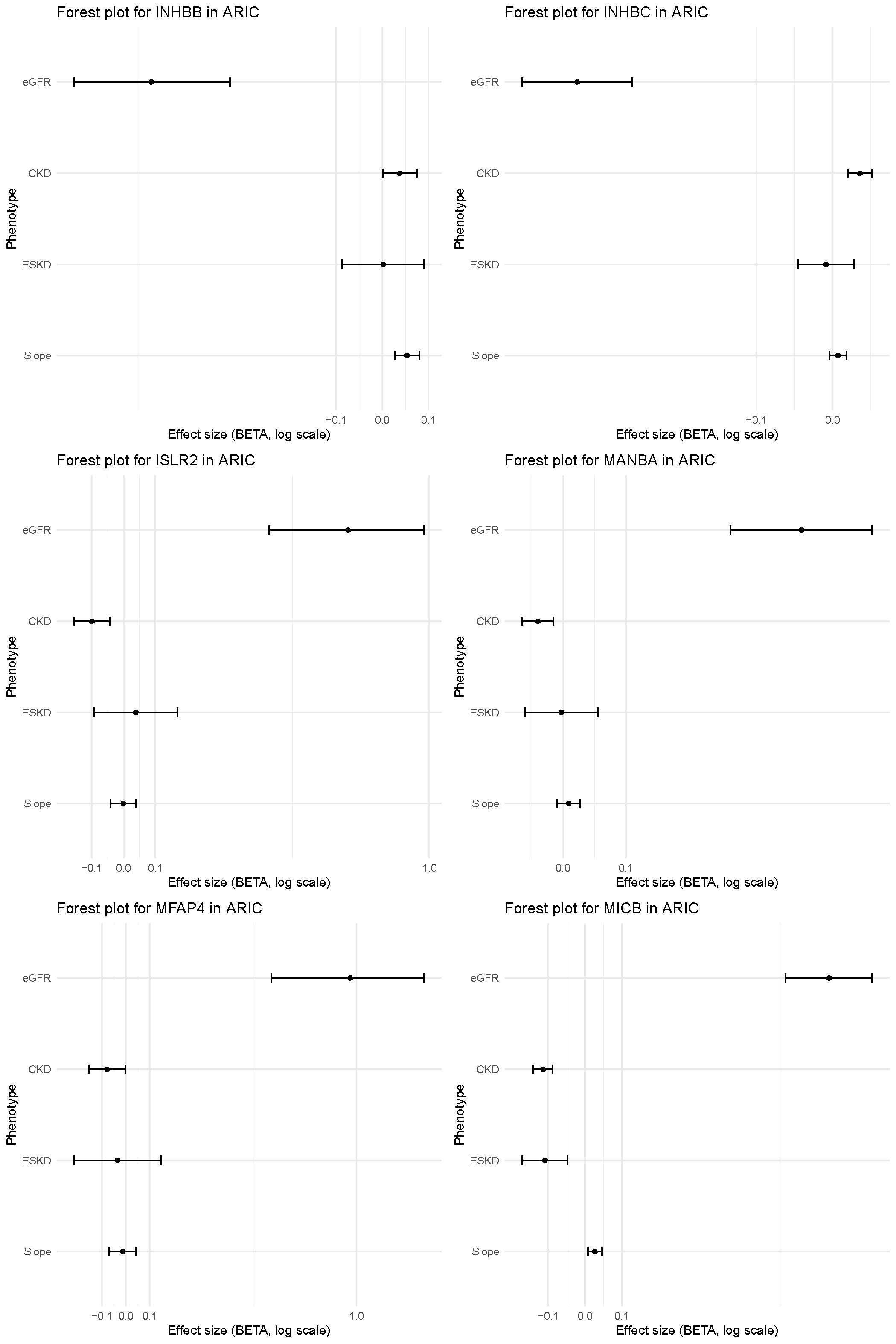

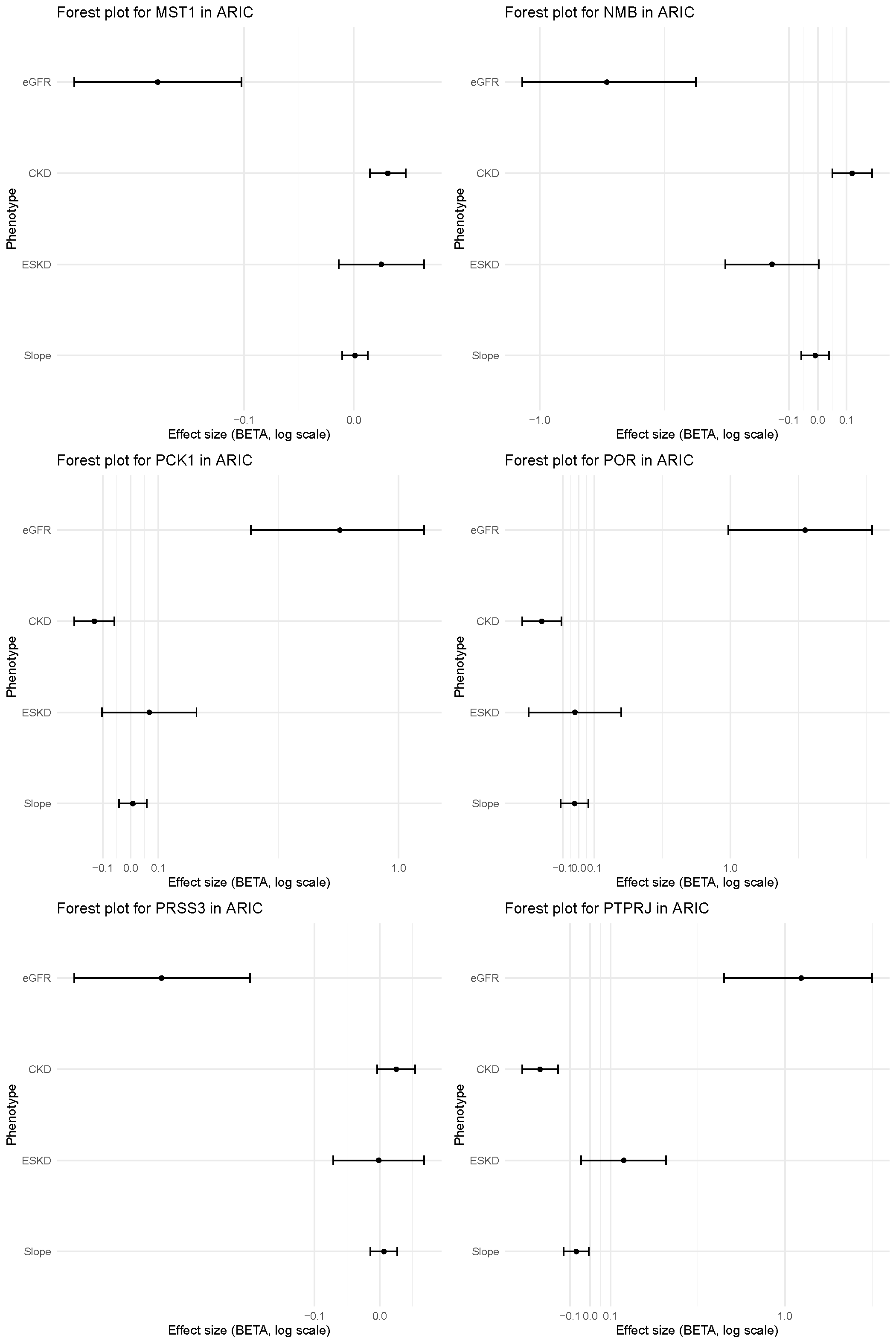

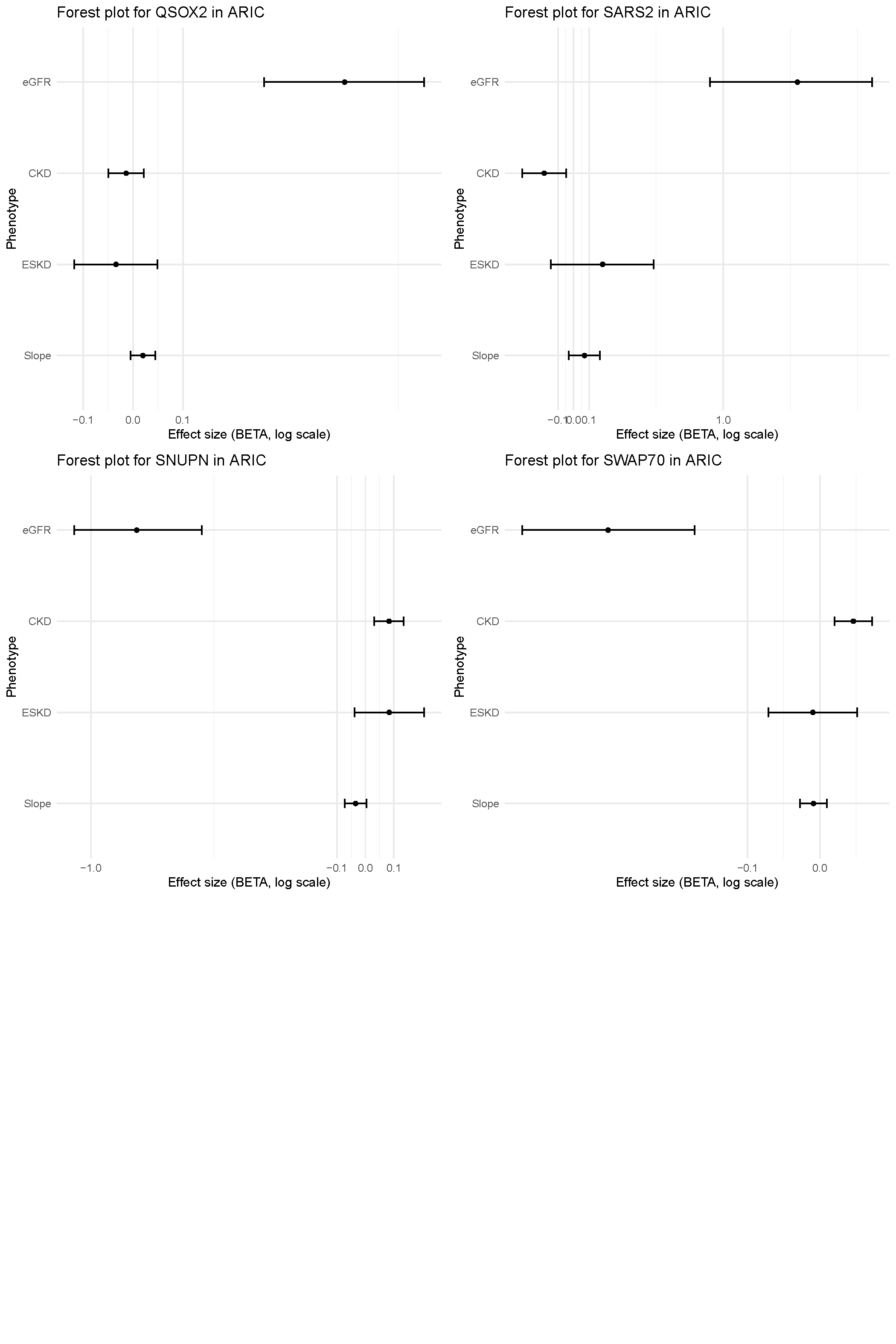

**Supplementary Figure 6. Forest plots of Fenland cis‑pQTLs that reached study‑wide significance and their association with the four MVP kidney outcomes.**

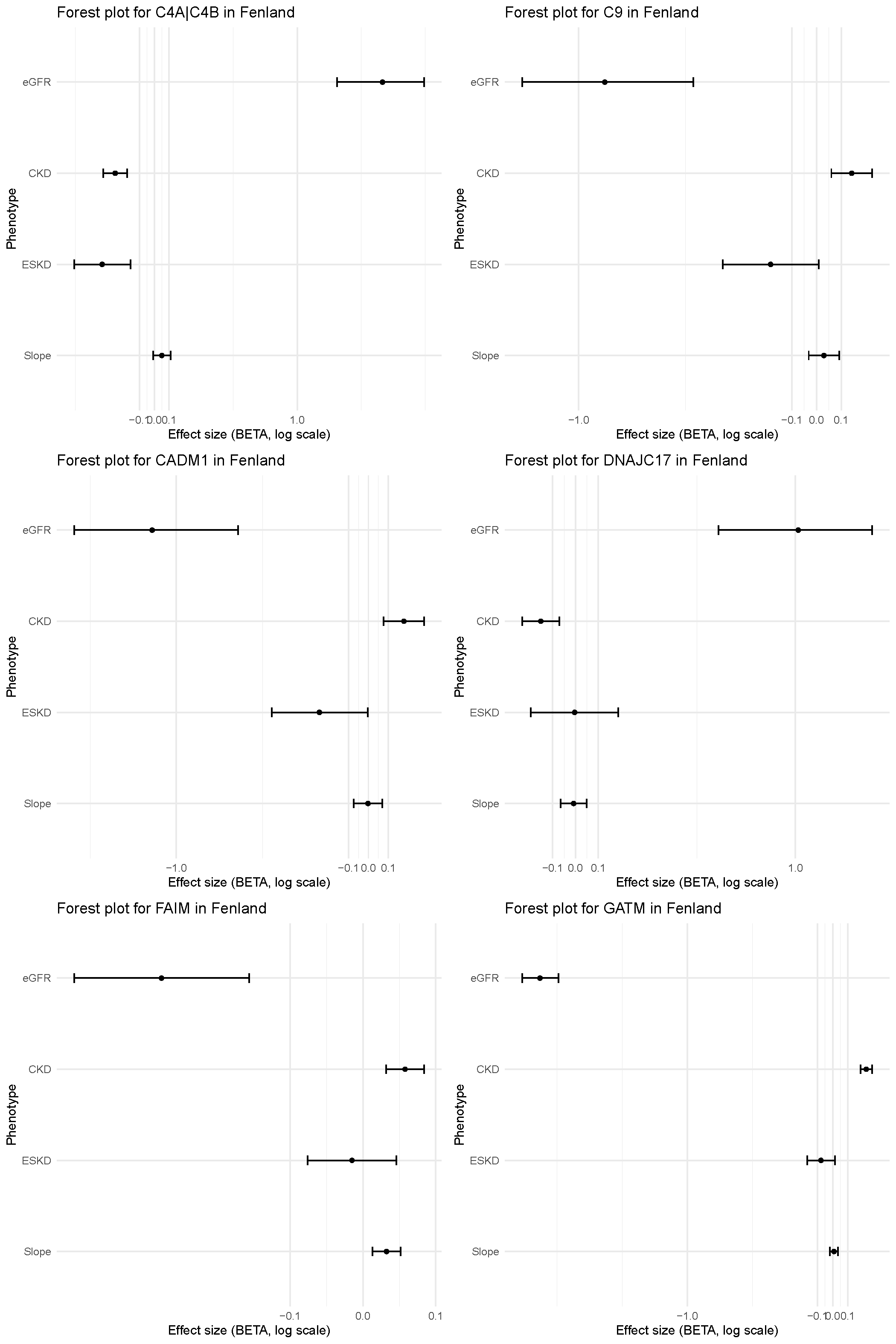

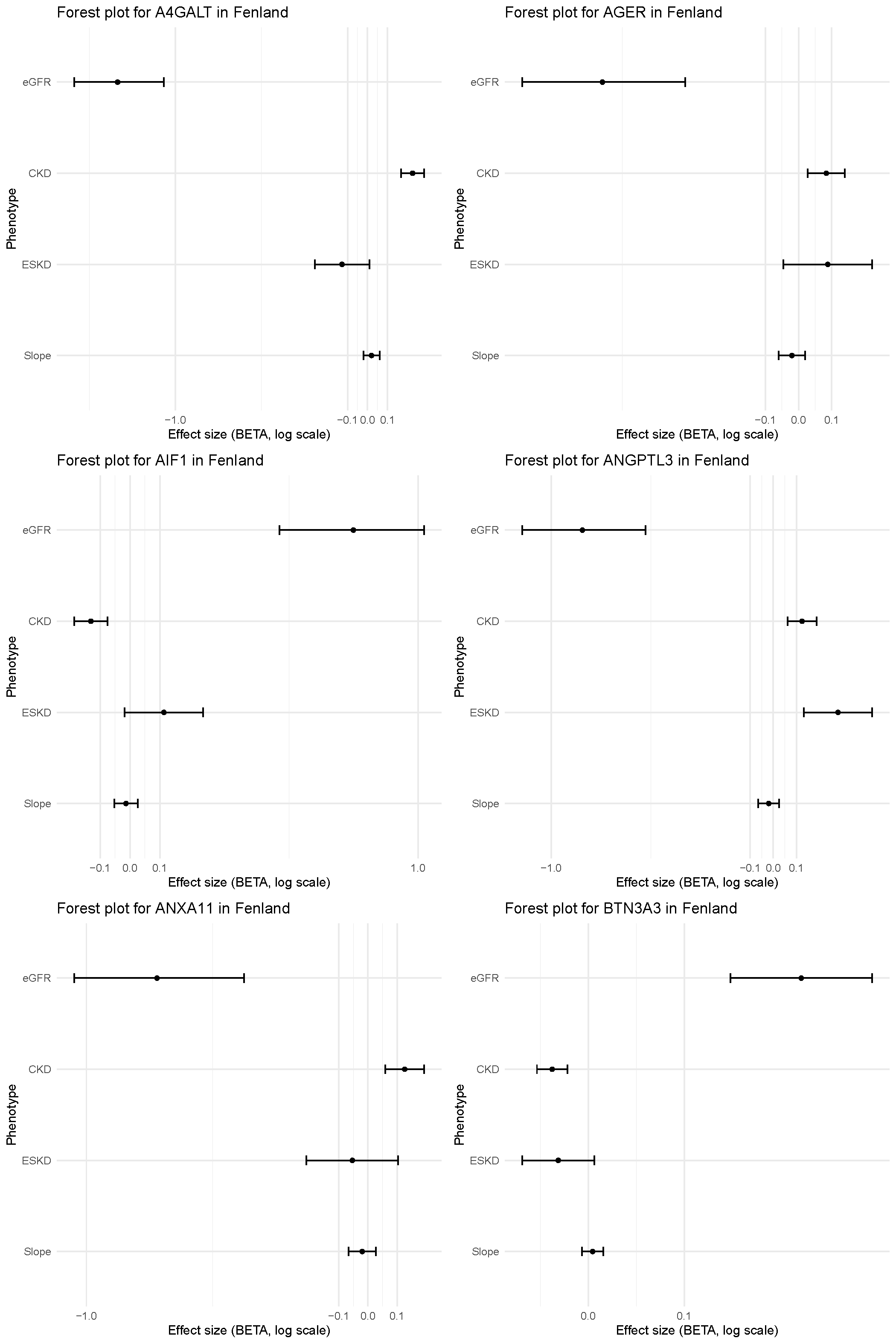

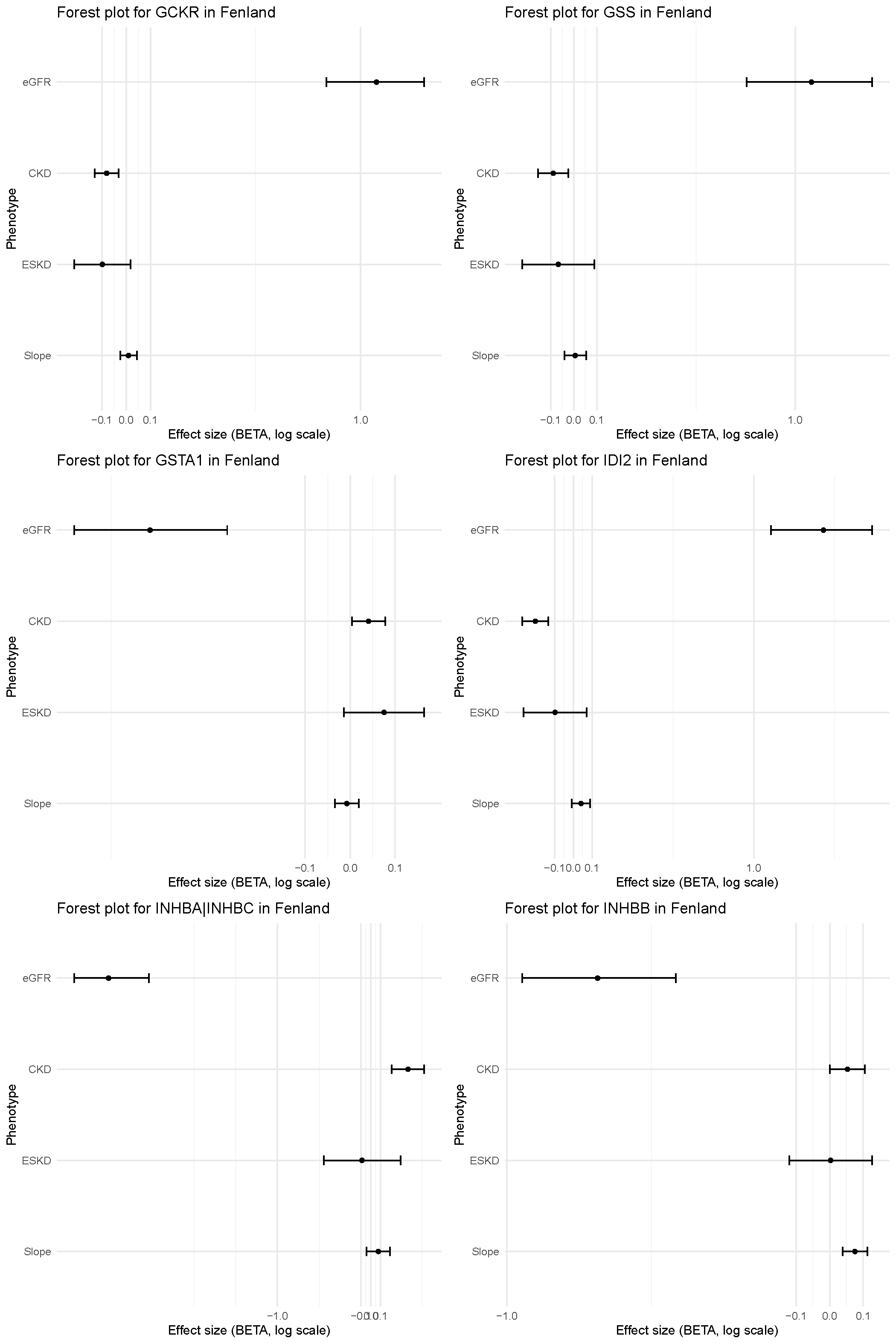

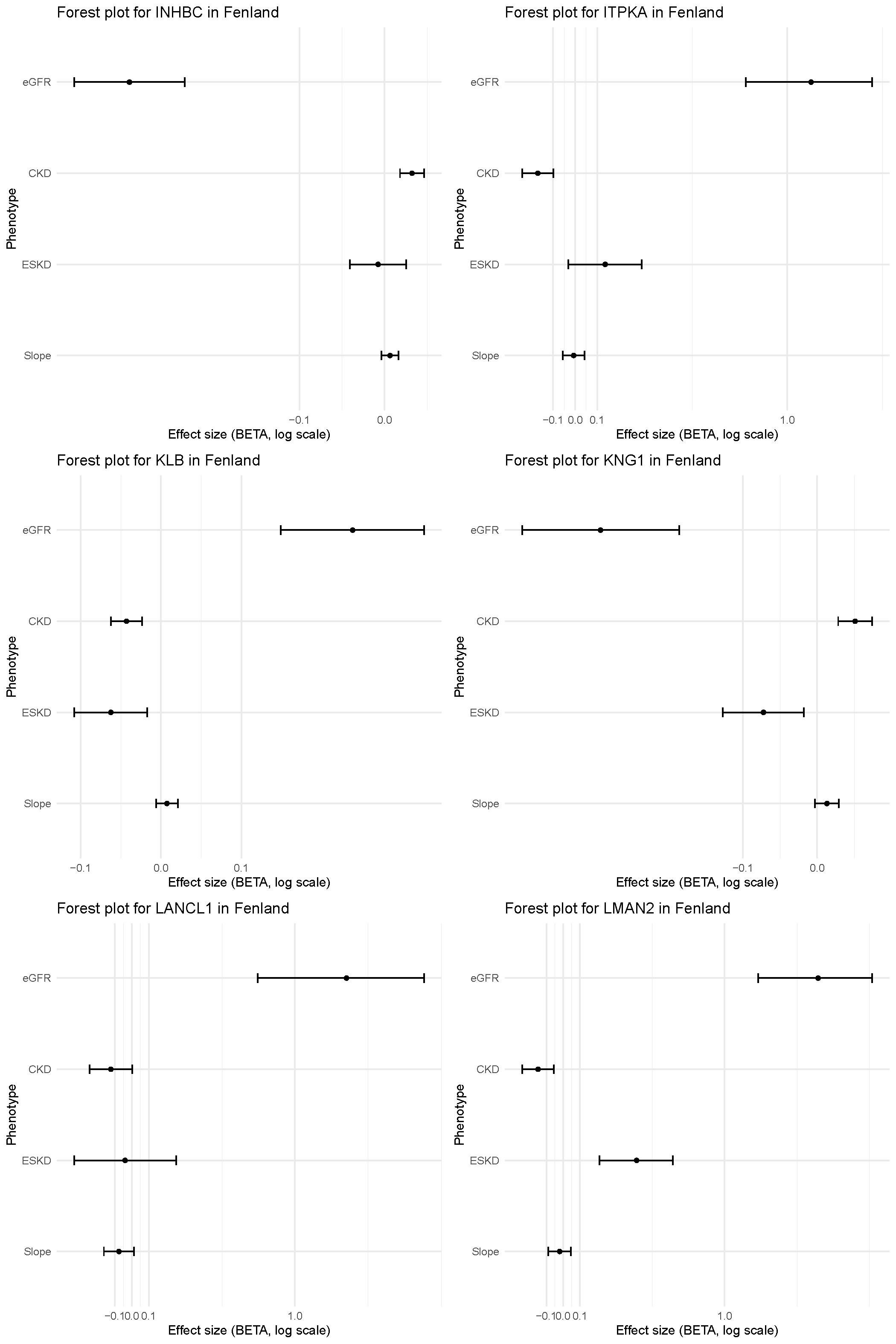

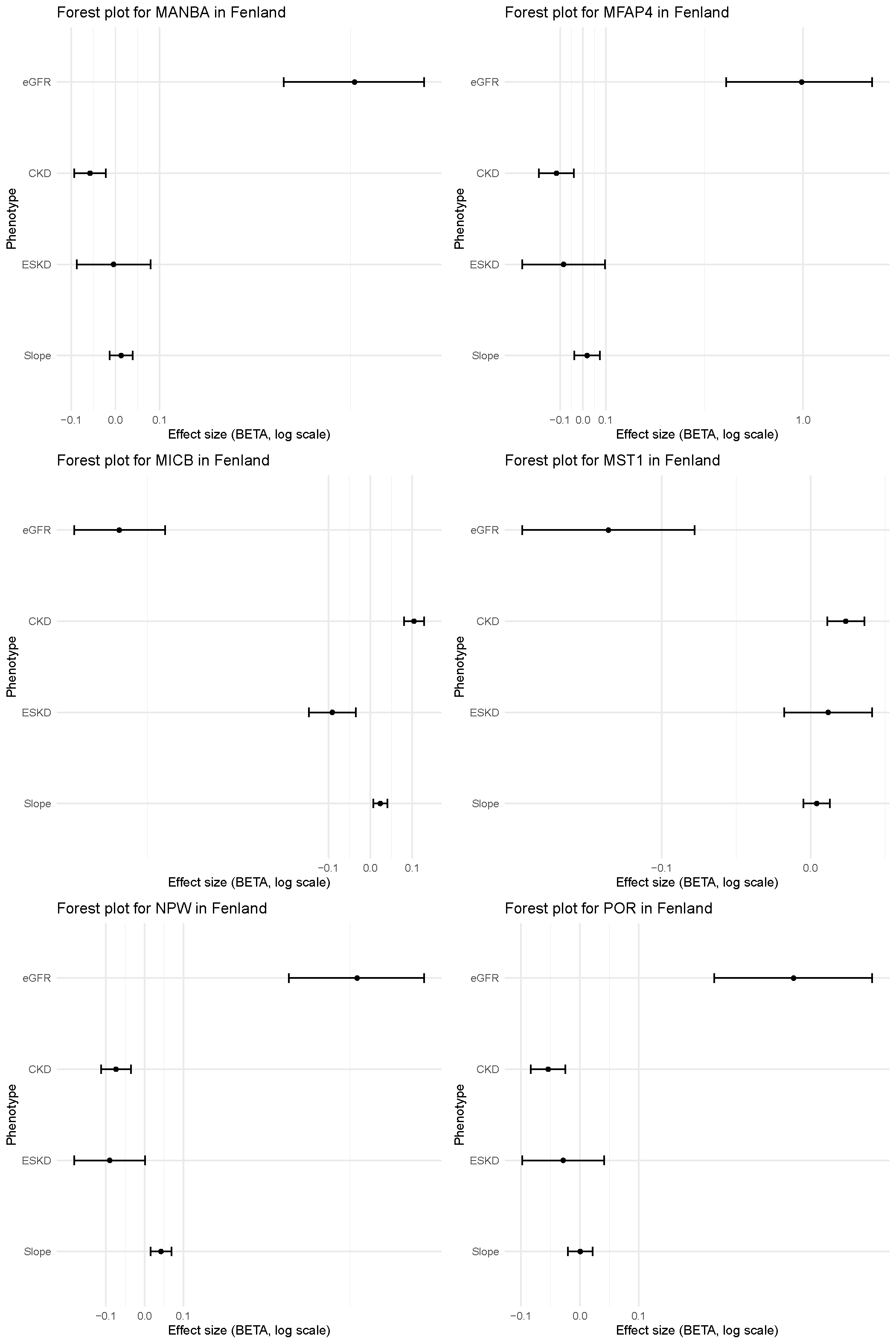

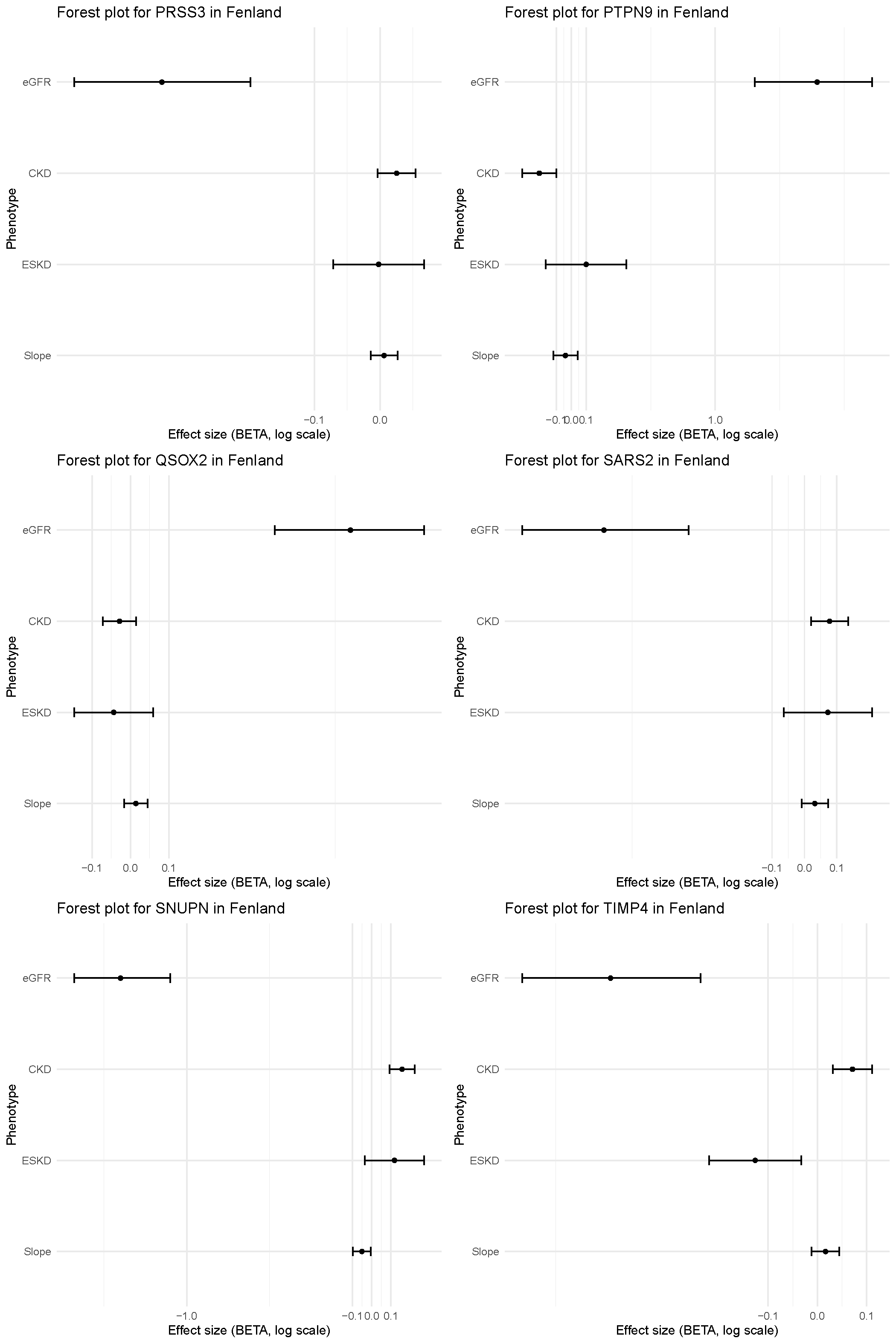

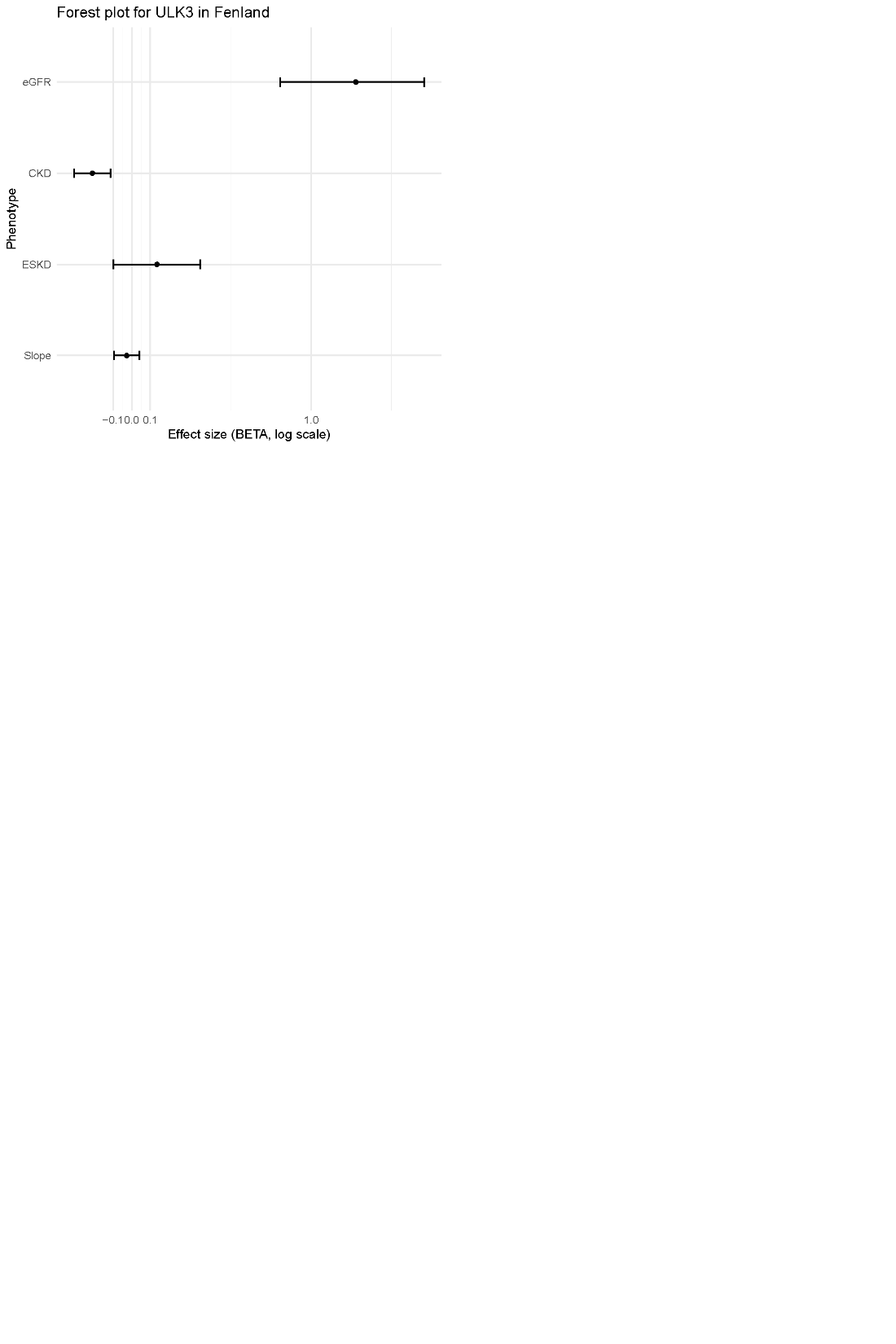

**Supplementary Figure 7. Forest plots of deCODE cis‑pQTLs that reached study‑wide significance and their association with the four MVP kidney outcomes.**

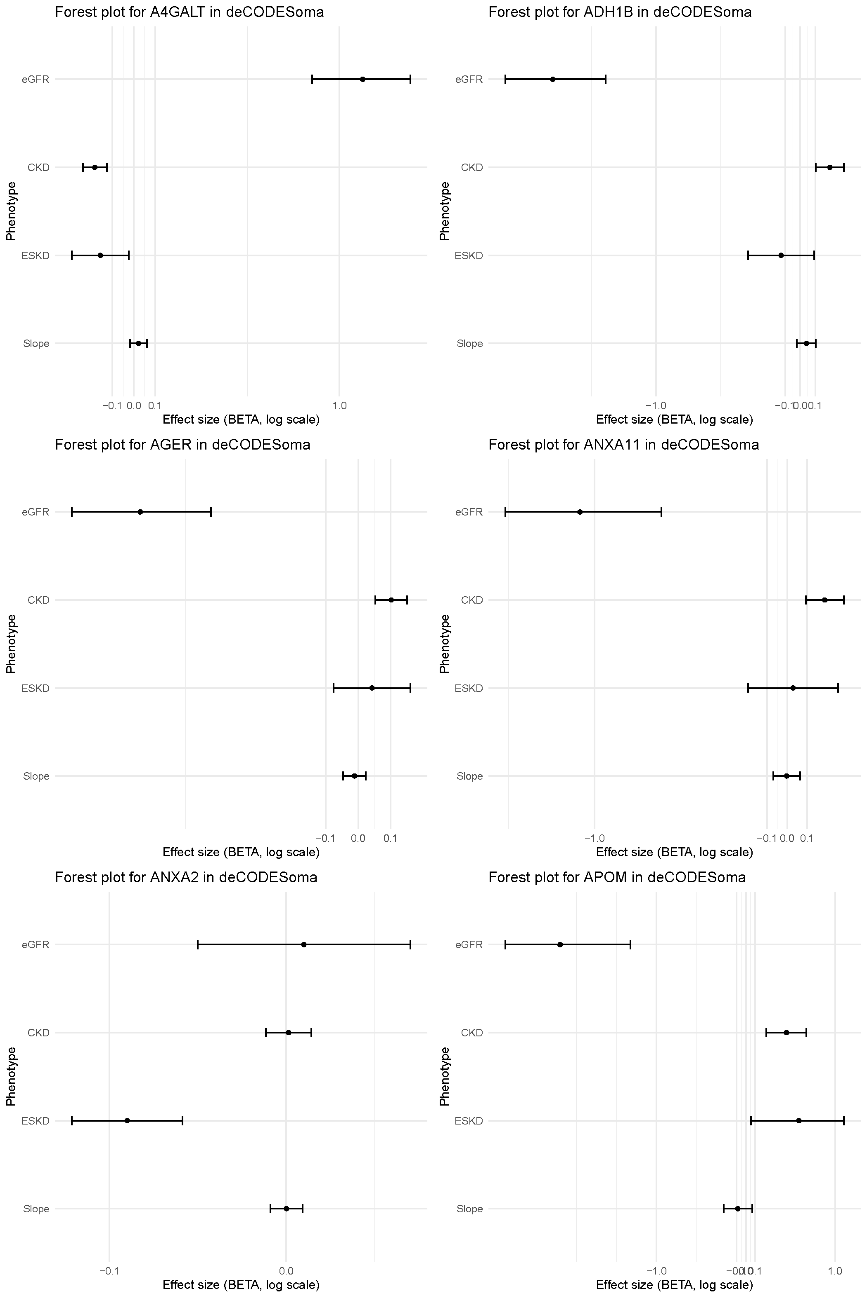

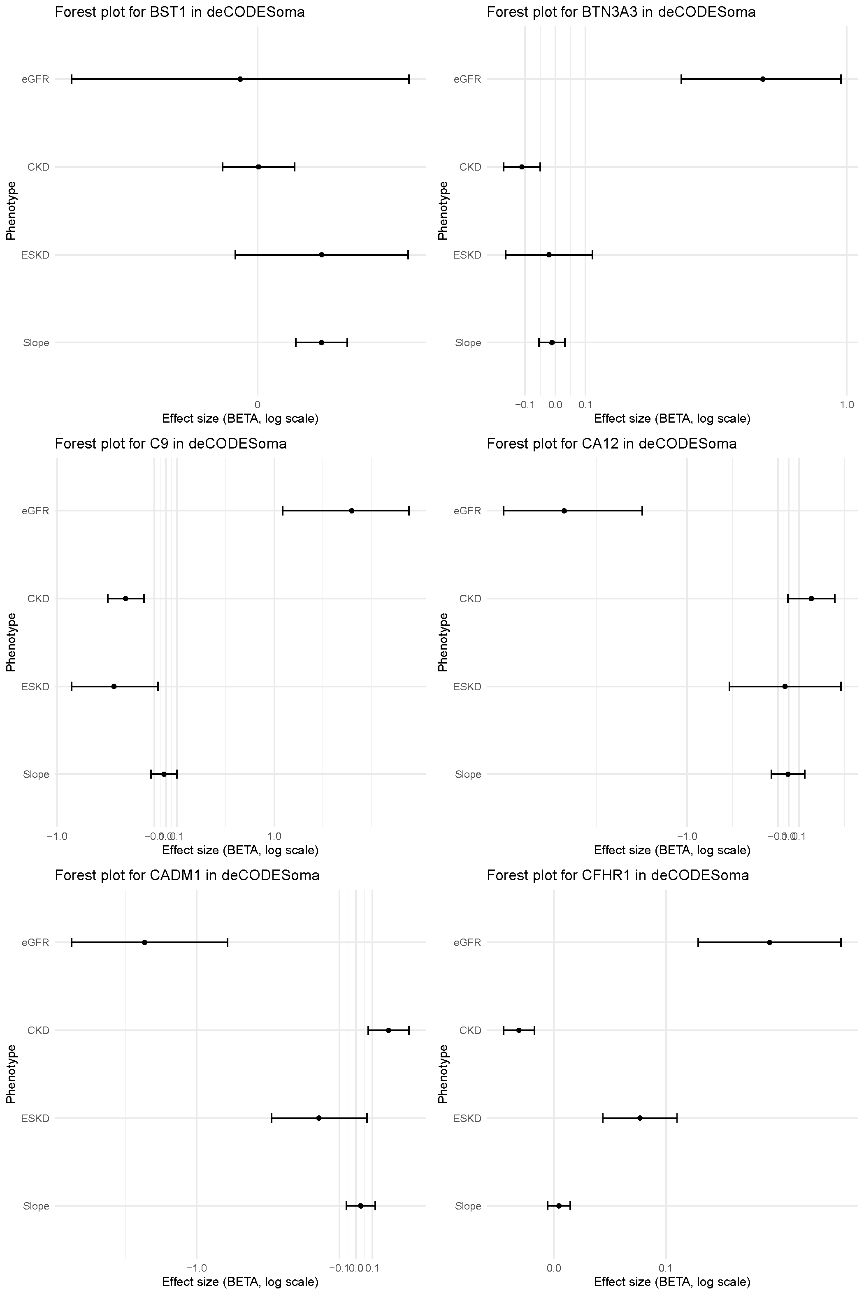

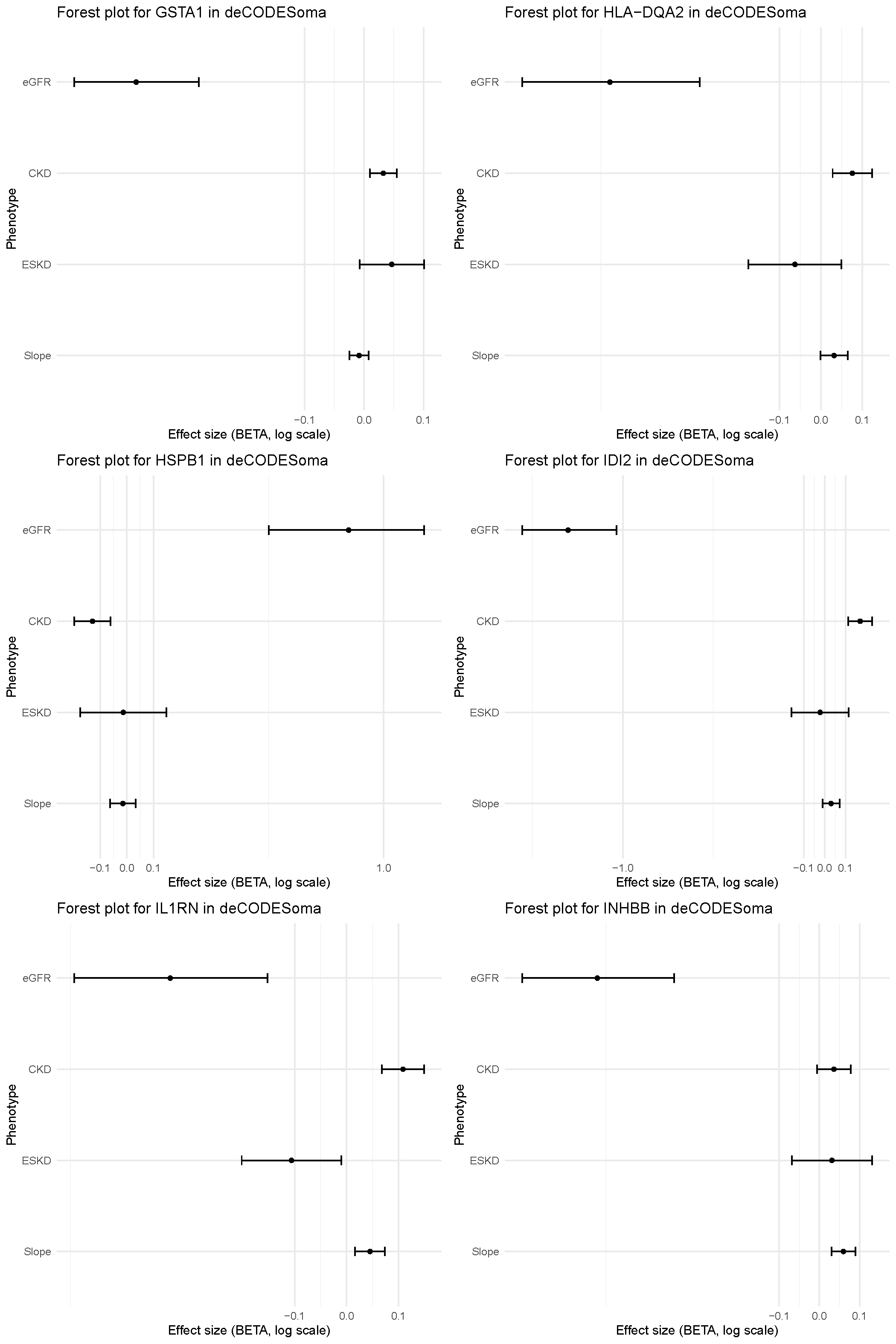

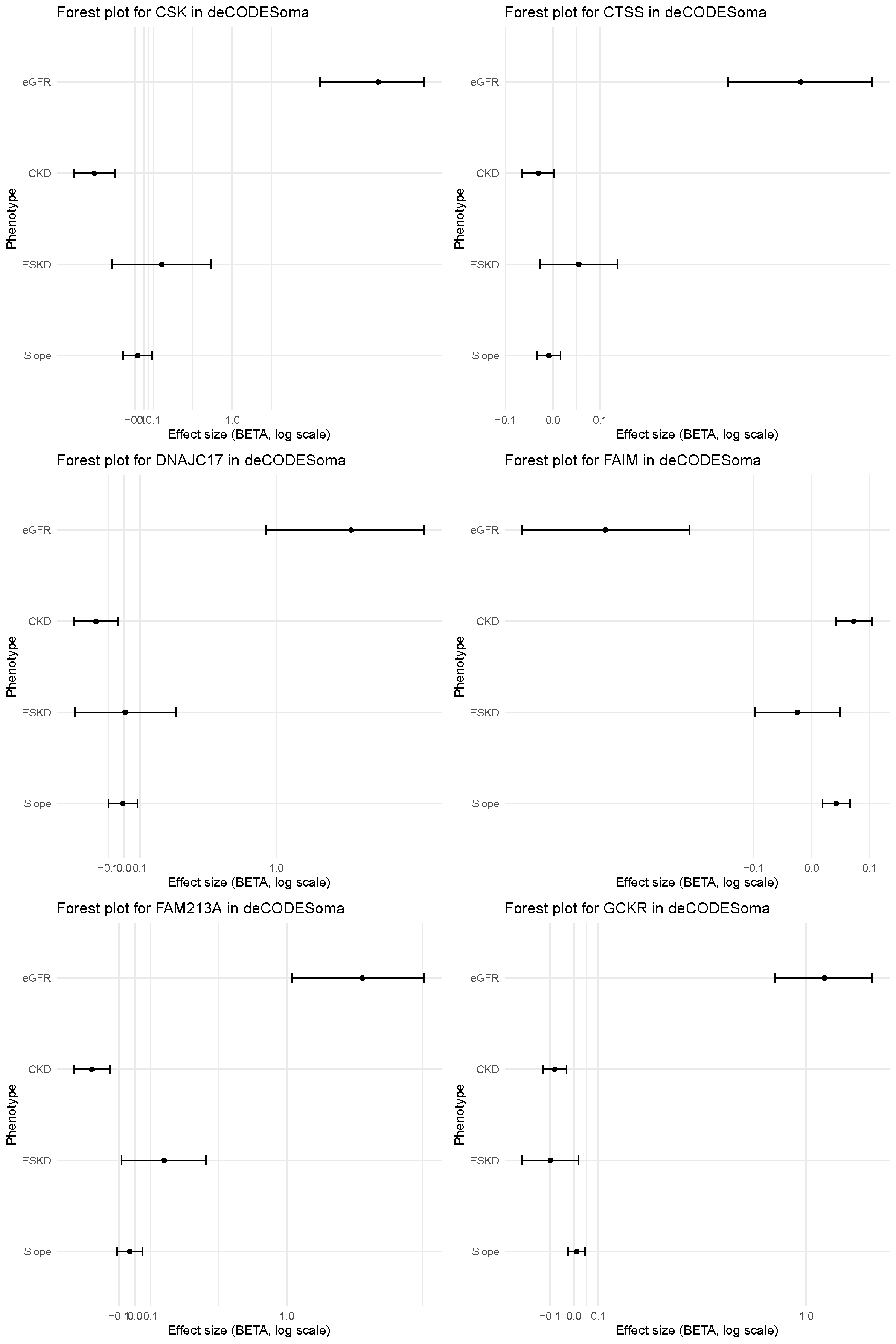

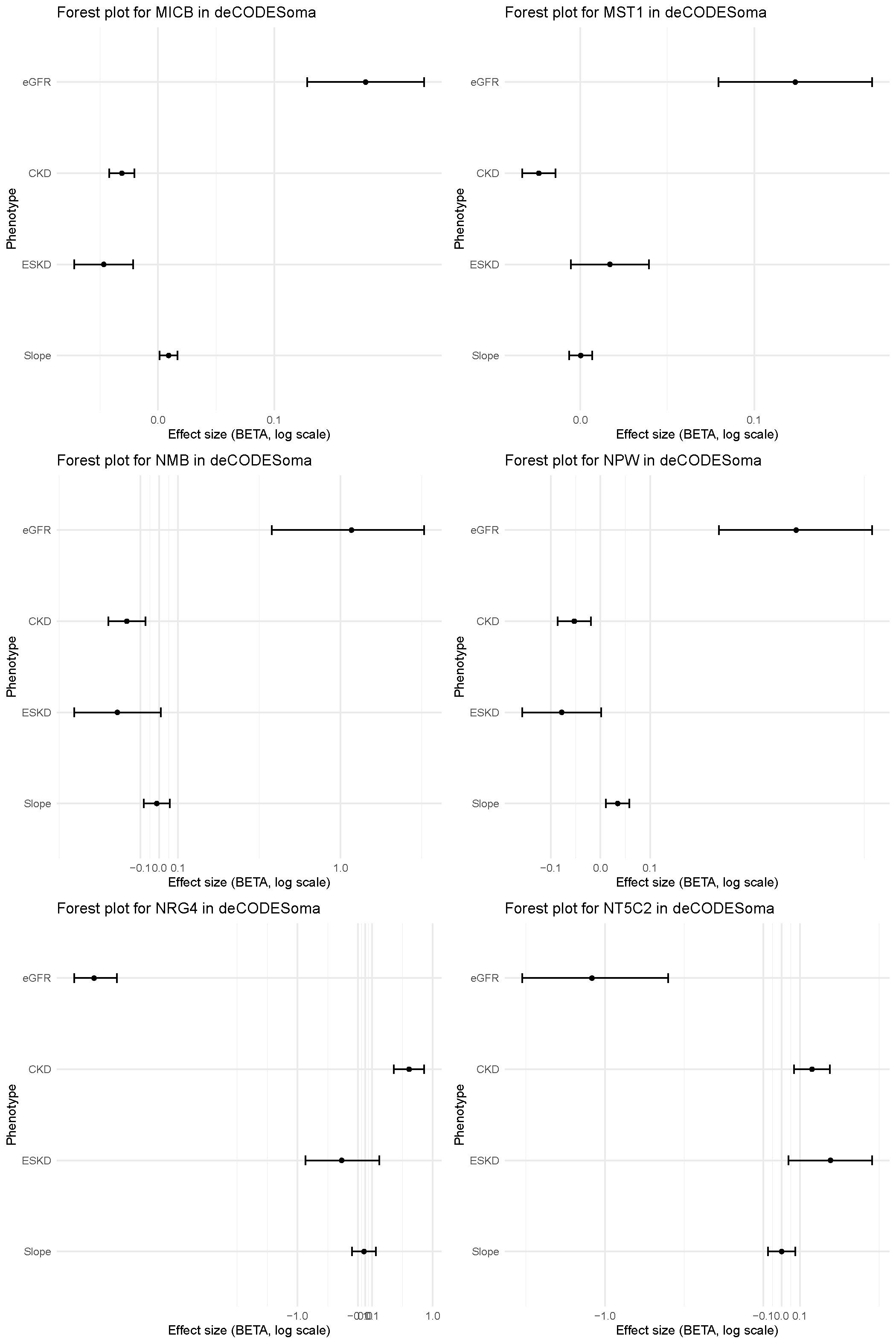

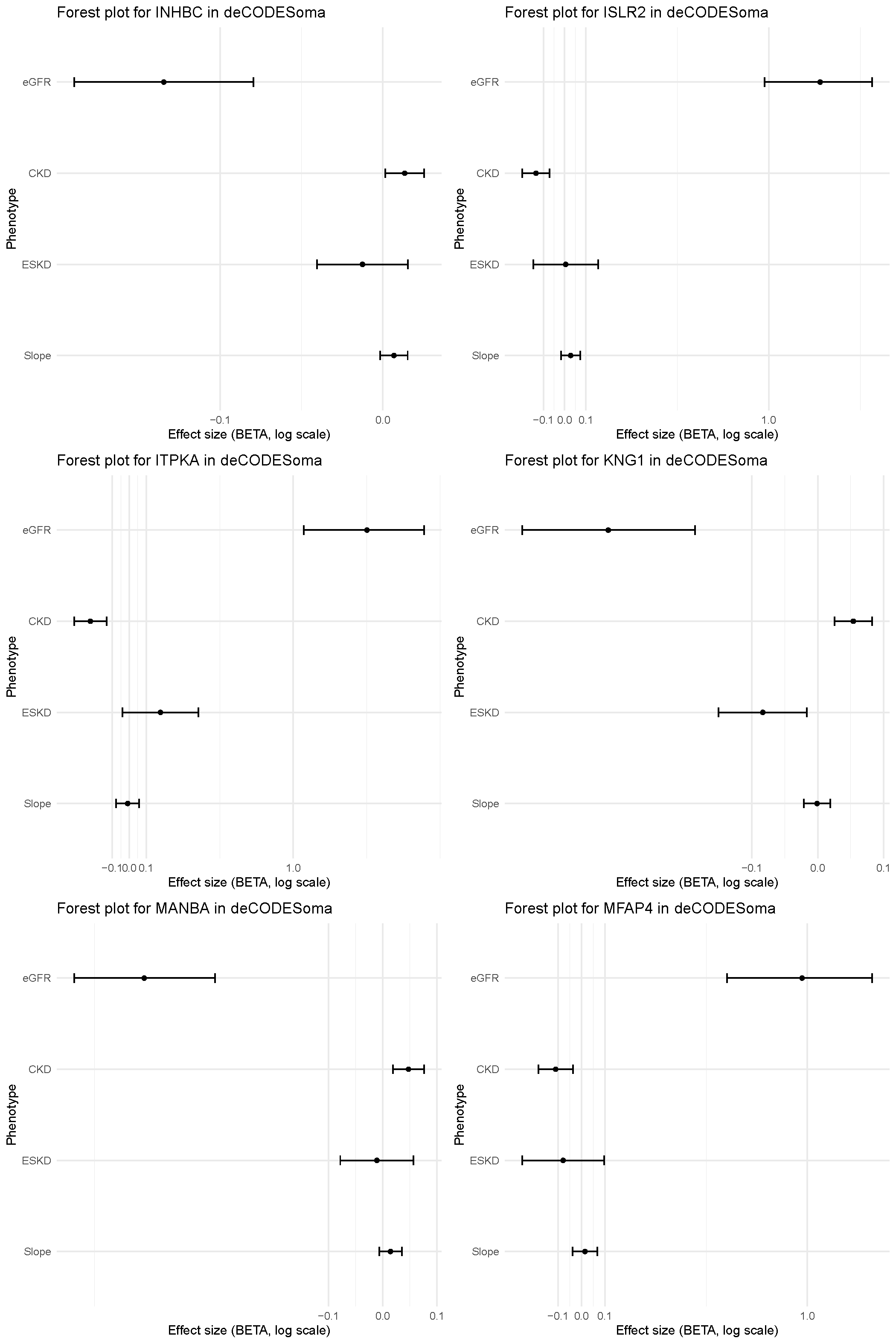

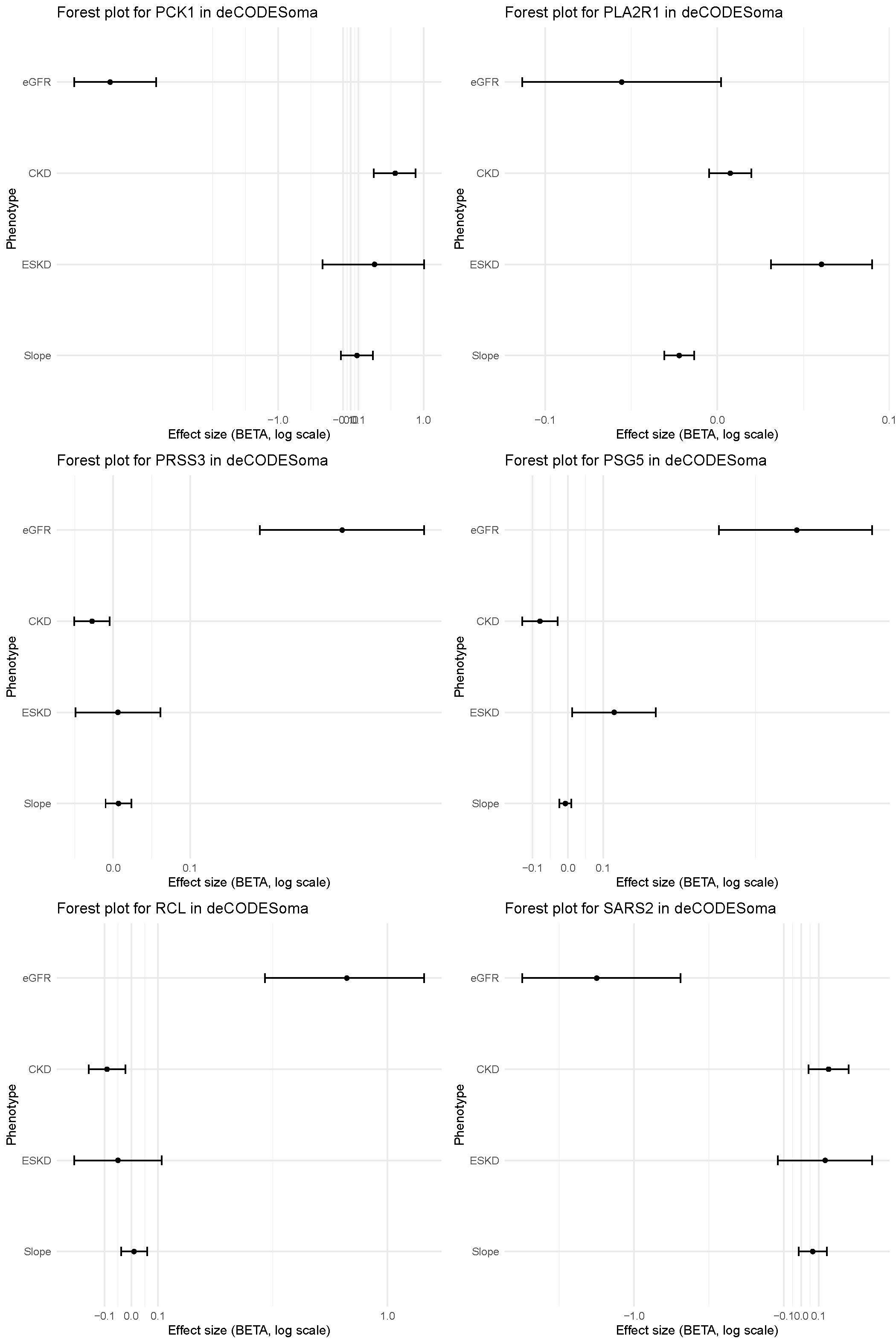

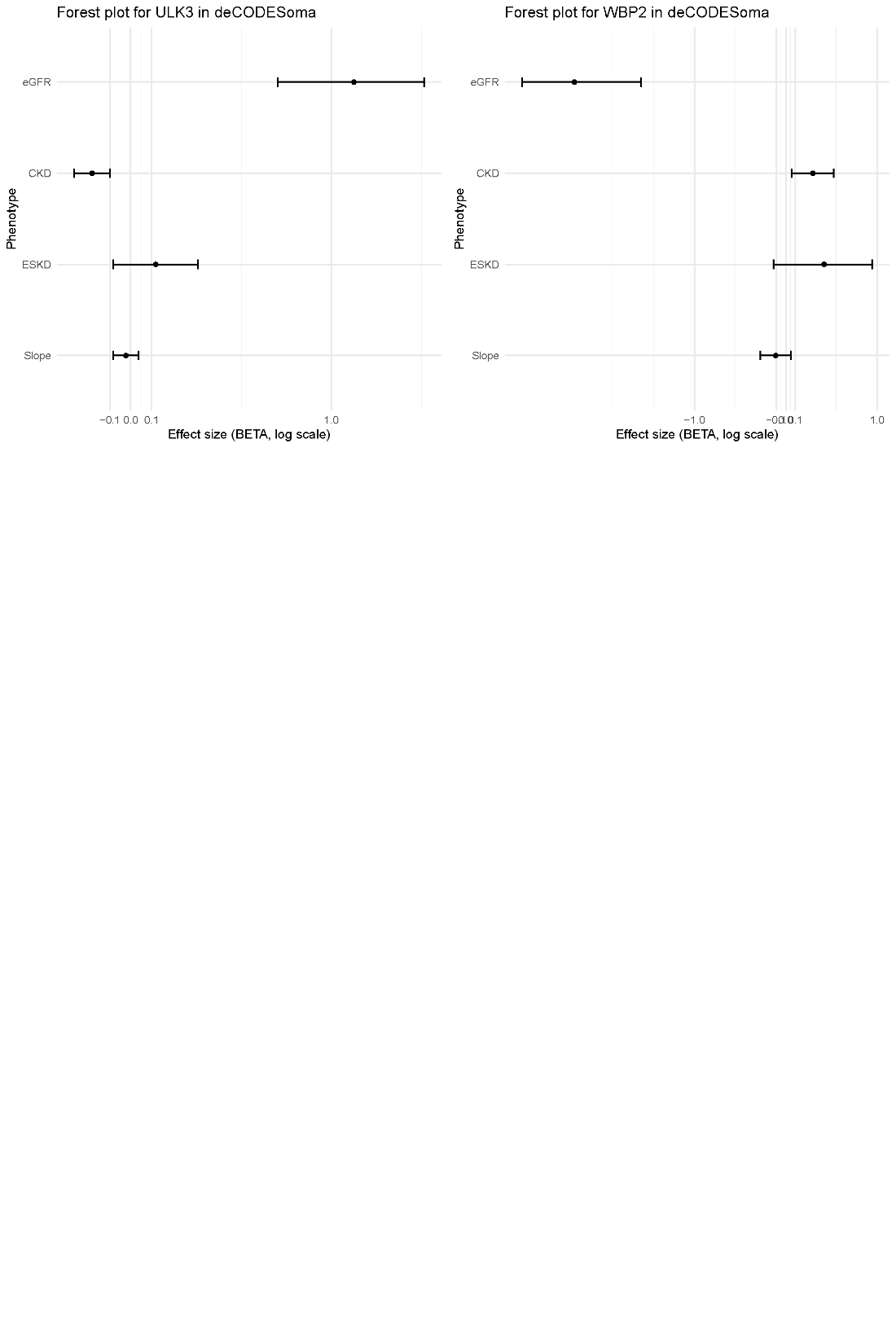

**Supplementary Figure 8. Forest plots of UK Biobank cis‑pQTLs that reached study‑wide significance and their association with the four MVP kidney outcomes.**

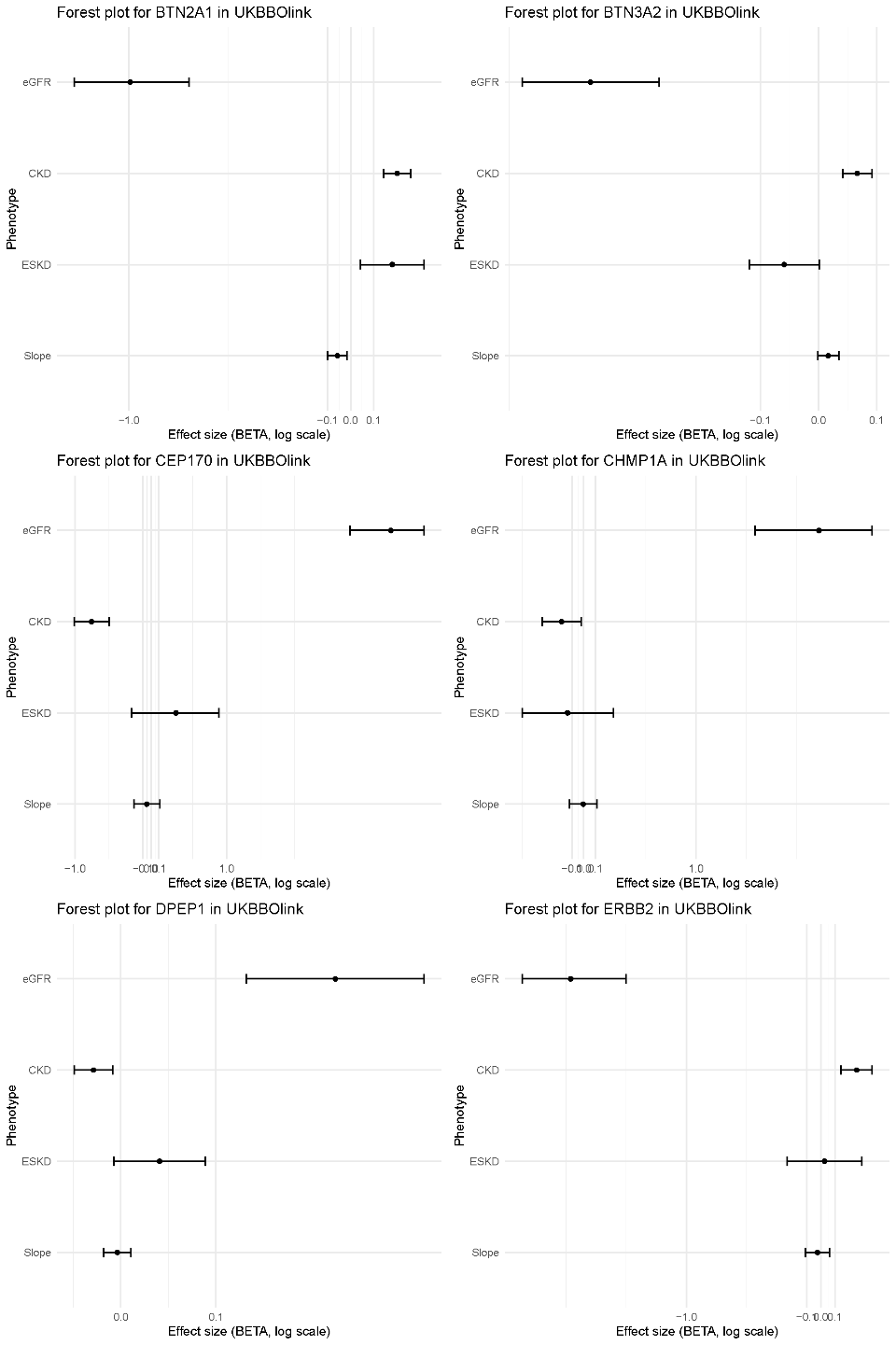

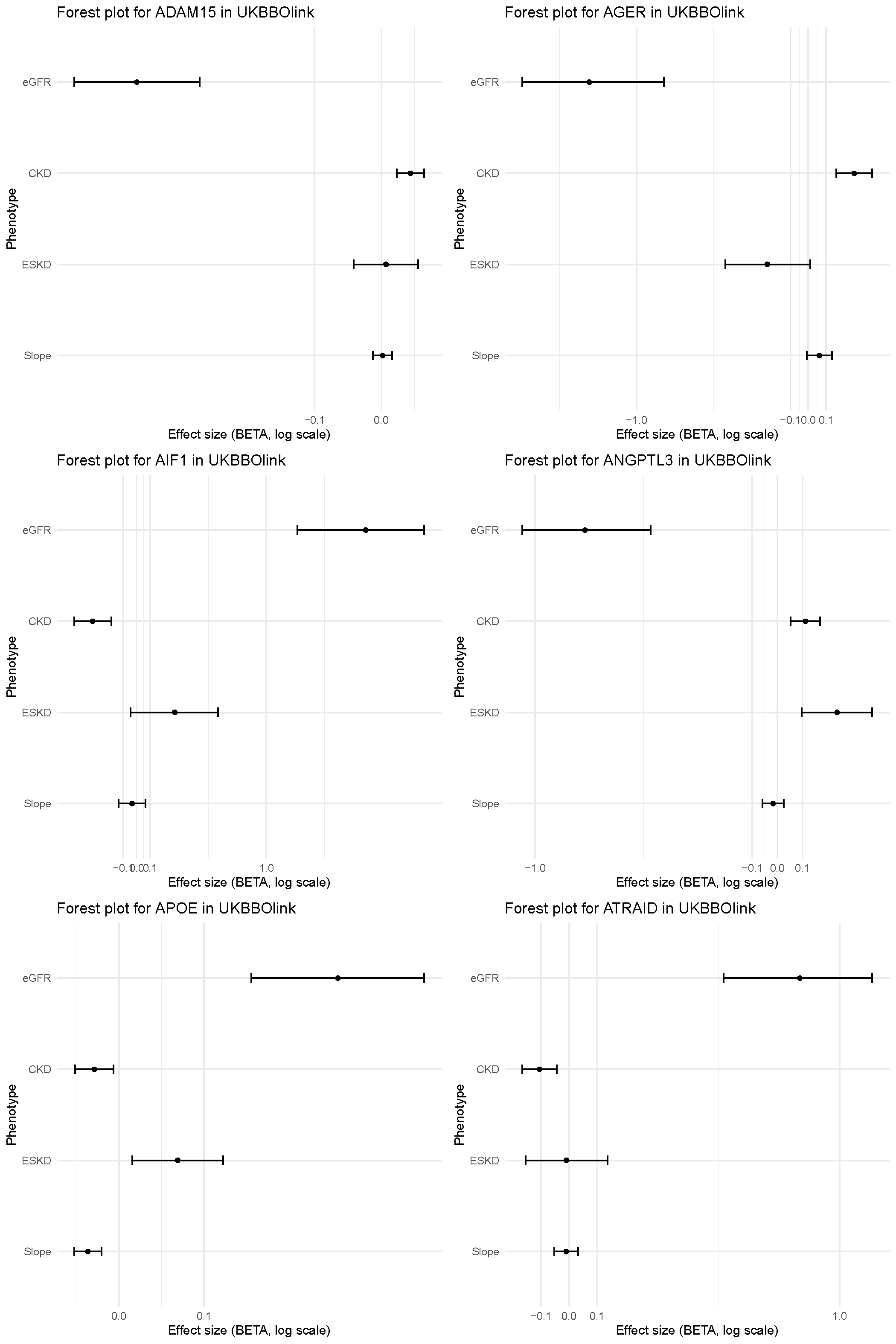

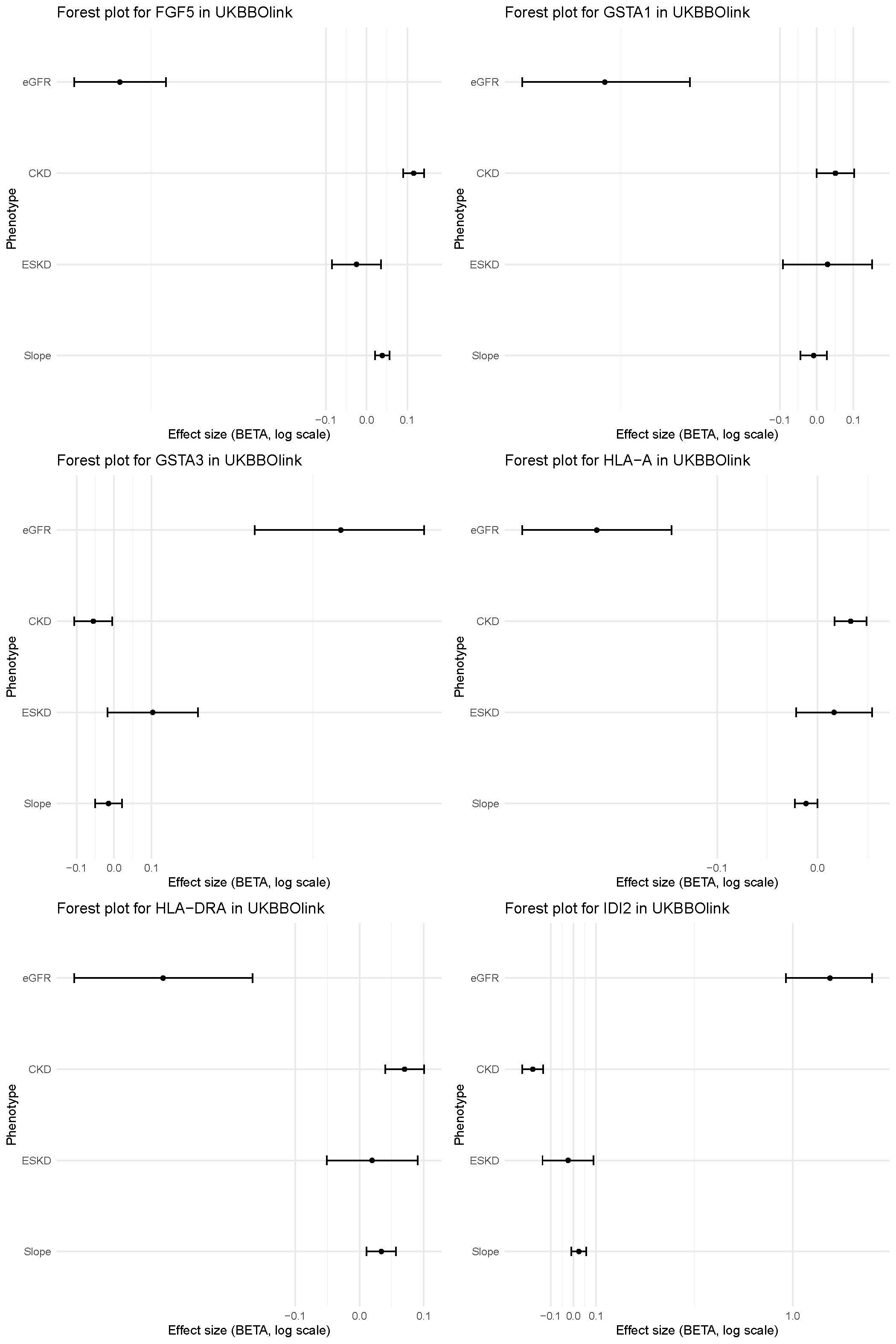

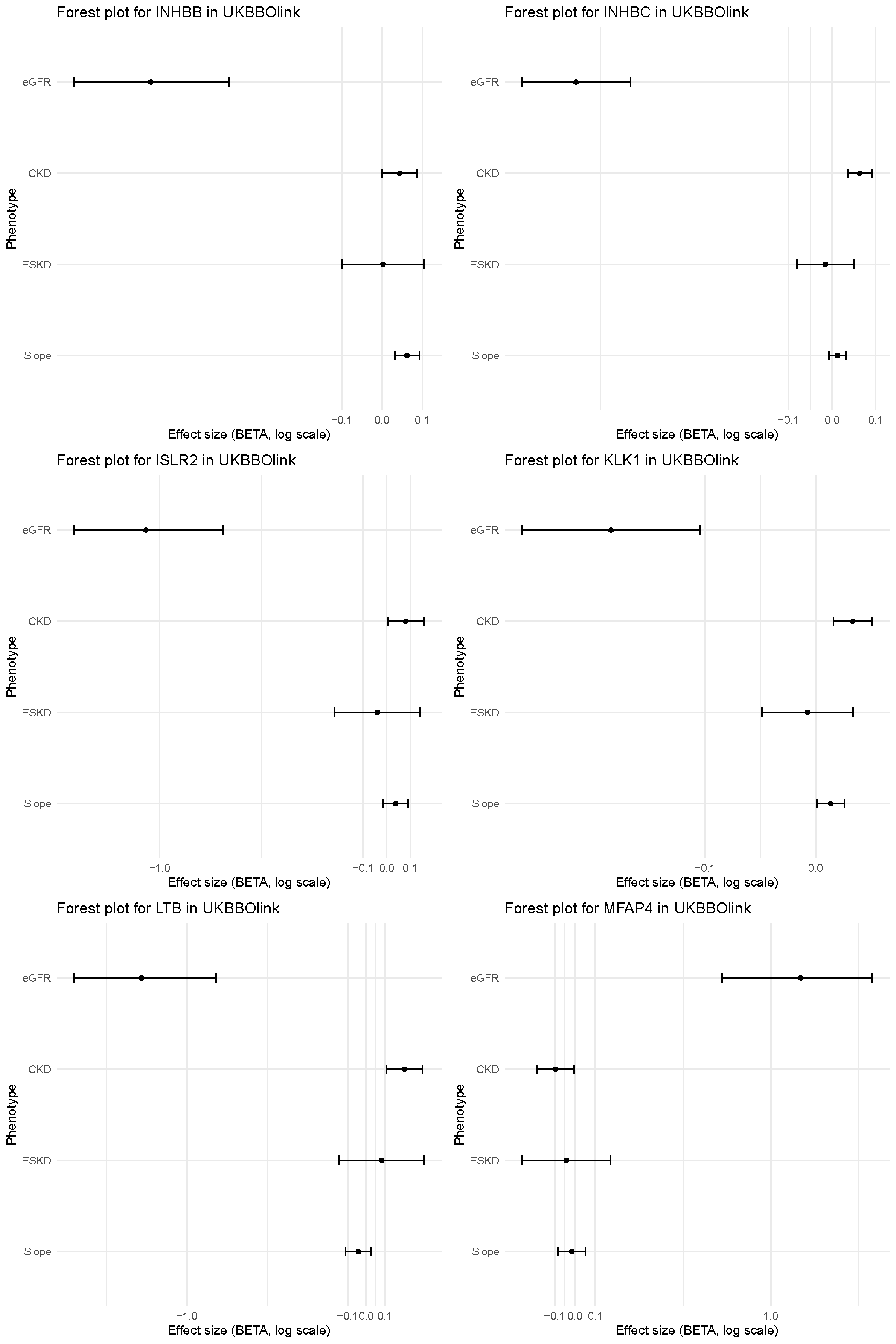

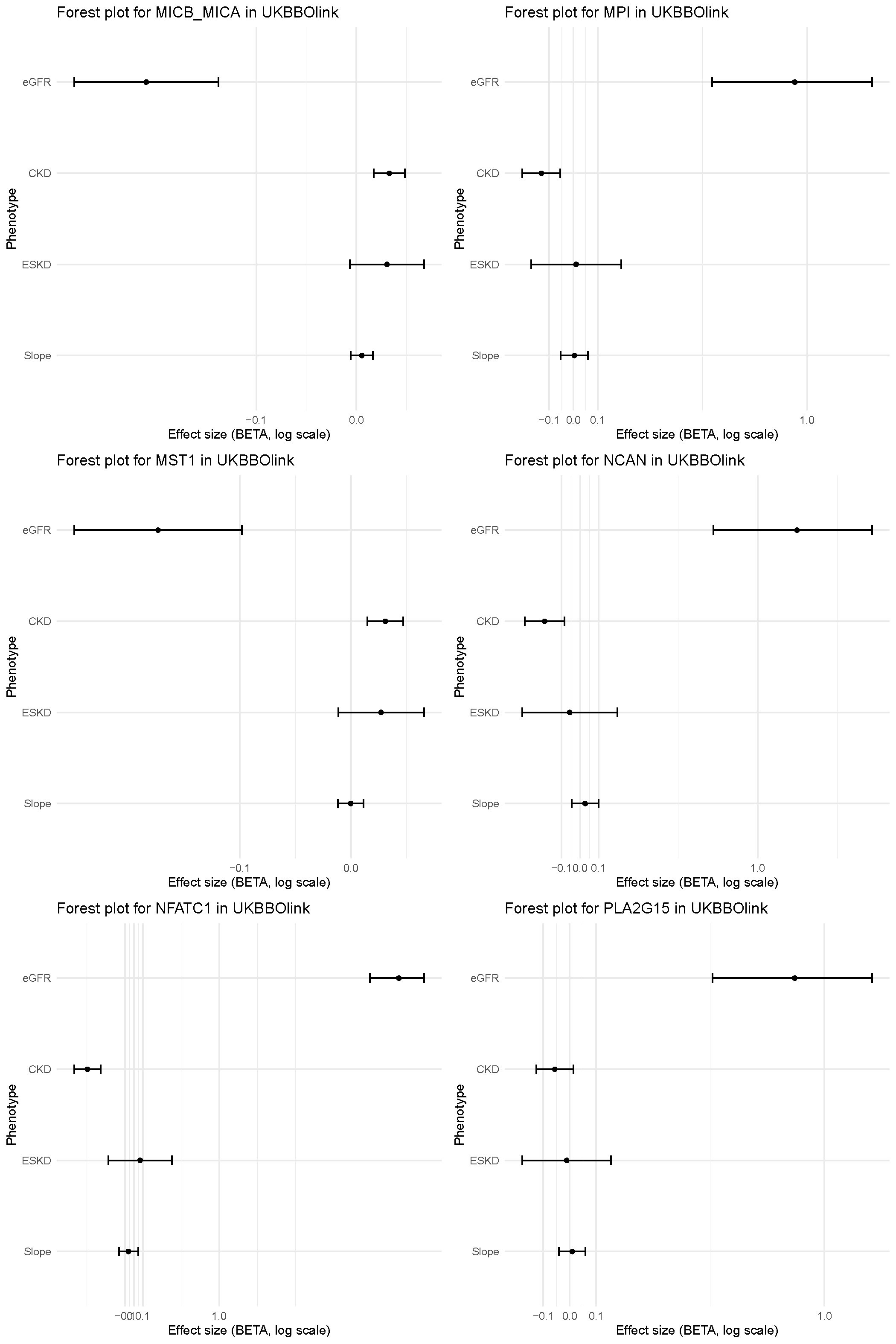

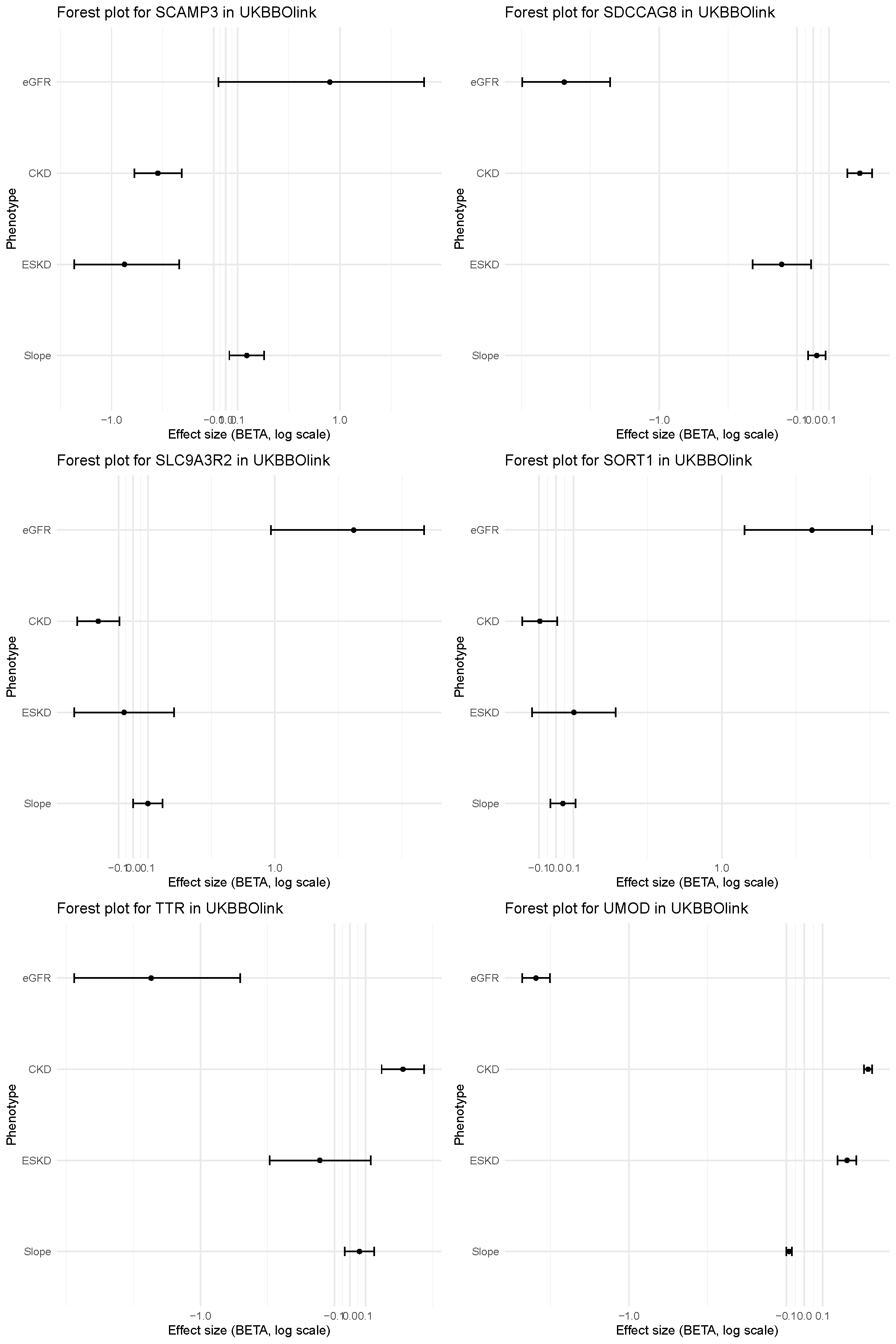

**Supplementary Figure 9. Scatter plot illustrates diabetes-dependent variability in genetic associations between plasma proteins and the four components of kidney outcome.** Proteins below the diagonal line have stronger negative associations with eGFR in diabetic compared to non-diabetic individuals. Each point represents the genetic effect estimate (Beta) of a plasma protein on estimated glomerular filtration rate (eGFR) in diabetic (y-axis) versus non-diabetic (x-axis) individuals. Proteins demonstrating statistically significant heterogeneity (P<0.05/93, Z-test) between diabetes strata are highlighted in red and labeled explicitly. The dashed diagonal reference line indicates identical effect sizes across diabetes strata. Deviation from this line reveals diabetes-specific modulation of the protein-kidney function relationship Proteins above the diagonal line indicate stronger negative associations in non-diabetic individuals.

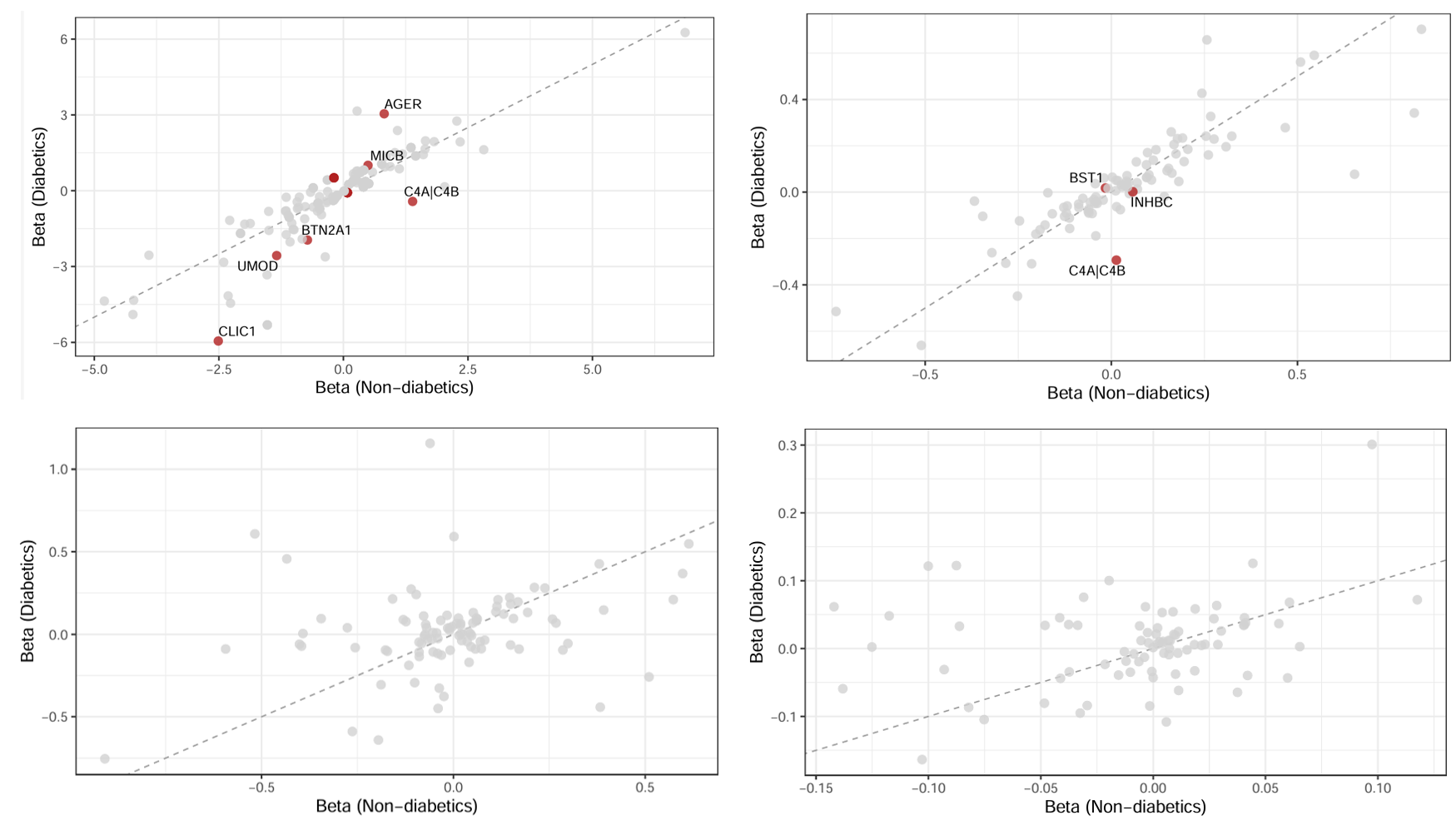

**Supplementary Figure 10. UpSet plot of phenome‑wide protein associations across clinical domains.** Shows pleiotropy among proteins that reached Bonferroni-adjusted significance in the PheWAS. Bars quantify the clinical domains number of proteins whose cis-pQTLs were significantly associated with one phecode in that domain. Kidney phecodes were separated as their own domain to highlight renal-specific signals. This is conditional on all being associated with the primary kidney function outcome.

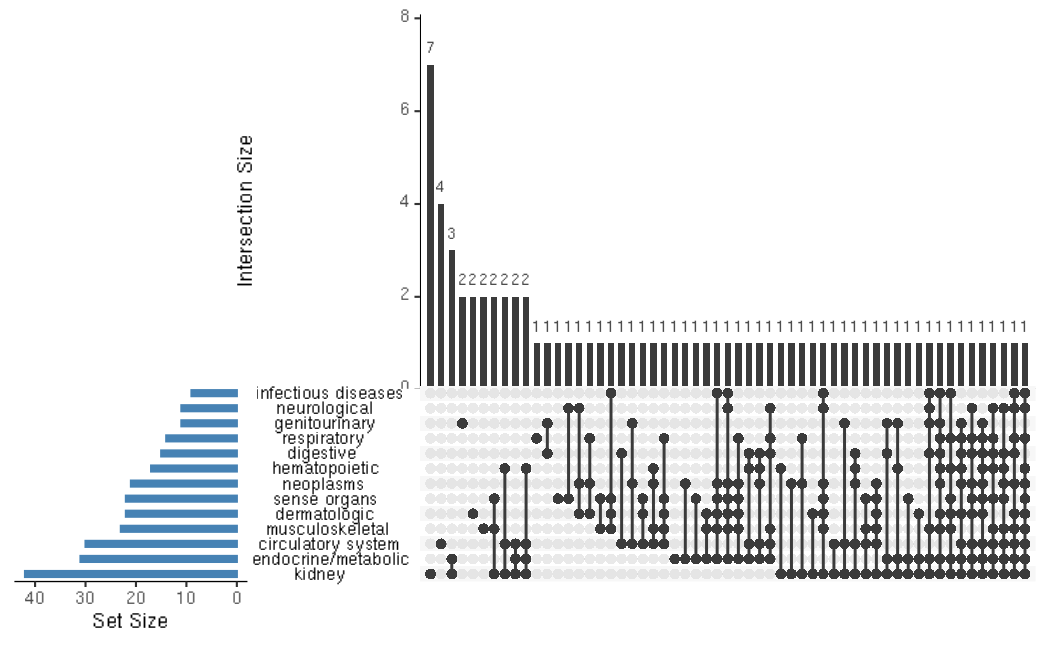

**Supplementary Figure 11. A PheWAS Volcano plot for each of the 93 proteins associated with the primary kidney outcome.**

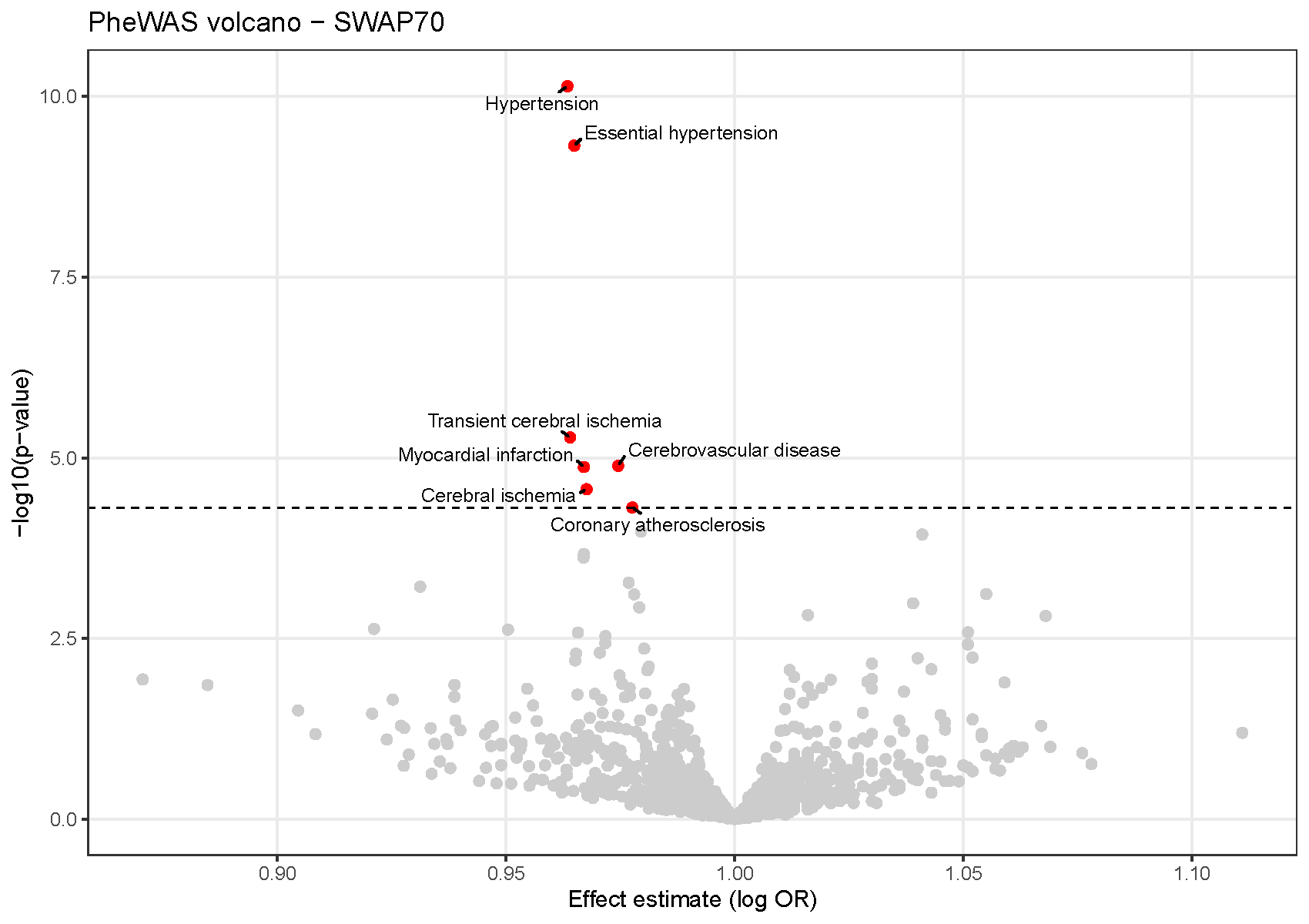

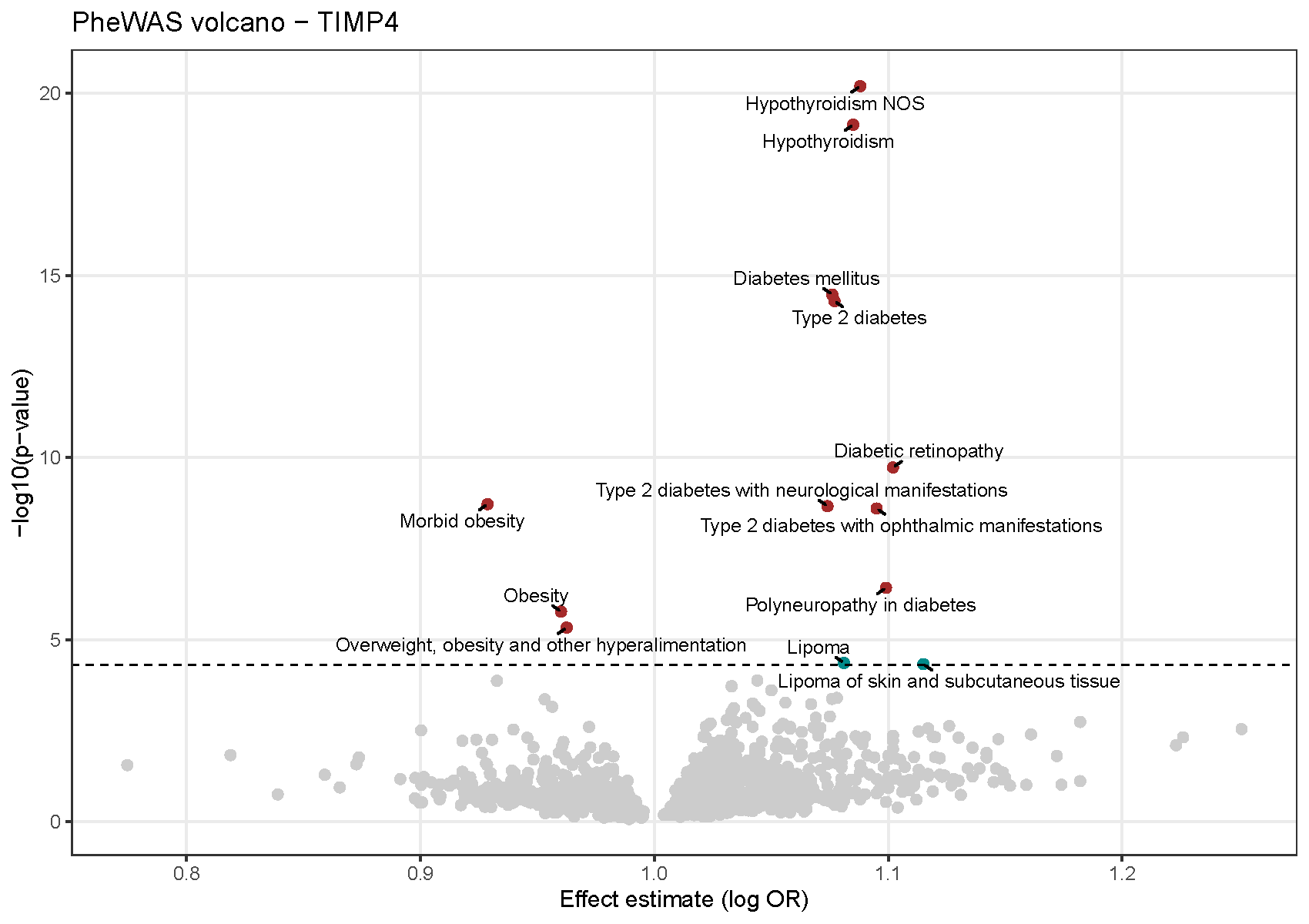

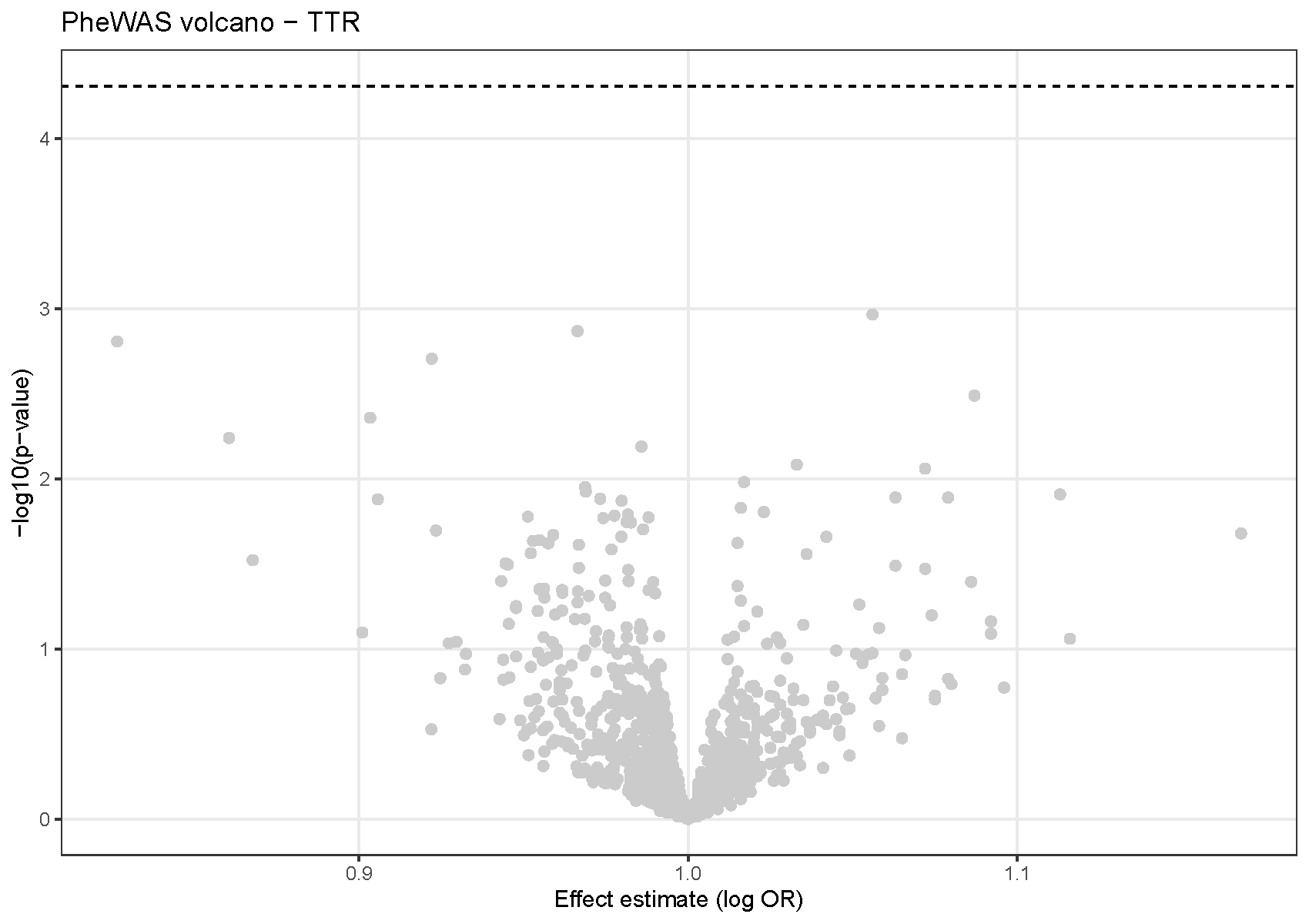

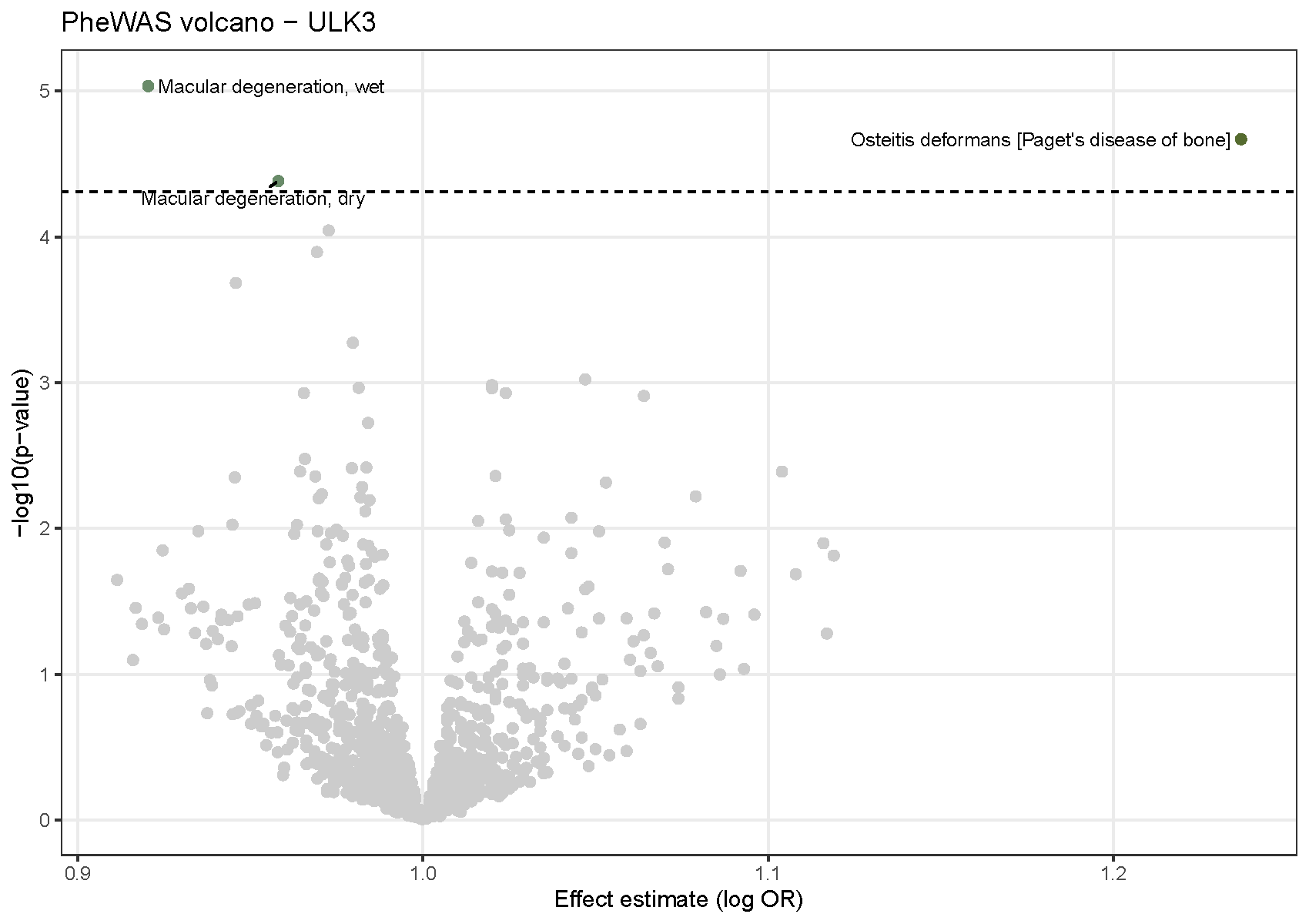

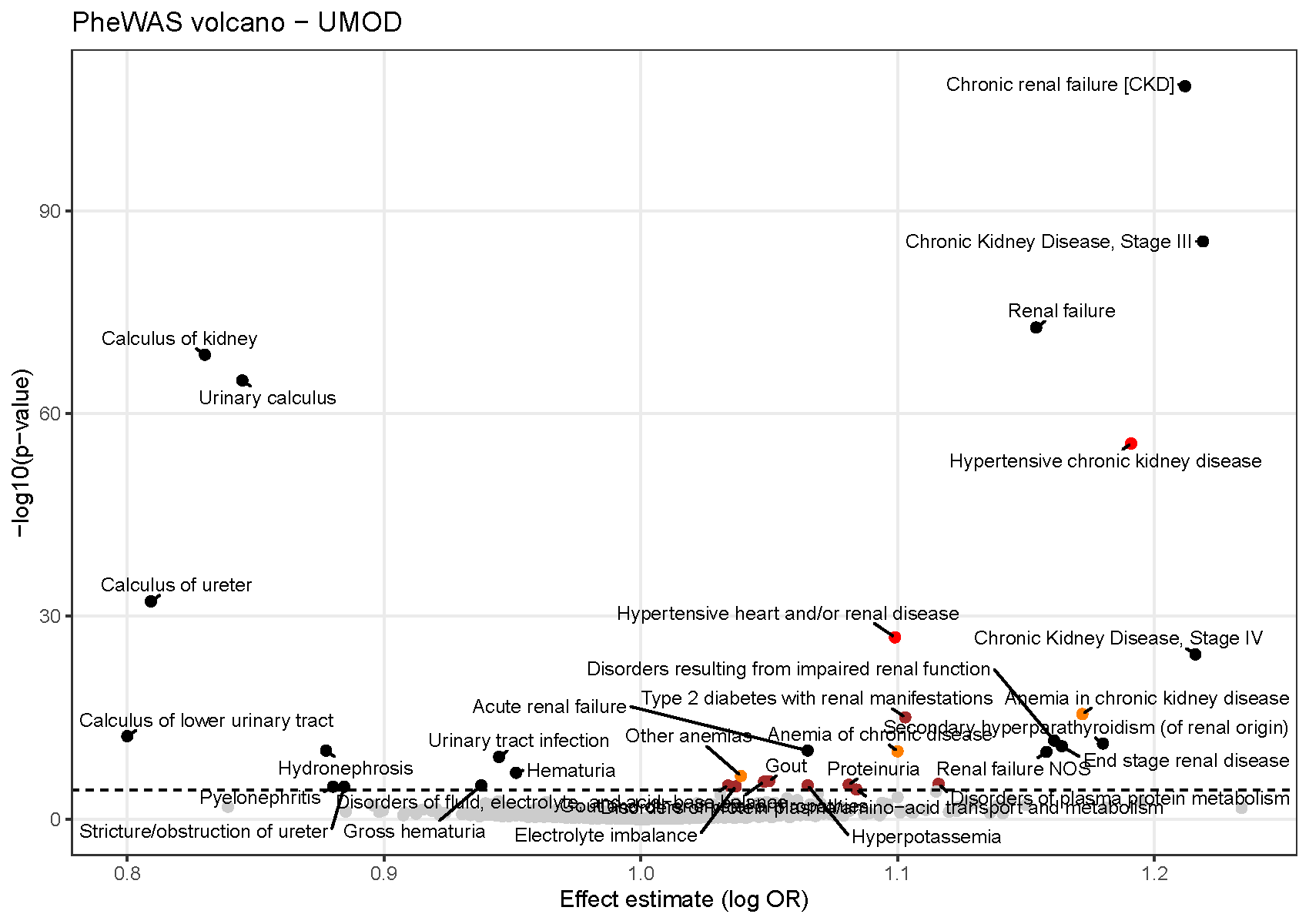

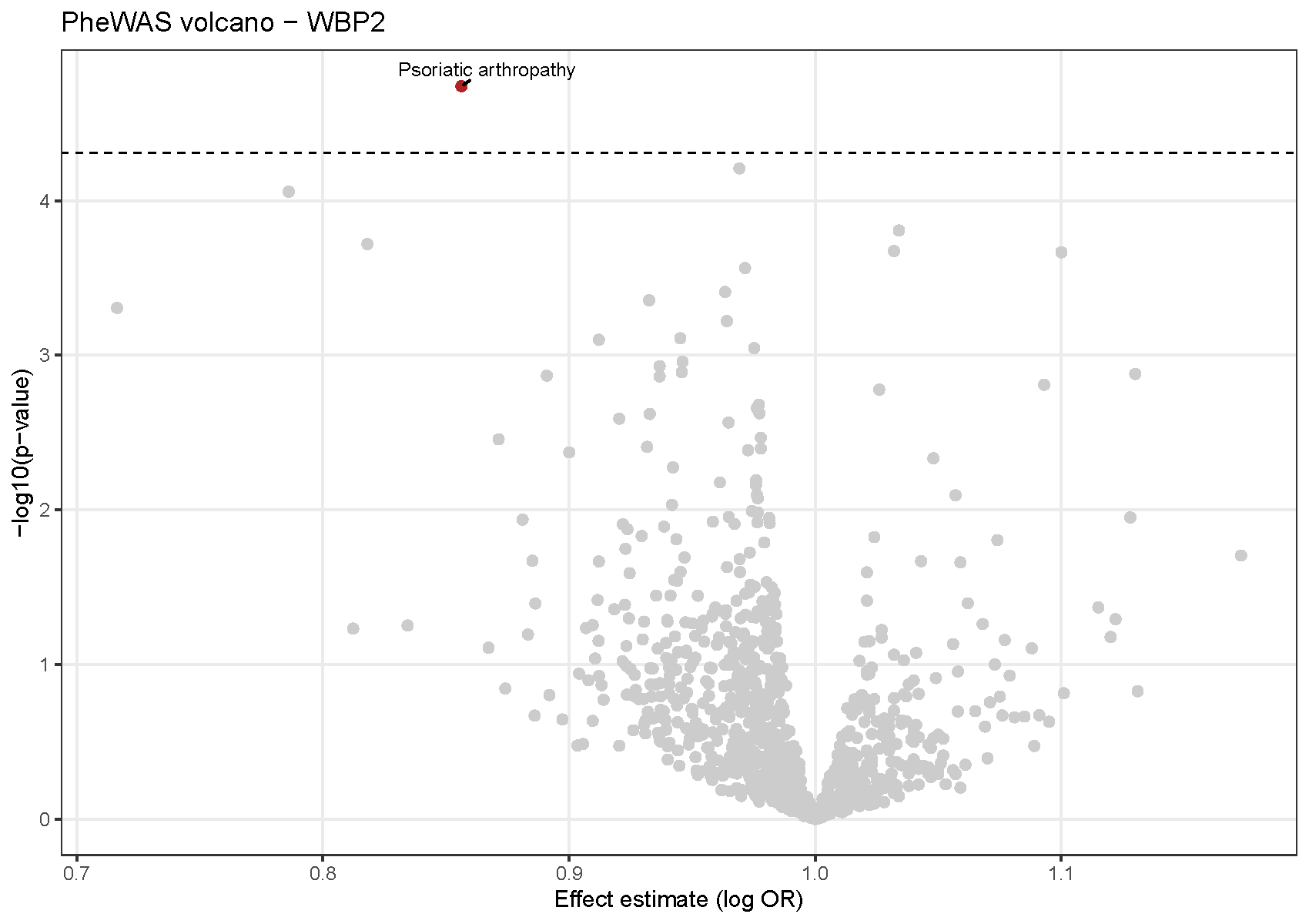

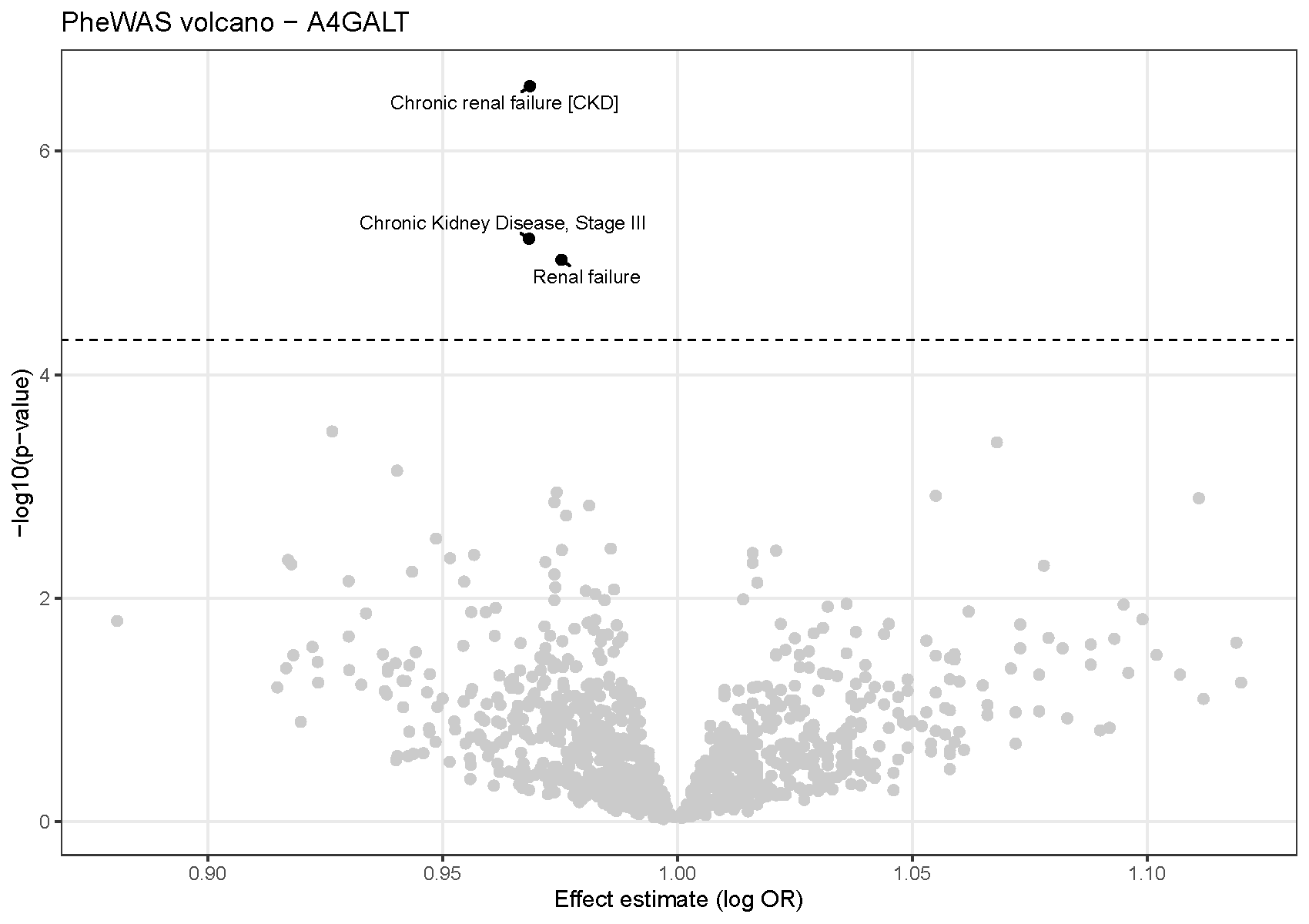

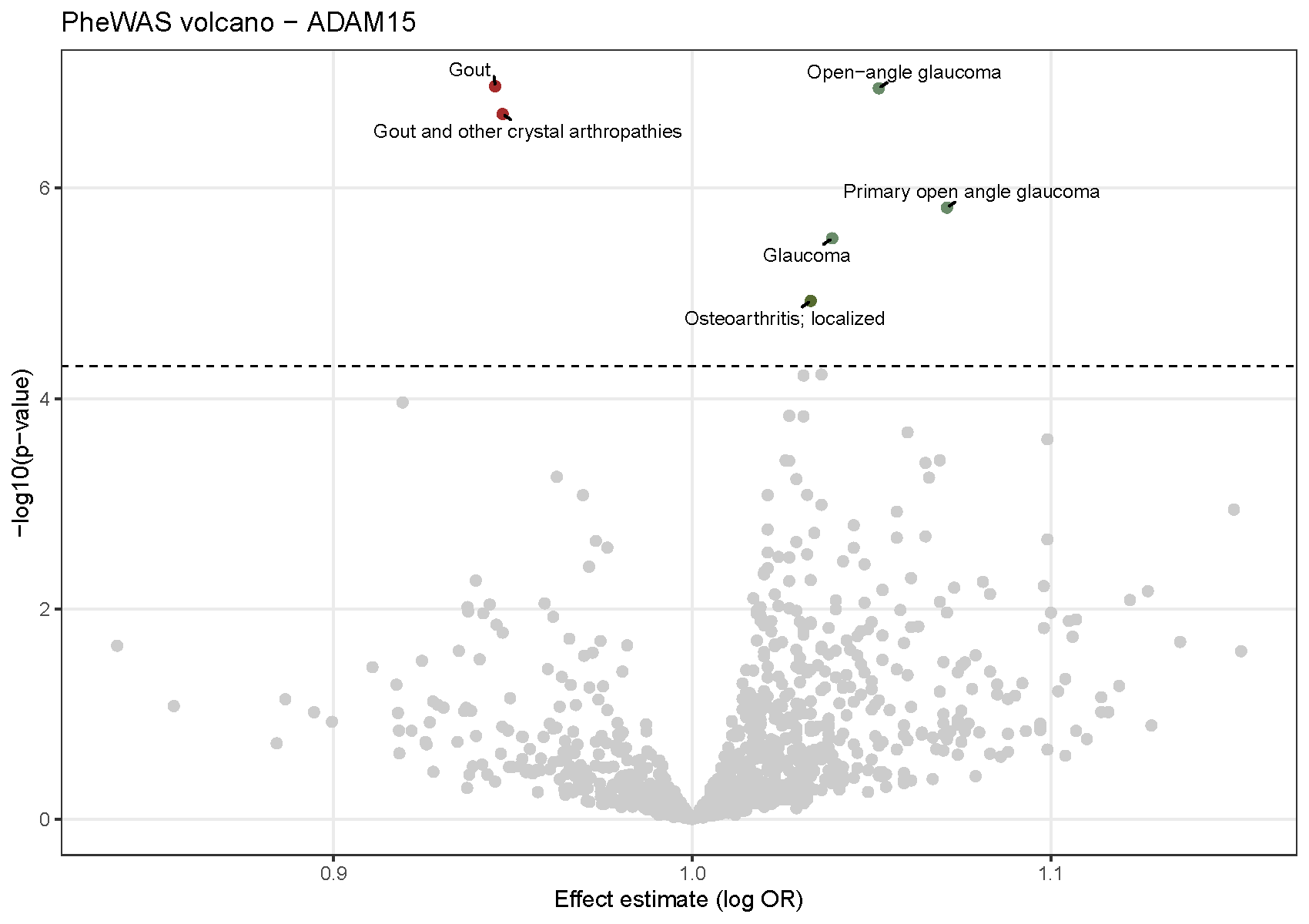

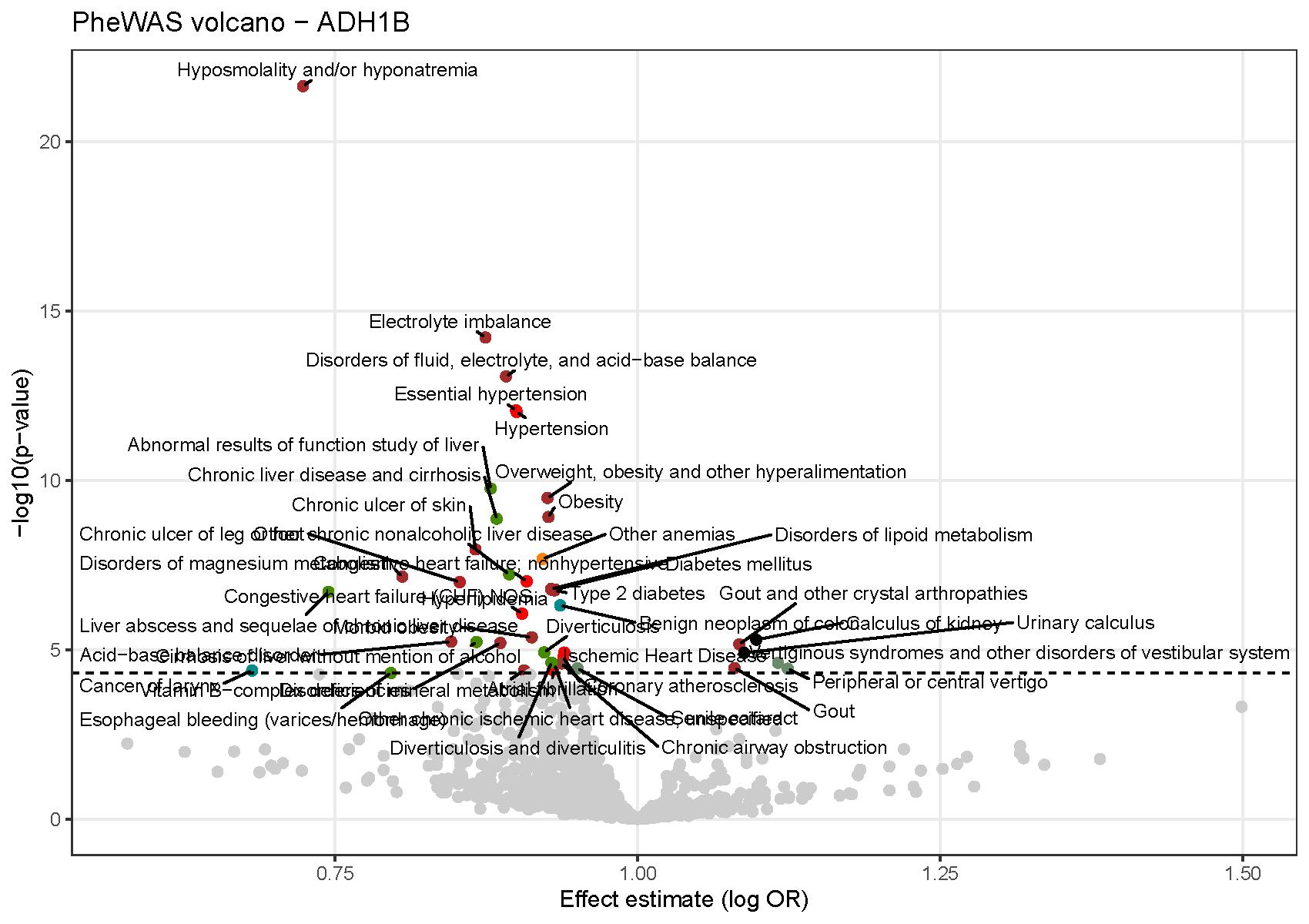

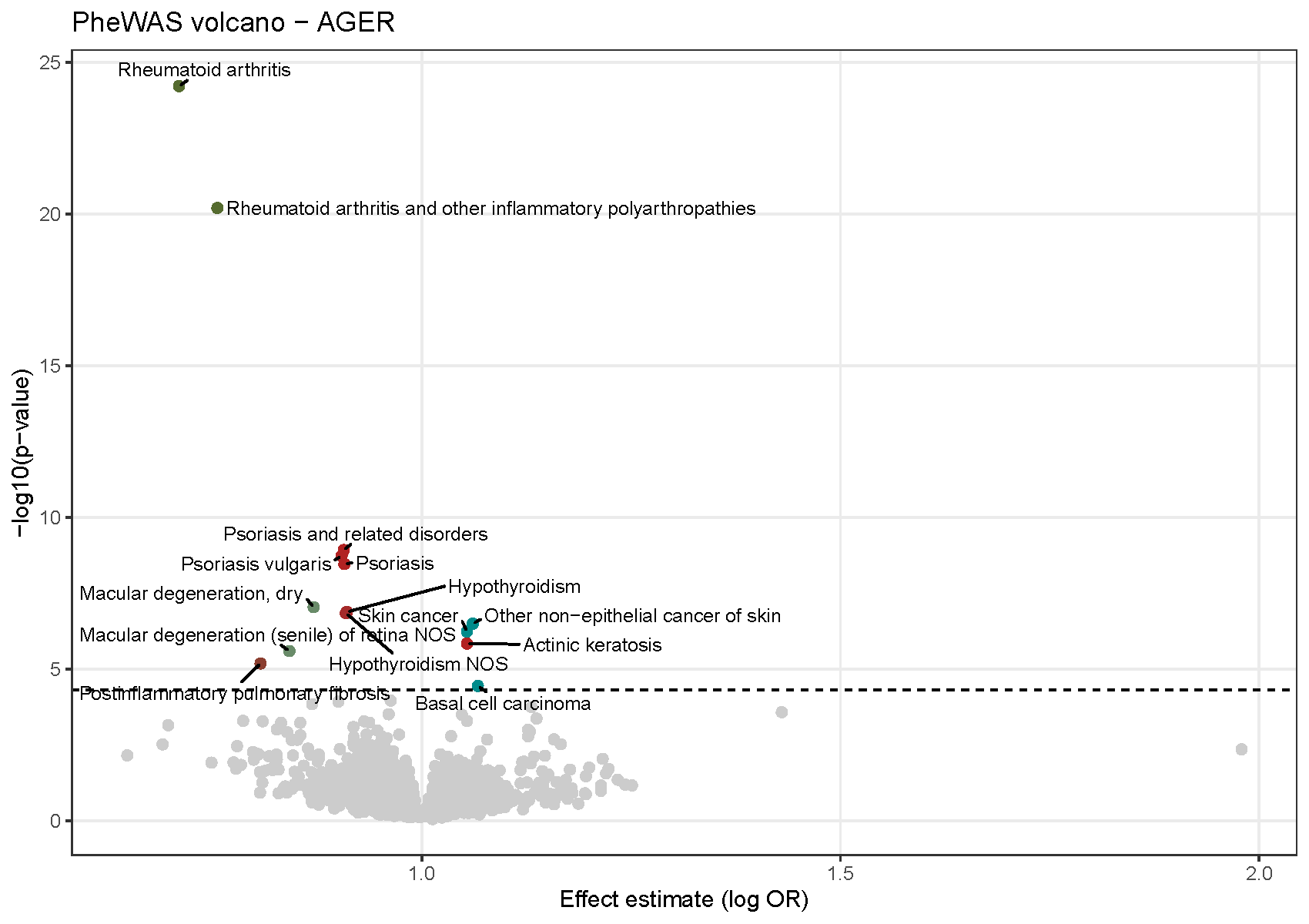

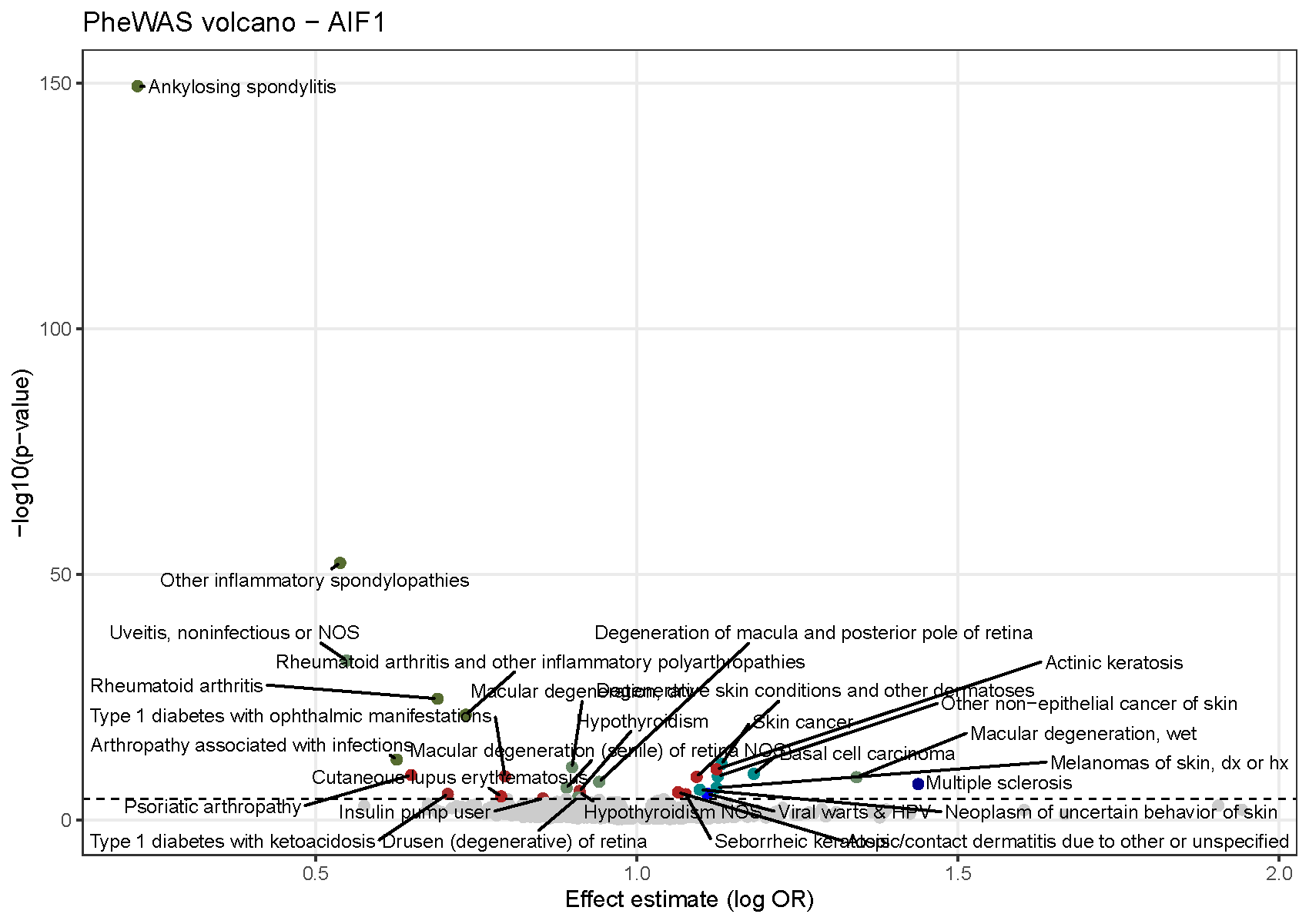

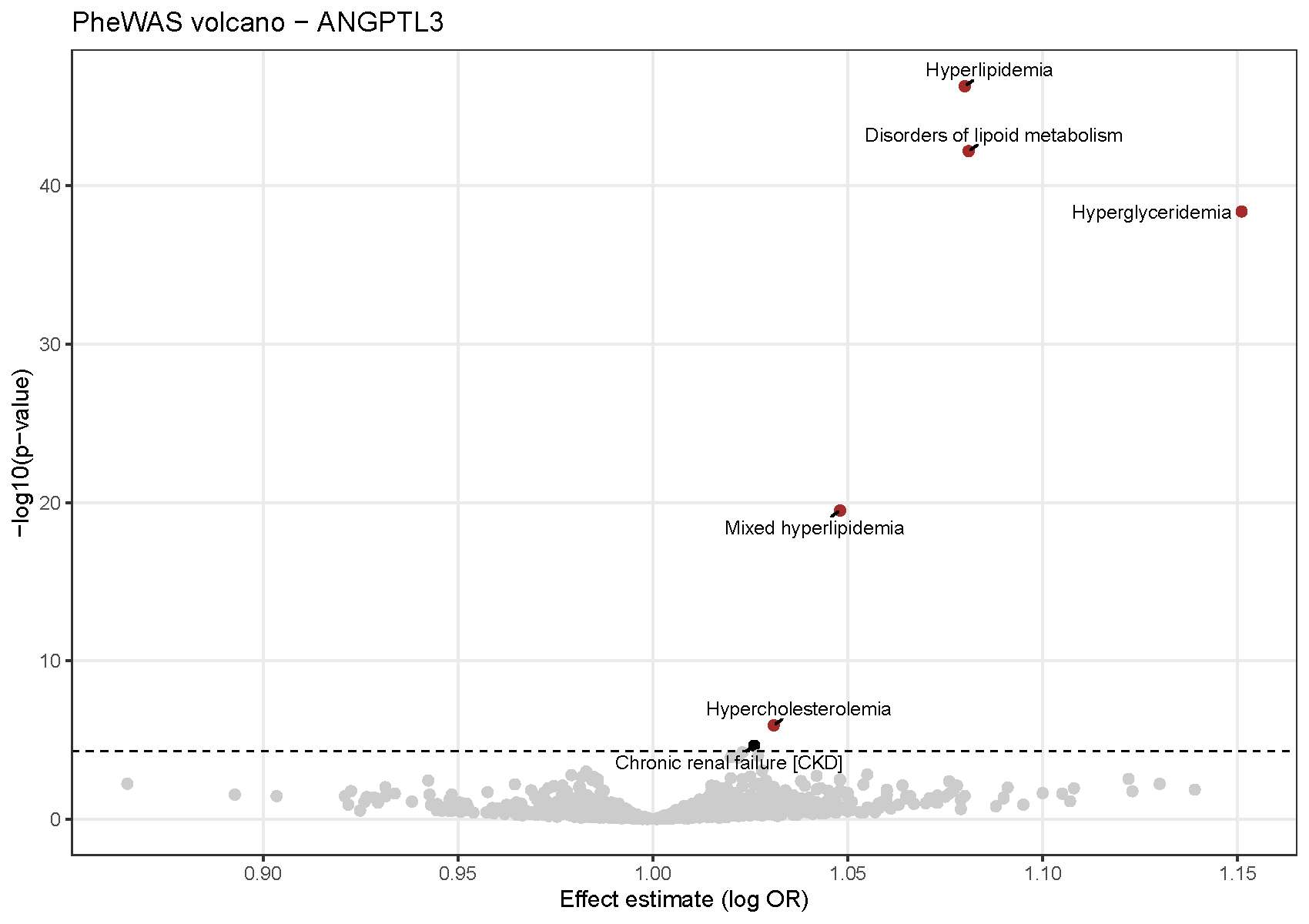

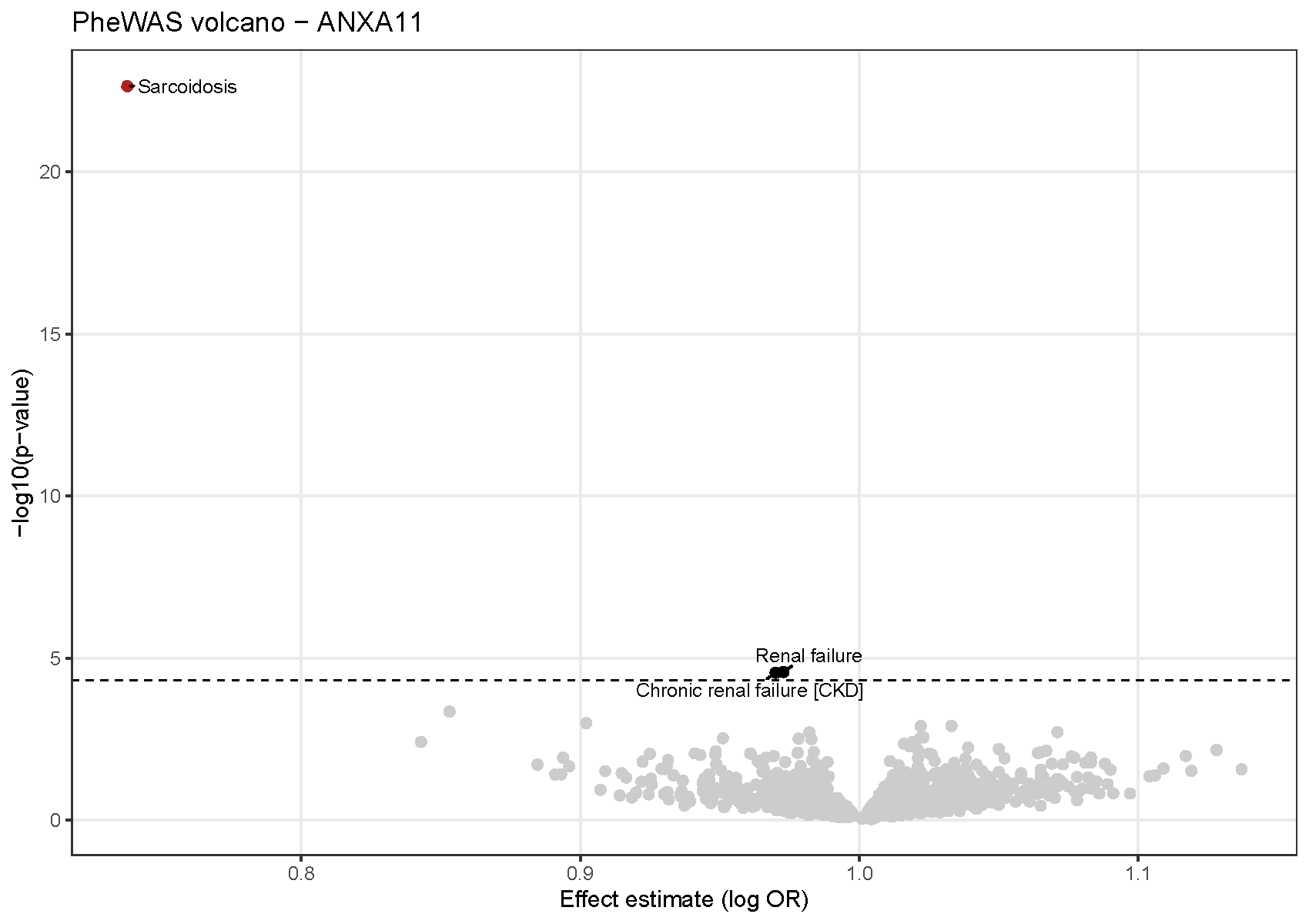

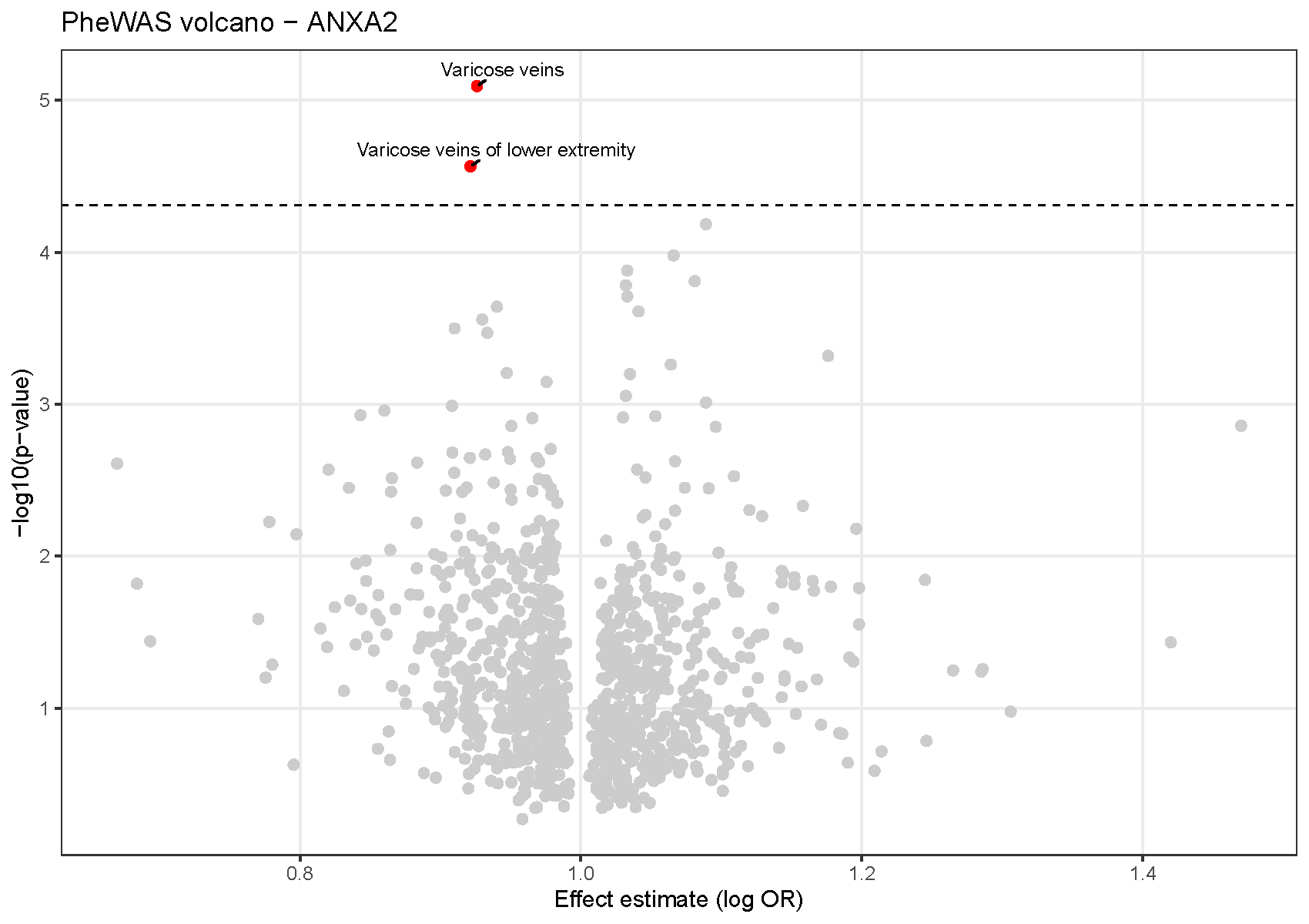

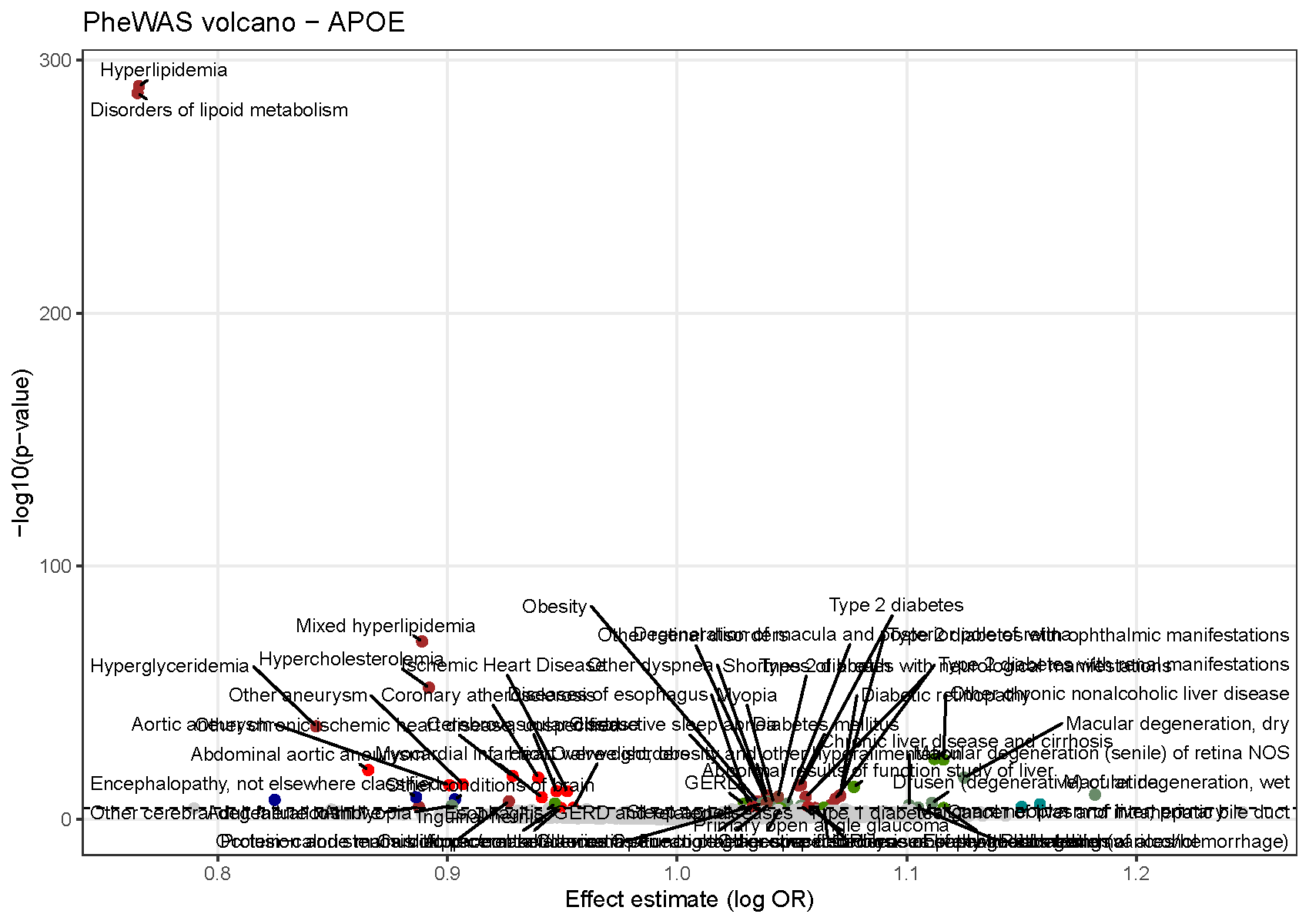

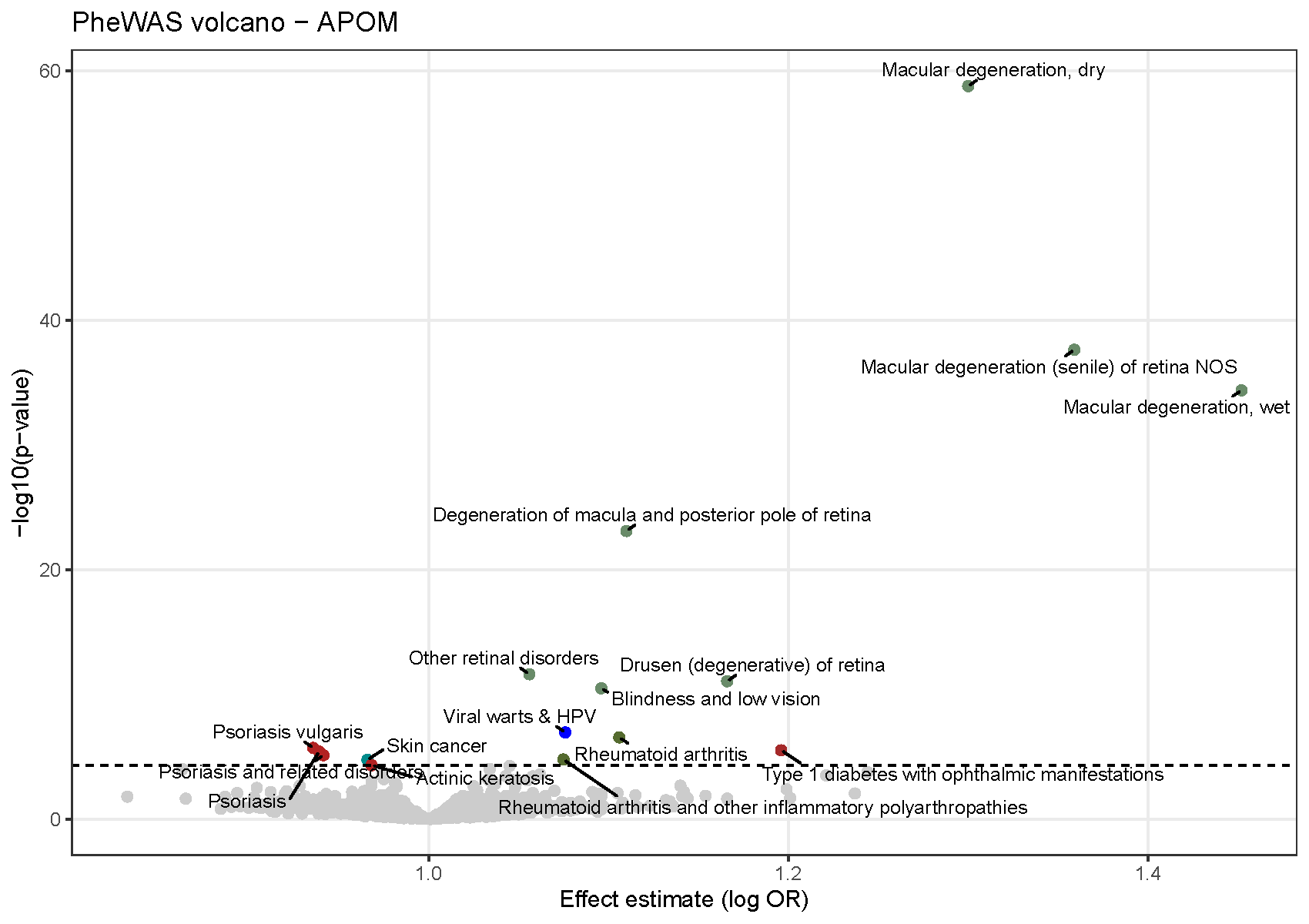

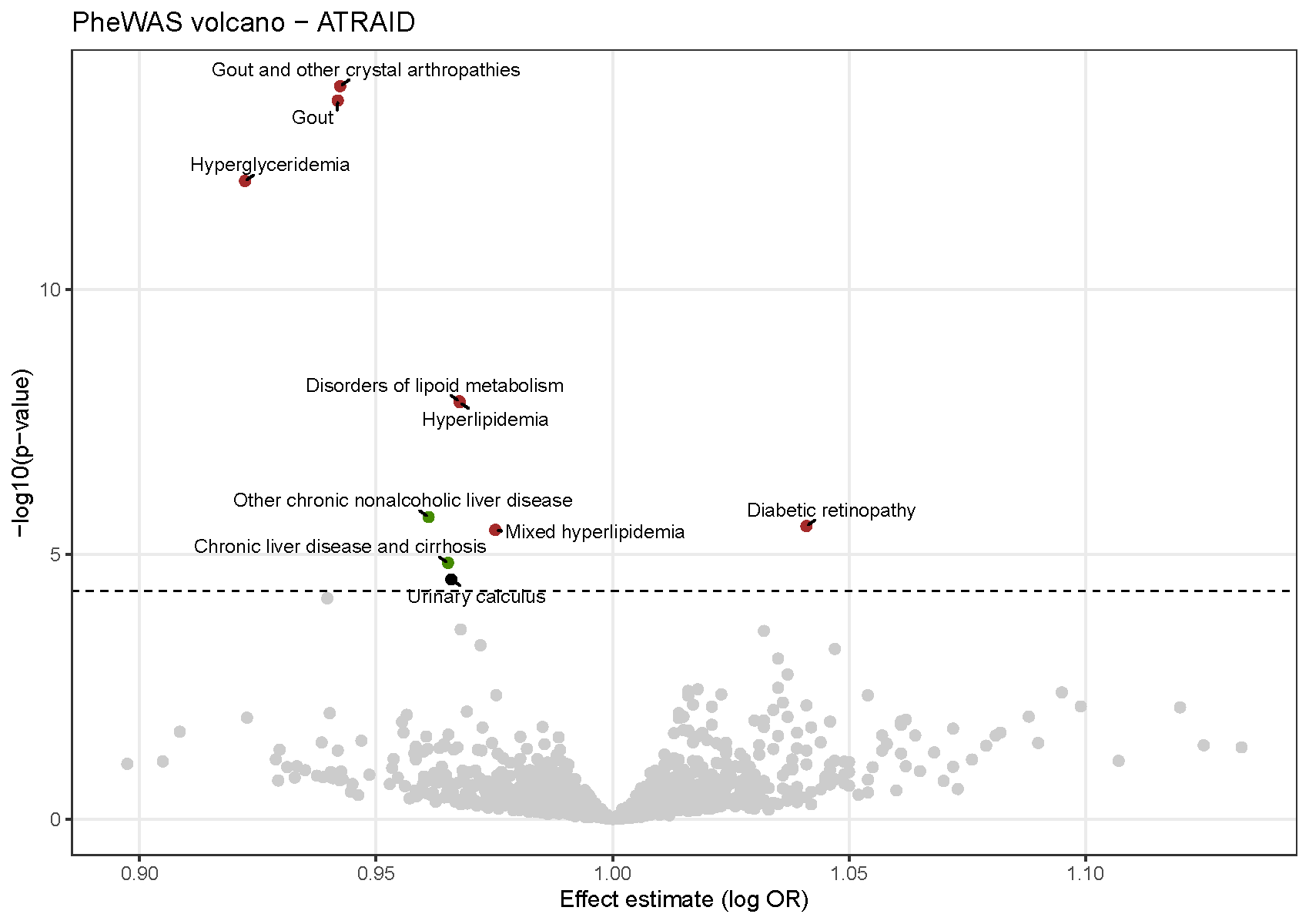

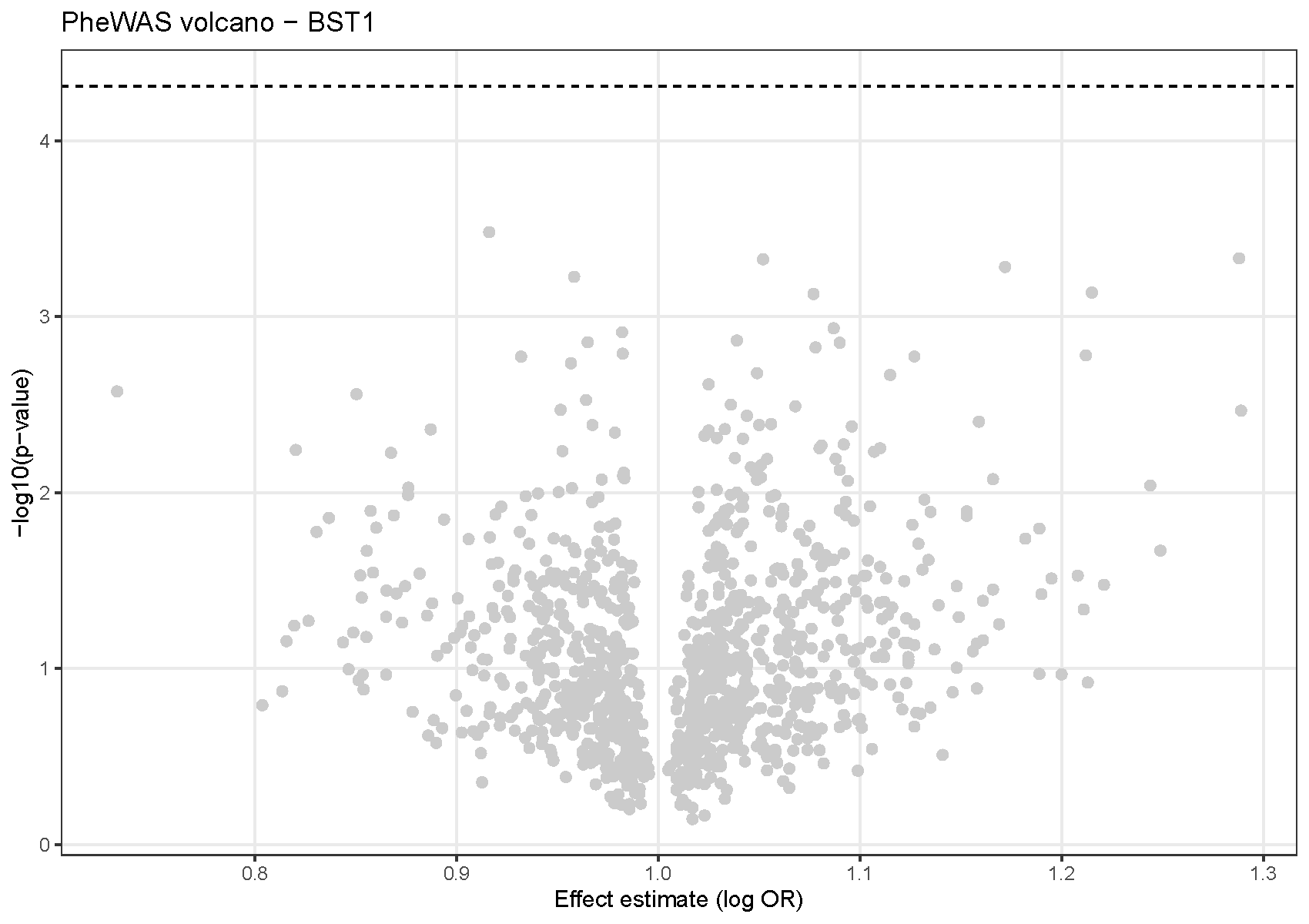

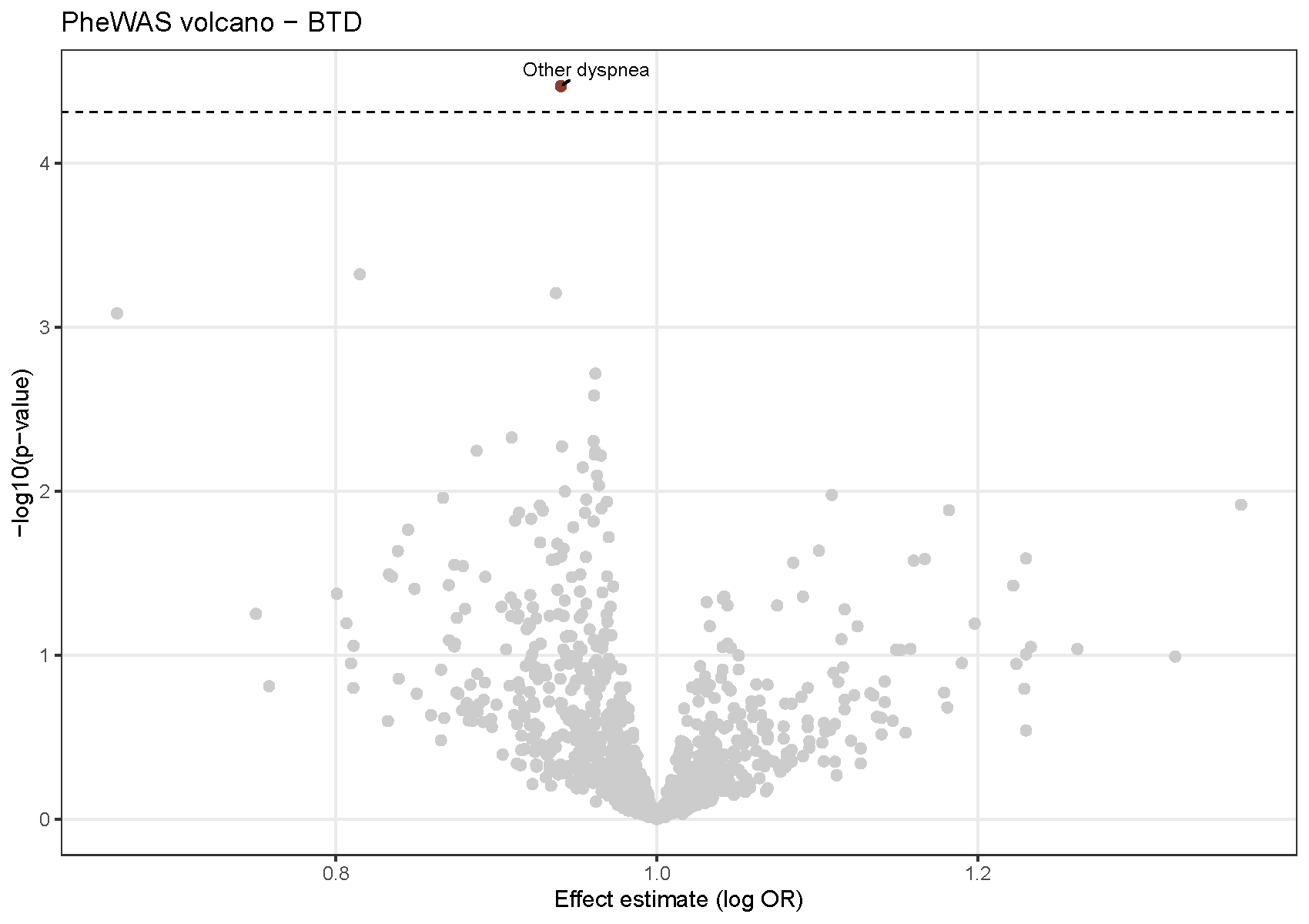

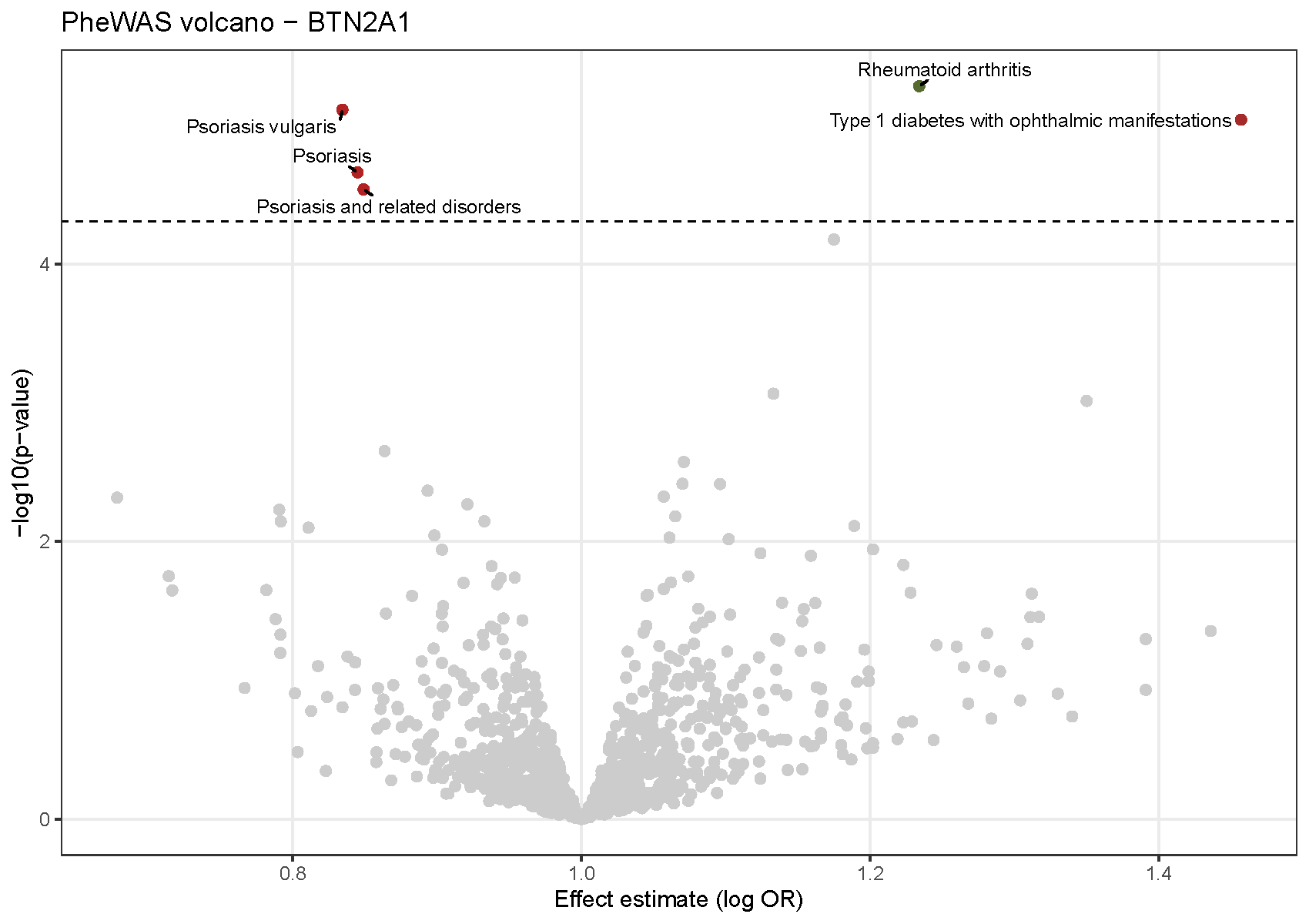

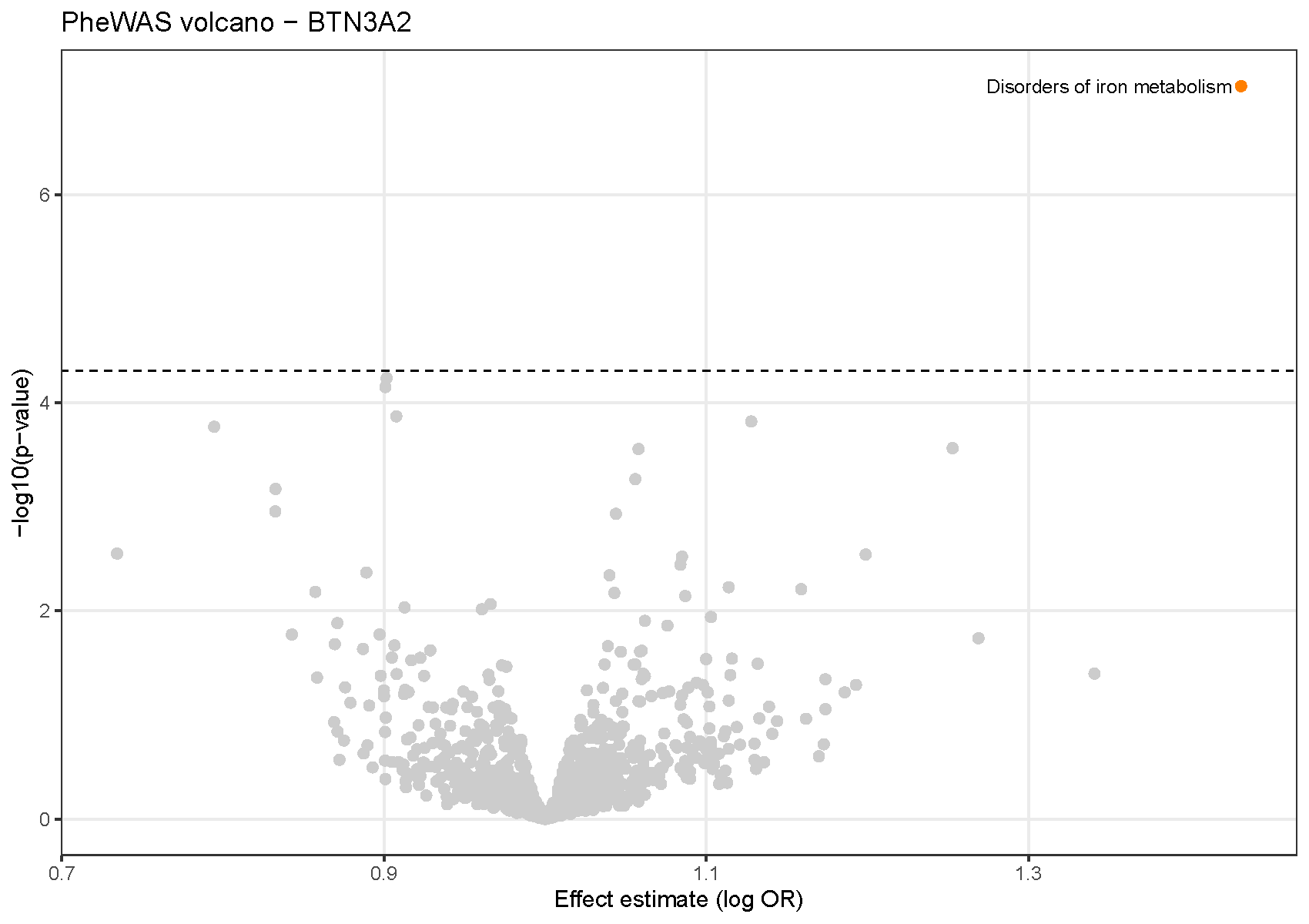

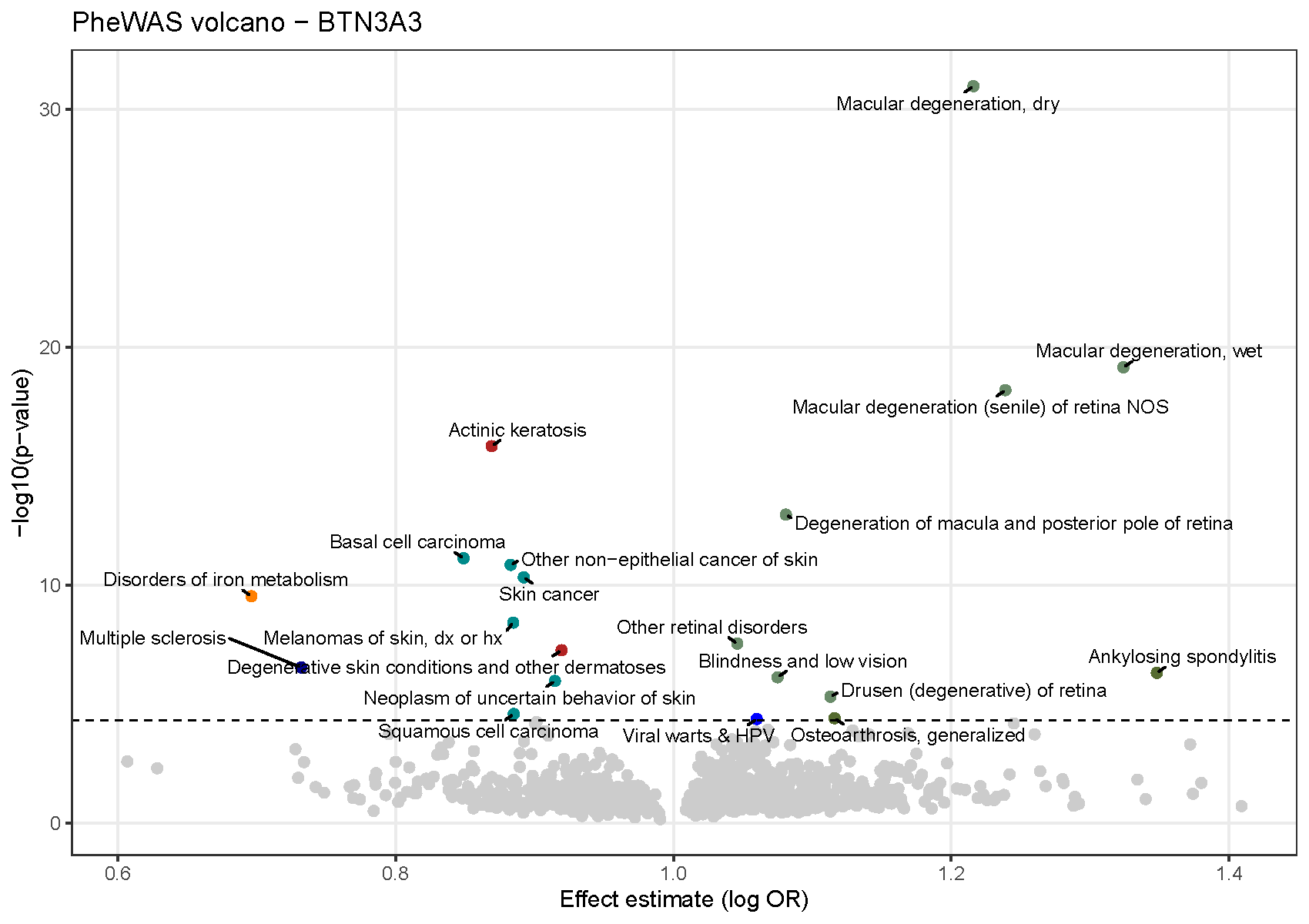

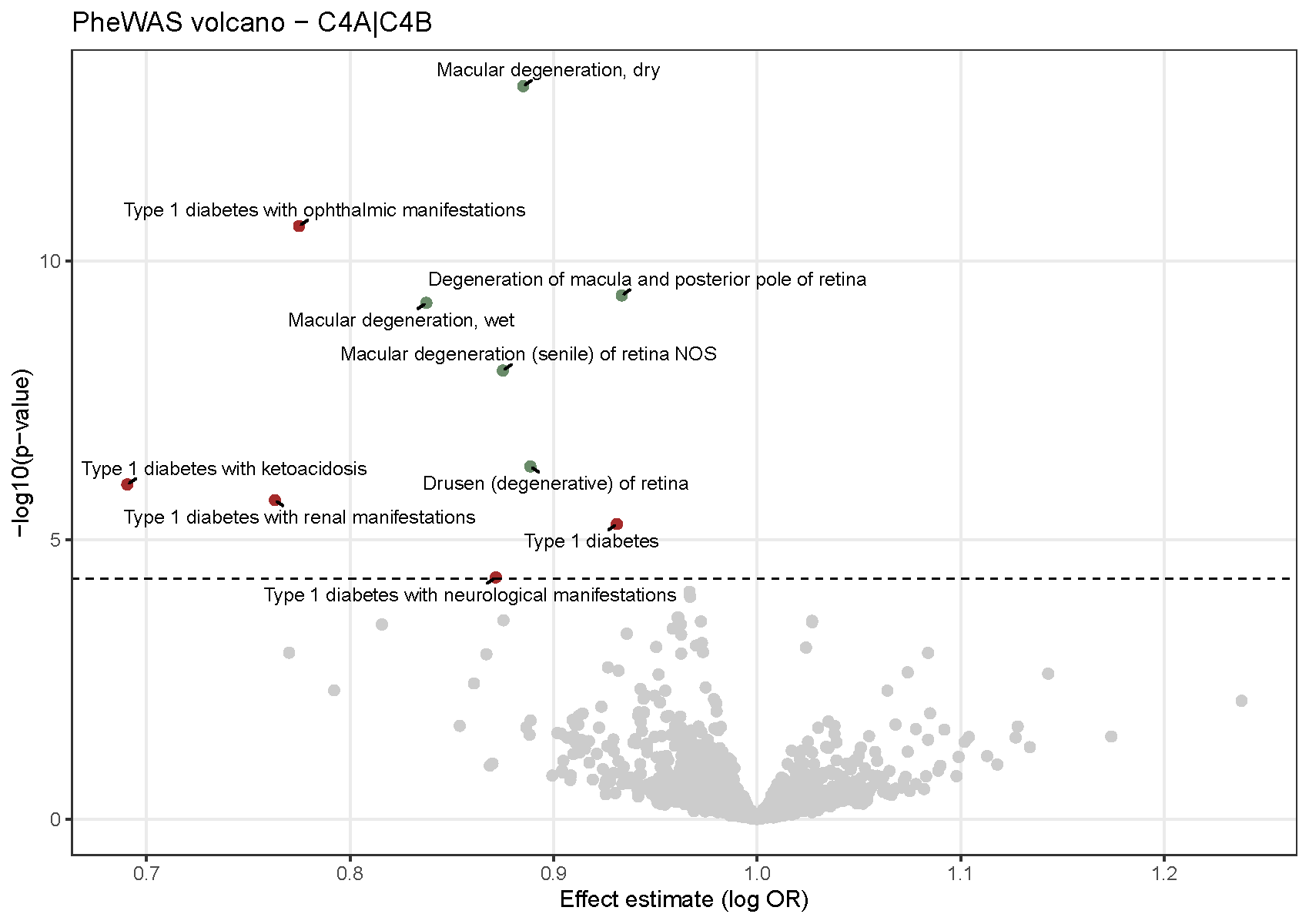

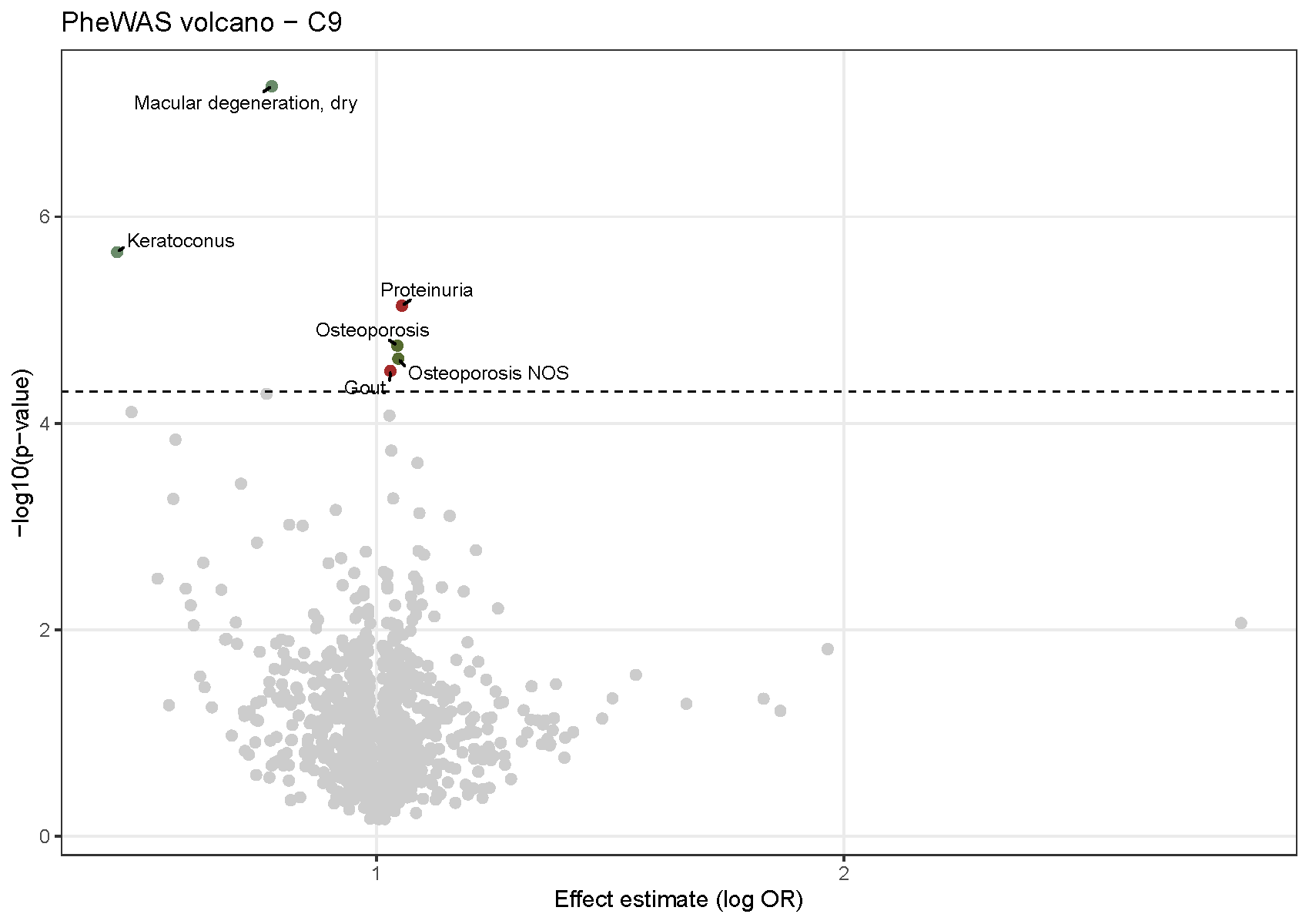

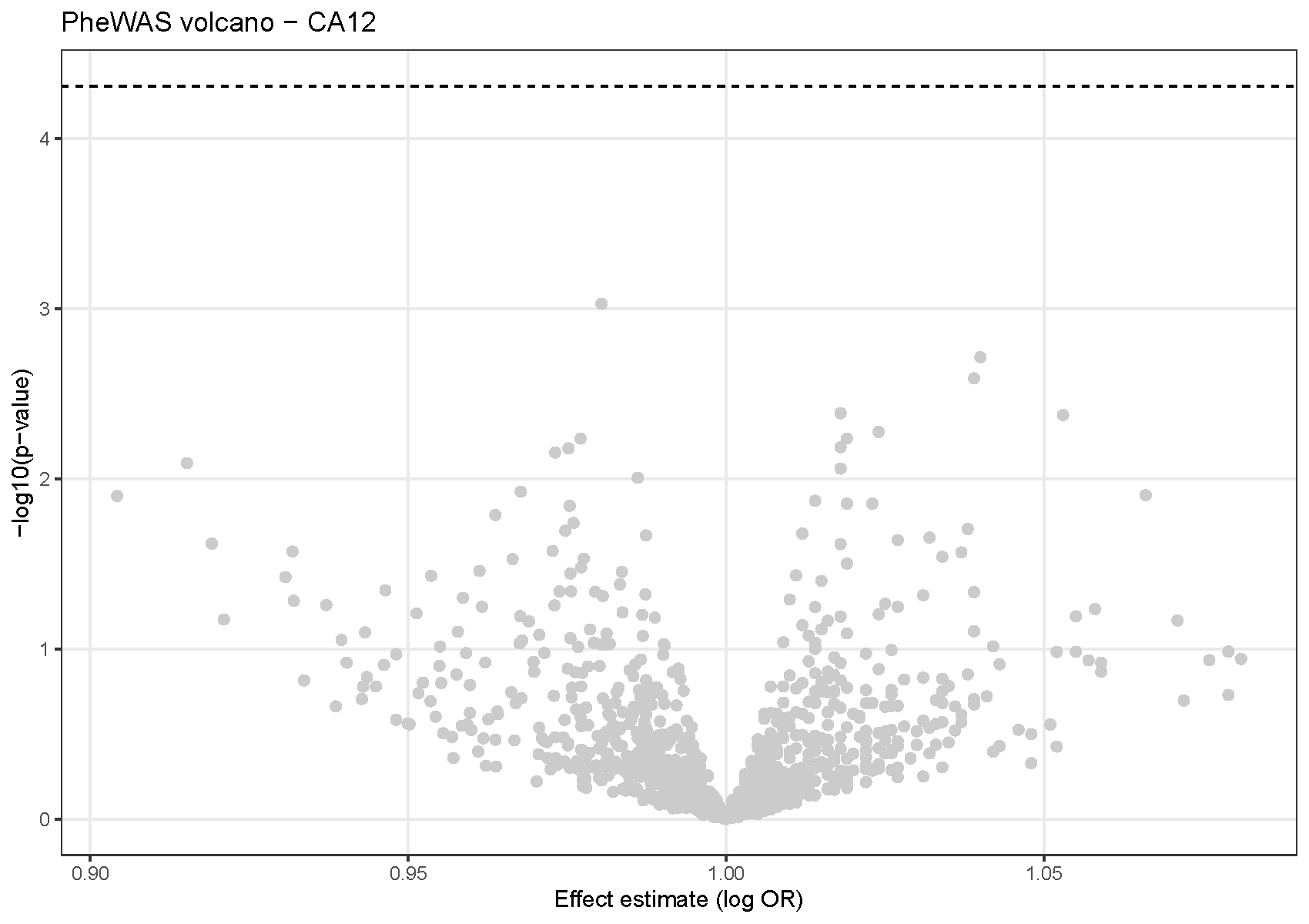

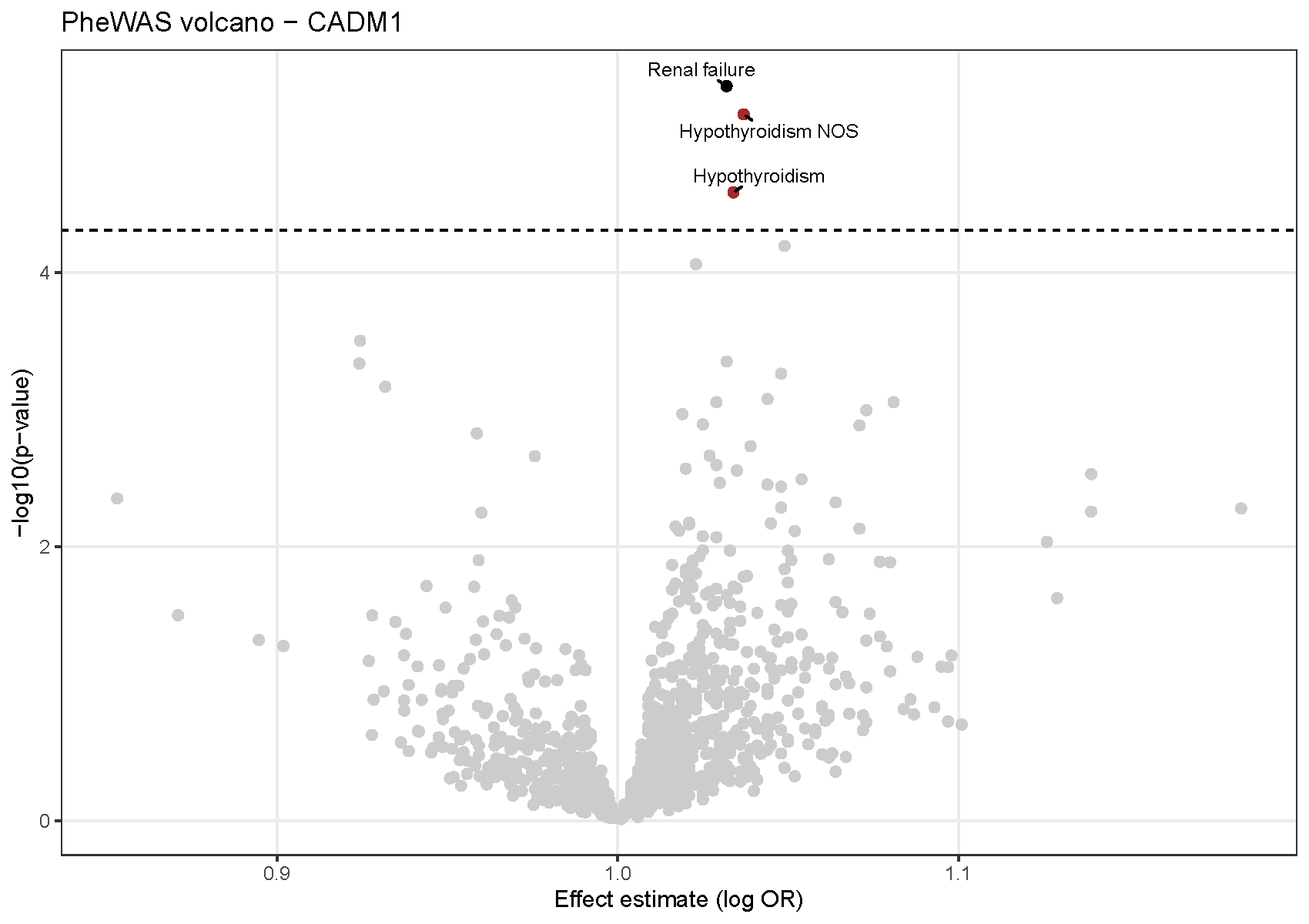

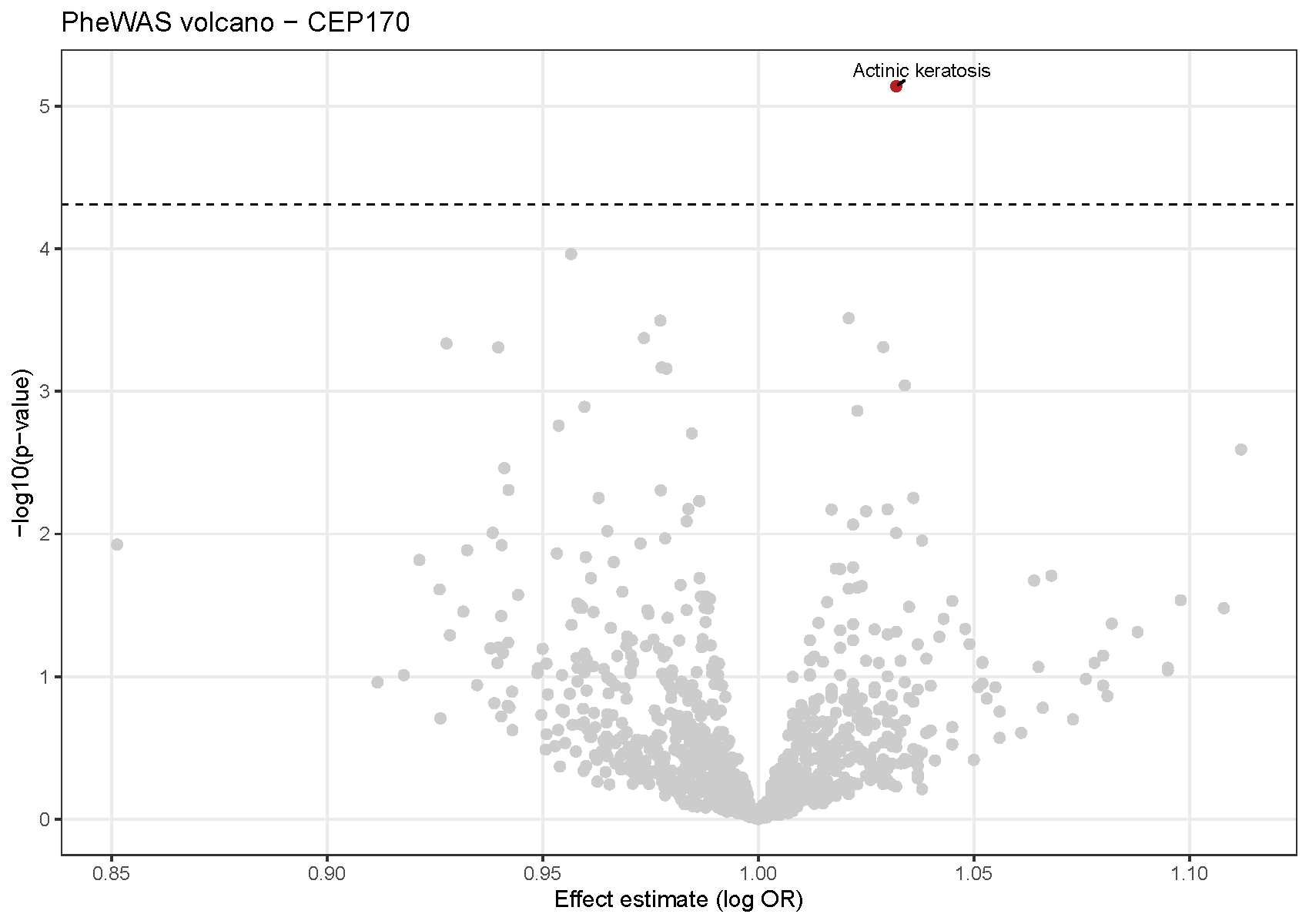

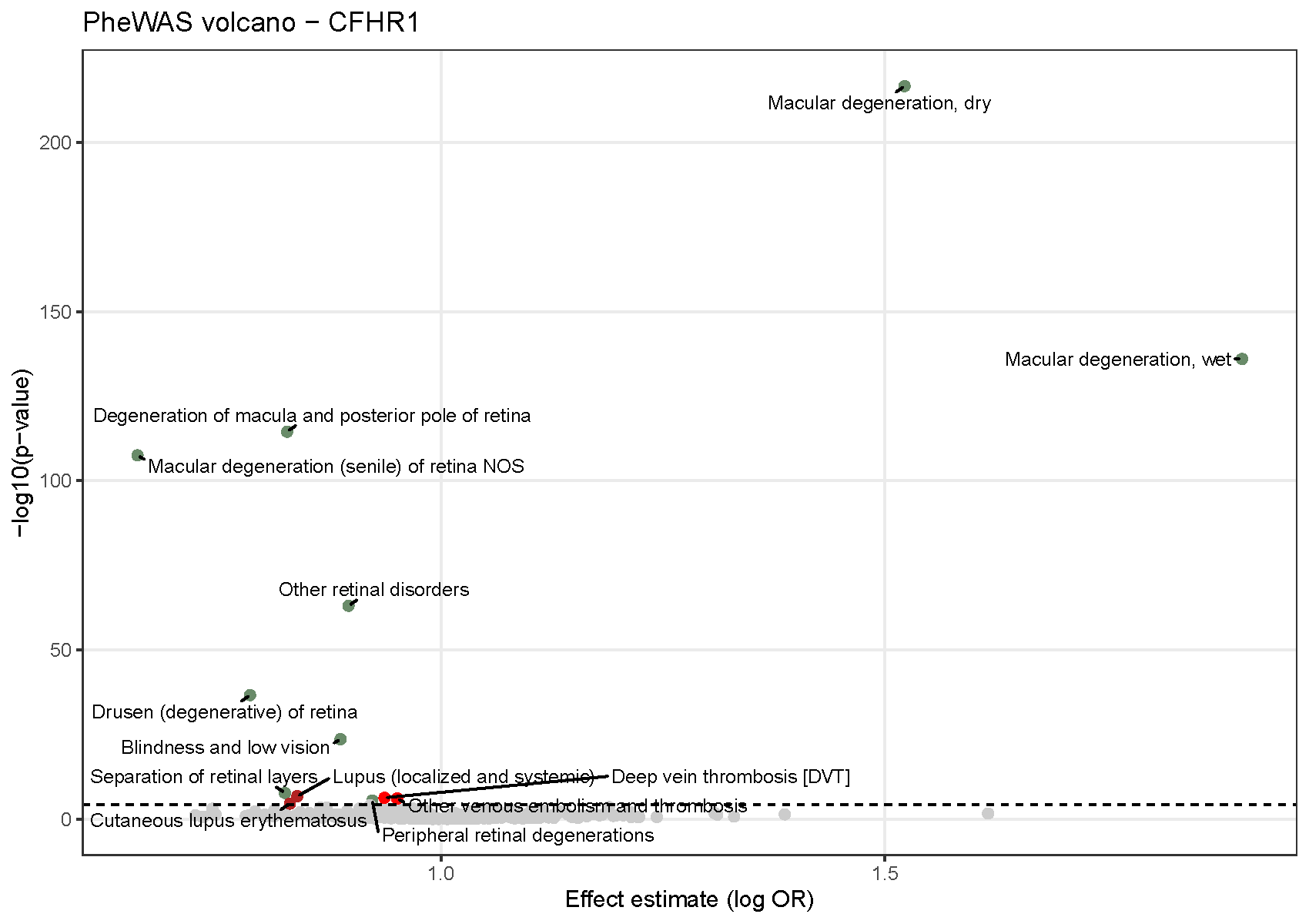

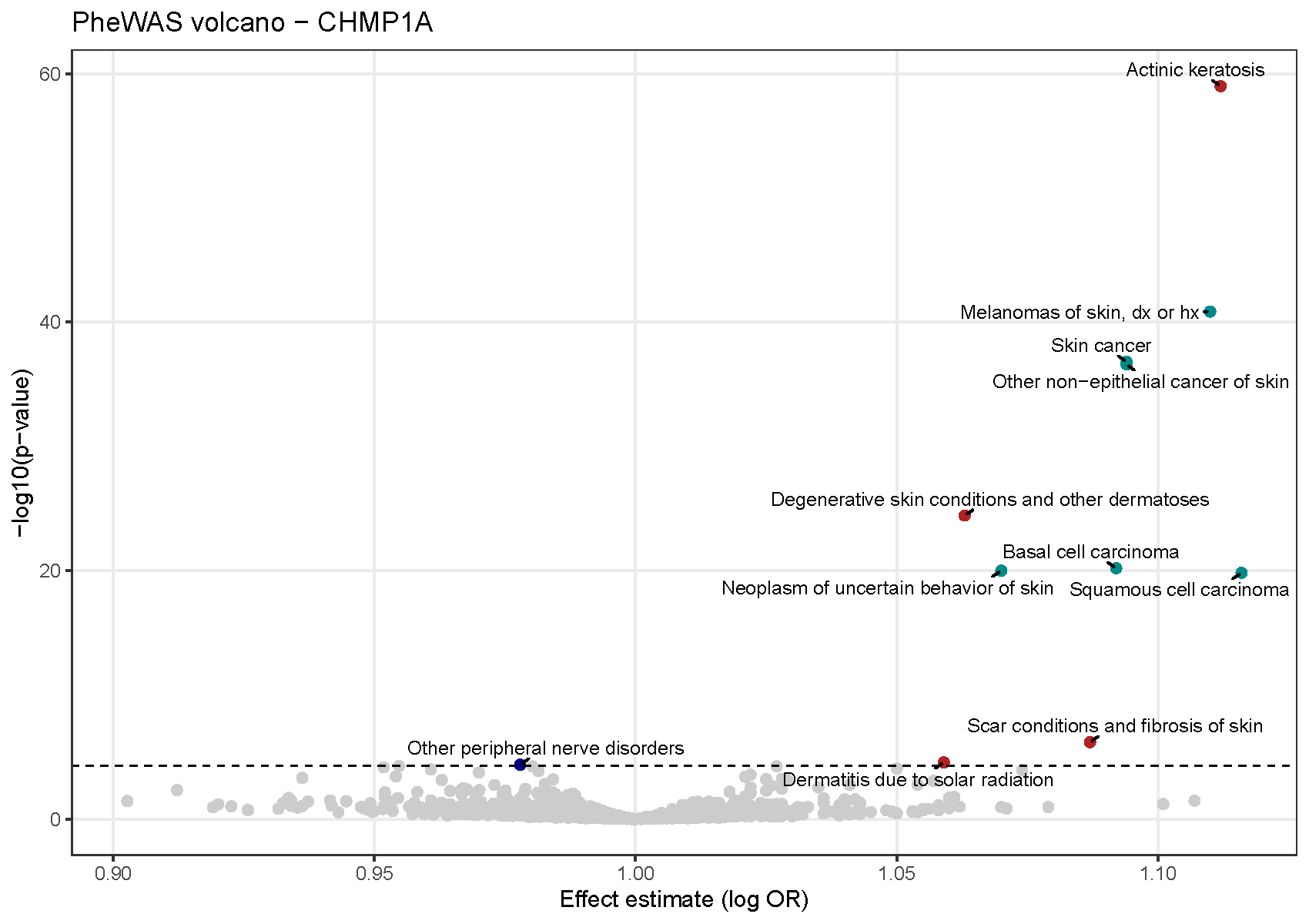

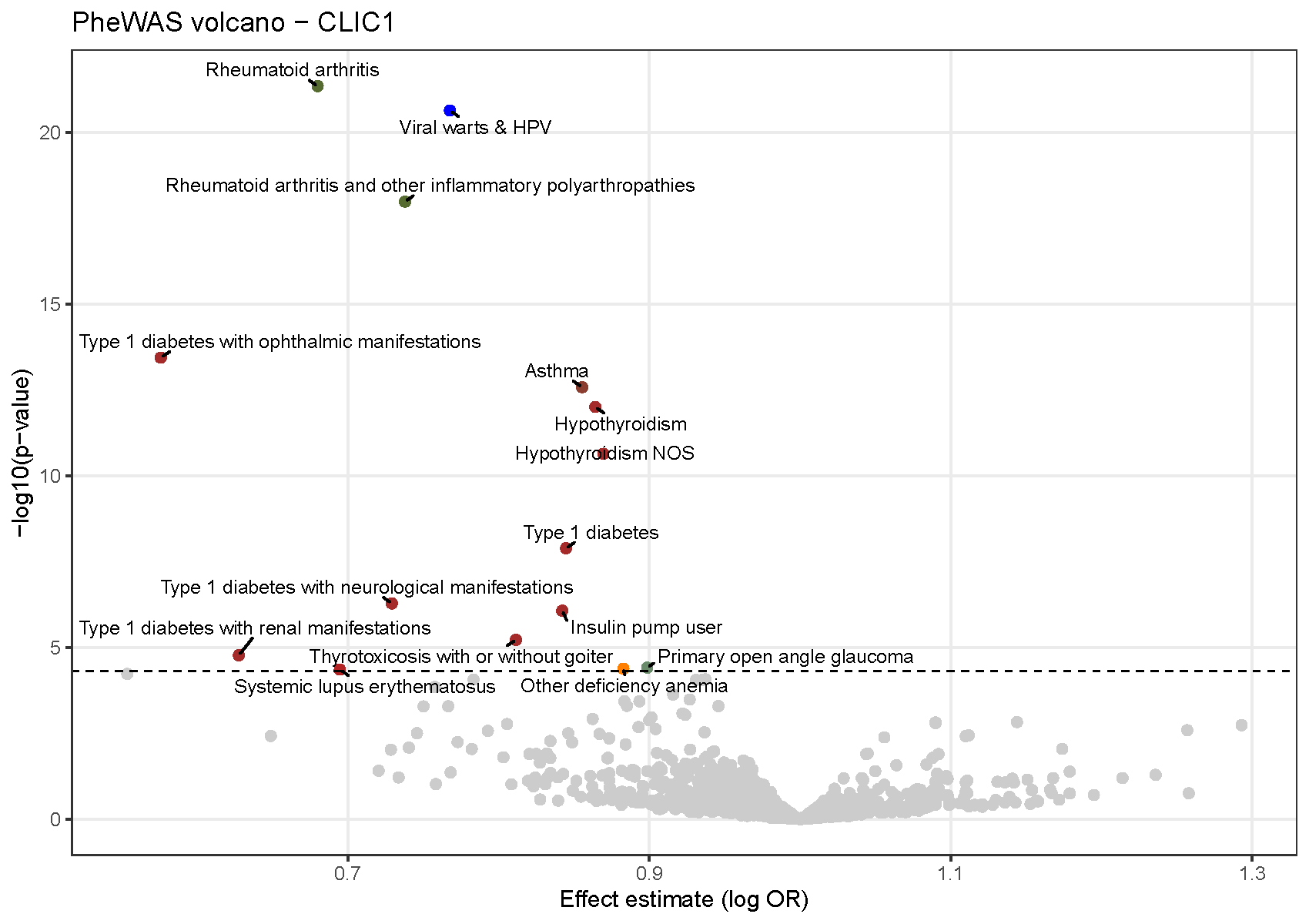

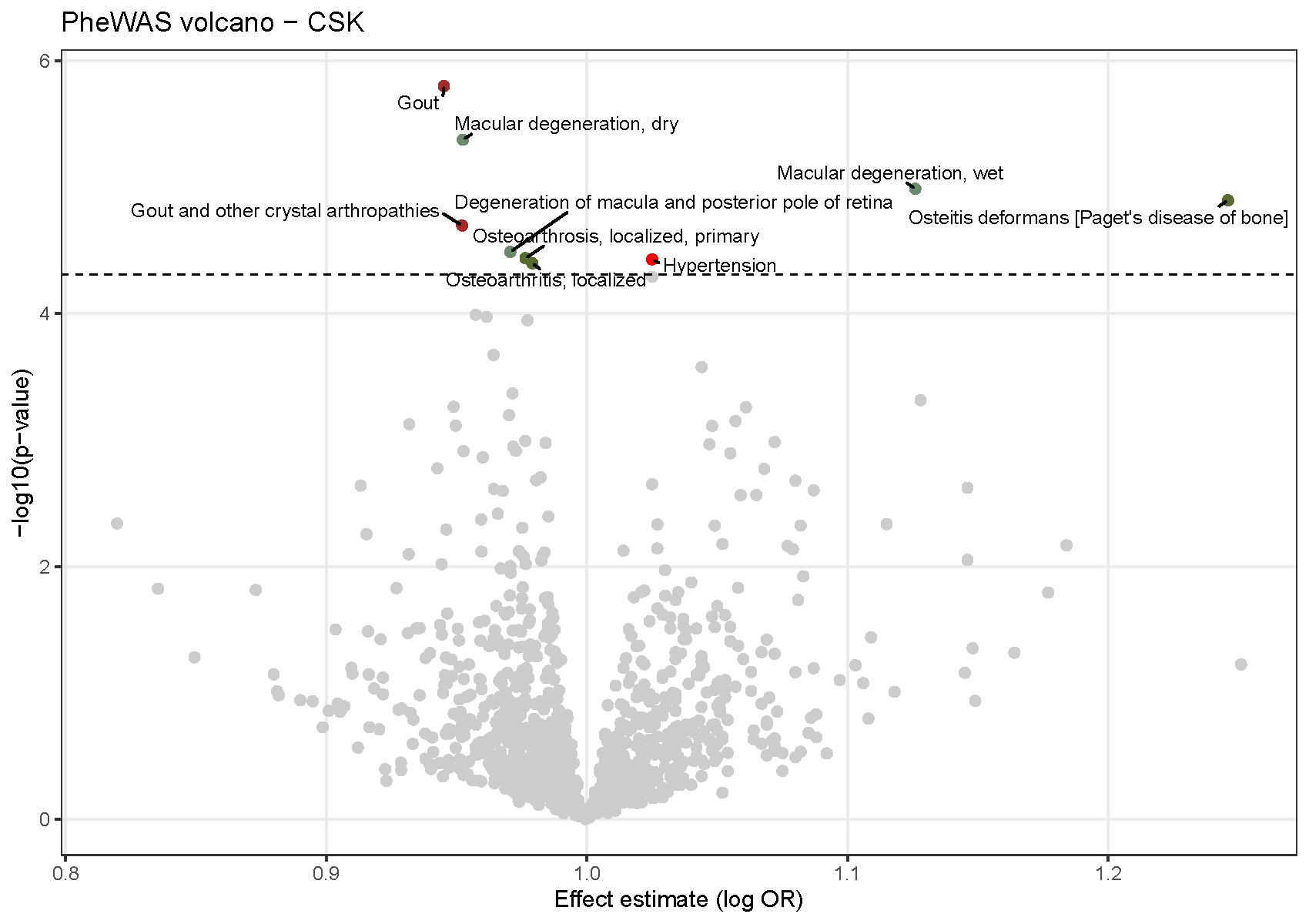

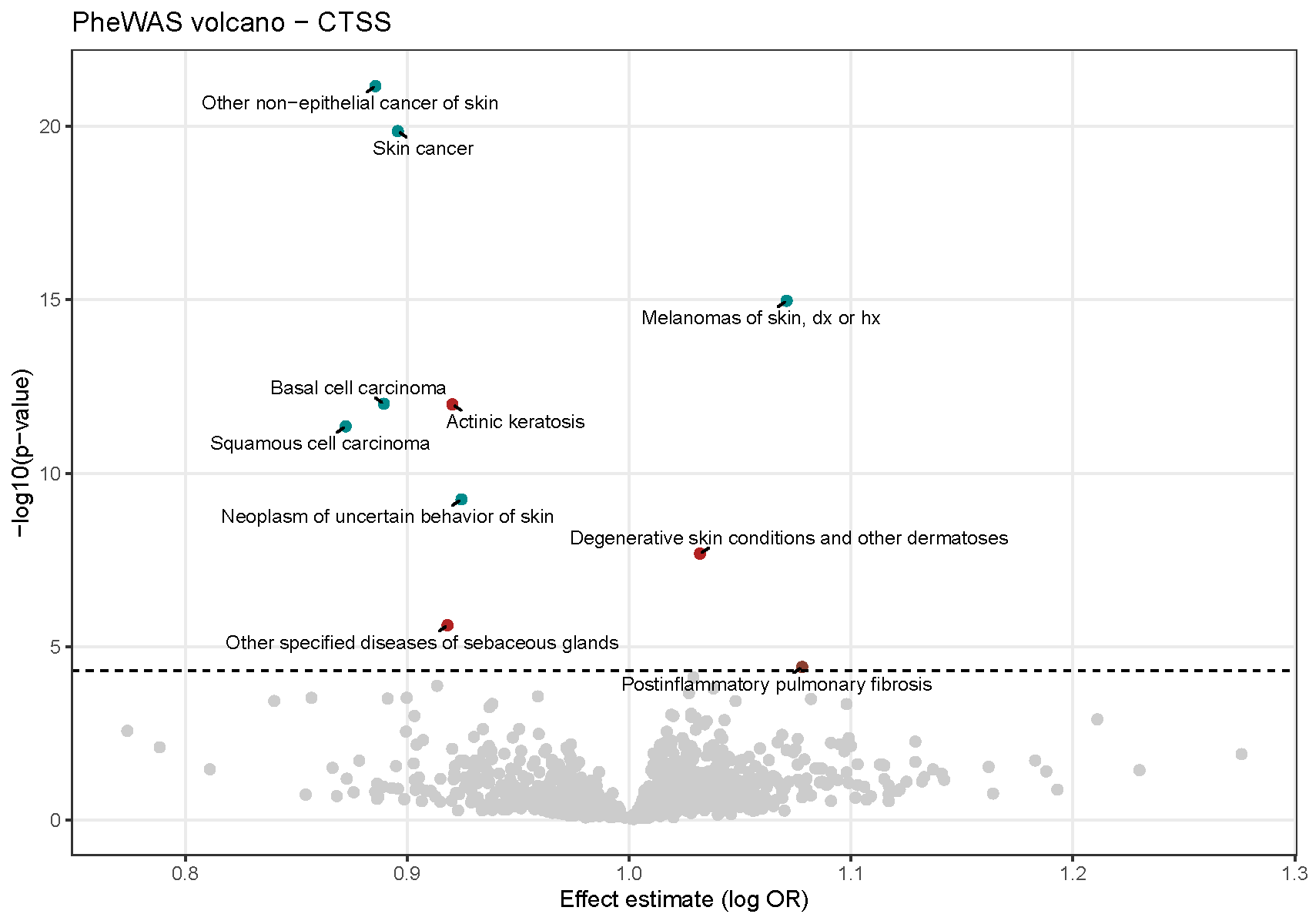

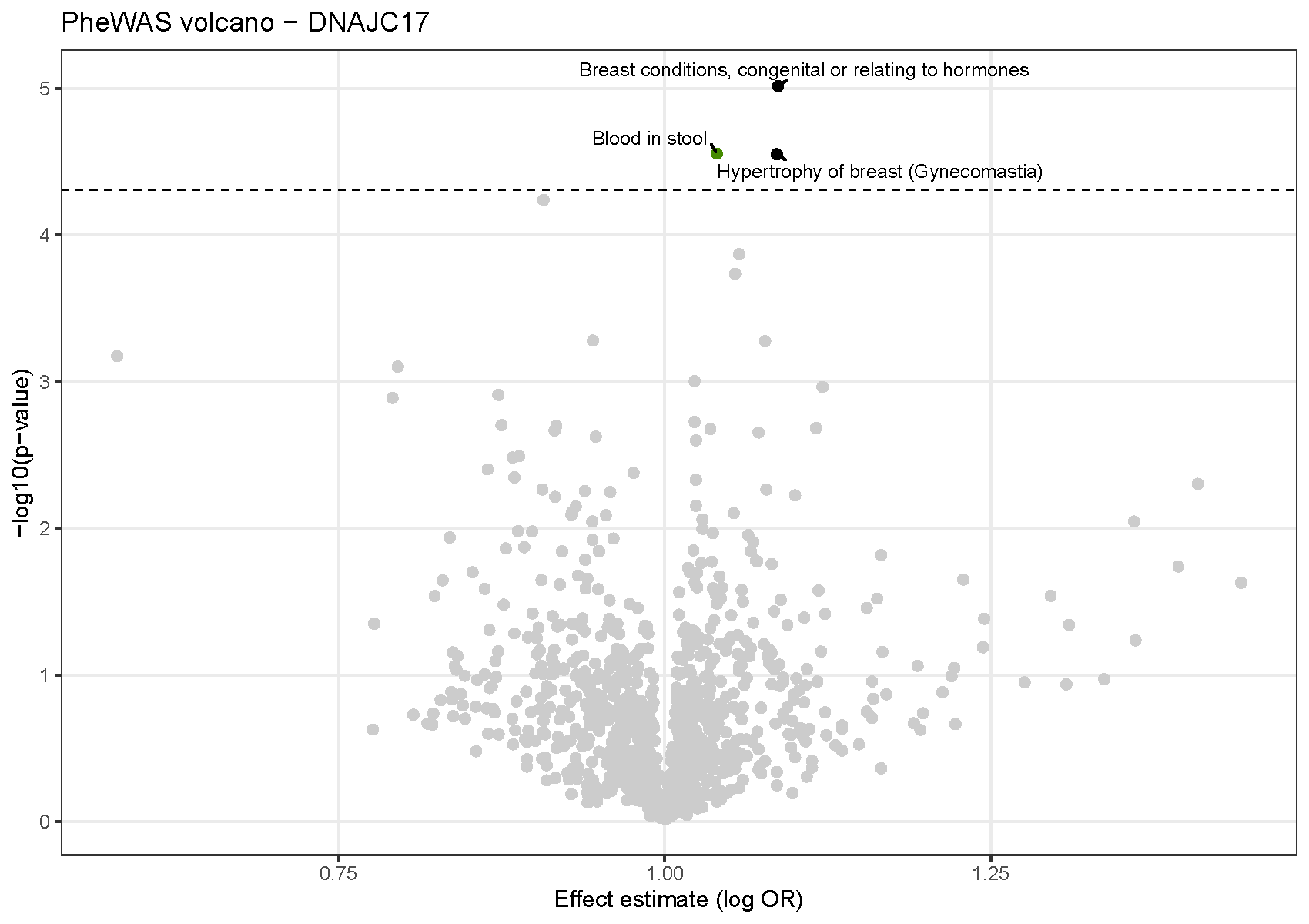

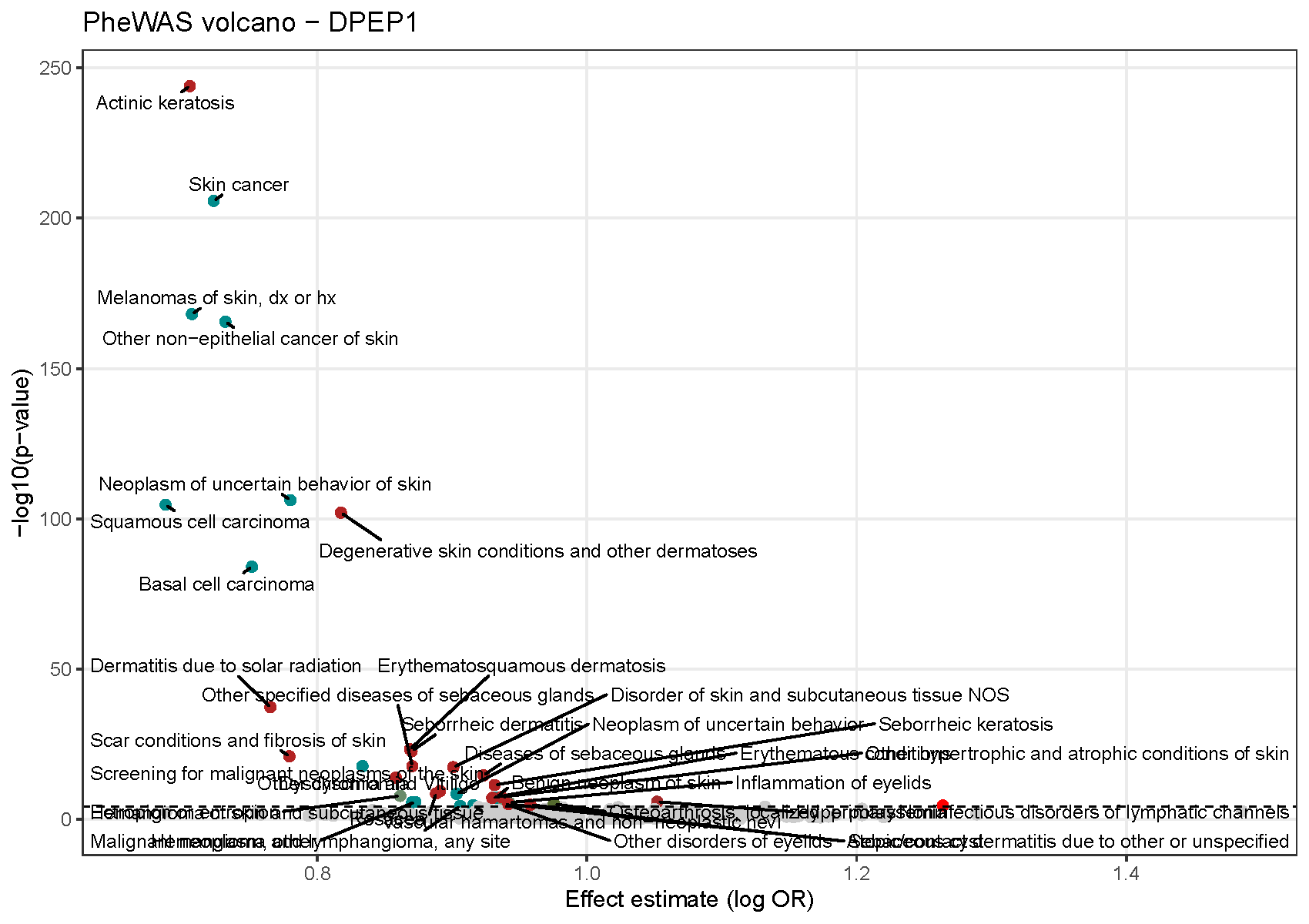

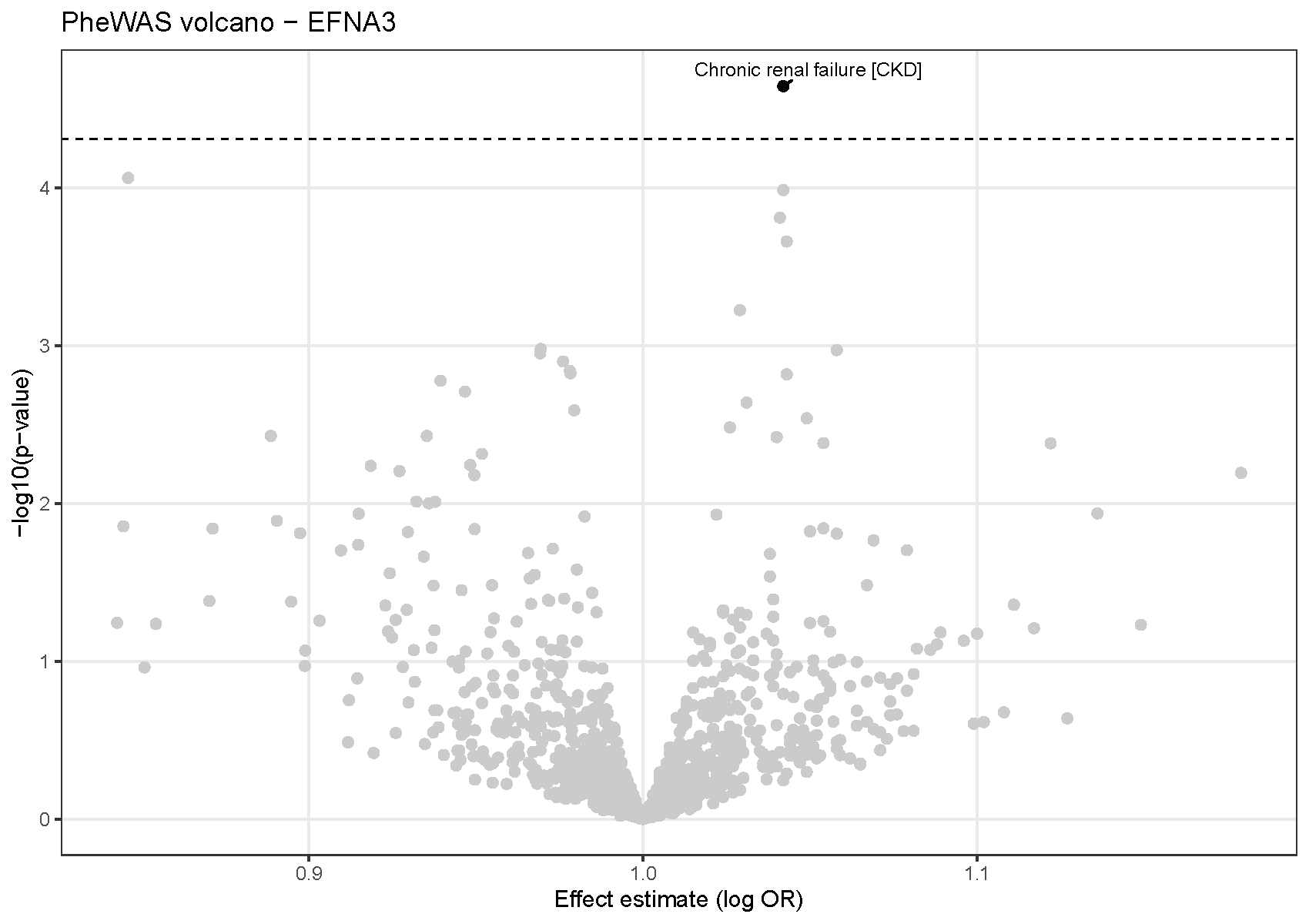

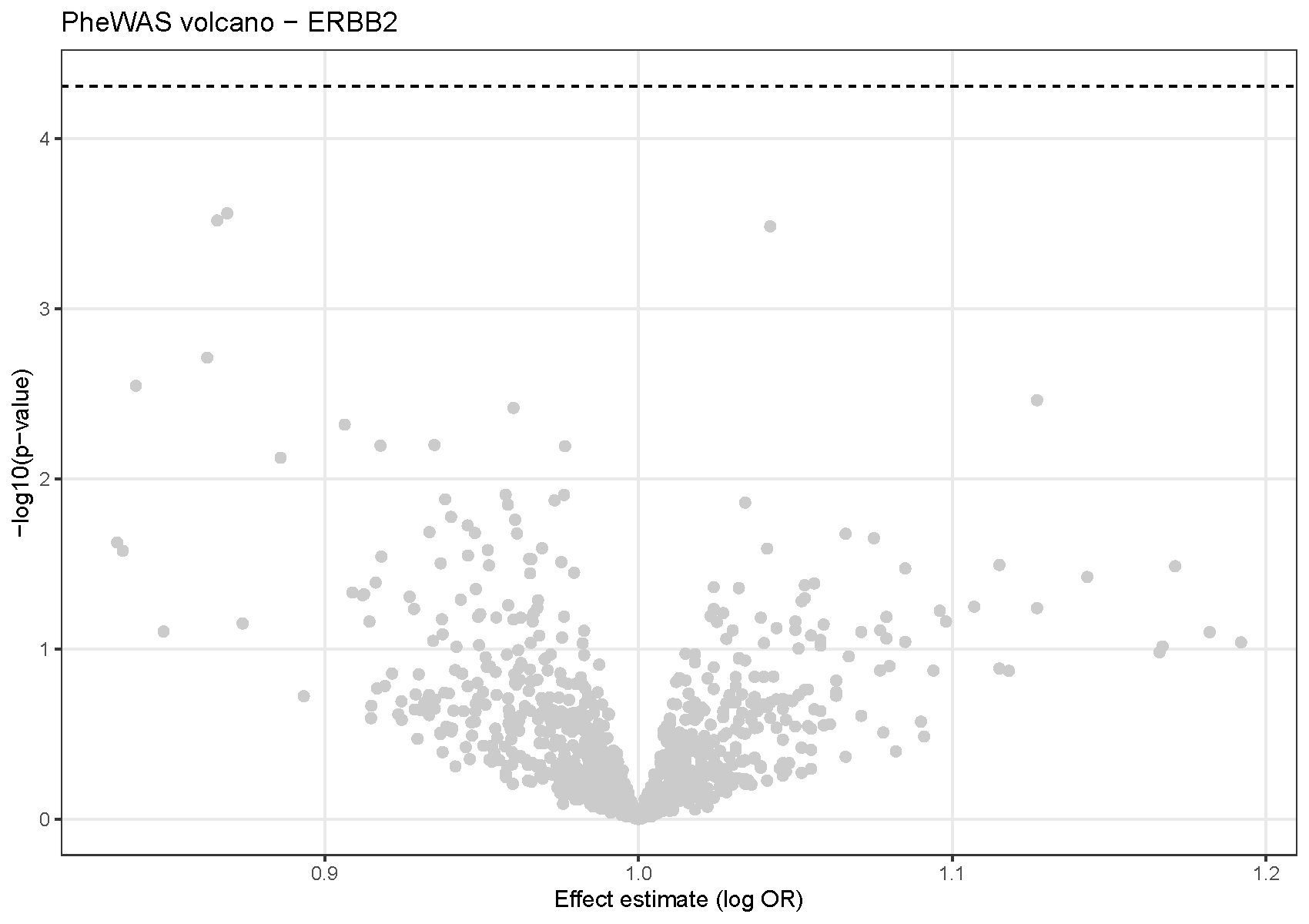

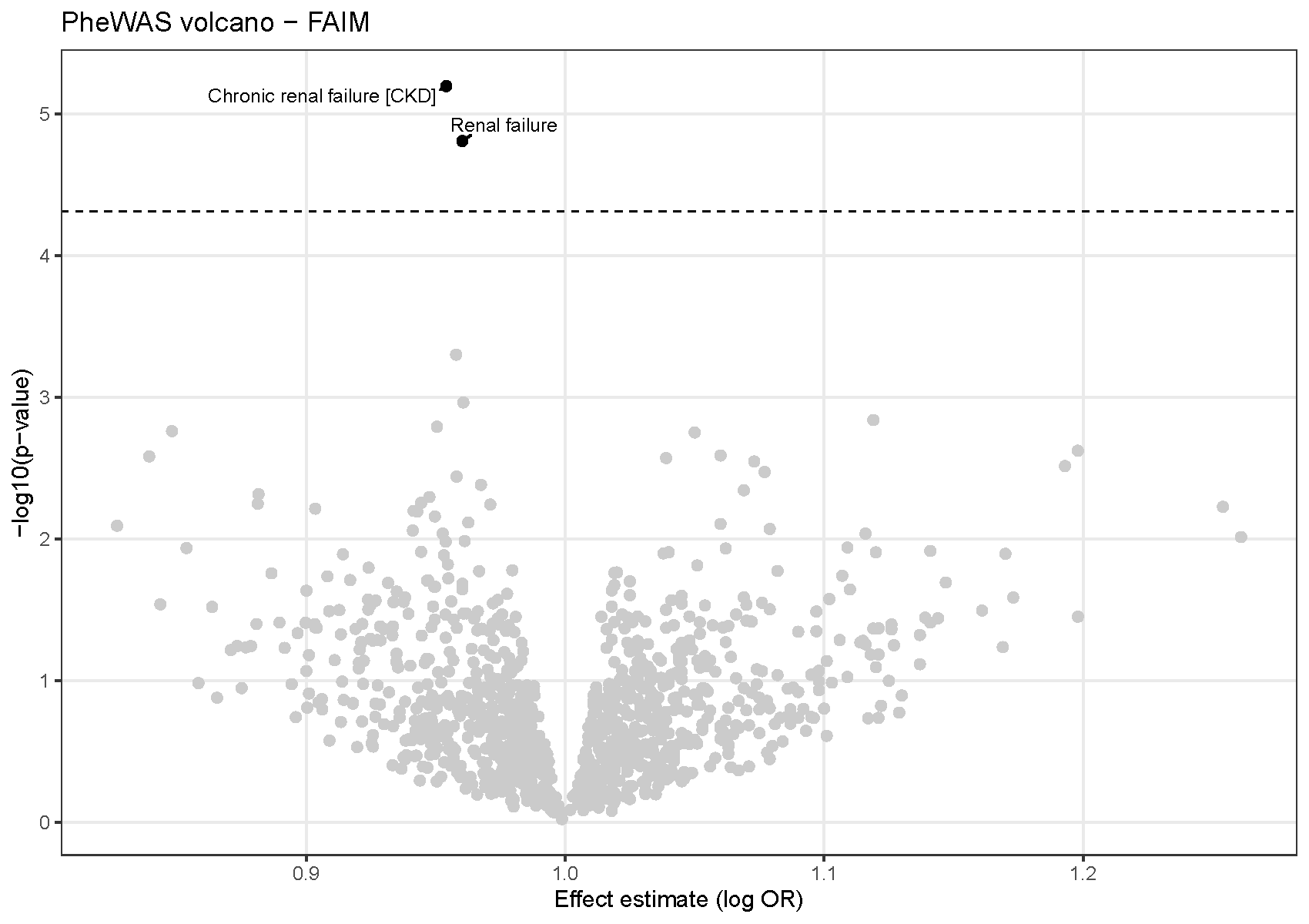

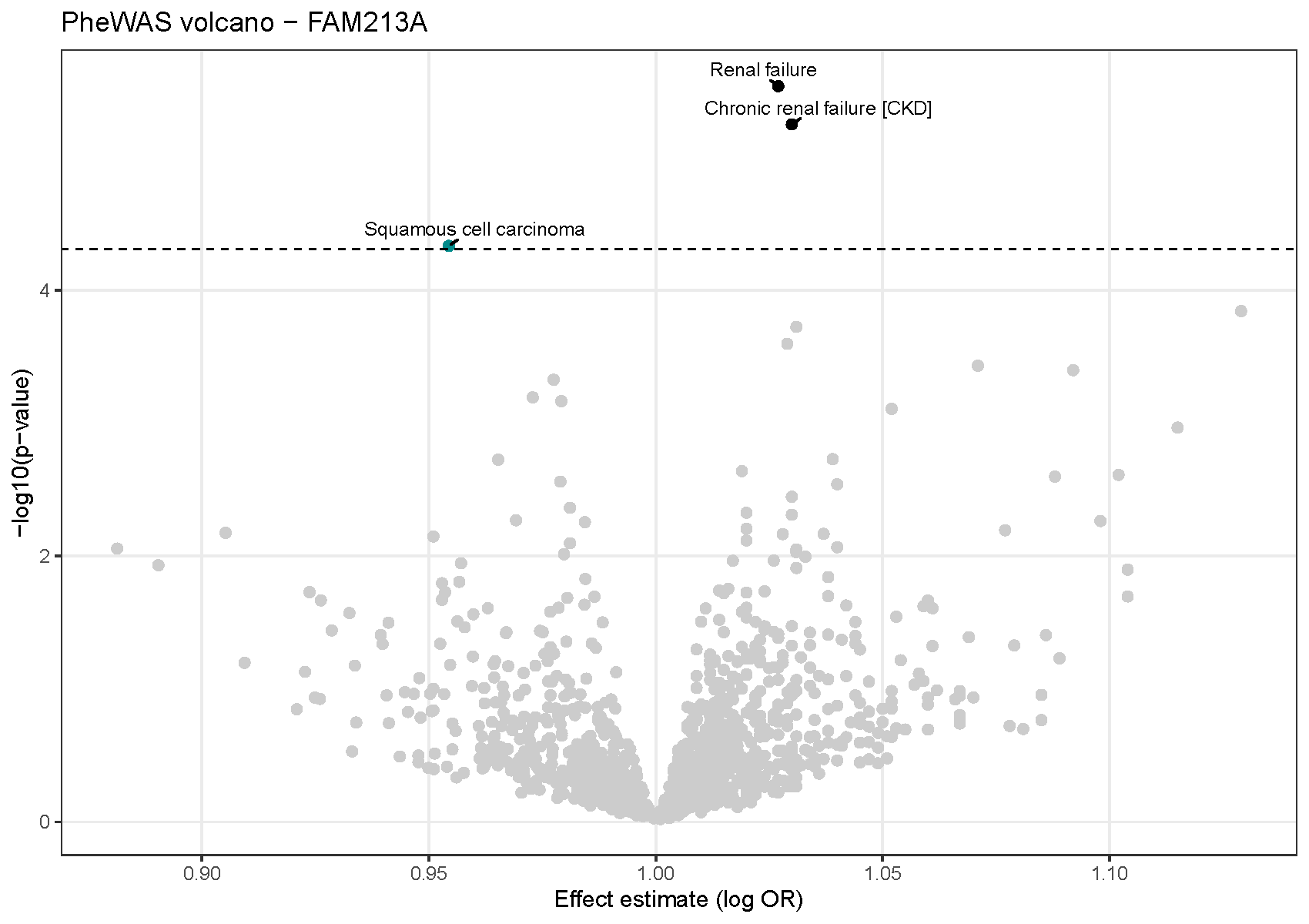

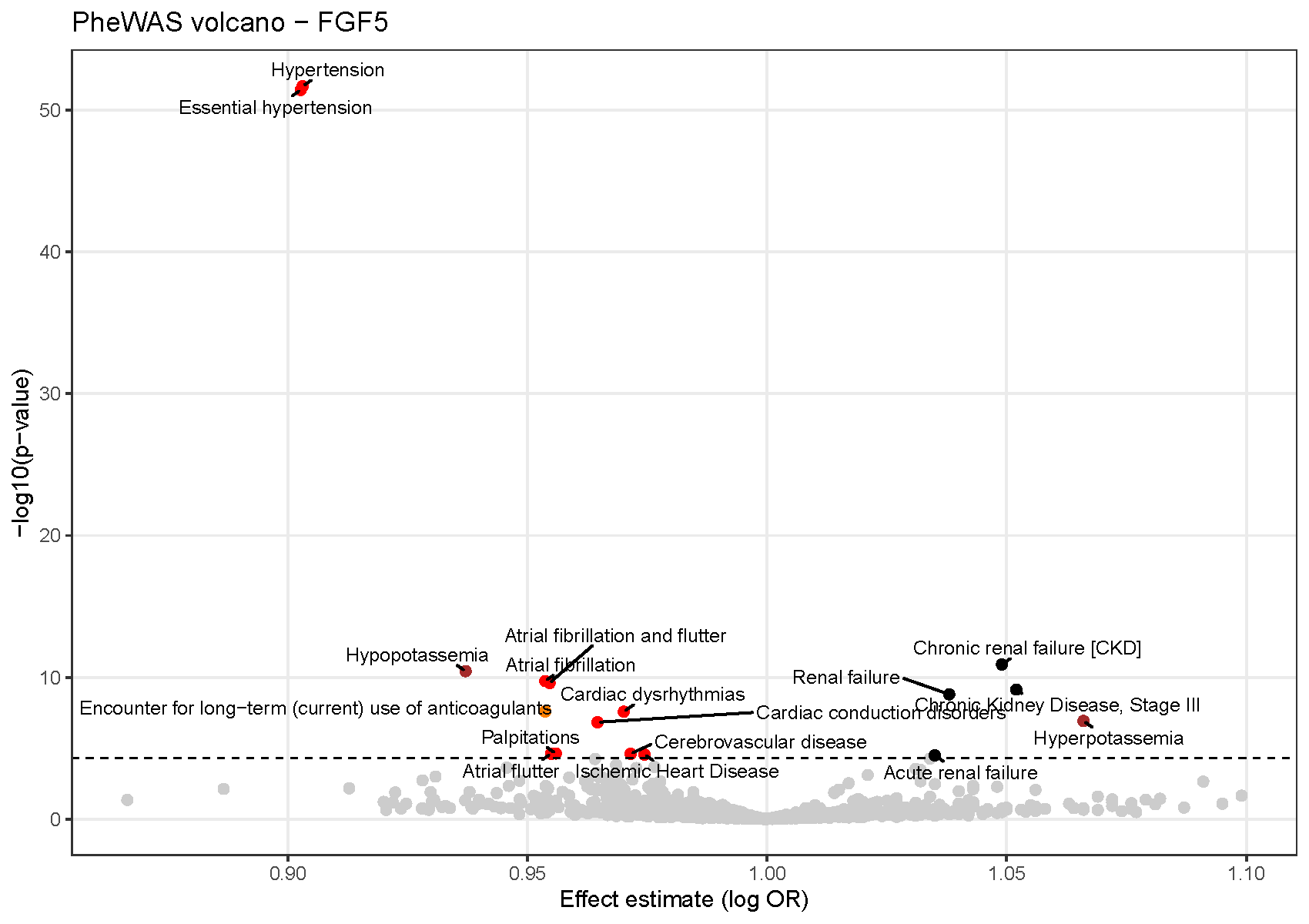

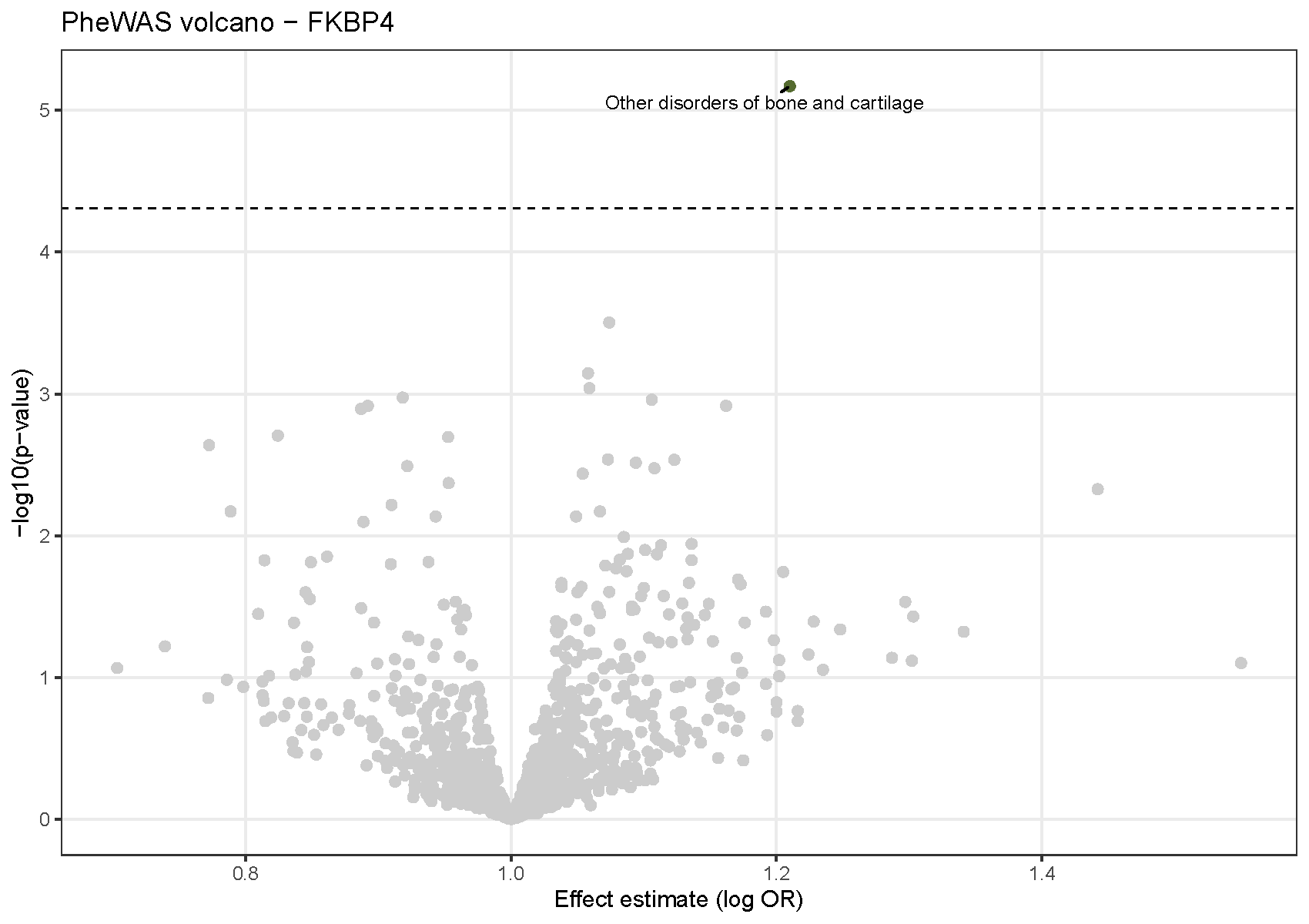

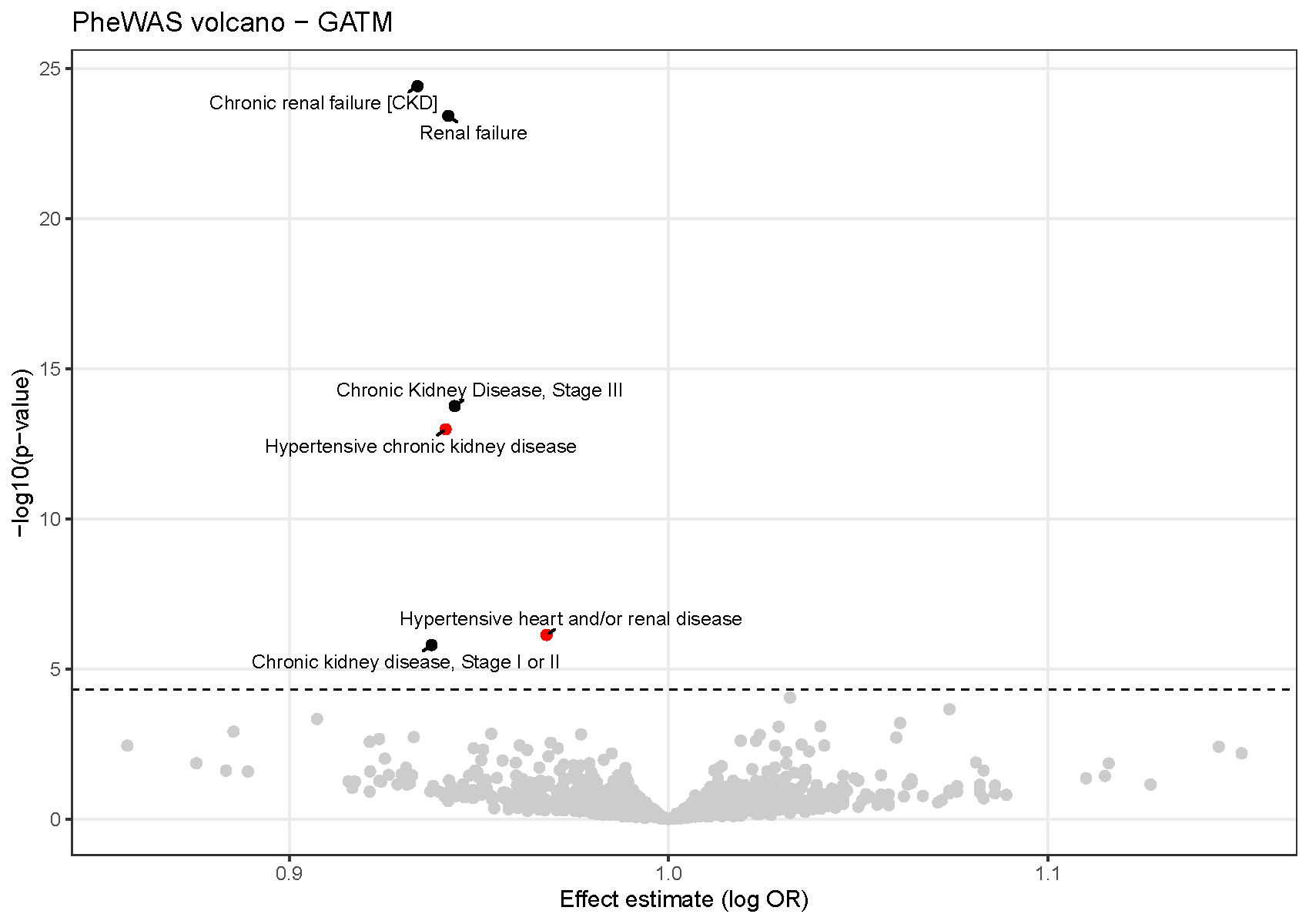

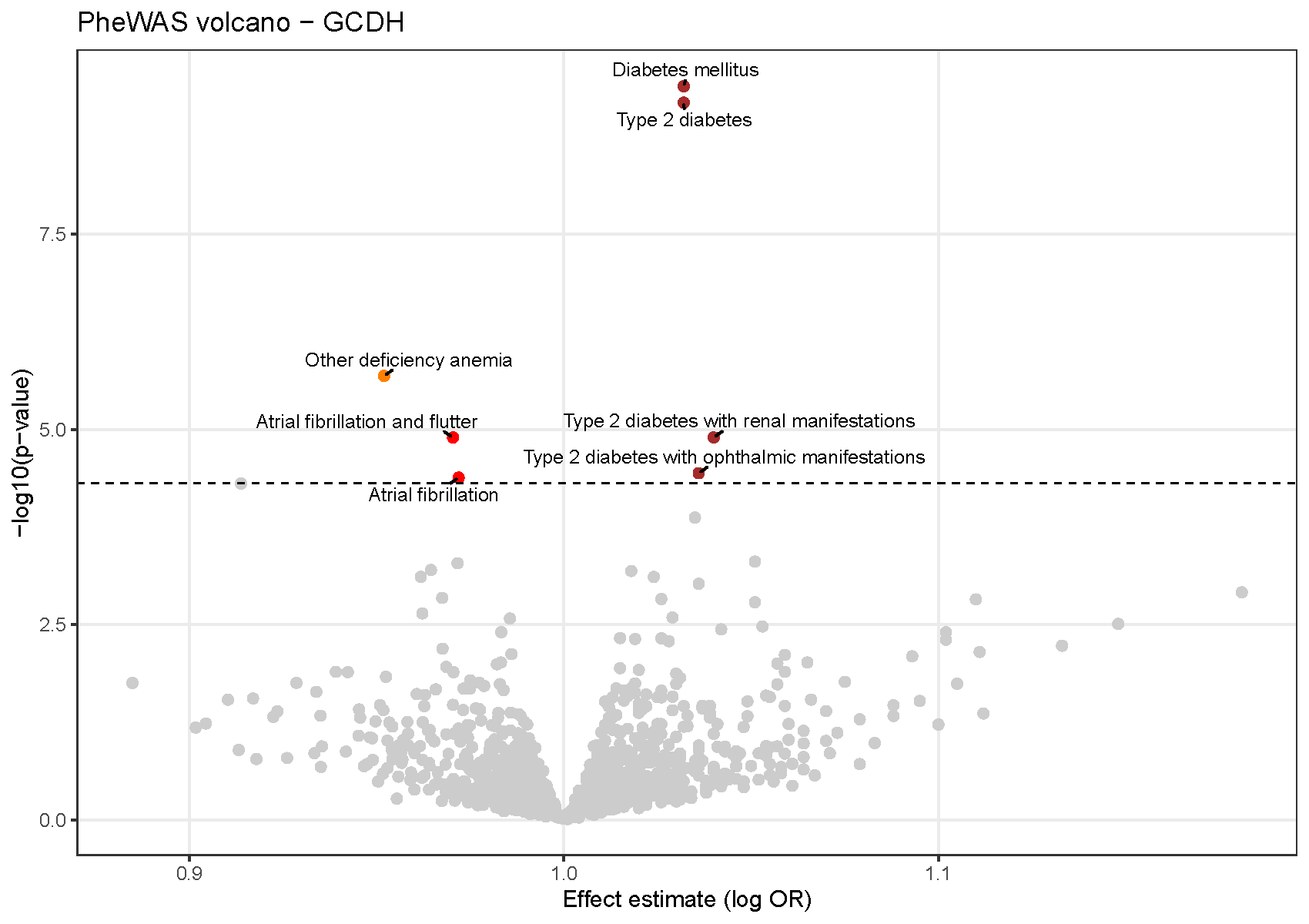

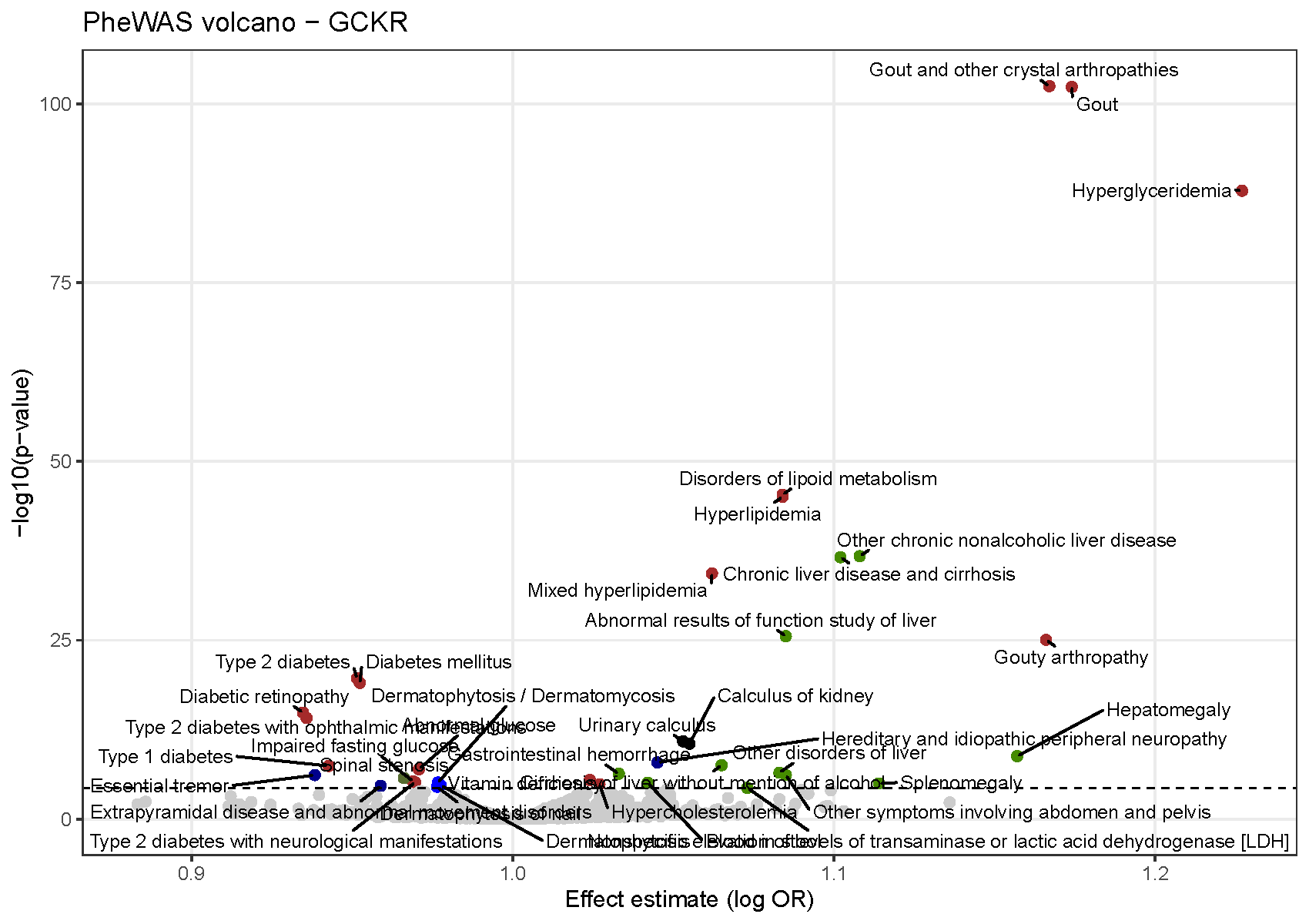

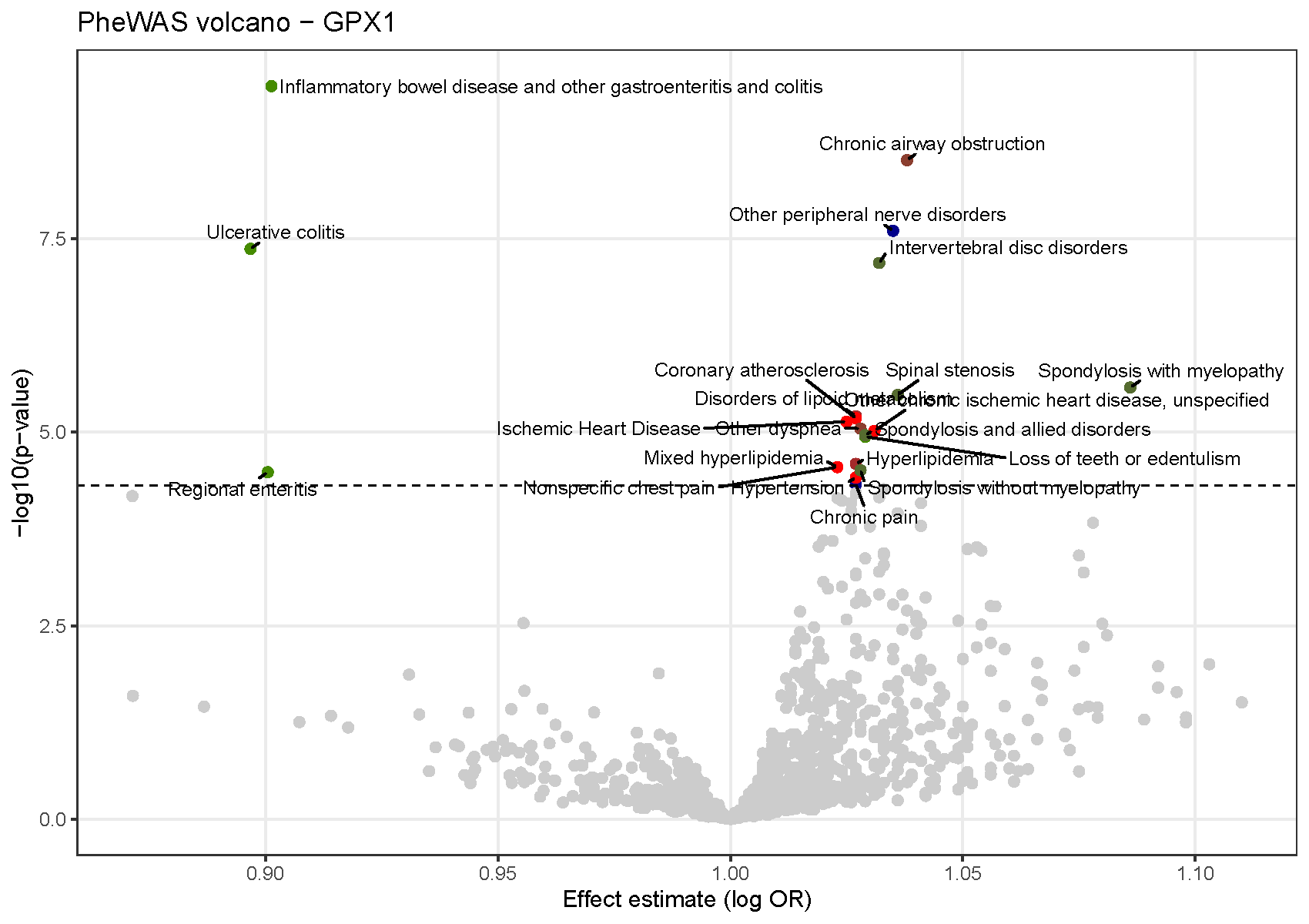

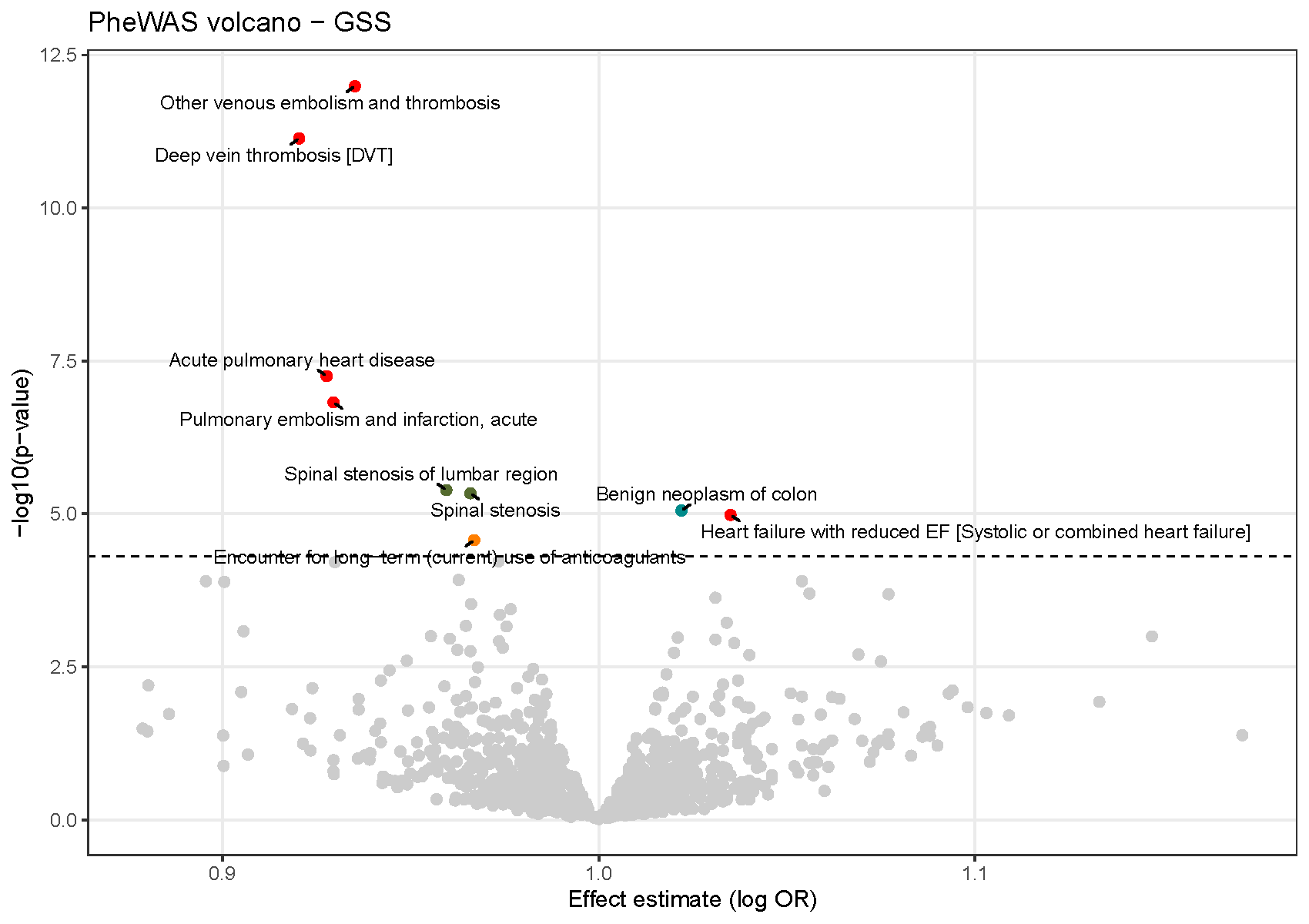

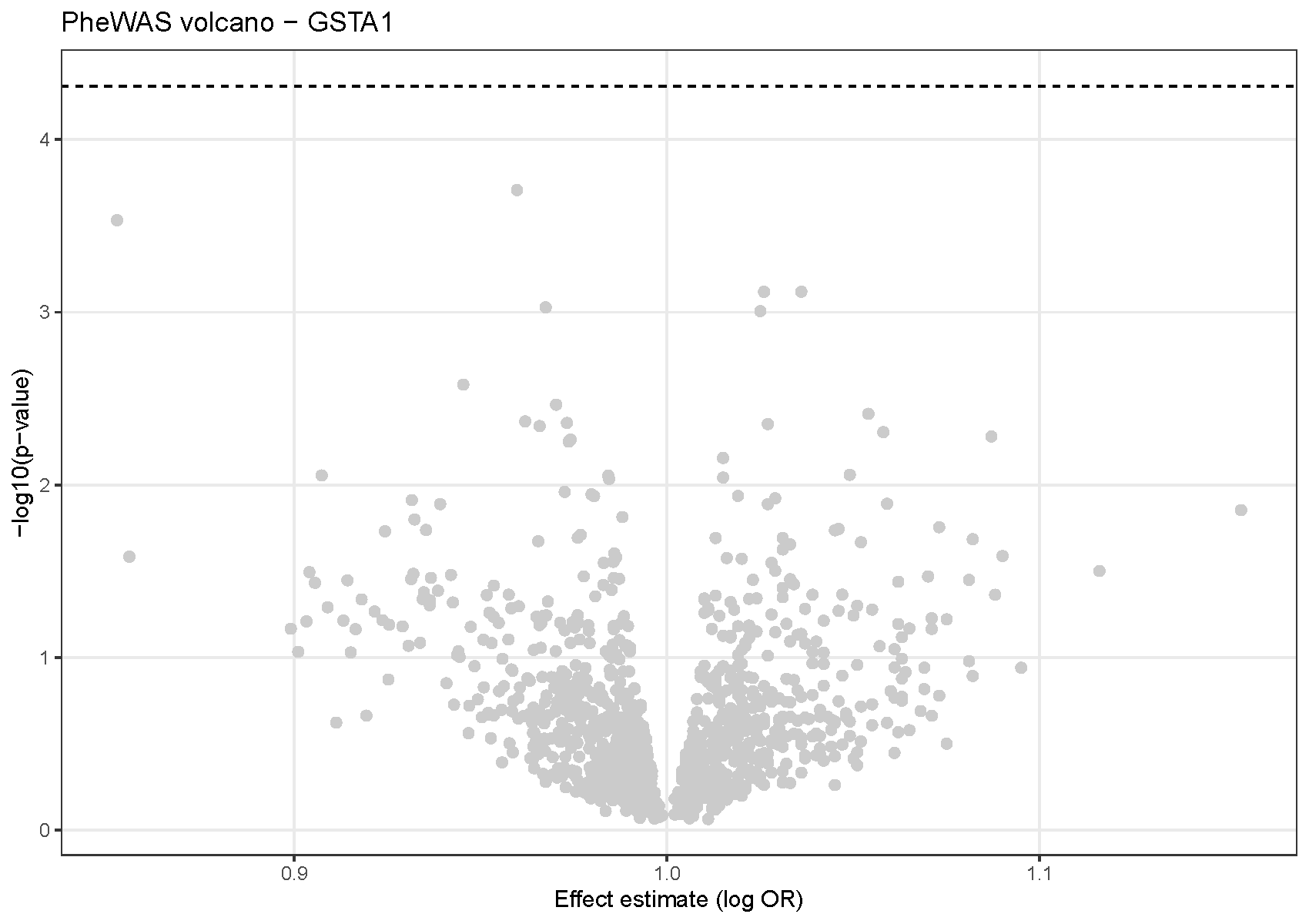

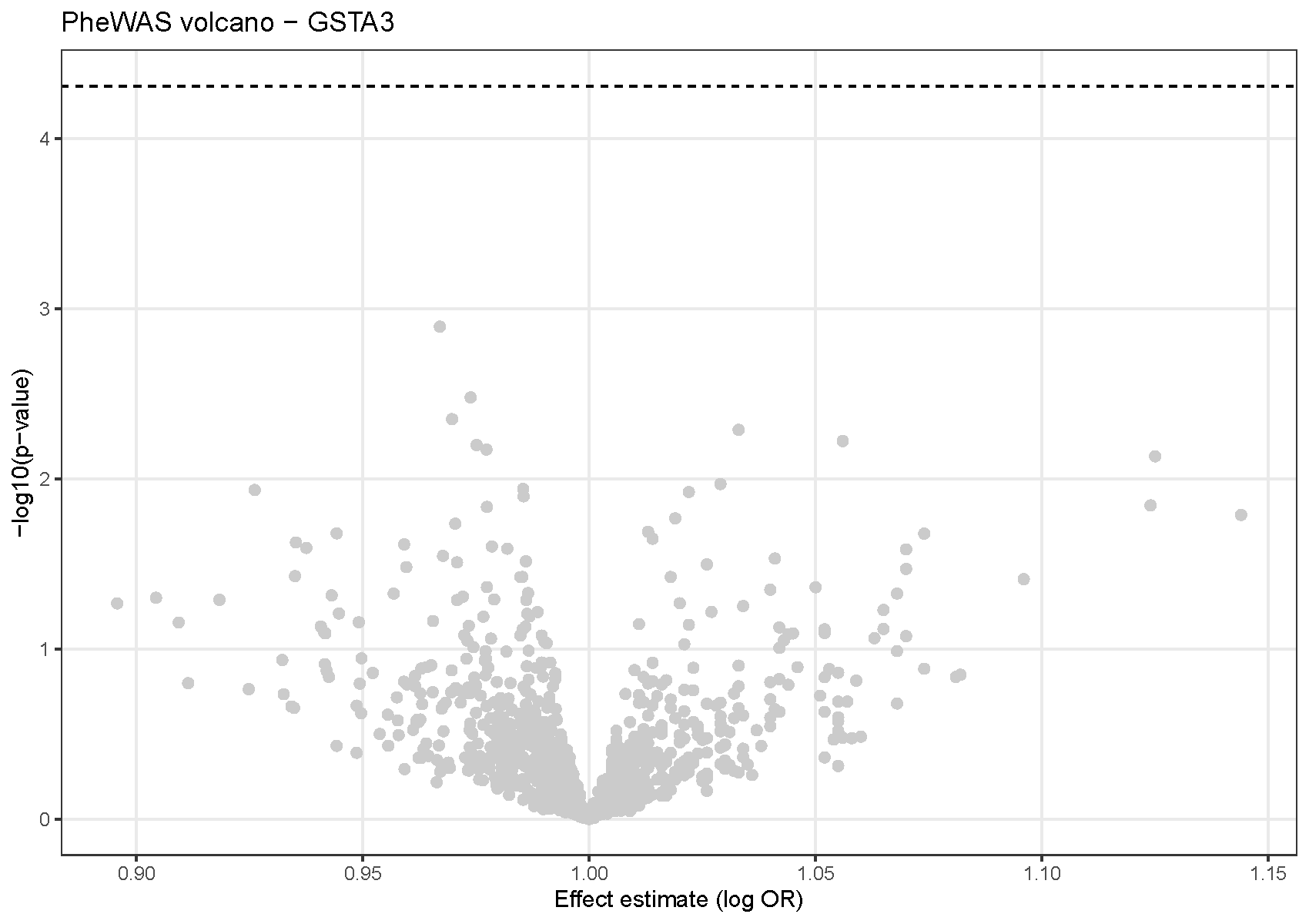

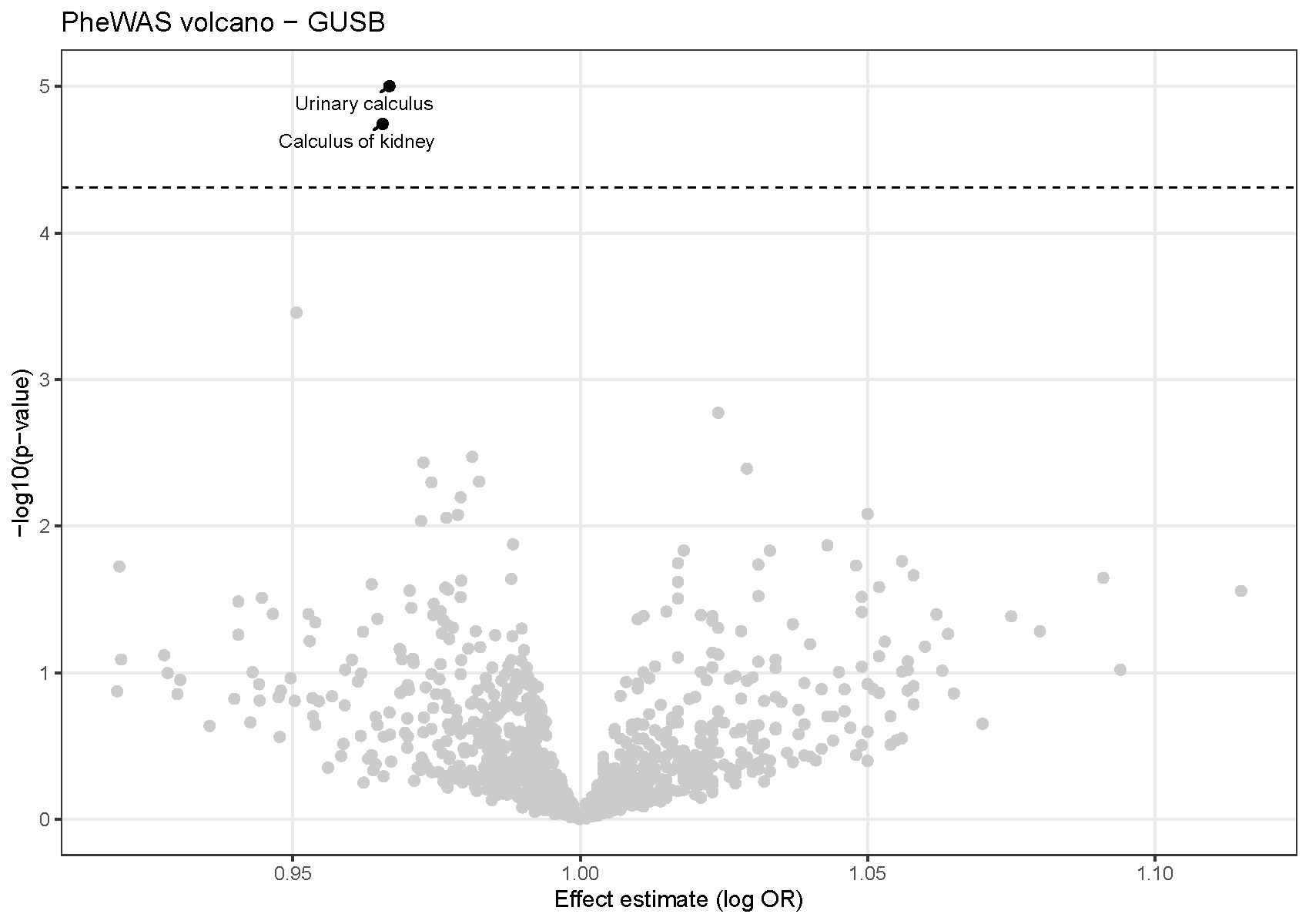

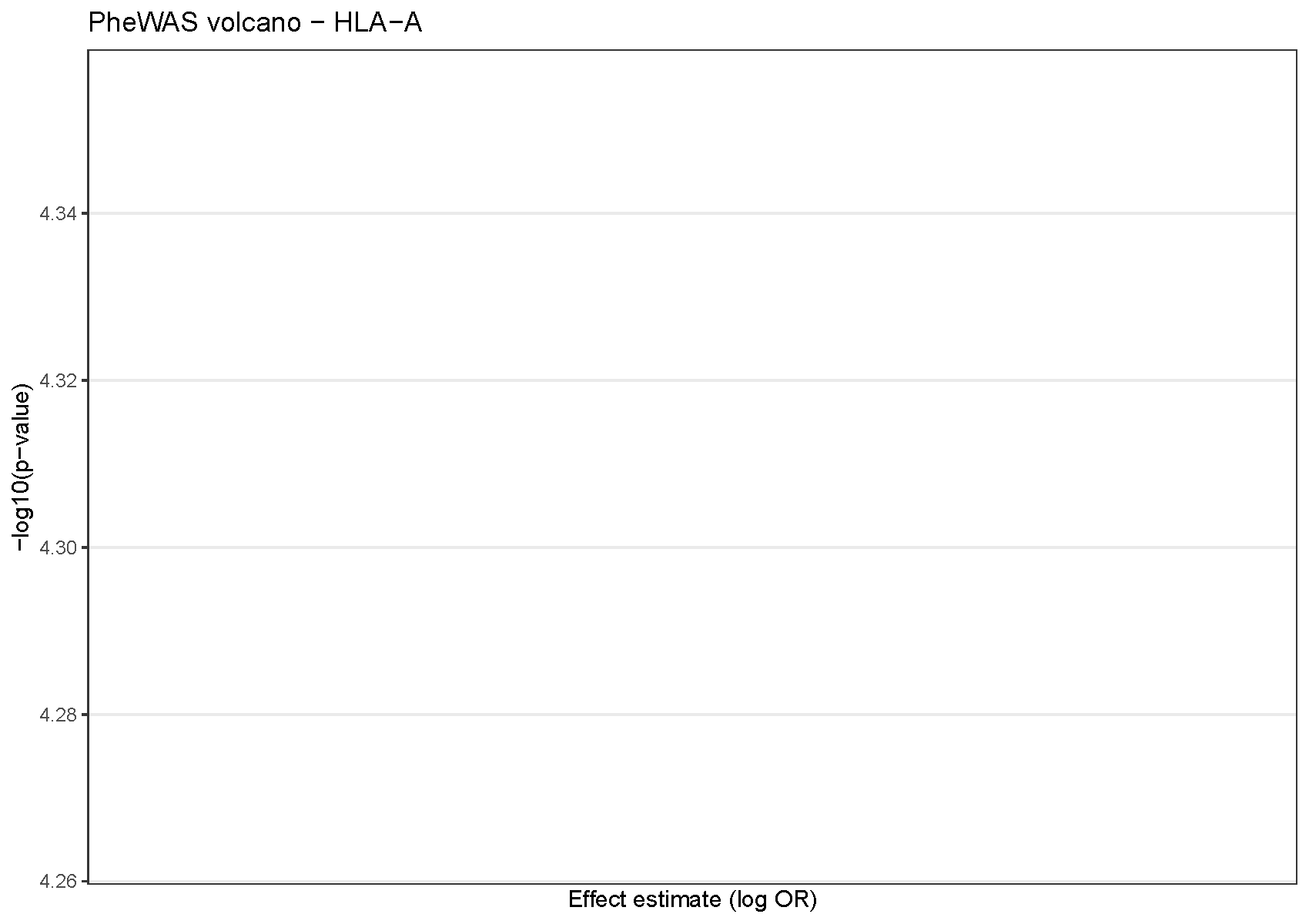

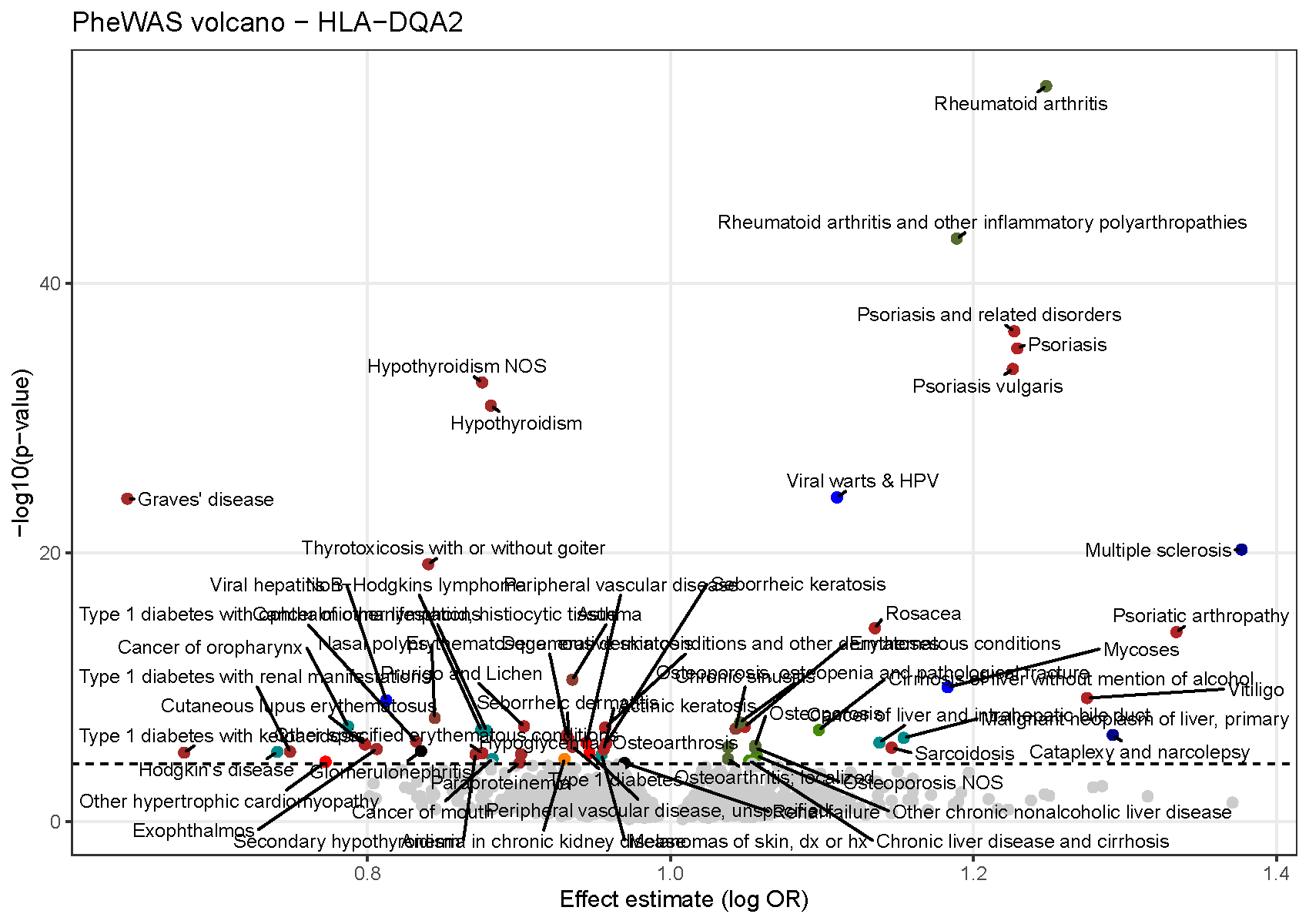

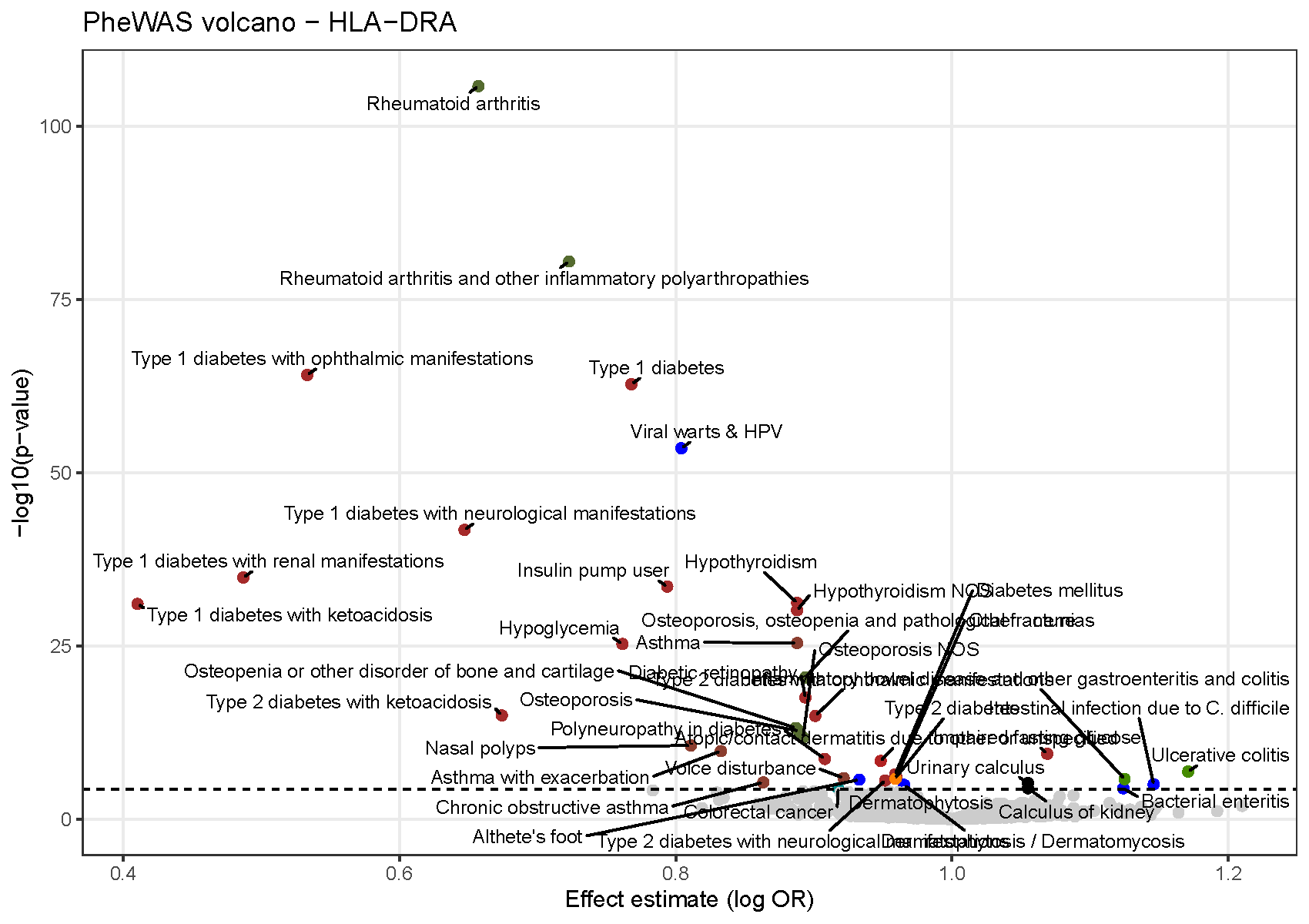

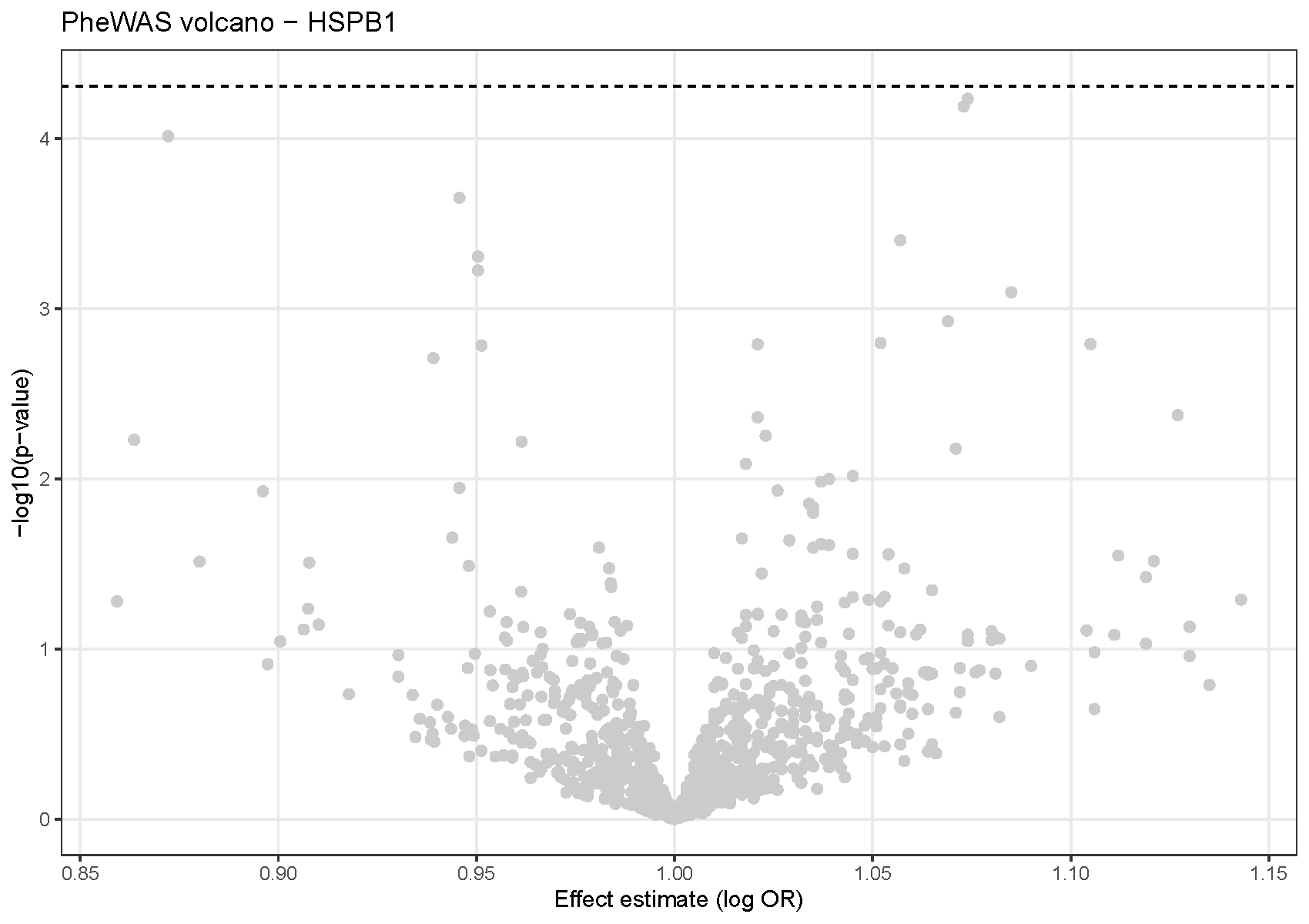

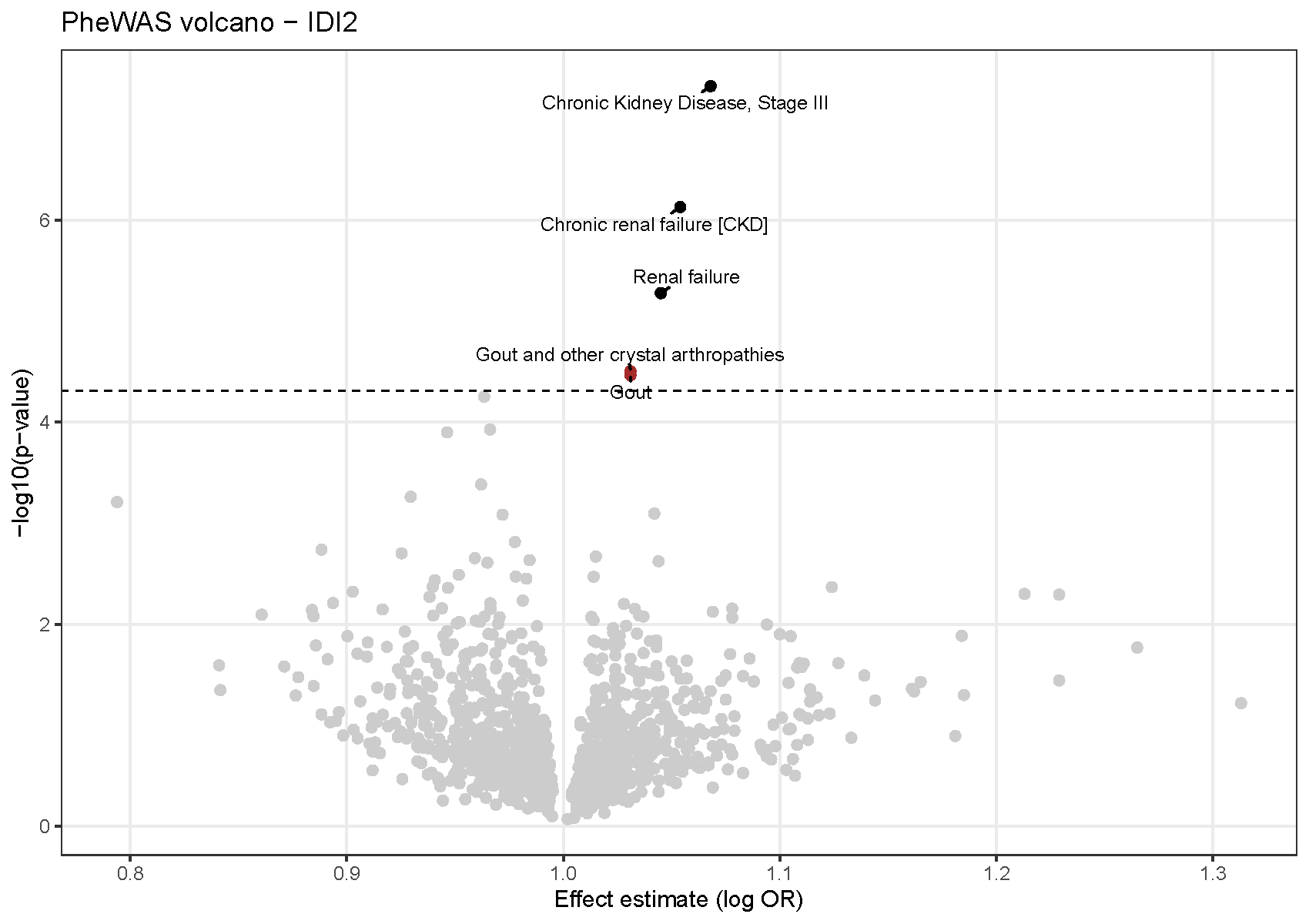

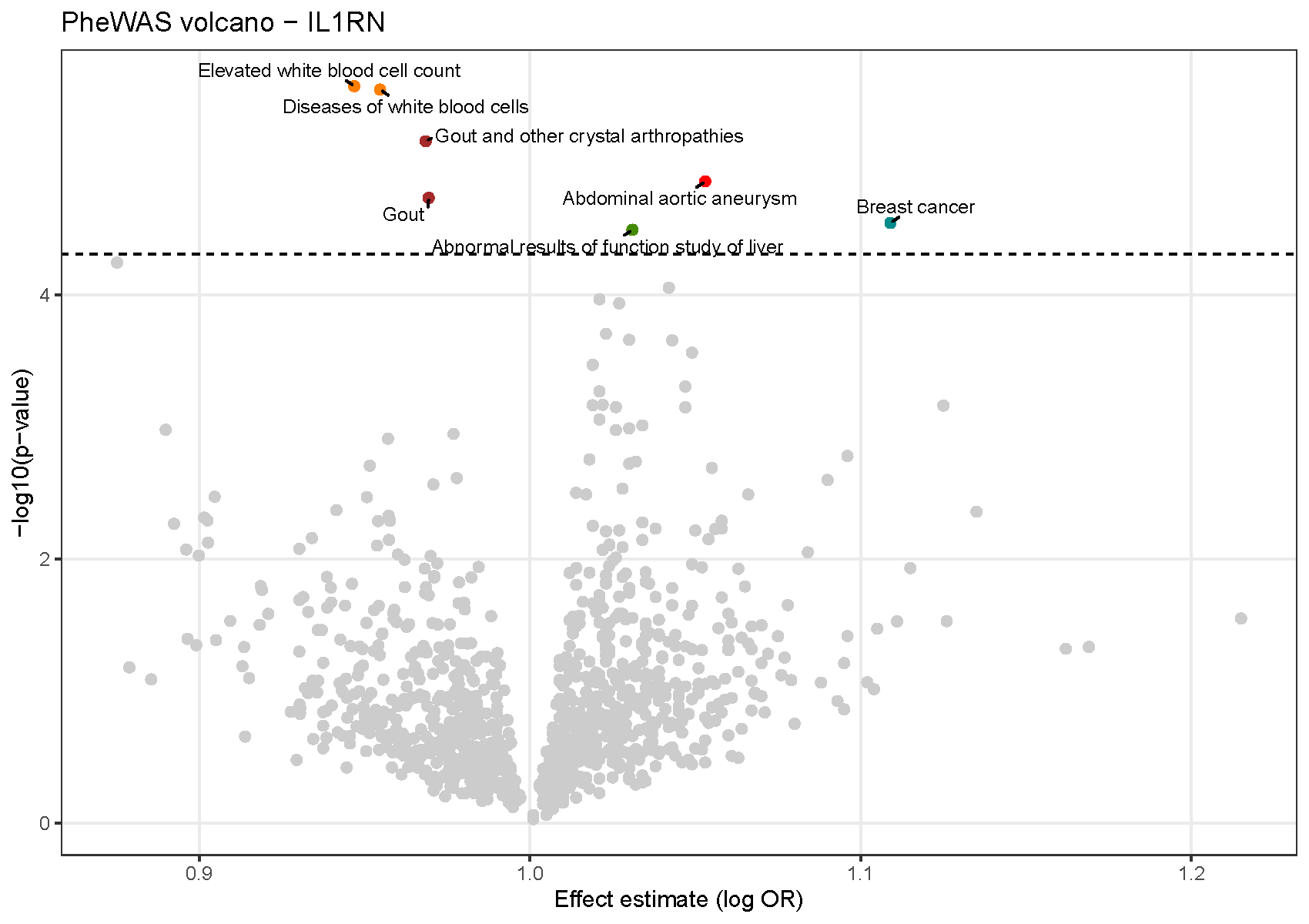

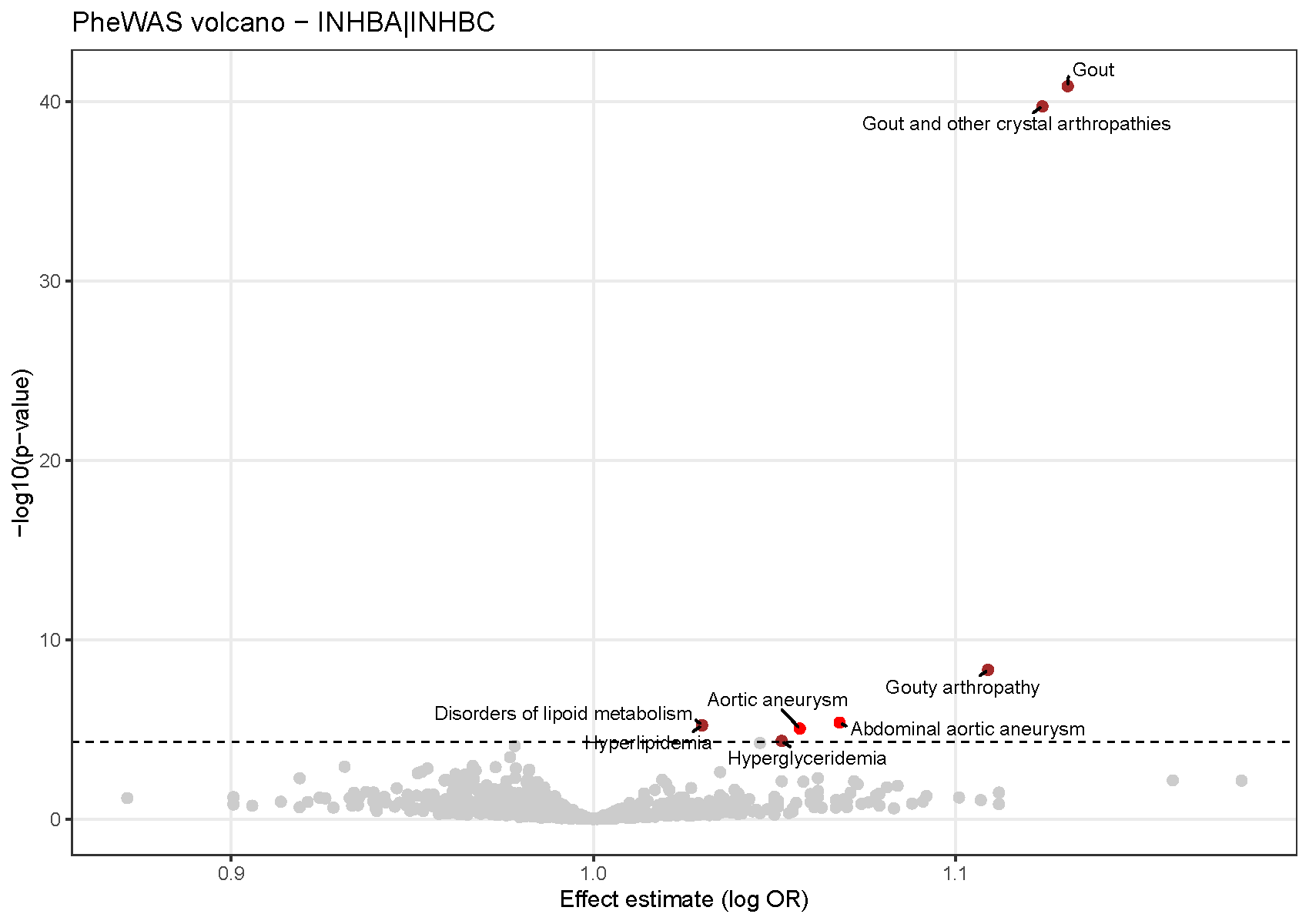

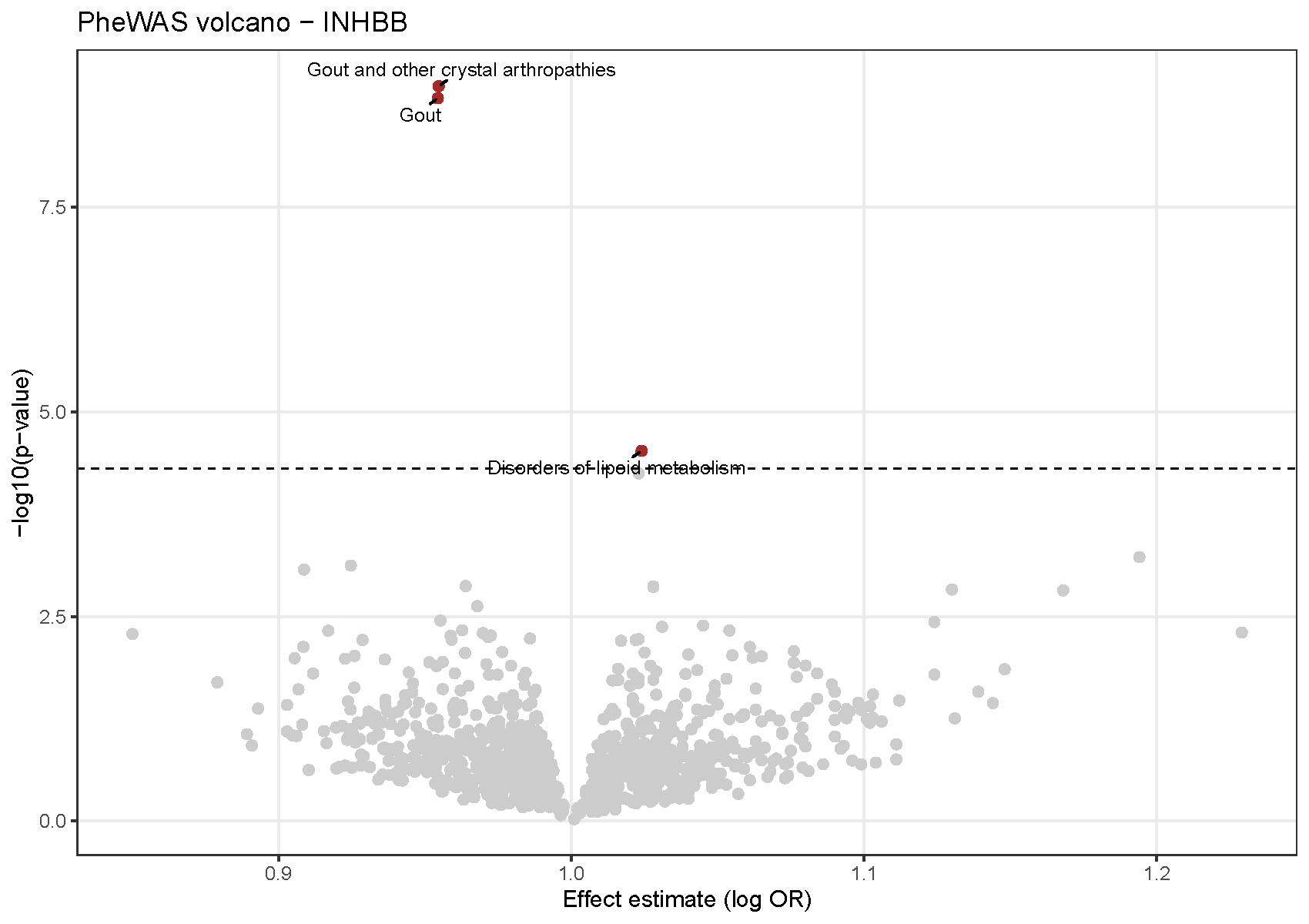

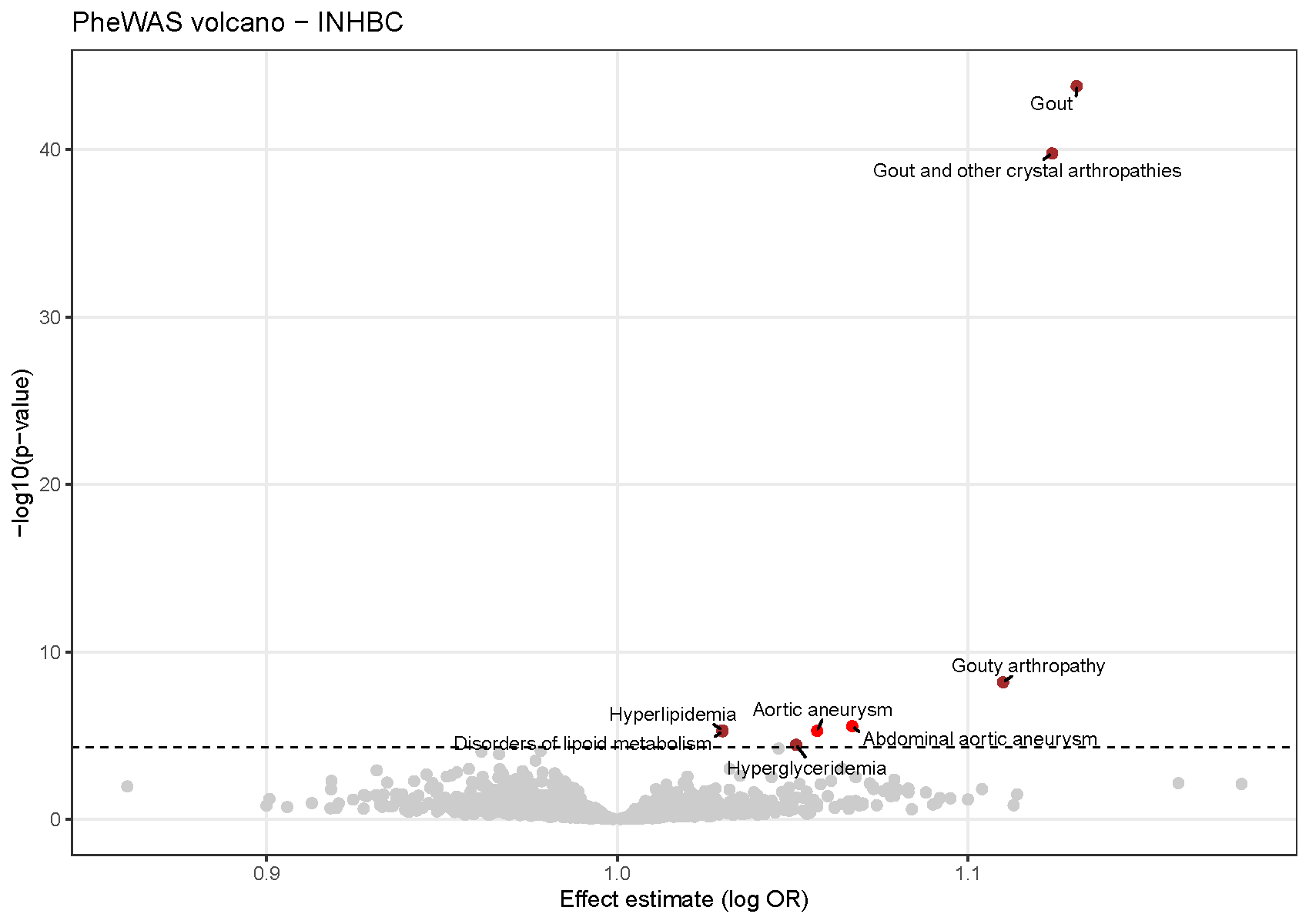

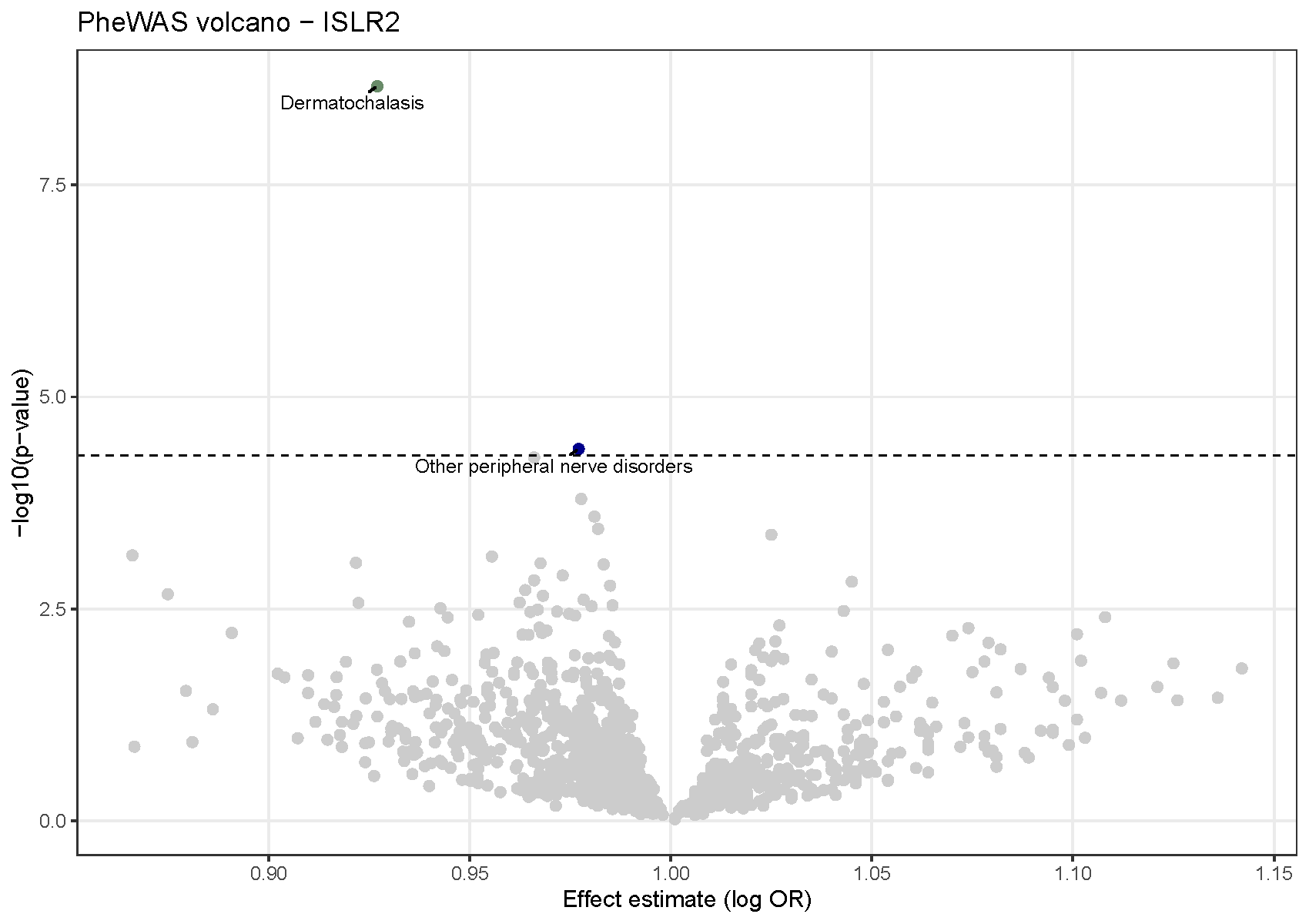

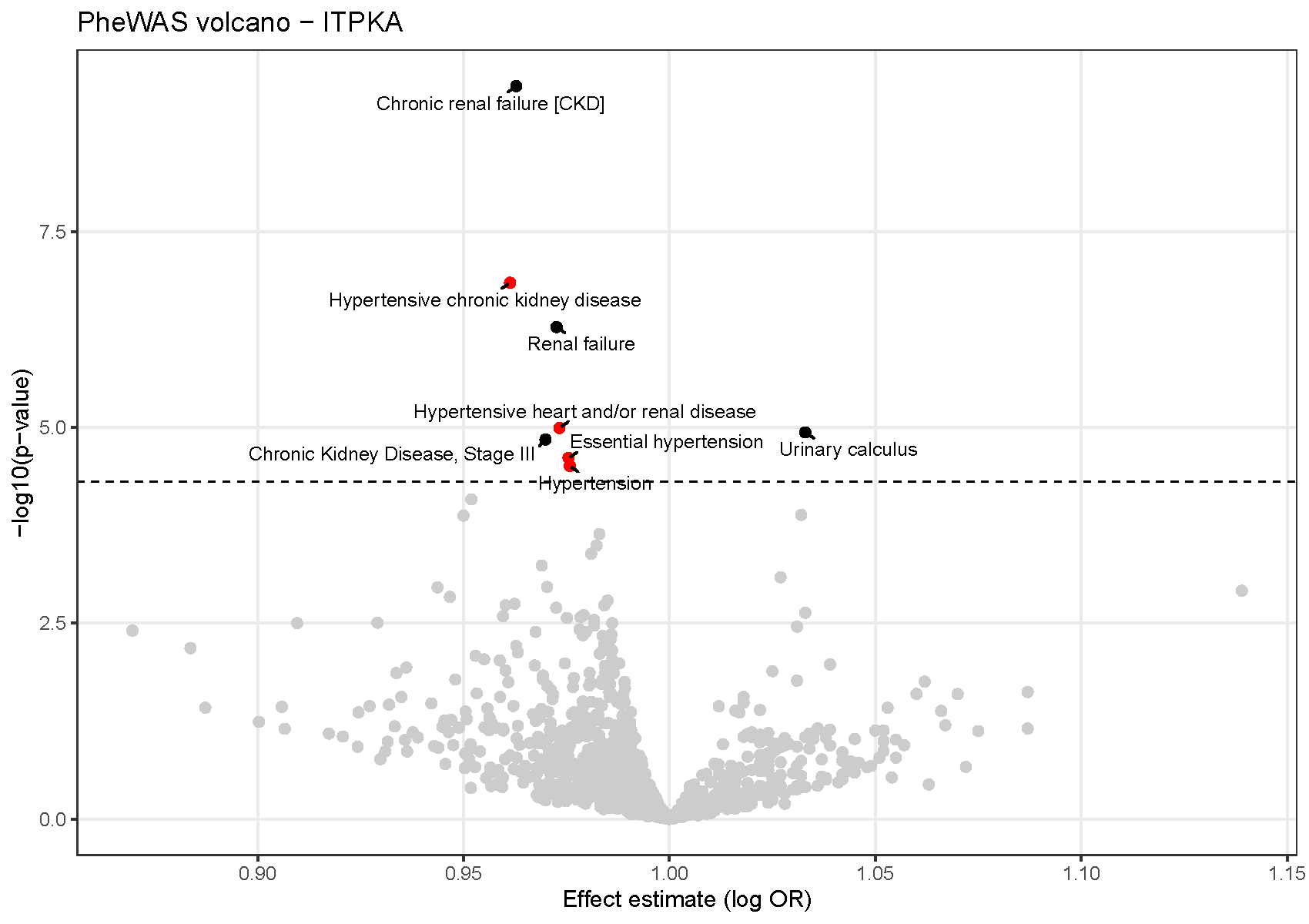

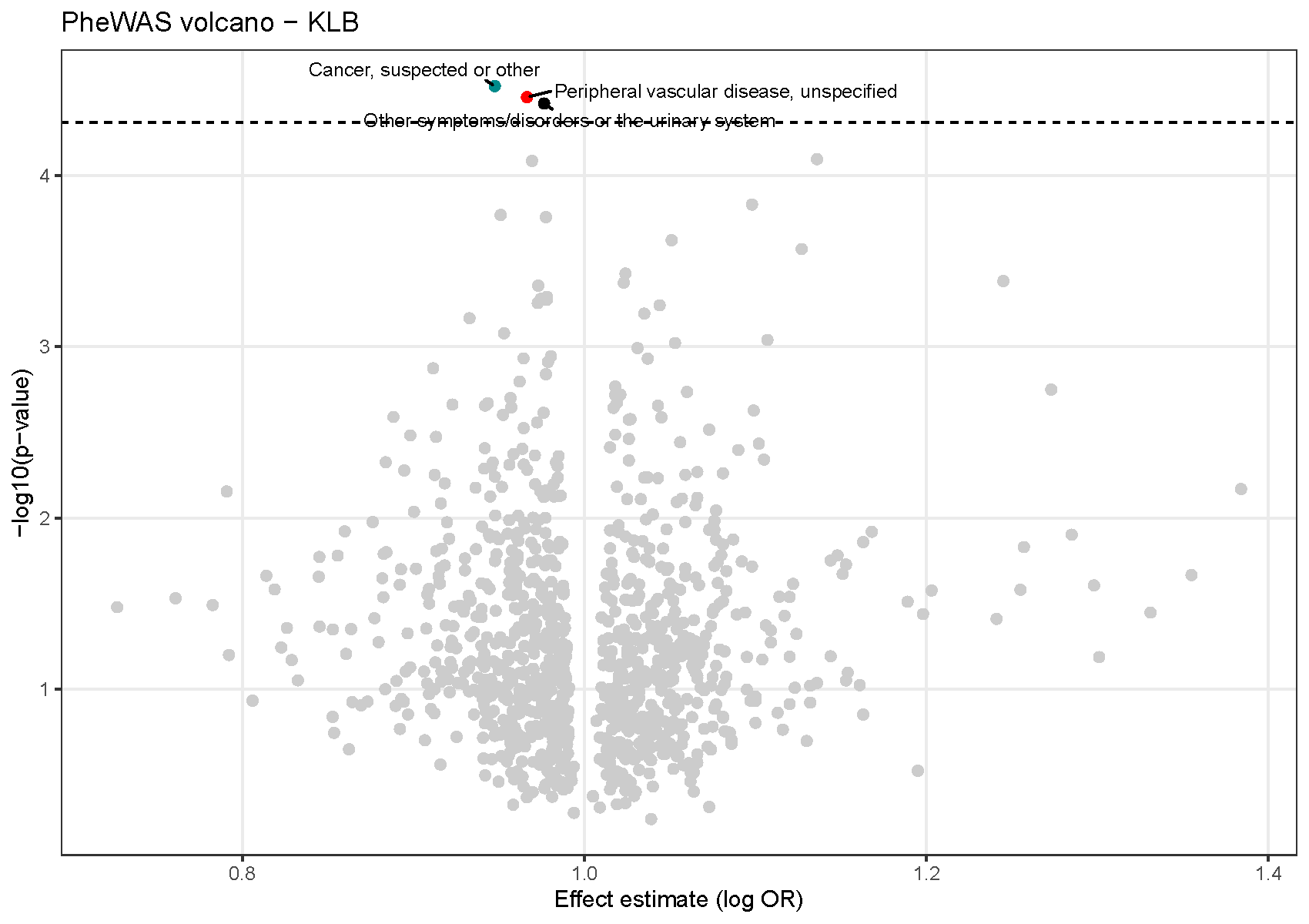

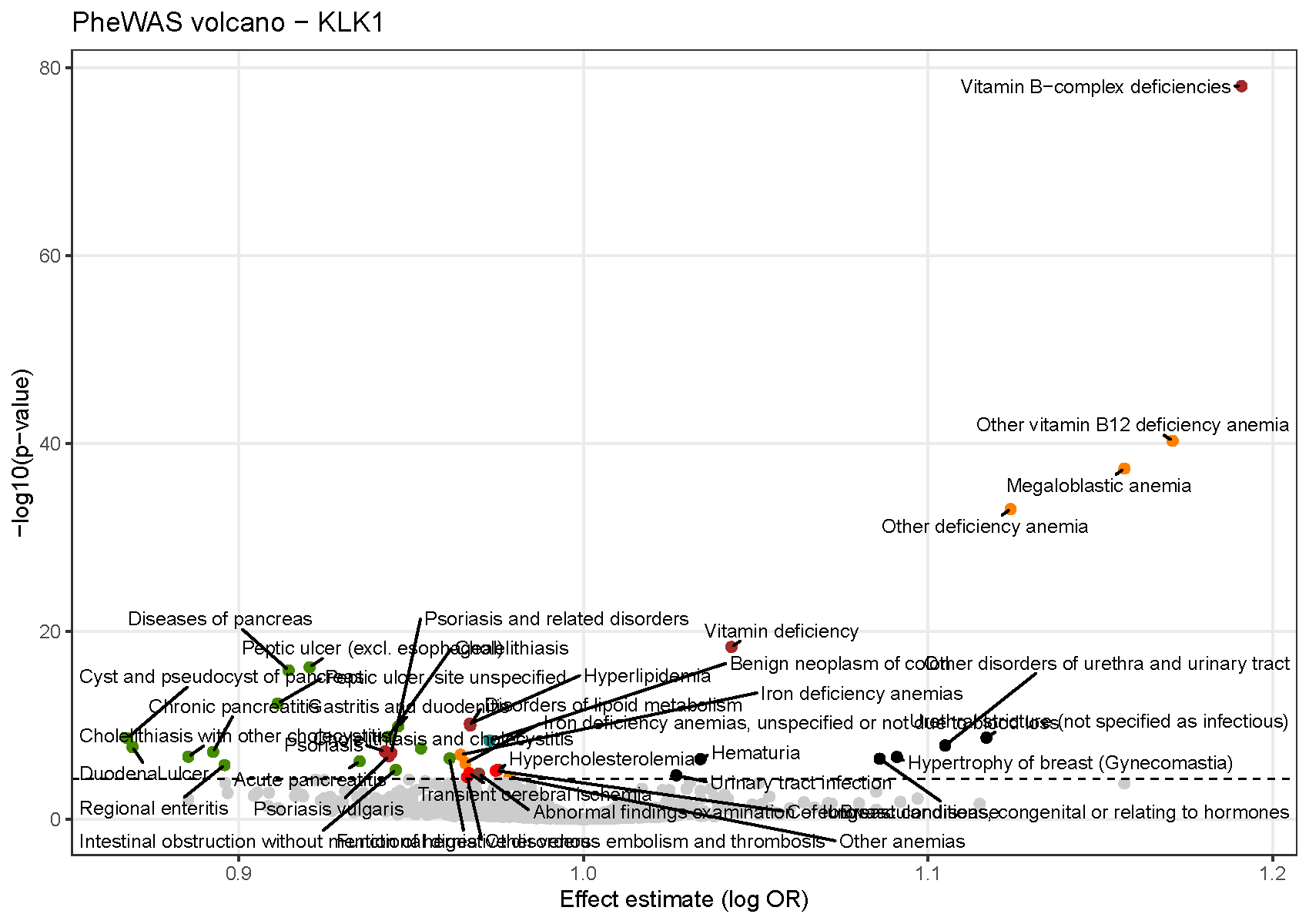

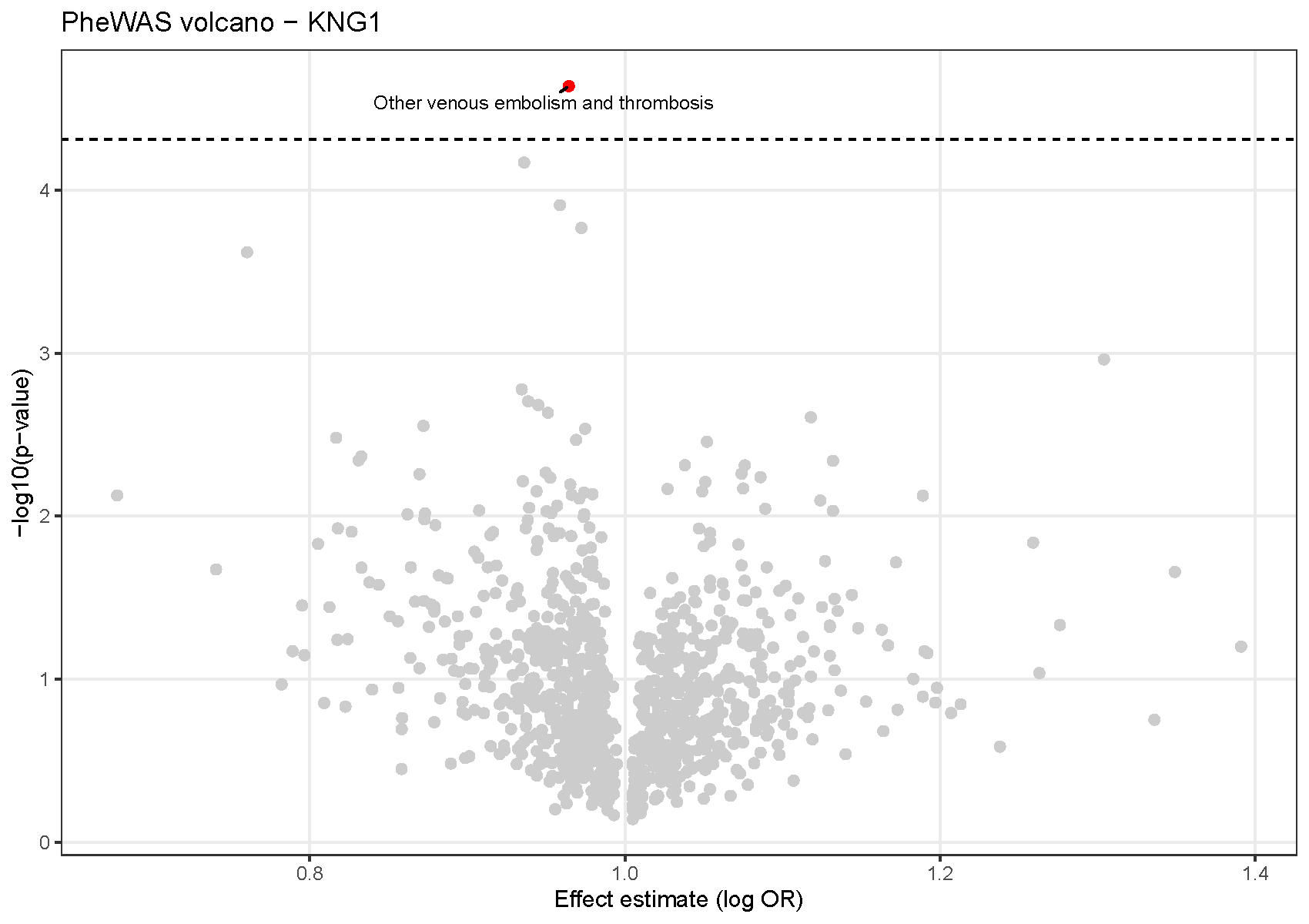

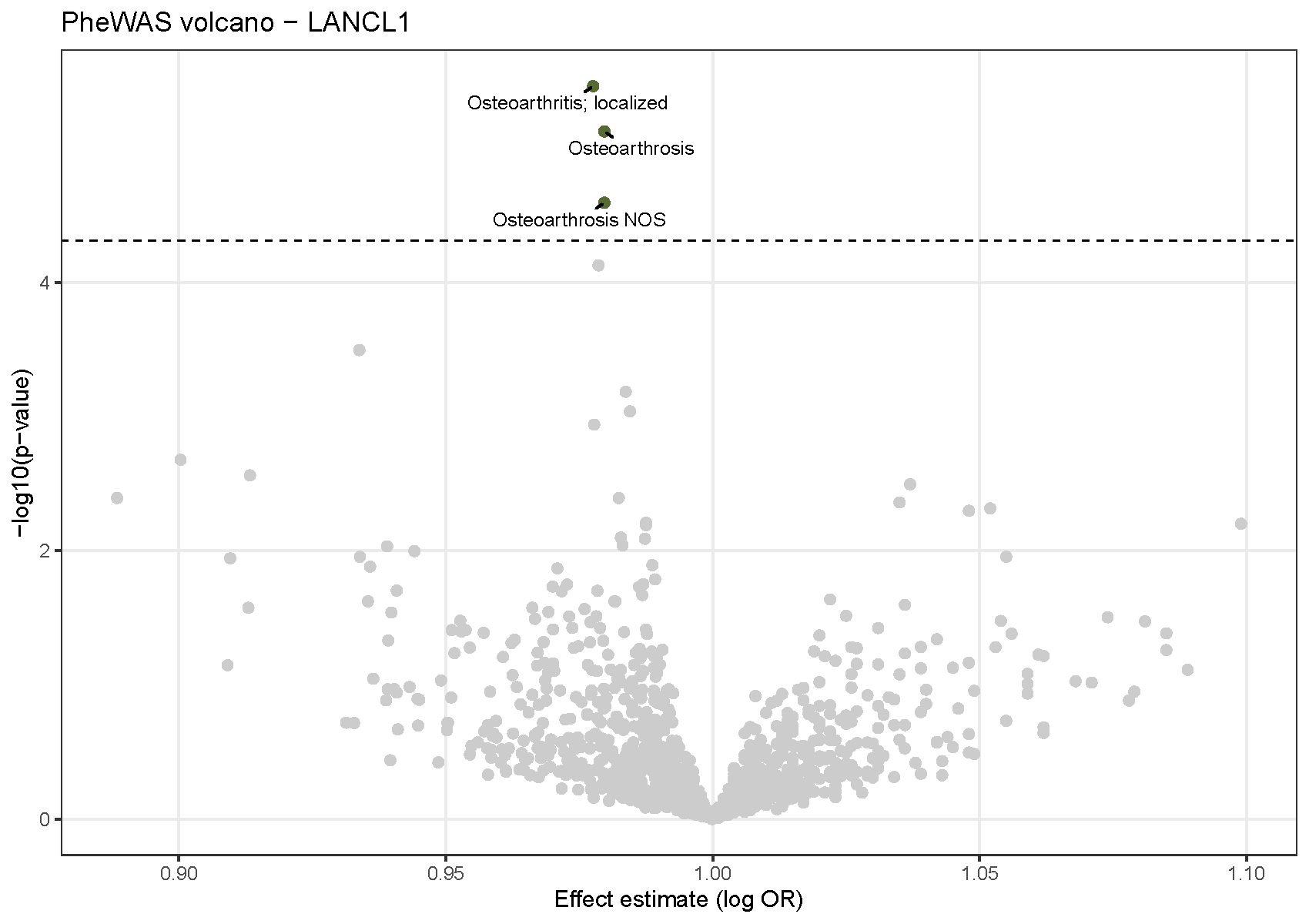

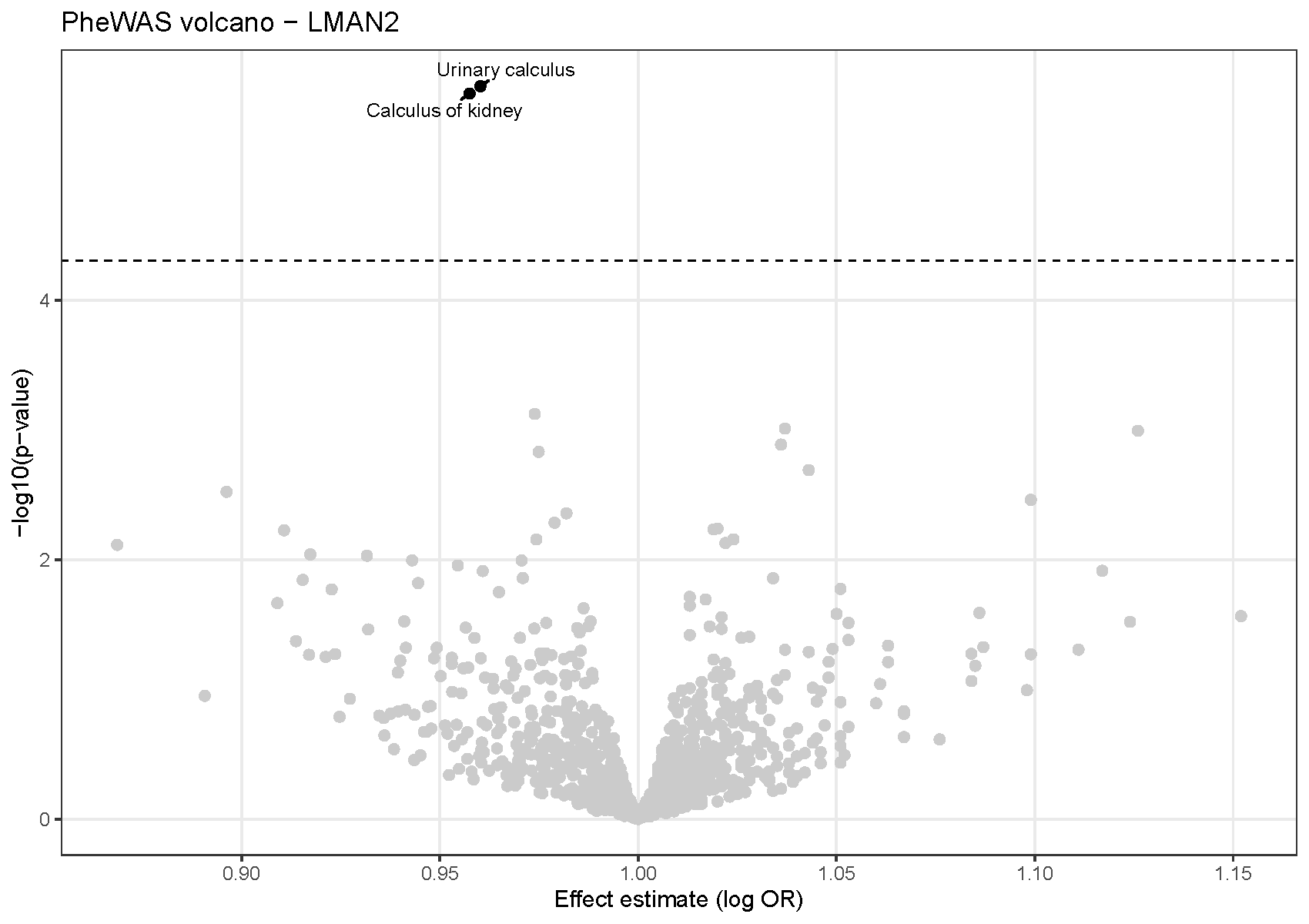

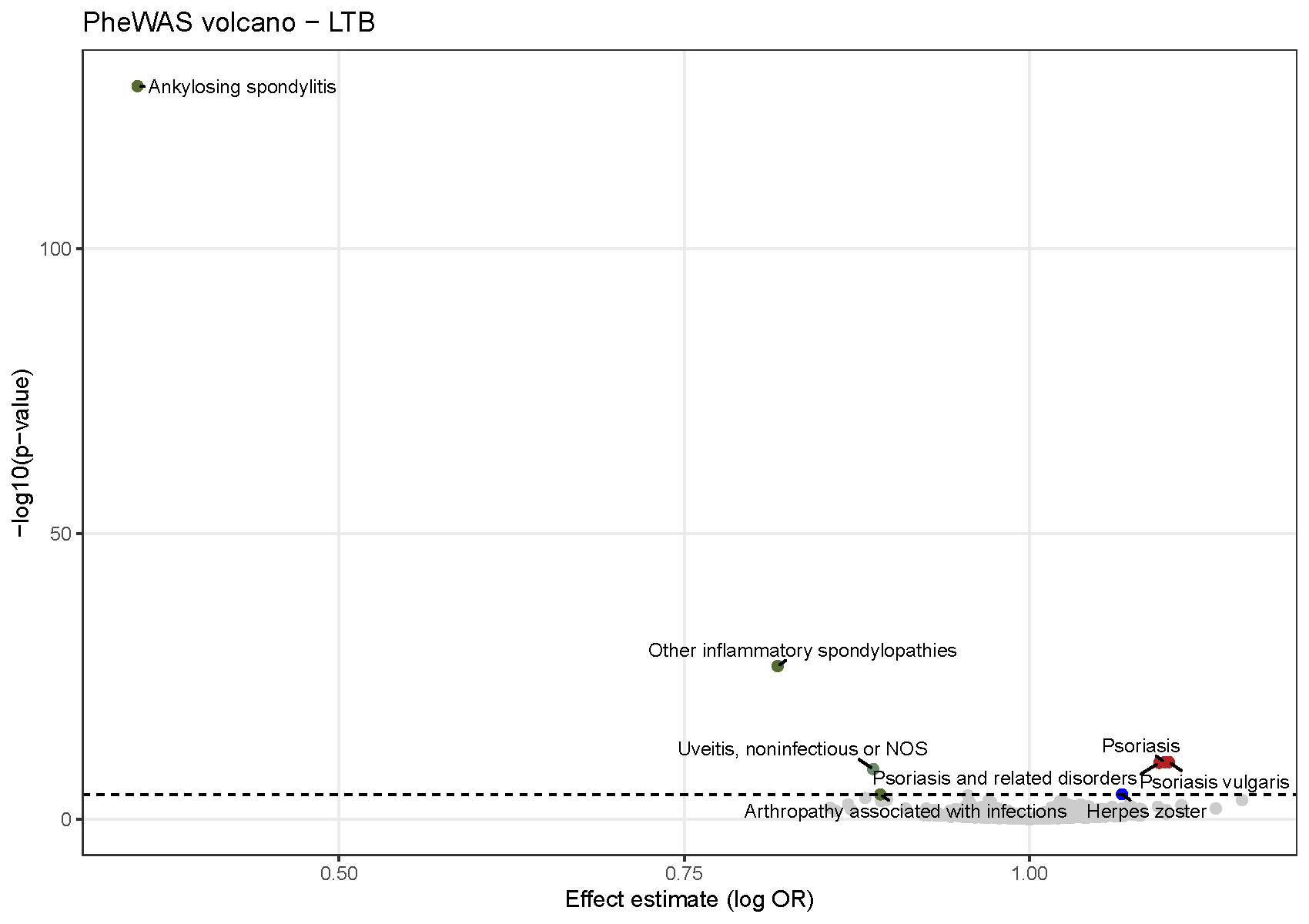

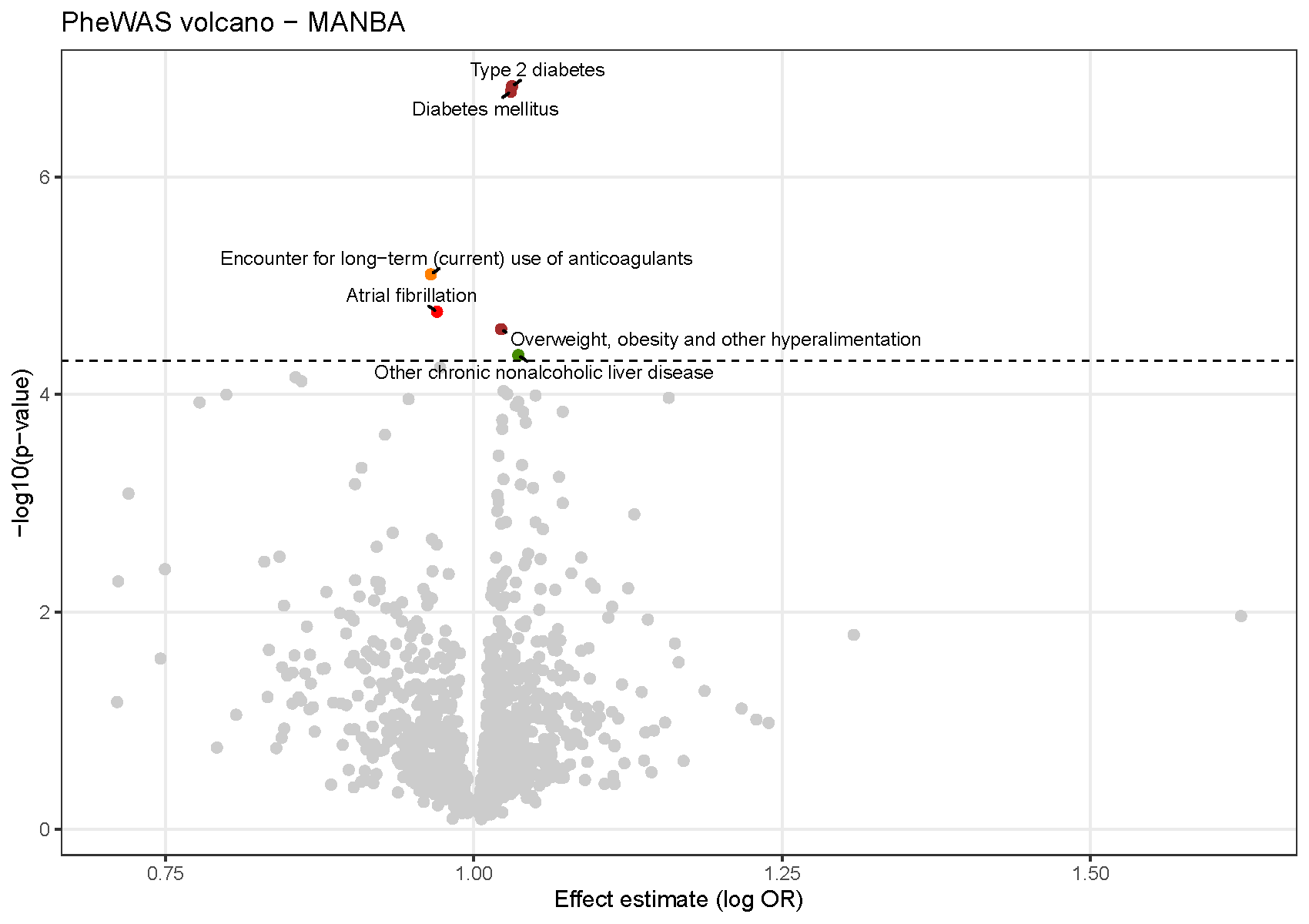

**Supplementary Figure 12.**

**Appendex 1. VA Million Veteran Program: Core Acknowledgements for Publications.**
